# Balanced Risk Integrity Index: a stratum-level metric for evaluating clinical prediction alerts under competing risks and censoring

**DOI:** 10.64898/2026.10.14.26357330

**Authors:** Inyong Jeong, Taeyeong Lee, Jin-Hyun Park, Yeongmin Kim, Hwamin Lee

**Author notes:** These authors contributed equally to this work (Inyong Jeong, Taeyeong Lee). **Corresponding author [Hwamin Lee]** Department of Biomedical Informatics, Korea University College of Medicine, 161 Jeongneung-ro, Seongbuk-gu, Seoul 02708, Republic of Korea.

## Abstract

**Background:** Conventional discrimination metrics, such as the area under the receiver operating characteristic curve (AUROC), do not characterize the composition of outcomes among patients flagged as high risk by clinical prediction models. This limitation is particularly relevant in critical care, where competing deaths and incomplete follow-up can complicate the interpretation of prediction alerts. We developed the Balanced Risk Integrity Index (BRII), a stratum-level metric defined as the proportion of target events among target events, competing deaths, and early-censored observations in a flagged high-risk stratum. BRII is algebraically related to positive predictive value (PPV) and is reported alongside its component decomposition to characterize the outcome composition of flagged patients.

**Methods:** We evaluated BRII using seven acute kidney injury (AKI) prediction models developed in MIMIC-IV and externally validated in MIMIC-III and eICU-CRD (116,010 patients in total). Additional simulations examined the effects of competing deaths and early censoring on BRII and conventional model performance metrics.

**Results:** For 48-hour AKI prediction in the top 10% risk stratum, BRII ranged from 0.90 to 0.96 in MIMIC-III, compared with less than 0.26 across all models in eICU-CRD, where 61.5% of evaluated patients were censored before the prediction horizon. In the eICU-CRD top 10% strata for the same target, 37.0% to 50.2% of patients were censored early, 12.2% to 15.5% had AKI, and 3.4% to 8.1% died before AKI. BRII and PPV ordered at least one model pair differently in 34 of 48 cohort-outcome-top-k groups. Under simulated random censoring for 48-hour AKI prediction, BRII declined by up to 0.80 while binary-label AUROC changed by less than 0.02.

**Conclusions:** BRII provided complementary information on high-risk stratum composition, yielding different model rankings from PPV and revealing substantial differences across external validation cohorts. Interpreted alongside PPV and its component decomposition, BRII offers a standardized approach to evaluating clinical prediction alerts across models and observation environments.

## Introduction

As clinical prediction models move into routine deployment, health systems increasingly need ways to verify that a model remains safe and useful after deployment, not only at the development stage [1, 2]. Yet critical-care prediction models are commonly evaluated using the area under the receiver operating characteristic curve (AUROC) and related discrimination metrics [3], which do not directly characterize the distribution of target events, competing deaths, and early censoring among patients flagged as high risk [4, 5]. Two processes further complicate the interpretation of high-risk alerts: competing death before the target event, which may be treated as a non-event in conventional binary outcome definitions [6], and discharge or transfer before the prediction horizon (early censoring), which can limit outcome ascertainment within the original intensive care unit (ICU) setting [7]. A multicenter randomized trial found that electronic AKI alerts did not improve patient outcomes [8]. Other studies have examined near-real-time AKI prediction [9], ward-based AKI prediction [10], external validation of a deployed sepsis model [11], and real-world performance of a commercial intracranial hemorrhage detector [12]. Together, these studies highlight the importance of evaluating clinical prediction systems beyond conventional discrimination metrics. A review of machine learning in acute respiratory distress syndrome emphasized the need to consider competing risks alongside discrimination [13], and a systematic review of COVID-19 prediction models highlighted deficiencies in accounting for competing events and censoring [14]. From discrimination alone, these reports cannot show whether a shortfall in benefit arises from ineffective intervention, workflow barriers, or the composition of the flagged stratum.

Competing-risk approaches, including cause-specific hazard models [15], Fine-Gray regression [16], and deep-learning methods such as DeepHit [17], allow competing events to be incorporated into prediction modeling. However, the availability of competing-risk prediction methods does not itself establish how the outcome composition of a clinically flagged high-risk stratum should be summarized. Competing-risk concordance indices and prediction-error measures [18–20], as well as time-dependent and inverse-probability-of-censoring-weighted (IPCW) AUROC variants [21], primarily assess predictive performance rather than explicitly describing the relative frequencies of target events, competing deaths, and early censoring within a flagged stratum.

Although positive predictive value (PPV) and net benefit provide complementary information about predictions and clinical decisions [22, 23], neither metric alone describes how competing deaths and early censoring contribute to the observed outcomes of a flagged stratum. Two strata with similar PPV may therefore differ substantially in their competing-death and censoring profiles. Cause-specific positive predictive values have been defined within marker-defined strata under competing risks, so that the target event and competing death are reported as separate stratum-level probabilities [24]. Kamran et al. highlighted a related limitation involving the timing of clinical prediction, emphasizing that conventional discrimination metrics may not adequately reflect opportunities for intervention [25]. The outcome composition of a flagged stratum matters particularly in intensive care, where competing mortality, short stays, and transfers may alter the distribution of outcomes and the completeness of follow-up among patients selected for clinical alerts. To our knowledge, a stratum-level proportion that also counts early censoring as a component and excludes event-free observed survivors from the denominator has not been reported.

Here we define the Balanced Risk Integrity Index (BRII) for a flagged high-risk stratum as the number of target events divided by the number of target events, competing deaths, and early-censored observations. BRII is computed from the same counts as PPV and equals PPV divided by the fraction of the flagged stratum that did not remain event-free and under observation through the prediction horizon. Many performance measures in routine use, the F-measure among them, are functions of the four counts of one confusion matrix, and this relation does not determine which measure suits a problem [26]. The accompanying decomposition reports competing death and early censoring separately. Complementing prior work on the temporal dimension of prediction, we asked what this proportion shows when it is reported with PPV and with its components. We evaluated it using seven AKI prediction models in three intensive care databases and under simulated competing-death and censoring conditions.

## Methods

### Study design and data sources

We conducted a retrospective cohort study using three PhysioNet critical care databases [27]: MIMIC-IV (version 2.2) [28], MIMIC-III (version 1.4, CareVue) [29], and the eICU Collaborative Research Database (eICU-CRD, v2.0) [30]. Because MIMIC-IV and MIMIC-III contain data from Beth Israel Deaconess Medical Center, MIMIC-III was restricted to CareVue records to reduce the possibility of patient overlap. The eICU-CRD includes data from more than 200 US hospitals.

### Cohort construction

We included adults aged 18 years or older with a first ICU stay lasting at least 24 hours. The prediction anchor was set at 24 hours after ICU admission, using data collected during the preceding 24-hour baseline window. Patients with AKI before or at the prediction anchor were excluded to focus on incident AKI after the prediction time. Patients without sufficient serum creatinine measurements to establish baseline kidney function were also excluded.

### Outcomes, predictors, and competing risks

AKI was defined using the KDIGO serum creatinine criteria [31] (**Supplementary Methods S1**). Four prediction targets were evaluated: any AKI and stage 2 or higher AKI within 48 or 168 hours after the prediction anchor (any_48h, any_7d, 2p_48h, and 2p_7d). Follow-up was limited to the corresponding prediction horizon. We used 42 predictors extracted during the first 24 hours of ICU admission, with missing values imputed using medians estimated exclusively from the training data within each cross-validation fold (**Supplementary Methods S2**). Death before the target event was treated as a negative label, a censoring event, or a competing event in binary, cause-specific, and competing-risk models, respectively. We additionally characterized the observed frequencies of target AKI, competing death, and early censoring at each horizon (**Supplementary Methods S1**).

### Model development

Seven models spanning the principal AKI-prediction paradigms [32] were trained in MIMIC-IV with five-fold stratified cross-validation: binary classifiers (logistic regression, XGBoost, LightGBM), cause-specific survival models (Cox proportional hazards, random survival forest [33]), and competing-risk models (Fine-Gray subdistribution hazard [34] and DeepHit [17]). Hyperparameters are given in **Supplementary Methods S3**.

Development-cohort results were computed from out-of-fold predictions. The seven models serve as a diverse testbed for evaluating BRII across paradigms rather than as individually optimized clinical tools. For external validation, each model was re-fitted on the full MIMIC-IV cohort and applied to MIMIC-III and eICU-CRD with frozen parameters [4, 35, 36], ranking patients by the model-specific risk quantity at each horizon (predicted probability, cumulative hazard, or cumulative incidence).

### Balanced Risk Integrity Index

BRII is the share of target events among flagged patients whose follow-up ended in a target event, a competing death, or early censoring. One minus BRII is the share of these patients whose follow-up ended in competing death or early censoring. BRII differs from PPV by excluding patients who remain event-free and under observation through the prediction horizon. The word balanced in the name refers to counting the target event against the two other ways in which follow-up ended, competing death and early censoring, with unit weights; the unit weights are a reference point and were not derived from a decision analysis. The accompanying component decomposition allows competing deaths and early censoring to be examined separately.

Within the top-k risk stratum (the k% of patients with the highest predicted risk), let *A*, *D*, *C*, and *S* denote the empirical proportions of patients classified by horizon *H* as having experienced the target event, competing death, early censoring, or remaining event-free and observed through *H*, respectively, such that *A* + *D* + *C* + *S* = 1. We define BRII as:

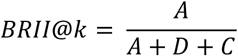

the proportion of target events among patients whose outcome resolved (target event or competing death) or whose observation ended (early censoring) by *H*; event-free observed survivors are counted in neither the numerator nor the denominator.

BRII can be expressed as *PPV*/(1 − *S*), where *S* is the event-free observed survivor fraction (derivation in **Supplementary Methods S4**). BRII is therefore an algebraic function of PPV and *S*, and given PPV and *S* it adds no information. Early censoring is a component of the denominator and is not a separate input. We report BRII because it expresses the two quantities as one proportion with a stated meaning, and we report it together with PPV and the four component proportions *A*, *D*, *C*, and *S*.

In the primary analysis, *A*, *D*, and *C* were empirical proportions within each stratum, so the same quantity is computed identically in development and validation cohorts. As a sensitivity analysis, *A* and *D* were re-estimated with the Aalen-Johansen estimator [37], with early-censored patients treated as censored at their last observation time. *C* was kept as an empirical proportion because it counts patients whose observation ended rather than a modeled event. The two estimators were compared across all 336 model-outcome-top-*k*-cohort combinations (**Supplementary Table 1**).

BRII ranges from 0 to 1 when *A* + *D* + *C* is positive. For fixed *A*, increasing *D* or *C* decreases BRII algebraically. When the number of patients contributing to *A* + *D* + *C* is small, BRII may have substantial sampling uncertainty. With m patients in the denominator, the normal-approximation half-width of a 95% interval is 1.96 × 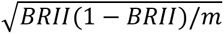. We therefore interpret estimates cautiously when event-free observed survivors exceed 90% of the stratum and report the denominator mass (*A* + *D* + *C*) alongside BRII.

We used a top-*k* approach to standardize the number of flagged patients across models, with *k* = 10% as the primary analysis and *k* = 1% to 20% in sensitivity analyses. Threshold-based analyses are provided in **Supplementary Table 2**. For descriptive purposes, we also defined four exploratory BRII zones based on the observed distribution in this study. These zones were not externally validated and should not be interpreted as clinical decision thresholds (zone boundaries in **Supplementary Methods S5**). We also described the decomposition of the top-10% stratum for the two 48-hour targets in each external cohort.

### Death-contamination sweep

To test whether BRII responds to competing-death burden beyond the population-level shift, we varied the target death fraction, defined as competing deaths divided by target events plus competing deaths, from 0.0 to 0.9 by resampling within the MIMIC-IV cohort while holding model predictions fixed, yielding 224,000 scenarios across four outcomes, seven models, and four top-*k* thresholds. Within an event type, patients were removed without regard to the model score. At each level we compared the slope of the target-event share *A*/(*A* + *D*) between the top-*k* and the complementary bottom stratum (**Supplementary Table 3**). Resampling procedure, iteration counts, and the derivation of death-fraction crossing points are detailed in **Supplementary Methods S6**.

### Censoring-sensitivity sweep

We examined incomplete follow-up in the MIMIC-IV cohort under three censoring mechanisms (random censoring, and informative censoring biased toward lower- or higher-risk patients) across rates from 0.0 to 0.9, yielding 168,000 scenarios at the top-10% threshold. Censoring times were drawn from a right-skewed distribution concentrating loss within the first quarter of the horizon, reflecting the early departures typical of ICU care. Because the informative-censoring bias signal used one of the evaluated models (LightGBM), we repeated the sweep with an alternative signal (Cox) to check for circularity. Full parameterization is given in **Supplementary Methods S7**.

### External validation and observation-window analysis

Models trained on MIMIC-IV were evaluated in MIMIC-III and eICU-CRD at top-*k* thresholds of 1%, 2%, 5%, and 10%, following guidance for survival models with competing risks [38, 39] and targeted validation in the intended population [40]. AUROC was computed against binary AKI labels (target event within the horizon = 1; all other outcomes, including competing death and censoring = 0) to allow comparison across paradigms. Because Fine-Gray score saturation could preclude meaningful ranking, we examined its predicted score distributions and ran the primary BRII-AUROC correlation analysis (56 model-outcome-cohort combinations) both with and without Fine-Gray (**Supplementary Table 4**). In eICU-CRD, we removed only patients censored before the horizon, re-identified the top-*k* stratum from the frozen predictions, and evaluated BRII at the top-10% threshold (score distributions in **Supplementary Fig. 1**). We compared the resulting fold change in BRII with two reference ratios. The first is the ratio of AKI incidence in the restricted cohort to that in the full cohort. The second is the ratio (A + D + C)/(A + D) in the original top-10% stratum, which is the fold change obtained by removing early-censored patients from the BRII denominator without re-identifying the stratum (**Supplementary Methods S8**).

### Complementary analyses

To position BRII against standard and decision-analytic metrics, we regressed BRII on AUROC, AUPRC, PPV, and F1 at top-10% across all 84 cohort-model-outcome conditions and separately by cohort (n = 28 each) and by outcome (n = 21 each). For each cohort, outcome, and top-k level, we counted the model pairs (21 per group) that BRII and PPV ordered differently; pairs with values equal to three decimals were counted as ties. We also compared BRII with net benefit computed under fixed-threshold and prevalence-matched specifications. To examine whether strata with equal net benefit differ in composition, we examined BRII decomposition within matched net-benefit bins. The primary BRII gives competing death and early censoring unit weights, so that it remains a proportion of patients. Sensitivity of BRII rankings to other weights was assessed with a weighted variant at five death-fraction levels of the death-contamination sweep. Formulae, net-benefit specifications, binning rules, and the weight grid are given in **Supplementary Methods S9**.

Within each top-k stratum, we estimated Aalen-Johansen cumulative incidences of the target event and competing death and their ratio, and compared model rankings by this ratio and by BRII (**Supplementary Methods S10**). We computed time-dependent AUROC at half the horizon and IPCW concordance, with weights from each cohort’s own censoring distribution and competing death treated as censoring, and correlated each with BRII at the top-10% threshold (**Supplementary Methods S11**). The death-contamination sweep was repeated with removal probability proportional to the rank of a reference score, in either direction (**Supplementary Methods S12**). Calibration was assessed in deciles of predicted risk, with observed risk taken as the binary proportion and as the Aalen-Johansen cumulative incidence (**Supplementary Methods S13**). Top-k BRII depends on the score only through its ranking and is invariant to monotone recalibration.

### Statistical analysis

All resampling used fixed random seeds. Point-estimate BRII values were reported with 95% confidence intervals from 1,000 within-stratum bootstrap samples (empirical 2.5th and 97.5th percentiles). These intervals are conditional on the given flagged stratum and do not include the uncertainty of selecting it; a bootstrap that also re-selected the stratum, values under extreme assignments of early-censored patients, and weights that include a censoring weight of zero are described in **Supplementary Methods S14**. Neither bootstrap includes the uncertainty of refitting the model. For the death-contamination and censoring sweeps, intervals came from empirical percentiles across 200 repetitions. Rank associations used Spearman correlation, and all analyses used Python 3.11.

## Results

### The three cohorts differ in competing-death and censoring profiles

The three cohorts (MIMIC-IV n = 33,455; MIMIC-III n = 14,514; eICU-CRD n = 68,041) showed distinct competing-risk profiles (Table 1). Death fractions were 0.118 (any_48h) and 0.579 (2p_48h) in MIMIC-IV and 0.344 and 0.826 in eICU-CRD. The eICU-CRD cohort also had shorter observation: for the 48-hour horizon, median follow-up among patients without a target event or death was 27.1 hours in eICU-CRD and 48.0 hours in both MIMIC cohorts. Cohort-level frequencies of the three outcomes are given in **Supplementary Table 5**.

**Table 1.** Cohort characteristics and competing-risk profiles across three databases.

| Characteristic | MIMIC-IV | MIMIC-III | eICU-CRD |
| --- | --- | --- | --- |
| **n** | 33,455 | 14,514 | 68,041 |
| **Age, median [IQR]** | 66 [54-77] | 64 [51-76] | 65 [53-75] |
| **Female, %** | 44.1 | 42.1 | 45.9 |
| **Any AKI, 48h: AKI rate** | 3,692 (11.0%) | 1,663 (11.5%) | 3,320 (4.9%) |
| **Any AKI, 48h: Deaths** | 493 | 225 | 1,739 |
| **Any AKI, 48h: $p_0$** | 0.118 | 0.119 | 0.344 |
| **Any AKI, 48h: Early censoring, %** | 6.7 | 7.6 | 61.5 |
| **Any AKI, 7d: AKI rate** | 4,907 (14.7%) | 2,335 (16.1%) | 4,145 (6.1%) |
| **Any AKI, 7d: Deaths** | 1,103 | 534 | 3,136 |
| **Any AKI, 7d: $p_0$** | 0.184 | 0.186 | 0.431 |
| **Any AKI, 7d: Early censoring, %** | 31.9 | 39.4 | 83.0 |
| **Stage $\geq 2$, 48h: AKI rate** | 405 (1.2%) | 166 (1.1%) | 402 (0.6%) |
| **Stage $\geq 2$, 48h: Deaths** | 558 | 246 | 1,910 |
| **Stage $\geq 2$, 48h: $p_0$** | 0.579 | 0.597 | 0.826 |
| **Stage $\geq 2$, 48h: Early censoring, %** | 6.9 | 7.8 | 62.6 |
| **Stage $\geq 2$, 7d: AKI rate** | 1,041 (3.1%) | 506 (3.5%) | 922 (1.4%) |
| **Stage $\geq 2$, 7d: Deaths** | 1,329 | 635 | 3,632 |
| **Stage $\geq 2$, 7d: $p_0$** | 0.561 | 0.557 | 0.798 |
| **Stage $\geq 2$, 7d: Early censoring, %** | 35.5 | 44.4 | 85.9 |

Demographic characteristics and outcome distributions for the development cohort (MIMIC-IV) and two external validation cohorts (MIMIC-III, eICU-CRD). Rate: target event incidence. $p_0$ : death fraction among resolved events, defined as death / (AKI + death). Age reported as median [IQR].

### BRII in the development cohort is high for any AKI at 48 hours and lower for stage 2 or higher AKI at 48 hours

In MIMIC-IV, top-10% BRII spanned 0.881 to 0.962 for any_48h and 0.224 to 0.545 for 2p_48h (**Table 2**). BRII and PPV diverge when event-free observed survivors dominate the stratum: LGBM reached BRII 0.962 but PPV 0.412 for any_48h, with 57.2% event-free observed survivors. The denominator mass *A* + *D* + *C* was large for most targets but fell to 6.8 to 14.7% for 2p_48h, around the caution threshold defined in Methods, so BRII for this target is interpreted with caution throughout.

**Table 2.** Development cohort BRII and standard performance metrics.

| Eval | Model | BRII<br>(95% CI) | AUROC | PPV | Sensitivity | F1 | Net Benefit | AKI% | Death% | Cens% | Event-free% |
| --- | --- | --- | --- | --- | --- | --- | --- | --- | --- | --- | --- |
| Any 48h | LR | 0.931 (0.919-0.944) | 0.788 | 0.412 | 0.373 | 0.391 | +0.035 | 41.2 | 1.8 | 1.2 | 55.8 |
| Any 48h | XGB | 0.955 (0.944-0.967) | 0.758 | 0.377 | 0.342 | 0.358 | +0.031 | 37.7 | 0.8 | 0.9 | 60.5 |
| Any 48h | LGBM | 0.962 (0.952-0.971) | 0.795 | 0.412 | 0.373 | 0.391 | +0.035 | 41.2 | 1.1 | 0.5 | 57.2 |
| Any 48h | CS_CoxPH | 0.920 (0.905-0.933) | 0.784 | 0.408 | 0.370 | 0.388 | +0.034 | 40.8 | 2.3 | 1.3 | 55.6 |
| Any 48h | CS_RSF | 0.881 (0.862-0.897) | 0.774 | 0.371 | 0.336 | 0.353 | +0.030 | 37.1 | 3.3 | 1.7 | 57.9 |
| Any 48h | DeepHit | 0.921 (0.905-0.935) | 0.747 | 0.365 | 0.331 | 0.347 | +0.029 | 36.5 | 2.0 | 1.1 | 60.4 |
| Any 48h | FineGray | 0.934 (0.921-0.946) | 0.784 | 0.409 | 0.371 | 0.389 | +0.034 | 40.9 | 1.7 | 1.2 | 56.2 |
| Any 7d | LR | 0.635 (0.613-0.654) | 0.738 | 0.437 | 0.298 | 0.354 | +0.037 | 43.7 | 4.3 | 20.8 | 31.2 |
| Any 7d | XGB | 0.594 (0.573-0.613) | 0.717 | 0.410 | 0.279 | 0.332 | +0.034 | 41.0 | 2.8 | 25.1 | 31.0 |
| Any 7d | LGBM | 0.611 (0.592-0.631) | 0.754 | 0.441 | 0.301 | 0.357 | +0.038 | 44.1 | 3.1 | 24.9 | 27.9 |
| Any 7d | CS_CoxPH | 0.634 (0.615-0.652) | 0.733 | 0.448 | 0.305 | 0.363 | +0.039 | 44.8 | 4.5 | 21.4 | 29.4 |
| Any 7d | CS_RSF | 0.600 (0.581-0.620) | 0.738 | 0.416 | 0.283 | 0.337 | +0.035 | 41.6 | 6.0 | 21.7 | 30.7 |
| Any 7d | DeepHit | 0.517 (0.498-0.537) | 0.704 | 0.390 | 0.266 | 0.316 | +0.032 | 39.0 | 3.4 | 32.9 | 24.7 |
| Any 7d | FineGray | 0.630 (0.611-0.650) | 0.733 | 0.448 | 0.305 | 0.363 | +0.039 | 44.8 | 3.6 | 22.7 | 28.9 |
| 2p 48h | LR | 0.545 (0.486-0.605) | 0.781 | 0.049 | 0.402 | 0.087 | -0.006 | 4.9 | 2.5 | 1.6 | 91.1 |
| 2p 48h | XGB | 0.498 (0.433-0.562) | 0.667 | 0.034 | 0.279 | 0.060 | -0.007 | 3.4 | 0.9 | 2.5 | 93.2 |
| 2p 48h | LGBM | 0.482 (0.421-0.550) | 0.666 | 0.033 | 0.272 | 0.059 | -0.007 | 3.3 | 1.3 | 2.2 | 93.2 |
| 2p 48h | CS_CoxPH | 0.377 (0.330-0.420) | 0.763 | 0.045 | 0.375 | 0.081 | -0.006 | 4.5 | 5.1 | 2.4 | 88.0 |
| 2p 48h | CS_RSF | 0.308 (0.268-0.348) | 0.747 | 0.042 | 0.348 | 0.075 | -0.006 | 4.2 | 6.3 | 3.2 | 86.3 |
| 2p 48h | DeepHit | 0.224 (0.189-0.259) | 0.686 | 0.033 | 0.272 | 0.059 | -0.007 | 3.3 | 8.3 | 3.1 | 85.3 |
| 2p 48h | FineGray | 0.427 (0.380-0.475) | 0.770 | 0.047 | 0.388 | 0.084 | -0.006 | 4.7 | 4.2 | 2.1 | 89.0 |
| 2p 7d | LR | 0.227 (0.206-0.249) | 0.714 | 0.096 | 0.309 | 0.147 | -0.000 | 9.6 | 10.4 | 22.3 | 57.6 |
| 2p 7d | XGB | 0.160 (0.141-0.178) | 0.625 | 0.069 | 0.223 | 0.106 | -0.003 | 6.9 | 3.7 | 32.8 | 56.6 |
| 2p 7d | LGBM | 0.210 (0.190-0.233) | 0.681 | 0.089 | 0.285 | 0.135 | -0.001 | 8.9 | 5.2 | 28.3 | 57.7 |
| 2p 7d | CS_CoxPH | 0.211 (0.191-0.231) | 0.709 | 0.094 | 0.301 | 0.143 | -0.001 | 9.4 | 10.3 | 24.6 | 55.7 |
| 2p 7d | CS_RSF | 0.211 (0.190-0.233) | 0.709 | 0.091 | 0.293 | 0.139 | -0.001 | 9.1 | 14.6 | 19.4 | 56.9 |
| 2p 7d | DeepHit | 0.165 (0.146-0.183) | 0.663 | 0.075 | 0.240 | 0.114 | -0.003 | 7.5 | 18.0 | 19.8 | 54.7 |
| 2p 7d | FineGray | 0.214 (0.193-0.236) | 0.709 | 0.094 | 0.304 | 0.144 | -0.001 | 9.4 | 8.9 | 25.7 | 55.9 |

BRII with 95% bootstrap confidence intervals (1,000 resamples), AUROC, PPV, sensitivity, F1, net benefit, BRII decomposition components (AKI%, Death%, Cens%), and the percentage of event-free observed survivors (Event-free%) for seven models evaluated at the 10% top- $k$ threshold in MIMIC-IV. PPV, sensitivity, and F1 were computed using the model-specific probability threshold corresponding to the top-10% stratum boundary. Net benefit was computed by flagging the top 10% of patients ( $k = 0.10$ ) and applying threshold probability $p_t = 0.10$ in the standard formulation $NB = TP/N - (FP/N) \times [p_t/(1 - p_t)]$ . BRII decomposition components were estimated using empirical event proportions within the top 10% stratum. Eval: prediction target (Any, any-stage AKI; 2p, stage 2 or higher AKI). FineGray: Fine-Gray model.

### BRII declines as the competing-death burden increases

In the death-contamination sweep (**Fig. 1**), BRII declined monotonically with the death fraction across all outcomes and model families. Part of this decline follows from the metric’s definition, because competing deaths enter the BRII denominator. The target-event share *A*/(*A* + *D*) fell more slowly in the top-*k* stratum than in the complementary bottom stratum in 24 of 28 model-outcome combinations when patients were removed without regard to the model score (mean slope difference 0.13; **Supplementary Table 3**). The difference was strongest for any-AKI targets and weaker or reversed for stage ≥ 2 AKI, which accounted for the four exceptions. For any_48h, BRII exceeded 0.88 with no competing deaths but fell below 0.50 beyond a death fraction of about 0.7, whereas BRII for the rarer 2p_48h target converged toward low values (**Supplementary Table 6** and **Supplementary Fig. 2**).

**Figure 1.**
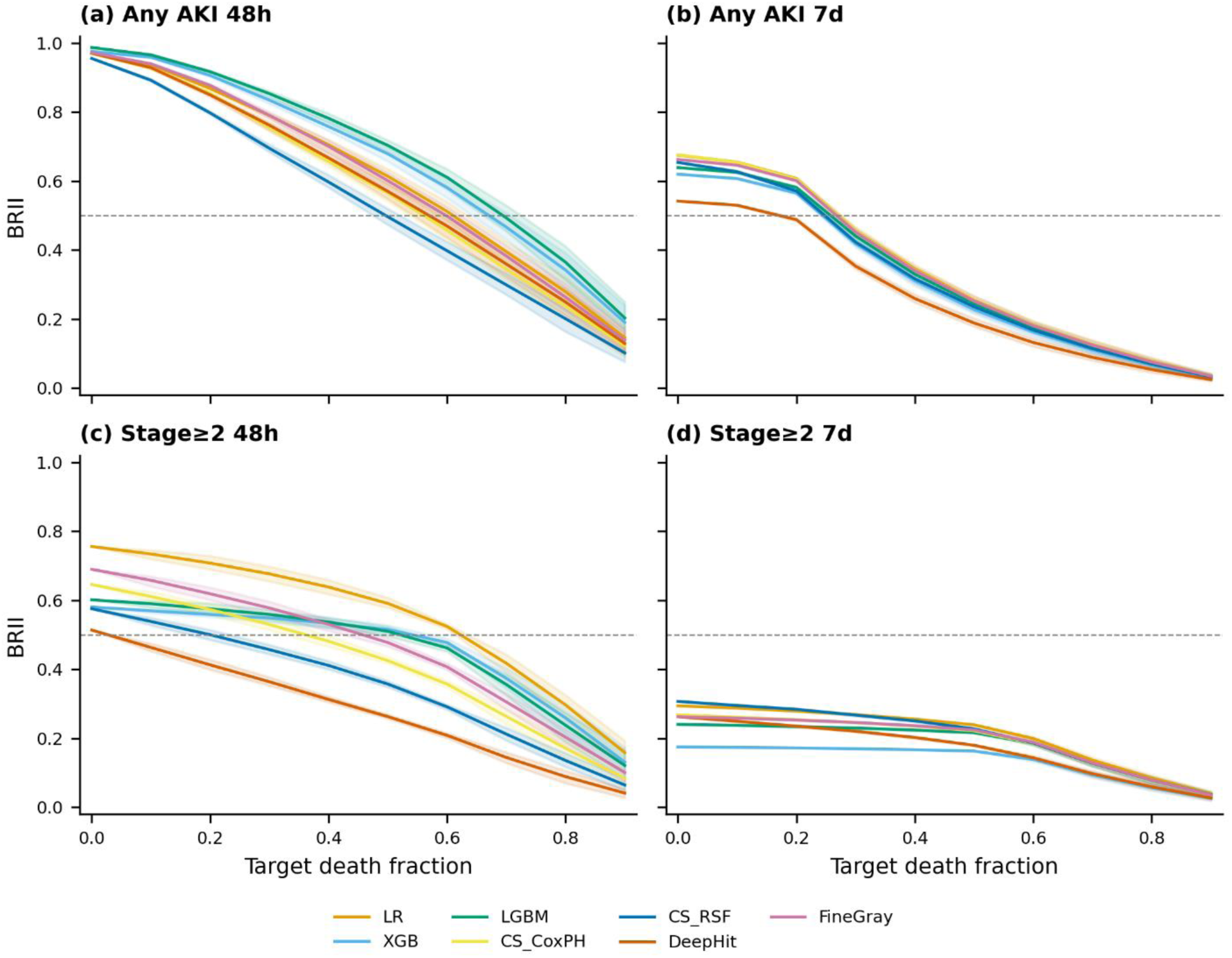
BRII declines monotonically with increasing competing-death burden. BRII at the top-10% threshold plotted against target death fraction (0.0-0.9) for seven models across four prediction targets (a, any_48h; b, any_7d; c, 2p_48h; d, 2p_7d). Points represent means across 200 resampling iterations; shaded bands denote empirical 95% intervals. Dashed horizontal line indicates BRII = 0.50.

### Censoring lowers BRII, and AUROC remains stable under random censoring

In the censoring sweep (**Fig. 2**), BRII declined with the censoring rate under all three mechanisms. Compared with random censoring, the decline was faster under censoring biased toward higher-risk patients and slower under censoring biased toward lower-risk patients. At a nominal censoring rate of 0.5 for any_48h, at which 51% to 52% of patients were censored early under each mechanism, mean BRII across the seven models was 0.34 to 0.39 under random censoring, 0.14 to 0.18 under censoring biased toward higher-risk patients, and 0.72 to 0.87 under censoring biased toward lower-risk patients. Under random censoring, AUROC changed little (**Fig. 3**). For any_48h, LGBM BRII fell from 0.962 to 0.169 while AUROC shifted only from 0.795 to 0.799, and this pattern was consistent across all seven models. Under censoring biased toward lower-risk and higher-risk patients, AUROC for any_48h changed by up to 0.08 and 0.10. In this sweep the flagged stratum was fixed and only the outcome labels changed, so the decline in BRII follows from the metric’s definition: censored patients enter the denominator.

**Figure 2.**
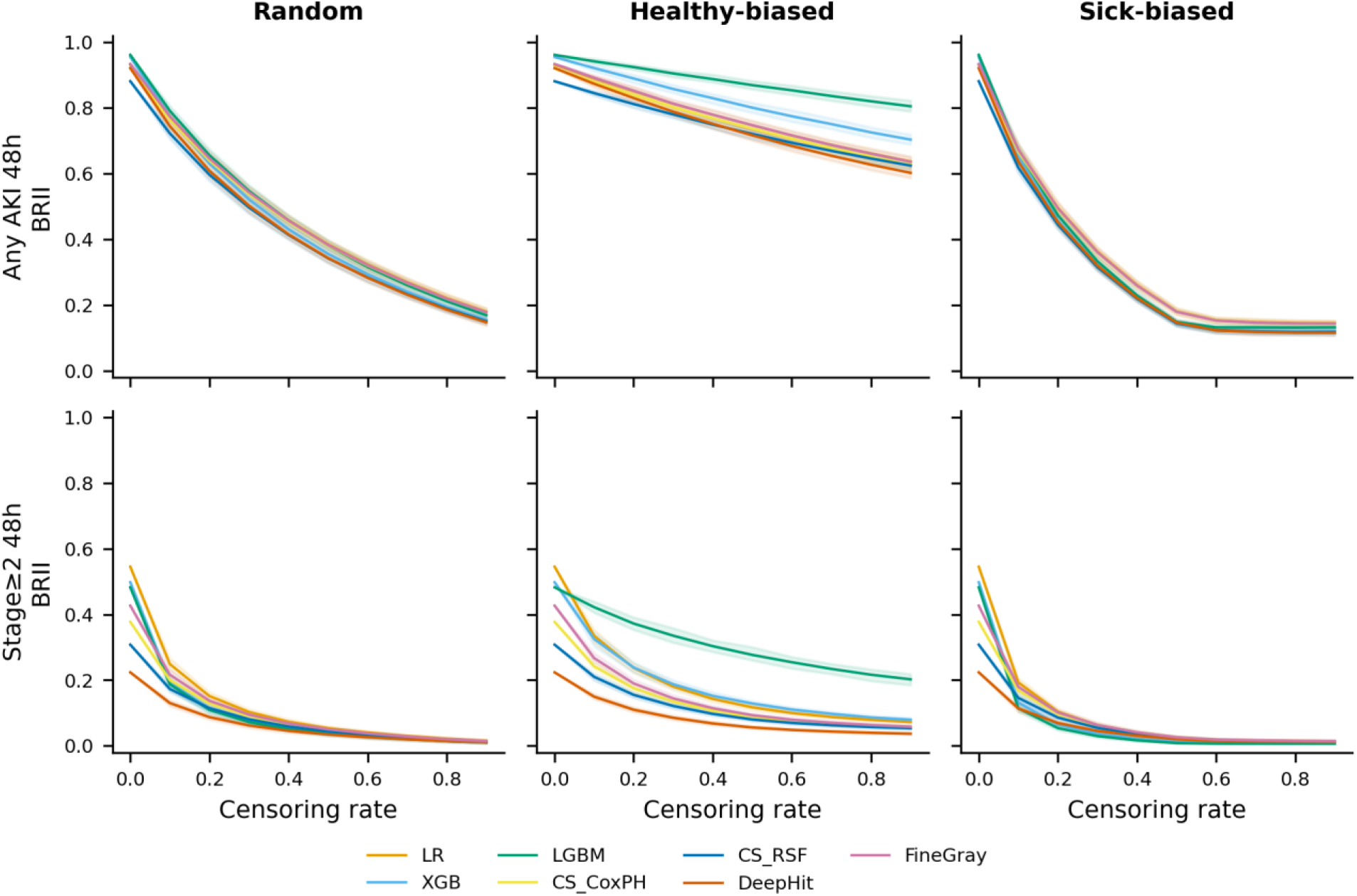
Compared with random censoring, BRII declines faster under censoring biased toward higher-risk patients and slower under censoring biased toward lower-risk patients. BRII at the top-10% threshold plotted against censoring rate (0.0-0.9) under three censoring mechanisms for seven models. Top row: any_48h. Bottom row: 2p_48h. Columns from left to right: random censoring (Random), informative censoring biased toward lower-risk patients (Healthy-biased), and informative censoring biased toward higher-risk patients (Sick-biased). Shaded bands denote empirical 95% intervals across 200 repetitions.

**Figure 3.**
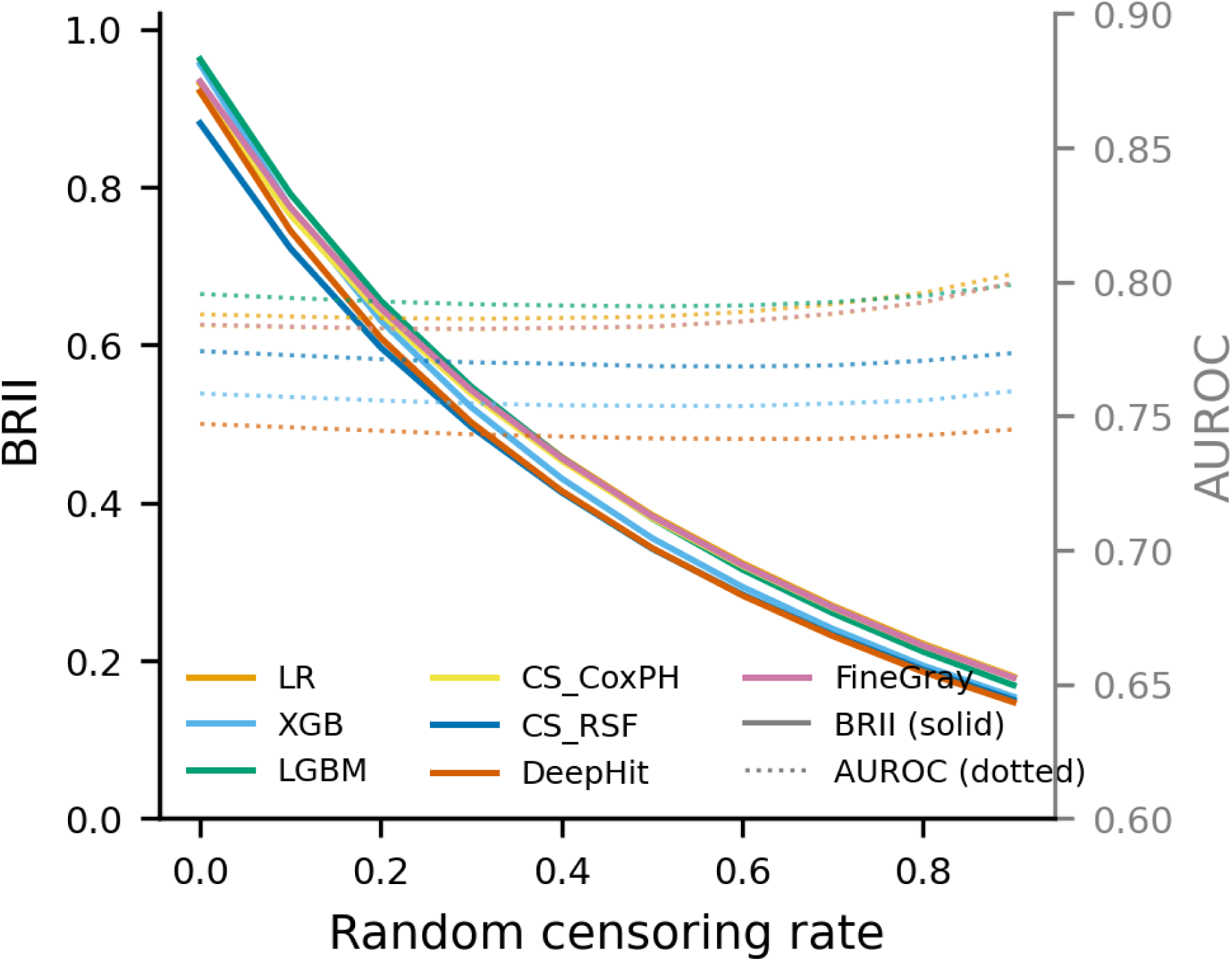
AUROC remains stable while BRII declines under increasing random censoring. BRII (solid lines, left axis) and AUROC (dotted lines, right axis) plotted against random censoring rate (0.0-0.9) for seven models in the any_48h target at top-10%. BRII declines by 0.73-0.80 across models while AUROC changes by less than 0.02.

### BRII differs between the two external validation cohorts

In MIMIC-III, BRII remained high for any_48h (0.899 to 0.960 across all seven models). For the rarer 2p_48h, XGB had a higher BRII than every cause-specific and competing-risk model despite a lower AUROC, with fewer deaths and censored patients in its top-10% stratum, whereas LGBM did not. In eICU-CRD, BRII was low across all models and outcomes: none exceeded 0.26 for any_48h, so target events made up at most about one quarter of the denominator mass. Binary classifiers had less competing death but far more early censoring in their top-10% strata than cause-specific models (**Table 3**).

**Table 3.**
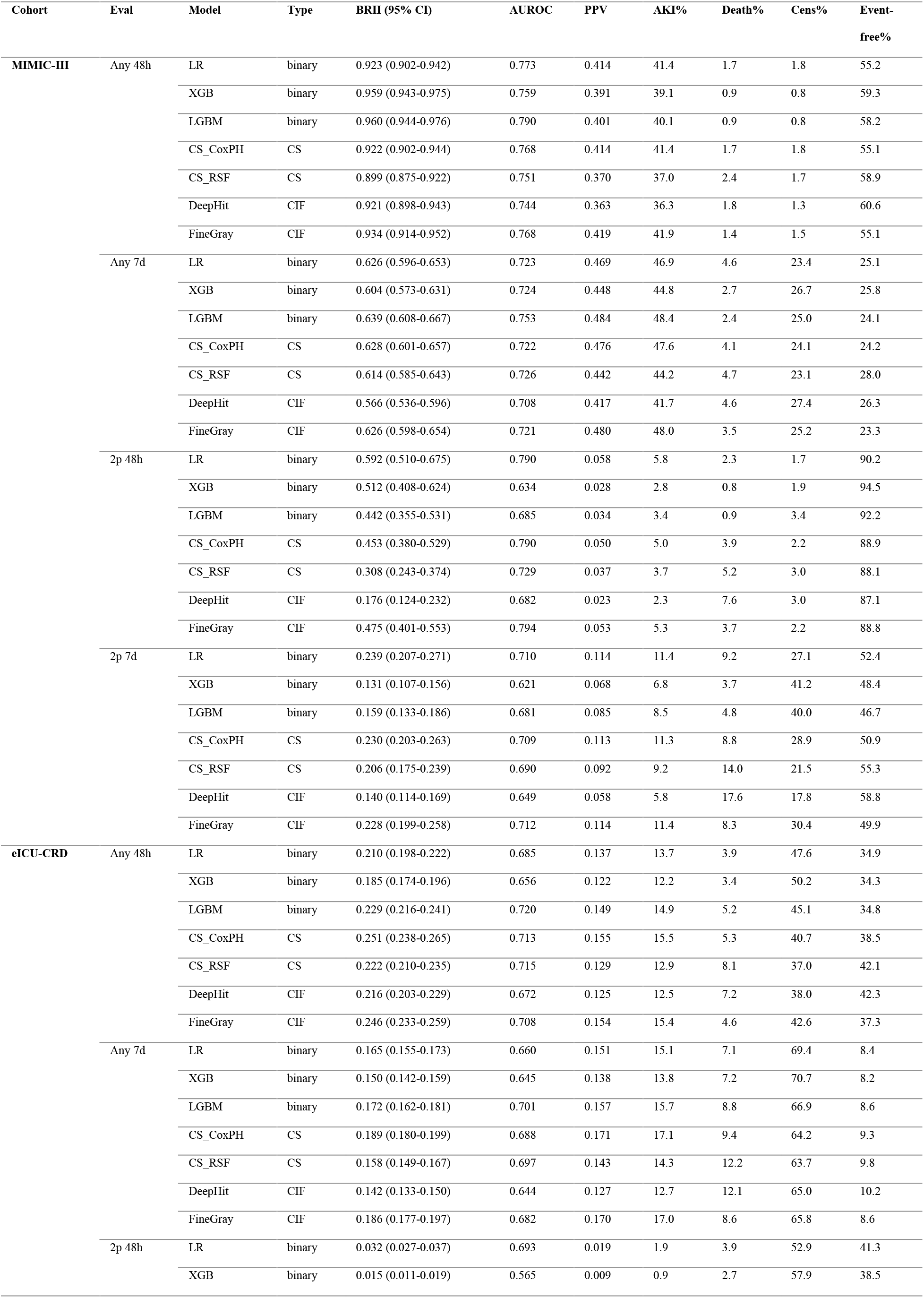
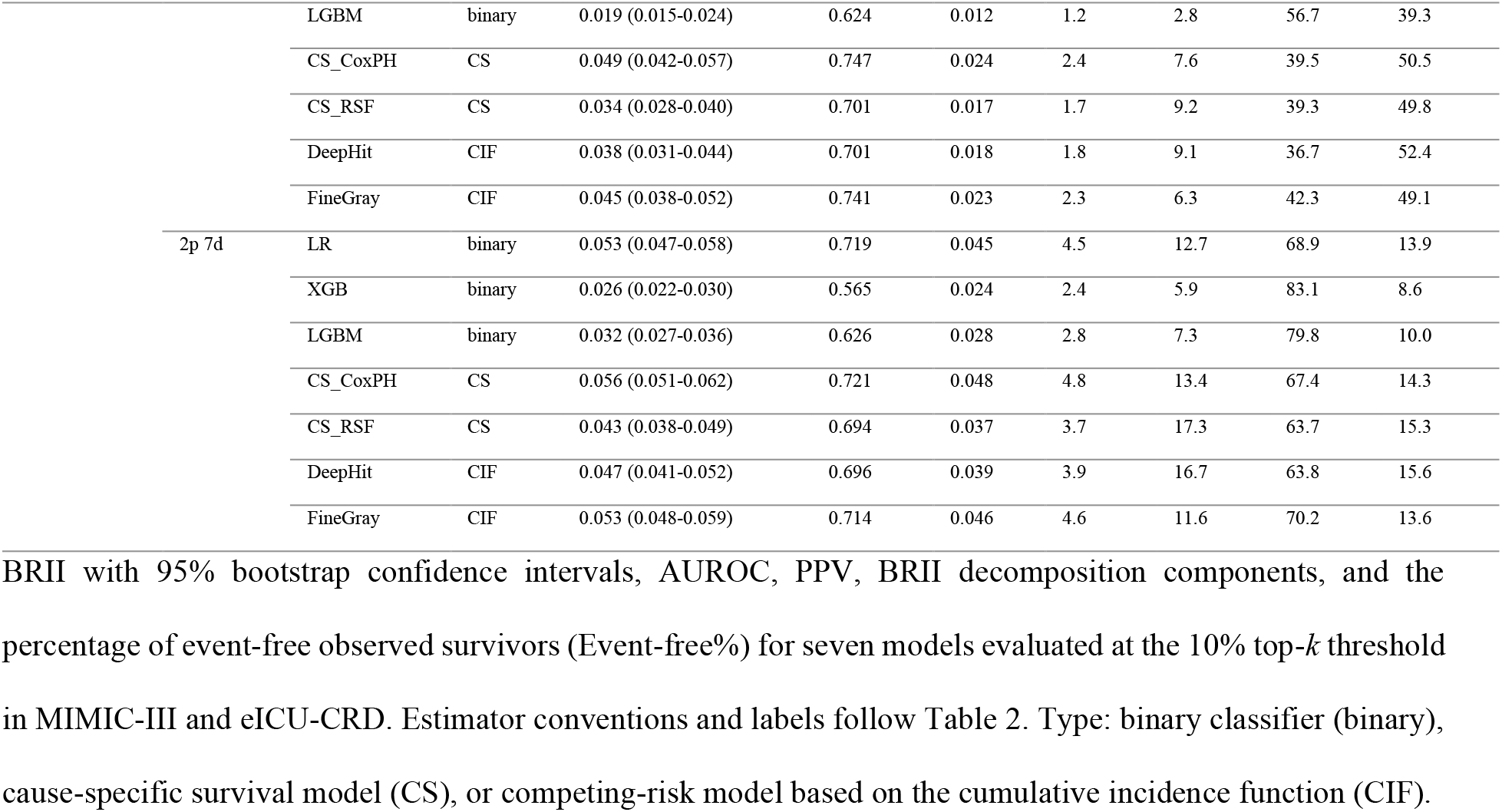
External validation BRII and standard performance metrics.

In eICU-CRD, BRII increased in all model-outcome combinations after patients censored before the horizon were removed and the top-*k* stratum was re-identified from the frozen predictions (**Fig. 4**). BRII rose 2.7-fold to 4.3-fold for any_48h and 4.5-fold to 17.5-fold for 2p_48h. In all 28 model-outcome combinations, the fold change was 0.97 to 1.12 times the ratio (*A* + *D* + *C*)/(*A* + *D*) in the original stratum, which is the change obtained by removing early-censored patients from the BRII denominator. The fold change was closest to this ratio for any_48h (0.99 to 1.03 times) and furthest above it for 2p_7d (1.08 to 1.12 times in four of seven models), where re-identifying the stratum added to the change. The fold change was above the AKI incidence ratio in all seven models for both 48-hour targets and in 2 of 14 model-outcome combinations for 168-hour targets (XGB and LGBM for 2p_7d; **Supplementary Table 7** and **Supplementary Fig. 1**).

**Figure 4.**
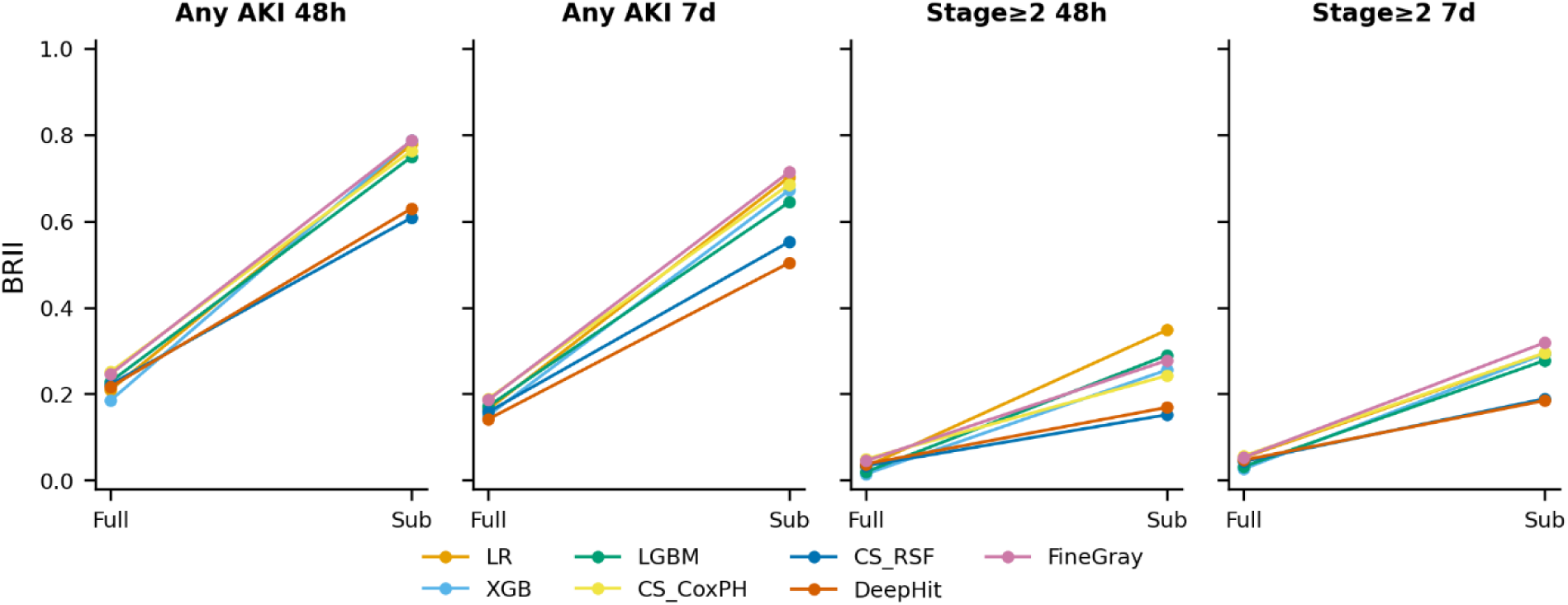
BRII in eICU-CRD increases after removal of patients censored before the horizon. BRII at top-10% for the full eICU-CRD cohort (left) versus the subcohort remaining after removal of patients censored before the horizon (right), for seven models across four outcomes. Connected lines link the same model across conditions.

### BRII decomposition reveals three distinct outcome profiles

Decomposition of the top-10% stratum showed three profiles (**Fig. 5**). High BRII occurred with predominantly target events (MIMIC-III any_48h). Lower BRII occurred in a rare target with few patients in any of the three BRII categories (MIMIC-III 2p_48h, XGB reaching 0.512 with minimal death and censoring). Low BRII occurred with more censoring than target events (eICU-CRD any_48h, XGB 12.2% target events against 50.2% censoring; eICU-CRD 2p_48h had the same profile). **Supplementary Table 8** gives the zone of each model-cohort-outcome combination, and **Supplementary Fig. 3** shows an example flowchart linking zones and decomposition profiles to possible responses.

**Figure 5.**
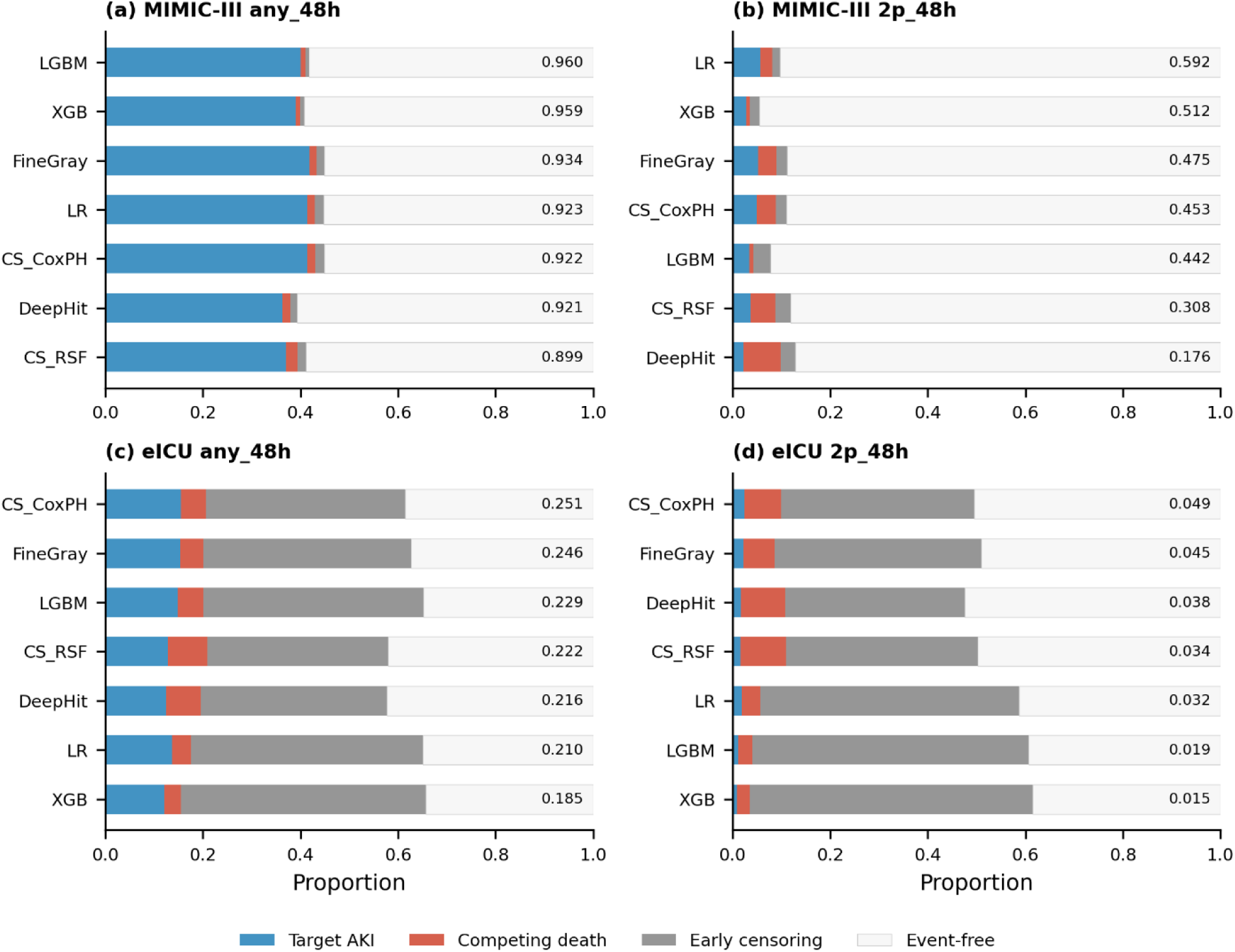
BRII decomposition reveals distinct outcome profiles across cohorts and models. Stacked bar plots showing the composition of the top-10% risk stratum for seven models in four cohort-outcome combinations. Components: target AKI (blue), competing death (red), early censoring (grey), event-free observed survivors (white, labeled Event-free). (a) MIMIC-III any_48h, (b) MIMIC-III 2p_48h, (c) eICU any_48h, (d) eICU 2p_48h. Models ordered by descending BRII within each panel; the number at the end of each bar is BRII.

### Binary-label AUROC alone explains less BRII variation than four standard metrics together

Across 56 external combinations at the top-10% threshold, the Spearman correlation between BRII and binary-label AUROC was *ρ* = 0.671. Within cohorts (n = 28 each), *ρ* was 0.705 in MIMIC-III and 0.304 in eICU-CRD, the cohort with the most early censoring, where the correlation was not statistically significant (p = 0.116 with the 28 combinations treated as independent; Fine-Gray sensitivity in **Supplementary Table 4**). In cohort-specific regressions of BRII on AUROC, AUPRC, PPV, and F1 (n = 28 per cohort), the adjusted R^2^ for AUROC alone was 50.3% in MIMIC-IV, 42.5% in MIMIC-III, and 1.5% in eICU-CRD, and the adjusted R^2^ for the four metrics together was 78.4%, 74.3%, and 91.3%. In outcome-specific regressions (n = 21 per outcome), the adjusted R^2^ for the four metrics together was lowest for the rarest target, 2p_48h (87.7%, versus 98.9% for any_48h and any_7d and 95.0% for 2p_7d), which had the highest death fraction.

### Estimator, weight, and bias-signal choices change model rankings in some conditions

Aalen-Johansen and empirical BRII differed by at most 0.014 in MIMIC-IV, 0.026 in MIMIC-III, and 0.073 in eICU-CRD, and the model rank changed in 64 of 336 combinations, by at most two positions (**Supplementary Table 1**). At a fixed probability threshold, the fraction of patients flagged differed across models, for example from 37% to 99% at a threshold of 0.10 for any_48h in MIMIC-IV (**Supplementary Table 2**). At top-k thresholds of 1%, 2%, and 5%, any_48h BRII was 0.903 to 1.000 in MIMIC-III and 0.235 to 0.447 in eICU-CRD (**Supplementary Table 9**). Fine-Gray scores had a compressed range in the external cohorts, and excluding Fine-Gray raised the BRII-AUROC rank correlation by 0.02 to 0.03 across thresholds (**Supplementary Table 4**).

Under alternative competing-death and censoring weights, model rankings were stable only for any_48h, for which the Spearman correlation with the unit-weight ranking was at least 0.93 in all 95 weight and death-fraction conditions. The correlation fell below 0.90 in 33 of 95 conditions for any_7d (minimum 0.21), 26 for 2p_48h (minimum 0.75), and 32 for 2p_7d (minimum −0.07; **Supplementary Table 10**). When the informative-censoring sweep was repeated with the alternative bias signal, the mean absolute difference in BRII between the two signals was at most 0.03 under censoring biased toward higher-risk patients and up to 0.12 under censoring biased toward lower-risk patients, and model rankings differed between the signals under both mechanisms (**Supplementary Table 11**). Comparisons between models under censoring biased toward lower-risk patients therefore depend on the bias signal.

In the death-contamination sweep for any_48h, BRII and net benefit had a Spearman correlation of 0.982. In 18 of 20 net-benefit bins, the 2.5th to 97.5th percentile range of BRII within the bin exceeded 0.05 or that of the death share exceeded 0.10; in these bins net benefit was between −0.001 and 0.036 and the BRII range was 0.06 to 0.27. The corresponding counts were 7, 20, and 16 of 20 bins for any_7d, 2p_48h, and 2p_7d (**Supplementary Fig. 4** and **Supplementary Table 12**).

### BRII and PPV order at least one model pair differently in 34 of 48 groups

BRII and PPV ordered at least one model pair differently in 34 of 48 cohort-outcome-top-k groups (138 of 1,008 pairs; 64 in MIMIC-IV, 63 in MIMIC-III, and 11 in eICU-CRD; **Supplementary Table 13**). The groups share the same seven models and nested outcomes, so these counts are descriptive. BRII and the within-stratum Aalen-Johansen ratio of target-event to target-event plus competing-death cumulative incidence ordered 329 of 1,008 pairs differently, in 45 of 48 groups (**Supplementary Table 14**). The two measures estimate different quantities: the ratio is the target-event share of cumulative incidences when early-censored patients are redistributed under independent censoring, and BRII is the target-event share of the observed composition that counts early-censored patients as a third component. Between eICU-CRD and MIMIC-III, the bootstrap interval of the difference excluded zero in 110 of 112 outcome-model-k combinations for BRII and in 104 for the Aalen-Johansen ratio, which was lower in eICU-CRD in all 104. In 79 of 112 combinations, the between-cohort difference in BRII differed from that in the Aalen-Johansen ratio (the interval of the difference between the two differences excluded zero).

### In eICU-CRD, three discrimination measures have correlations of 0.30 to 0.39 with BRII

At the top-10% threshold (n = 28 combinations per cohort), using each cohort’s own censoring distribution for the inverse-probability weights, the Spearman correlation of BRII with time-dependent AUROC at H/2 was 0.681 in MIMIC-III and 0.352 in eICU-CRD, and with IPCW concordance 0.699 and 0.389; the corresponding correlations with binary AUROC were 0.705 and 0.304 (**Supplementary Table 15**). In eICU-CRD, the correlation with time-dependent AUROC exceeded that with binary AUROC by 0.048 (95% bootstrap interval −0.279 to 0.354) and the correlation with IPCW concordance exceeded it by 0.085 (−0.180 to 0.346), so the three correlations could not be distinguished. For the eICU-CRD minus MIMIC-III difference in the correlation with time-dependent AUROC and in that with IPCW concordance, the unpaired bootstrap interval included zero and the paired interval excluded it, so the order of the cohorts could not be established (**Supplementary Methods S11**).

### The top-versus-bottom slope difference in the death sweep depends on which patients are removed

When patients were removed in a score-dependent manner (competing deaths were removed when the target death fraction was at or below the observed fraction, and target-event patients when it was above; removal probability proportional to the rank of the reference score, in either direction, with linear and cubic weights), the target-event share declined more slowly in the top-k stratum than in the bottom 90% stratum in 2, 1, 28, 28 of 28 model-outcome combinations (score-independent removal: 24 of 28). The first two counts are for removal weighted toward higher scores with linear and cubic weights, and the last two for removal weighted toward lower scores. With the evaluated model’s own score as the reference, the counts were 1 and 28 of 28. The mean slope difference was 0.13 under score-independent removal, −0.17 to −0.28 under removal weighted toward higher scores, and 0.49 to 0.67 under removal weighted toward lower scores (**Supplementary Table 16**).

### Observed risk in eICU-CRD differs between the binary and Aalen-Johansen definitions

Across 84 cohort-outcome-model units and 840 risk deciles, the sign of observed minus mean predicted risk differed between the binary proportion and the Aalen-Johansen cumulative incidence in 23 deciles in MIMIC-IV, 32 in MIMIC-III, and 97 in eICU-CRD (280 deciles per cohort). The Aalen-Johansen interval excluded the mean prediction in 0, 0, and 58 of these deciles (**Supplementary Table 17**). In the eICU-CRD top decile for any_48h, Aalen-Johansen observed risk minus the mean prediction was −0.64, −0.43, and −0.46 for LR, XGB, and LGBM and −0.08 to −0.01 for the four survival models. In three of eight eICU-CRD competing-risk model units (DeepHit for any_48h; Fine-Gray for 2p_48h and 2p_7d), the Aalen-Johansen interval of the top decile included the mean prediction, and BRII was 0.216, 0.045, and 0.053, against 0.921, 0.475, and 0.228 for the same models in MIMIC-III.

### Intervals that re-select the stratum are 0.92 to 1.13 times as wide as stratum-conditional intervals

A bootstrap that resampled patients and re-selected the top-10% stratum in each resample gave intervals 0.92 to 1.13 times as wide as the stratum-conditional bootstrap (n = 84 model-outcome-cohort units); 0 of 84,000 resamples (in 0 of 84 units) produced an empty denominator. The interval of XGB for 2p_48h in MIMIC-III included the Fine-Gray and Cox point estimates under both bootstraps, and the MIMIC-III and eICU-CRD intervals did not overlap for any of the 28 outcome-model pairs under either bootstrap (**Supplementary Table 18**). Values obtained by assigning all early-censored patients to no target event or to the target event are given in **Supplementary Table 19**, and half-widths by denominator size in **Supplementary Table 20**. With a censoring weight of zero, any_48h BRII was lower in eICU-CRD than in MIMIC-III in all seven models (**Supplementary Table 21**).

## Discussion

BRII is an algebraic function of PPV and the event-free observed survivor fraction of the flagged stratum. It gives one proportion: among flagged patients whose follow-up ended in a target event, a competing death, or early censoring, the share with the target event. Across three databases and seven models, BRII and PPV ordered at least one model pair differently in 34 of 48 cohort-outcome-top-k groups. For any AKI within 48 hours, BRII in the observed composition was high in MIMIC-III and low in eICU-CRD in all seven models, and early censoring was the largest of the three BRII components in eICU-CRD. Under simulated random censoring, BRII fell while binary-label AUROC changed by less than 0.02. In the death-contamination sweep, the target-event share declined more slowly in the top-k stratum than in the bottom stratum in 24 of 28 model-outcome combinations when patients were removed without regard to the model score; under score-dependent removal the count was 2 or 1 of 28 in one direction and 28 of 28 in the other. In eICU-CRD, the correlation of BRII with binary-label AUROC, time-dependent AUROC, and IPCW concordance was 0.30 to 0.39, and the three could not be distinguished.

Modeling competing risks did not by itself yield a high BRII: survival and competing-risk models also had low BRII under heavy competing-death burden or limited follow-up [7], and the ranking of models by BRII differed between validation cohorts. In MIMIC-III, XGB had a higher point estimate of BRII for 2p_48h than any cause-specific or competing-risk model, with a confidence interval that included the Fine-Gray and Cox estimates, but in eICU-CRD the gradient boosting classifiers had the lowest BRII for this target, and binary classifiers had more early censoring in their top-10% strata than cause-specific models. The 2p_48h ranking uses unit weights and rests on a small denominator mass. Features predictive of adverse outcomes may also be associated with early departure from observation.

AUROC and BRII address different questions: AUROC summarizes ranking across the whole population, whereas BRII summarizes the outcome composition of the flagged stratum. Under simulated random censoring for any_48h, BRII declined by up to 0.80 while binary-label AUROC shifted by less than 0.02, so binary-label AUROC did not show the change in the outcome composition of the flagged stratum [11, 25]. In eICU-CRD, the correlation of BRII with binary-label AUROC was 0.304; with time-dependent AUROC and IPCW concordance it was 0.352 and 0.389, and the three values could not be distinguished (**Supplementary Discussion**). PPV is computed within the flagged stratum, as BRII is, and AUPRC and F1 also depend on the predicted-positive set, which may explain why the four metrics together explained far more BRII variance in eICU-CRD than binary-label AUROC alone.

PPV counts patients who die and those who leave observation alike as non-events, and net benefit counts both in the false-positive term, so neither identifies which of the two lowers the value. The BRII decomposition separates this shortfall into competing-death and early-censoring components. These components point to possible contributors, including the model, the patient population, and the observation environment, but the contributors can interact, so the decomposition alone does not assign the shortfall to one of them. BRII and its decomposition could be logged at deployment and recomputed during monitoring as an algorithmovigilance signal (a measure used to monitor deployed algorithms for unintended effects), in line with the view that trust in ICU AI rests on reliability, safety, and auditability across the model lifecycle [41].

Existing competing-risk tools such as cause-specific concordance [18] and the competing-risk Brier score [19, 20], and recent evaluation guidance [4, 5, 35], assess discrimination or calibration across the full population [42, 43] but not the composition of the flagged stratum. BRII complements rather than replaces these: a model with adequate calibration and discrimination may still have a low BRII, and a high BRII does not guarantee calibration or predictive accuracy [38]. Cause-specific positive predictive values within marker-defined strata do report the target event and competing death as separate stratum-level probabilities [24]. BRII counts early-censored patients as an observed category of the flagged stratum instead of redistributing them under an independent-censoring assumption. Within-stratum Aalen-Johansen cumulative incidences of the target event and competing death, and their ratio, are reported alongside BRII in **Supplementary Table 14**. The ratio assumes independent censoring, which may not hold for ICU discharge. We recommend reporting BRII alongside PPV and evidence of intervention efficacy, because BRII excludes event-free observed survivors and can remain high when they dominate the stratum. Four recent preprints evaluate predictions over all cases in the evaluation data or summarize the event burden of a whole group (**Supplementary Discussion**).

Other reported measures are functions of quantities that are also reported separately. For two risk categories, the event and non-event components of the net reclassification index equal the increase in true-positive rate and the decrease in false-positive rate, and the weighted index equals the change in net benefit [44]. That review recommends reporting the two components separately and keeping the existing descriptive terms. We report BRII because it combines PPV and the event-free observed survivor fraction into one proportion, the share of target events among flagged patients other than event-free observed survivors. The components reported beside this proportion show which outcomes lower it. Tables 2 and 3 report PPV, the percentage of event-free observed survivors, and the three other component percentages alongside BRII.

Table 2 also reports net benefit, which is derived from the relation in which the threshold probability sets the relative harm of false-positive and false-negative results [22]. BRII is not derived from a decision analysis.

Clinical reachability, whether a flagged patient can still receive and benefit from an intervention, requires that the target event can still occur while the patient remains in the alerting setting, and the competing-death and early-censoring components provide information on this precondition. Reachability further requires that the alert arrives in time, that a clinician can act, and that an efficacious intervention exists, and BRII does not measure these conditions.

When competing death is the dominant component, the model may be flagging broad acute risk rather than the target event, and a two-gate approach could screen for short-term mortality before issuing the target-event alert, using existing estimates without a new model. When early censoring dominates and observation ends through transfer or the end of data collection rather than discharge after recovery, one possible response lies in the observation environment, for example restricting deployment to units where patients remain observed through the horizon [40]. Neither response was tested in this study.

Because BRII is defined over the flagged stratum of any model, it is computed in the same way for binary, cause-specific, and competing-risk models, as PPV at a top-k cut-off is. The same construction can be applied to other alert strata with competing death and early censoring, such as those for sepsis, delirium, and respiratory failure. Guidance on performance measures for survival outcomes is written for Cox proportional hazards models [39], and guidance on validation of competing-risk models is given separately [38].

BRII counts early-censored patients in its denominator, which makes it an operational metric rather than an event-probability summary. Because BRII does not count event-free observed survivors, a model that flags patients likely to remain event-free and observed can have a higher BRII than one that ranks patients by target-event risk. The evaluation covers one worked case, AKI in North American ICU cohorts with death as the principal competing event, and the behavior of BRII in non-ICU, non-US, and structurally different settings remains to be established [45]. The unit weights can be read as implicit costs and were not derived from a decision analysis. BRII rankings depended on the weights for all targets except any_48h. Calibration was assessed in risk deciles only, and its direction in eICU-CRD depended on how censored patients were counted.

The BRII increase after removal of early-censored patients mostly restates their share of the denominator and does not estimate BRII under complete follow-up. Binary-label AUROC counts early-censored patients as non-events, so it is not an estimate of discrimination under complete follow-up; time-dependent AUROC and IPCW concordance, which treat early censoring and competing death as censoring and target a cause-specific quantity (**Supplementary Methods S11**), are reported in **Supplementary Table 15**. We did not distinguish reasons for early censoring (discharge after recovery, transfer, or end of data collection), assess sex or age subgroups, or run an outcome-model-based simulation. Because the targets are nested, the sweep scenarios represent variation across conditions rather than independent observations.

Priority extensions include generalizing the denominator to multiple competing pathways and the numerator to composite targets, whose interpretability and ranking stability remain to be established [46]. A further extension is a prospective study of the effect of BRII-informed alert composition on clinician decisions, as reported for prognostic tools in colorectal liver metastases [47] and AI documentation tools in primary care [48]. Validation targets include settings with competing-event structures that differ from AKI, such as cardiovascular or oncology cohorts and non-ICU wards. We did not examine BRII as a training objective (**Supplementary Discussion**).

## Conclusions

The Balanced Risk Integrity Index (BRII) provides a standardized, stratum-level assessment of target-event occurrence relative to competing deaths and early censoring among patients flagged by clinical prediction models. Across three intensive care databases, BRII and PPV yielded different model rankings in 34 of 48 cohort-outcome-top-k groups (138 of 1,008 model pairs). For 48-hour AKI prediction in the top 10% risk stratum, BRII ranged from 0.90 to 0.96 in MIMIC-III but remained below 0.26 across all seven models in eICU-CRD, where early censoring was the largest component. Under simulated random censoring, BRII decreased by up to 0.80 while binary-label AUROC changed by less than 0.02. Although algebraically related to PPV, BRII and its component decomposition provide complementary information for comparing prediction alerts across models and observation environments. Its clinical decision-making utility remains to be established prospectively.

## Data Availability

MIMIC-IV, MIMIC-III, and eICU-CRD are publicly available through PhysioNet (https://physionet.org). Source data for all figures and tables are provided with this paper.

## List of abbreviations

AKI: Acute kidney injury;
AUPRC: Area under the precision-recall curve;
AUROC: Area under the receiver operating characteristic curve;
BRII: Balanced Risk Integrity Index;
CIF: Cumulative incidence function;
CS: Cause-specific;
CS_CoxPH: Cause-specific Cox proportional hazards;
CS_RSF: Cause-specific random survival forest;
eICU-CRD: eICU Collaborative Research Database;
ICU: Intensive care unit;
IPCW: Inverse-probability-of-censoring-weighted;
KDIGO: Kidney Disease: Improving Global Outcomes;
LGBM: LightGBM;
LR: Logistic regression;
MIMIC: Medical Information Mart for Intensive Care;
PPV: Positive predictive value;
XGB: XGBoost.

## Declarations

### Ethics approval and consent to participate

All three databases (MIMIC-IV, MIMIC-III, and eICU-CRD) contain de-identified data, and their use is exempt from institutional review board approval under 45 CFR 46.101(b)(4). Access was obtained through PhysioNet Credentialed Health Data Use Agreements. Consent to participate is not applicable, as the study used previously collected de-identified data.

### Consent for publication

Not applicable.

### Availability of data and materials

MIMIC-IV, MIMIC-III, and eICU-CRD are publicly available through PhysioNet (https://physionet.org). The source data underlying all figures and tables are provided with this article. Code for all analyses is available at https://github.com/taeyeong1209/BRII; the repository will be made publicly available upon acceptance. For the review process, editors and reviewers can temporarily access the materials via https://gitfront.io/r/InyongJeong/NFTJBNEsBTga/BRII/.

### Authors’ contributions

I.J. and T.L. contributed equally to this work. I.J. conceived the study, developed the BRII framework, designed and conducted all analyses, and wrote the original manuscript. T.L. contributed to study design, conducted analyses, and contributed to manuscript writing. J.P. contributed to data extraction and manuscript revision. Y.K. contributed to data curation and validation. H.L. supervised the study, contributed to study design and interpretation, and critically revised the manuscript. All authors read and approved the final manuscript.

## Supplementary Methods

### S1. Baseline kidney function and AKI definitions

Baseline serum creatinine was defined as the minimum value recorded within the first 24 hours after admission. AKI was defined according to KDIGO serum creatinine criteria. Stage ≥ 1 AKI required either an increase of at least 0.3 mg/dL within any 48-hour window or an increase to at least 1.5 times baseline within 7 days. Stage ≥ 2 AKI required an increase to at least 2.0 times baseline. Urine output criteria were not used.

We evaluated four targets: any AKI within 48 hours (any_48h), any AKI within 168 hours (any_7d), stage ≥ 2 AKI within 48 hours (2p_48h), and stage ≥ 2 AKI within 168 hours (2p_7d). Follow-up was limited to the prediction horizon of each target (at most 168 hours from the prediction anchor). Death before the target event was treated as a negative label in binary models, as a censoring event in cause-specific models, and as a competing event in competing-risk models.

To characterize the competing-risk profile of each cohort prior to model evaluation, we also computed cohort-level three-cause cumulative incidence functions (target AKI, competing death, and early censoring, with event-free survival as the residual) at each horizon using a three-cause Aalen-Johansen estimator. Because early censoring is treated as a cause and no other observation is censored before the horizon, these estimates equal the observed proportions of the three outcomes.

### S2. Features and preprocessing

We extracted 42 features from the first 24 hours after ICU admission using harmonized variable definitions across databases. These included demographics, baseline creatinine, vital signs, anthropometrics, Glasgow Coma Scale components, laboratory values, and the blood urea nitrogen-to-creatinine ratio. For time-varying measurements, summary statistics (minimum, maximum, and last observed values) were used where applicable. Missing values were imputed using fold-specific training-set medians.

### S3. Model hyperparameters

Seven models spanning the principal paradigms used in AKI prediction were trained in MIMIC-IV using five-fold stratified cross-validation. Binary classifiers comprised logistic regression (balanced class weights), XGBoost, and LightGBM (class imbalance weighting adjusted via the positive-class scale parameter). Cause-specific survival models comprised Cox proportional hazards regression (ridge penalty α = 10^-4^) and random survival forest (maximum tree depth = 10). Competing-risk models comprised Fine-Gray subdistribution hazard and DeepHit (64 hidden units, 80 time bins, 60 epochs, learning rate = 10^-4^). DeepHit hyperparameters were adopted from the original publication with minor adaptation for feature dimensionality. The seven models provide a diverse testbed for evaluating BRII across modeling paradigms rather than as individually optimized or calibrated clinical tools.

### S4. Closed-form relationship between BRII and PPV

Let *N_k_* denote the number of patients in the top-*k* stratum, and let *H* denote the prediction horizon. Within this stratum, let *nA*, *nD*, and *nC* denote, respectively, the counts of patients who experienced the target event by *H*, died of a competing cause before the target event by *H*, and were censored before *H* without either event; the remaining *nS* = *N_k_* − *nA* − *nD* − *nC* are event-free observed survivors, that is, patients without either event who were observed through *H*. Define *A* = *nA*/*N_k_*, *D* = *nD*/*N_k_* and *C* = *nC*/*N_k_*. BRII was then defined as:

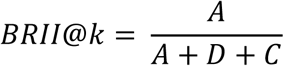

This equals the proportion of target events among patients whose outcome was resolved or whose observation ended by H; event-free observed survivors (*nS*/*N_k_*) contribute to neither the numerator nor the denominator. Relative to PPV, BRII can be written in closed form as *BRII*@*k* = *PPV*/(1 − *nS*/*N_k_*), because *PPV* = *nA*/*N_k_* and 1 − *nS*/*N_k_* = (*nA* + *nD* + *nC*)/*N_k_*. BRII is therefore an algebraic function of PPV and the event-free observed survivor fraction: the two coincide only when *nS* = 0, and the factor 1 − *nS*/*N_k_* is the quantity that PPV does not show. Early censoring is a component of the denominator and is not a separate input. Given PPV and the survivor fraction, BRII adds no information. We report BRII as one proportion together with PPV and the four component proportions.

### S5. Exploratory BRII zones

To facilitate cross-model and cross-cohort interpretation, we grouped top-10% BRII values into four exploratory BRII zones: Zone A (BRII ≥ 0.80, high), Zone B (0.50 to 0.80, moderate), Zone C (0.30 to 0.50, limited), and Zone D (below 0.30, minimal). Zone boundaries were set as preliminary anchors derived from the empirical distribution in this study rather than from external clinical validation; they are intended to support comparative interpretation rather than to prescribe deployment decisions.

### S6. Death-contamination sweep

We varied the target death fraction from 0.0-0.9 in increments of 0.1 using two-sided resampling. When the target death fraction was below the observed value, death events were downsampled while preserving AKI events and censored observations; when above, AKI events were downsampled. Within the downsampled event type, patients were removed at random, without regard to the model score. This preserved the original feature-prediction relationship among retained patients. Predictions were not re-fitted; original out-of-fold predictions were retained and the top-k stratum was redefined within each resampled dataset.

Resampling was repeated 200 times per condition, selected on the basis of pilot analyses confirming that 95% confidence interval widths stabilized within 0.01 by 150 iterations. Across four outcomes, seven models, 10 death-fraction levels, 200 repeats, and four top-k thresholds (5%, 10%, 15%, 20%), this yielded 224,000 scenarios.

We computed *A*/(*A* + *D*) within both the top-k and complementary bottom strata at each death-fraction level and compared the slope of this ratio across strata. From the resulting mean BRII trajectories across death fractions, we identified for each model-outcome combination at the 10% top-k threshold the death fraction at which BRII first fell below each of five anchor values (0.80, 0.70, 0.50, 0.30, 0.20), using linear interpolation between adjacent death-fraction grid points.

### S7. Censoring-sensitivity sweep

We examined the effect of incomplete follow-up under three censoring mechanisms: random, healthy-biased informative, and sick-biased informative. Censoring rates, defined as per-patient draw probabilities, ranged from 0.0-0.9 in increments of 0.1. Under random censoring, each patient was independently assigned a censoring draw probability equal to the target rate. Under informative censoring, lower-risk (healthy-biased) or higher-risk (sick-biased) patients were preferentially censored, using LightGBM prediction scores as the bias signal. Censoring was applied only when the drawn censoring time preceded the patient’s observed event or follow-up time, so the effective censored fraction was lower than the draw probability and varied with the underlying event-time distribution.

Because LightGBM is one of the seven models under evaluation, this design introduces potential circularity. To assess the magnitude of this concern, we repeated the informative-censoring sweep using Cox proportional hazards predictions as an alternative bias signal. Censoring times were drawn from a Beta(1, 3) distribution scaled to the prediction horizon, a right-skewed distribution with mean at approximately 0.25*H*. This choice stress-tests BRII under an early-censoring regime representative of ICU settings where patients frequently leave observation within the first quarter of the horizon. Across three censoring mechanisms, ten censoring rates, seven models, four outcomes, and 200 repetitions at the top-10% threshold, this yielded 168,000 scenarios. AUROC in the sweep was computed in the whole cohort against binary labels, with target events that fell after the drawn censoring time relabeled as 0.

### S8. Observation-window reference ratios

We compared the BRII fold change after removal of early-censored patients with two reference ratios. The first is the ratio of subcohort to full-cohort AKI incidence. The second is (A + D + C)/(A + D) in the original top-10% stratum, computed from the stratum counts. This second ratio is the fold change obtained when early-censored patients are removed from the BRII denominator and the stratum is not re-identified. Across the 28 model-outcome combinations, the observed fold change was 0.97 to 1.12 times the second ratio (0.99 to 1.03 times for any_48h). For 2p_7d it was 1.08 to 1.12 times the second ratio in four of seven models, which reflects re-identification of the stratum (Supplementary Table 7).

### S9. Complementary analyses: regression, net benefit, and weight sensitivity

To describe how much of the variation in BRII is explained by standard metrics, we regressed BRII on AUROC, area under the precision-recall curve (AUPRC), PPV, and F1 score, individually and in combination, at the 10% top-*k* threshold across all cohort-model-outcome conditions (n = 84), by cohort (n = 28 each), and by outcome (n = 21 each), and report adjusted R² values. To position BRII relative to decision-analytic metrics, we computed net benefit under two specifications using the standard formulation *NB* = *TP*/*N* − (*FP*/*N*) × [*p_t_*/(1 − *p_t_*)]. In the fixed-threshold specification, patients in the top 10% of predicted scores were flagged (*k* = 0.10) and the threshold probability *p_t_* = 0.10 was applied. In the prevalence-matched specification, patients in the top *π* fraction (where *π* is cohort prevalence of the target event) were flagged and *p_t_* = *π* was applied.

We examined BRII decomposition within matched net-benefit bins across death-contamination levels using the prevalence-matched specification, because this places models on a shared clinical threshold and is the specification reported in sensitivity analyses. For each outcome, net-benefit values of the seven models across the death-contamination sweep (200 repetitions per death-fraction level) were grouped into 20 quantile bins. A bin was counted as divergent when the 2.5th to 97.5th percentile range of BRII within the bin exceeded 0.05 or that of the death share exceeded 0.10. To assess sensitivity of BRII rankings to weight specification, we computed a weighted variant BRII_*w* = *A*/(*A* + *w_D_* · *D* + *w_C_* · *C*) across *w_D_* ∈ {0.5, 0.75, 1.0, 1.25, 1.5} and *w_C_* ∈ {0.25, 0.5, 0.75, 1.0} at the top-10% threshold. Model rankings under each weight combination were compared against the unit-weight specification using Spearman rank correlation, at five target death fractions (0.00, 0.10, 0.20, 0.30, and 0.50) of the death-contamination sweep. In the pooled analysis across all conditions, the adjusted R² was 79.7% for the four metrics together and 45.7% for AUROC alone; cohort-specific and outcome-specific values are reported in the main text.

### S10. Rank comparisons and within-stratum Aalen-Johansen cumulative incidence

For each cohort, outcome, and top-k level (1%, 2%, 5%, and 10%; 48 groups), the seven models give 21 model pairs. A pair was counted as discordant between BRII and PPV when the two measures ordered the two models in opposite directions, and as tied when either measure was equal for the two models to three decimals (1,008 pairs; Supplementary Table 13). Within each stratum we also estimated the Aalen-Johansen cumulative incidence of the target event (F1) and of competing death (F2) at the horizon, with early-censored patients treated as censored, and the ratio R = F1/(F1 + F2). Pairs were classified in the same way for BRII and R, using unrounded values (Supplementary Table 14). For each outcome, model, and top-k level (112 combinations), we computed the difference between eICU-CRD and MIMIC-III in BRII, the difference in R, and the difference between these two differences, with 95% intervals from 1,000 within-stratum bootstrap resamples drawn independently in the two cohorts. The Aalen-Johansen estimator assumes independent censoring, which may not hold for ICU discharge.

### S11. Time-dependent AUROC, IPCW concordance, and IPCW Brier score

Time-dependent AUROC (cumulative/dynamic definition) at half the horizon, IPCW concordance truncated at the horizon, and the IPCW Brier score at 0.99 times the horizon were computed for the 84 cohort-outcome-model units. Competing death and early censoring were both treated as censoring, and the censoring distribution was estimated with the Kaplan-Meier method. In the primary version the inverse-probability weights for MIMIC-III and eICU-CRD came from each cohort’s own censoring distribution. A version with weights from the MIMIC-IV censoring distribution is reported for comparison. The three estimators assume that early censoring and competing death are independent of the time to the target event; treating competing death as censoring targets a cause-specific quantity, which differs from the observed composition that BRII reports.

Within each cohort (n = 28 outcome-model combinations), each measure was correlated with top-10% BRII using Spearman correlation. Differences between correlations are reported with 95% intervals from 1,000 bootstrap resamples of the 28 combinations. For the difference in a correlation between eICU-CRD and MIMIC-III, the 28 combinations were resampled independently in each cohort (unpaired) and as the same combinations in both cohorts (paired). Before the results were available, we specified that the order of the two cohorts would be stated only when both intervals excluded zero. The combinations share models across outcomes, so the intervals are descriptive (Supplementary Table 15).

### S12. Score-dependent removal in the death-contamination sweep

The death-contamination sweep of S6 removes patients at random within an event type. We repeated it in MIMIC-IV at the top-10% threshold (200 repetitions) with removal probability proportional to the rank of a reference score. Competing deaths were removed when the target death fraction was at or below the observed fraction, and target-event patients when it was above, and the number removed at each level was the same as in S6. Removal was weighted toward higher scores (weight r, the within-cohort rank of the score scaled to 0 to 1) or toward lower scores (weight 1 − r). The reference score was the LightGBM prediction. Two sensitivity settings used cubic weights (r³ or (1 − r)³) or the evaluated model’s own score as the reference. For each setting we report the number of the 28 model-outcome combinations in which the slope of A/(A + D) was less steep in the top-10% stratum than in the bottom 90% stratum (Supplementary Table 16). Score-dependent removal is one mechanism and does not represent all changes in patient composition.

### S13. Decile calibration with binary and Aalen-Johansen observed risk

Calibration was assessed for the 84 cohort-outcome-model units; all predictions were within 0 to 1, so no unit was excluded. Patients were ranked by predicted risk. The top decile was the top-10% stratum, and the remaining patients were divided into nine groups of equal size. Observed risk in each decile was computed in two ways: the binary proportion, in which early-censored patients and competing deaths count as no event, and the Aalen-Johansen cumulative incidence at the horizon, in which death is a competing event and early-censored patients are censored. The 95% interval of the Aalen-Johansen estimate came from 1,000 within-decile bootstrap resamples. Unit-level summaries were the decile-size-weighted mean absolute difference between observed and mean predicted risk (weighted MAE), the observed minus expected event proportion, and the calibration slope and intercept against binary labels. Models were not recalibrated. Predictions of binary classifiers are probabilities of the binary label, those of cause-specific models ignore the competing risk, and those of competing-risk models are cumulative incidences, so the three are not on the same scale (Supplementary Table 17).

### S14. Stratum-selection bootstrap, values under extreme assignments of early-censored patients, and sampling uncertainty

The primary 95% intervals resample patients within the given top-10% stratum. As a sensitivity analysis, for each of the 84 units we resampled all patients with replacement 1,000 times, re-selected the top-10% stratum from the fixed predictions in each resample, and recomputed BRII (Supplementary Table 18). Neither bootstrap includes the uncertainty of refitting the model. For each external top-10% stratum we also report A/(A + D + C), A/(A + D), and (A + C)/(A + D + C); the first and third are the values obtained by assigning all early-censored patients to no target event or to the target event (Supplementary Table 19). With m patients in A + D + C, the normal-approximation half-width of a 95% interval for BRII is 1.96 × √(BRII(1 − BRII)/m) (Supplementary Table 20). Weighted BRII, A/(A + wD·D + wC·C) with wD = 1, was additionally computed for the external any_48h top-10% strata with wC from 0 to 1 (Supplementary Table 21).

### Supplementary Discussion. BRII in relation to AUROC and existing stratum-level metrics

AUROC and BRII address different questions: ranking discrimination across the full population versus the outcome composition of the flagged stratum. Under simulated random censoring for any_48h, BRII declined by up to 0.80 while binary-label AUROC shifted by less than 0.02, and in external validation the correlation between BRII and binary-label AUROC was *ρ* = 0.705 in MIMIC-III and *ρ* = 0.304 in eICU-CRD at the top-10% threshold. Under random censoring, binary-label AUROC did not show the change in the outcome composition of the flagged stratum.

AUROC in the primary analysis was computed against binary labels across all model families, to enable uniform comparison across binary, cause-specific, and competing-risk paradigms and to reflect the binary ascertainment that typically governs deployed alert systems. By definition, BRII does not change if censored patients are reweighted to obtain an unbiased discrimination estimate. With weights from each cohort’s own censoring distribution, the correlation of BRII with time-dependent AUROC and with IPCW concordance in eICU-CRD was 0.352 and 0.389, against 0.304 for binary-label AUROC, and the three values could not be distinguished (Supplementary Table 15). BRII describes the outcome composition of the flagged stratum through *H*; this is a different question from population-level ranking accuracy.

Reporting competing death and early censoring separately distinguishes BRII from PPV and net benefit. PPV and net benefit quantify the overall value of the flagged population, but neither separates competing death from early censoring. In binary PPV, patients who die and patients who leave observation are both counted as non-events; net benefit aggregates competing death and censoring contributions into the false-positive term, so a low net benefit cannot distinguish whether the flagged stratum is dominated by patients who die before the target event or by those who leave observation. The decomposition separates this shortfall into competing-death and early-censoring components, which indicates possible contributors when PPV or net benefit is low, including the model, the patient population, and the observation environment. Because these contributors can interact, the decomposition alone does not assign the shortfall to one of them. This aligns with the view that trust in ICU AI rests not on explainability alone but on reliability, safety, and auditability assessed across the model lifecycle; BRII contributes a reportable, auditable measure of one such property.

Existing competing-risk evaluation tools, including cause-specific concordance and the competing-risk Brier score, assess discrimination or calibration across the full population. These measures indicate whether a model ranks or predicts accurately on average, but do not evaluate the composition of the flagged stratum. Cause-specific positive predictive values within marker-defined strata report the target event and competing death as separate probabilities [1] and do not include early censoring as a component. Recent guidance has systematized the evaluation of clinical prediction models through discrimination, calibration, and clinical utility assessments, but does not address outcome composition within the flagged stratum in the presence of competing events. BRII extends this evaluation to the outcome composition of the flagged stratum, including the proportions of flagged patients who die or leave observation before the prediction horizon, complementing rather than replacing population-level measures. A model with adequate calibration and discrimination may still have a low BRII; conversely, high BRII does not guarantee calibration or overall predictive accuracy.

### Recent preprints on censoring and competing risks

Four recent arXiv reports address censoring or competing risks in time-to-event data. Two evaluate predictions under censoring. A copula-based dependent Brier score and its integrated version evaluate single-event survival models over all patients under an assumed Archimedean copula, a model of the dependence between the censoring time and the event time [2]. Threshold-weighted continuous ranked probability scores (a weighted integral of the Brier score over time thresholds) and logarithmic scores evaluate predictive distributions when realized event times or forecasts are known only through values right-censored at a common fixed horizon [3]. That work does not consider random censoring.

The other two address competing risks. The average cause-specific hazard is the cause-specific cumulative incidence divided by the restricted mean event-free time, the average time free of any event up to a horizon. This hazard is a rate-scale summary of a group whose population definition does not involve the censoring distribution [4]. Its estimator assumes independent right-censoring. A benchmarking framework for competing-risk models reports, among other metrics, calibration, time-dependent AUROC, the Brier score, and net benefit [5]. Each of the four reports evaluates predictions over all cases in the evaluation data or summarizes the event burden of a whole group.

### Number needed to evaluate and training-objective use

The number needed to evaluate for an early warning score is the number of patients evaluated per detected outcome, and a PPV of 20% corresponds to a value of 5 [6].

A differentiable approximation of the Wilcoxon-Mann-Whitney statistic, whose normalized form equals the AUROC, has been used as a training objective [7]. We did not examine BRII as a training objective; such use would require a differentiable surrogate for the rank-based top-k selection, a constraint on the event-free observed survivor fraction or joint use with PPV, and separate reporting of calibration and net benefit, and we make no claim that it would improve a model.

### Supplementary Tables

**Supplementary Table 1.**
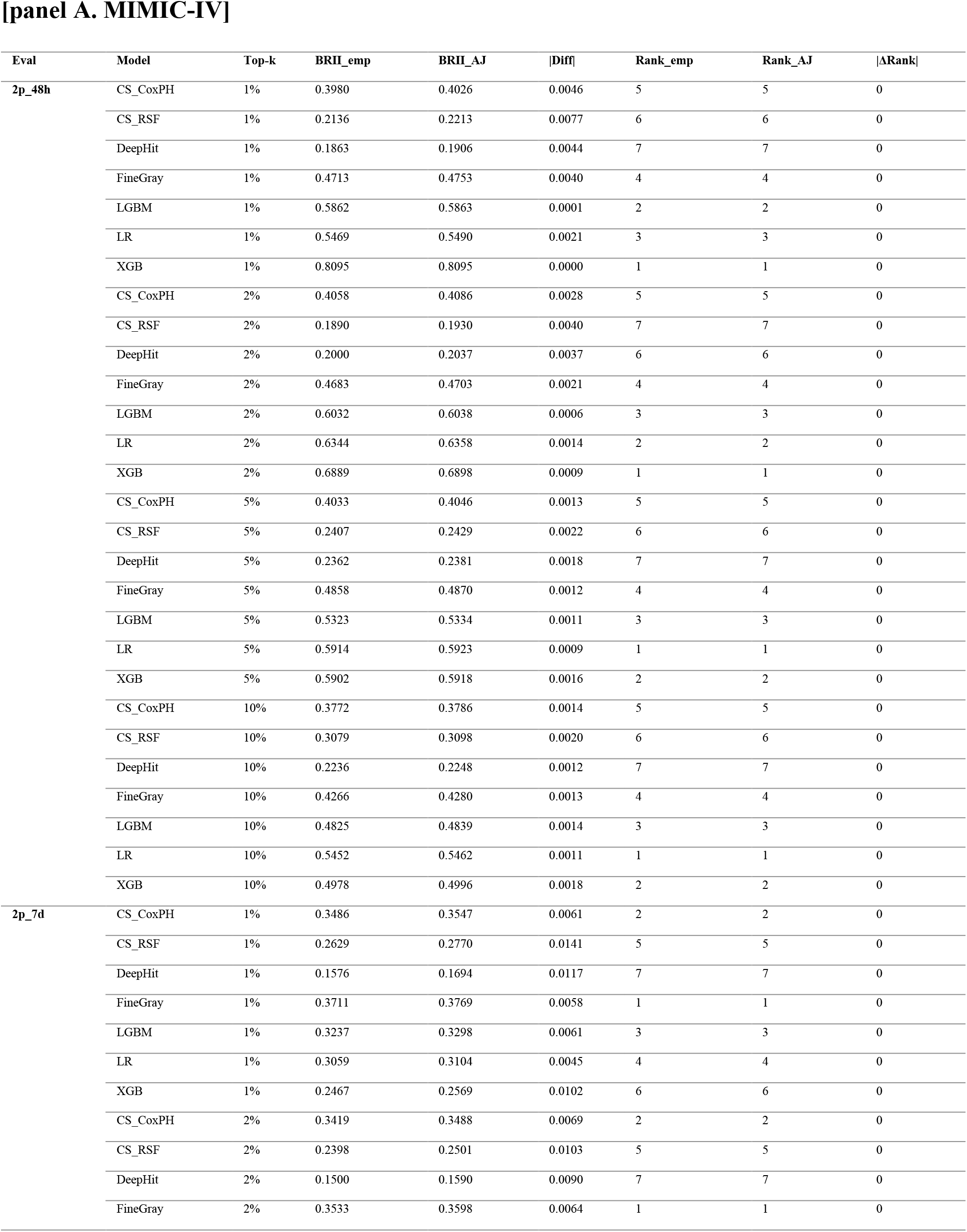
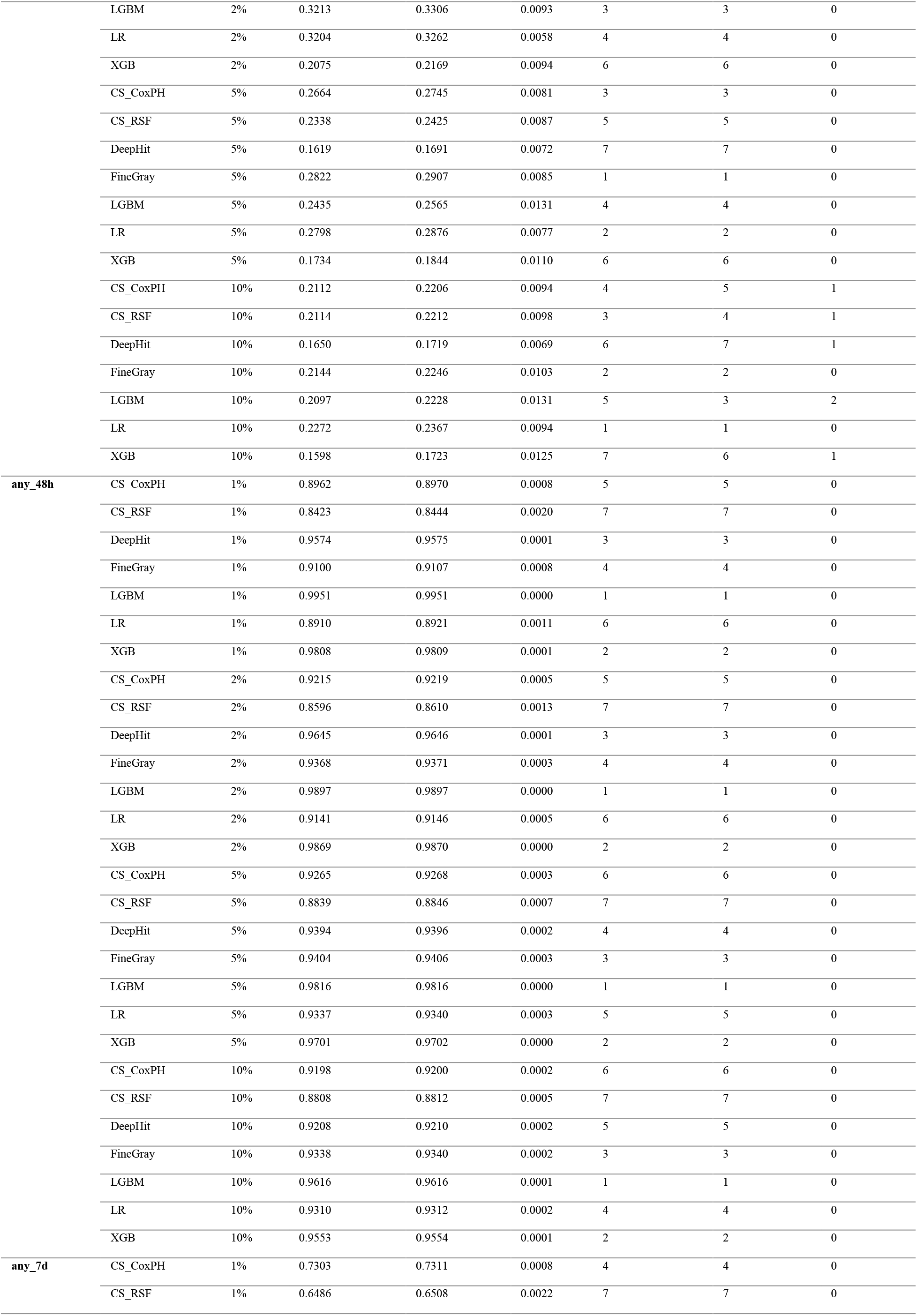
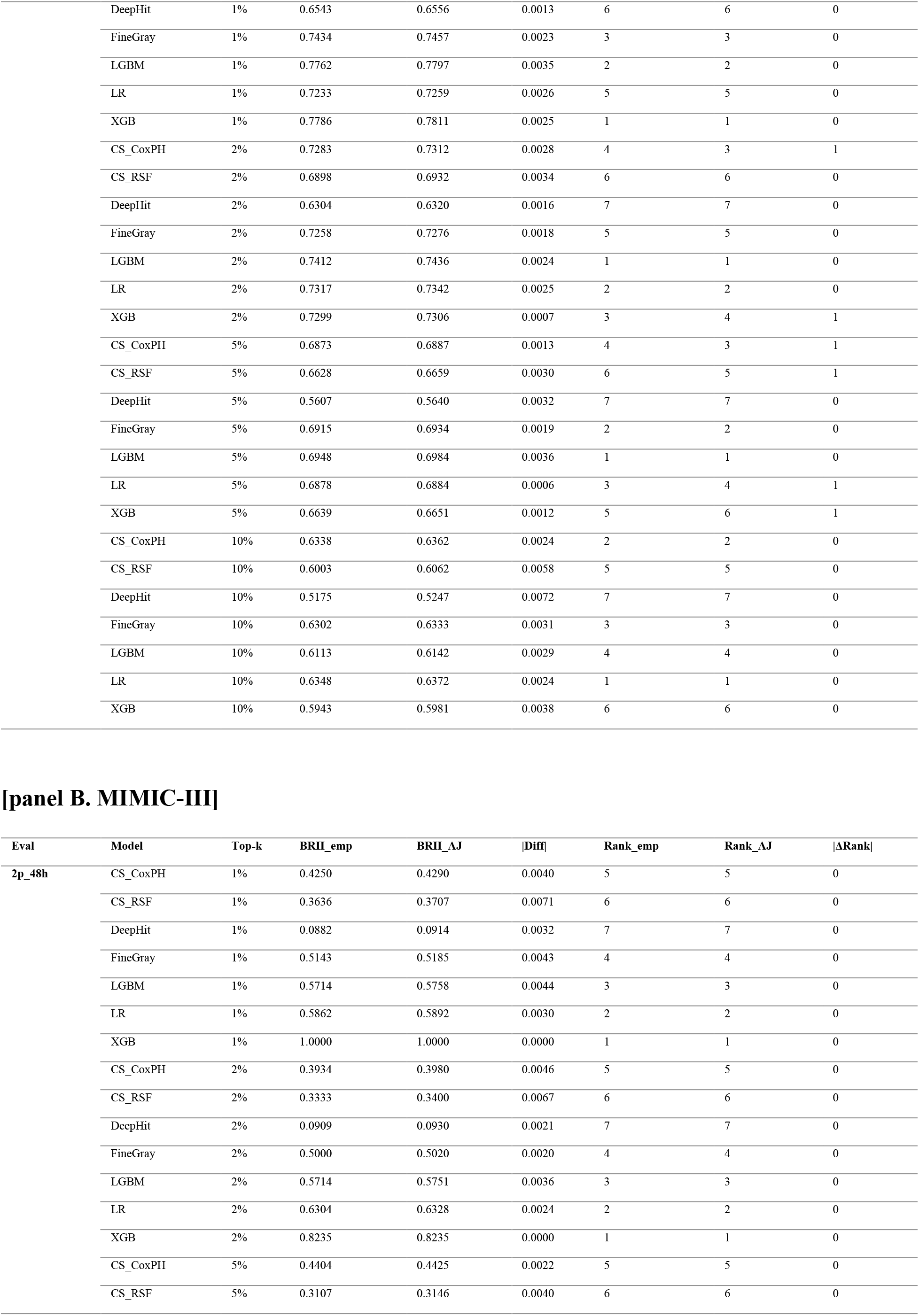
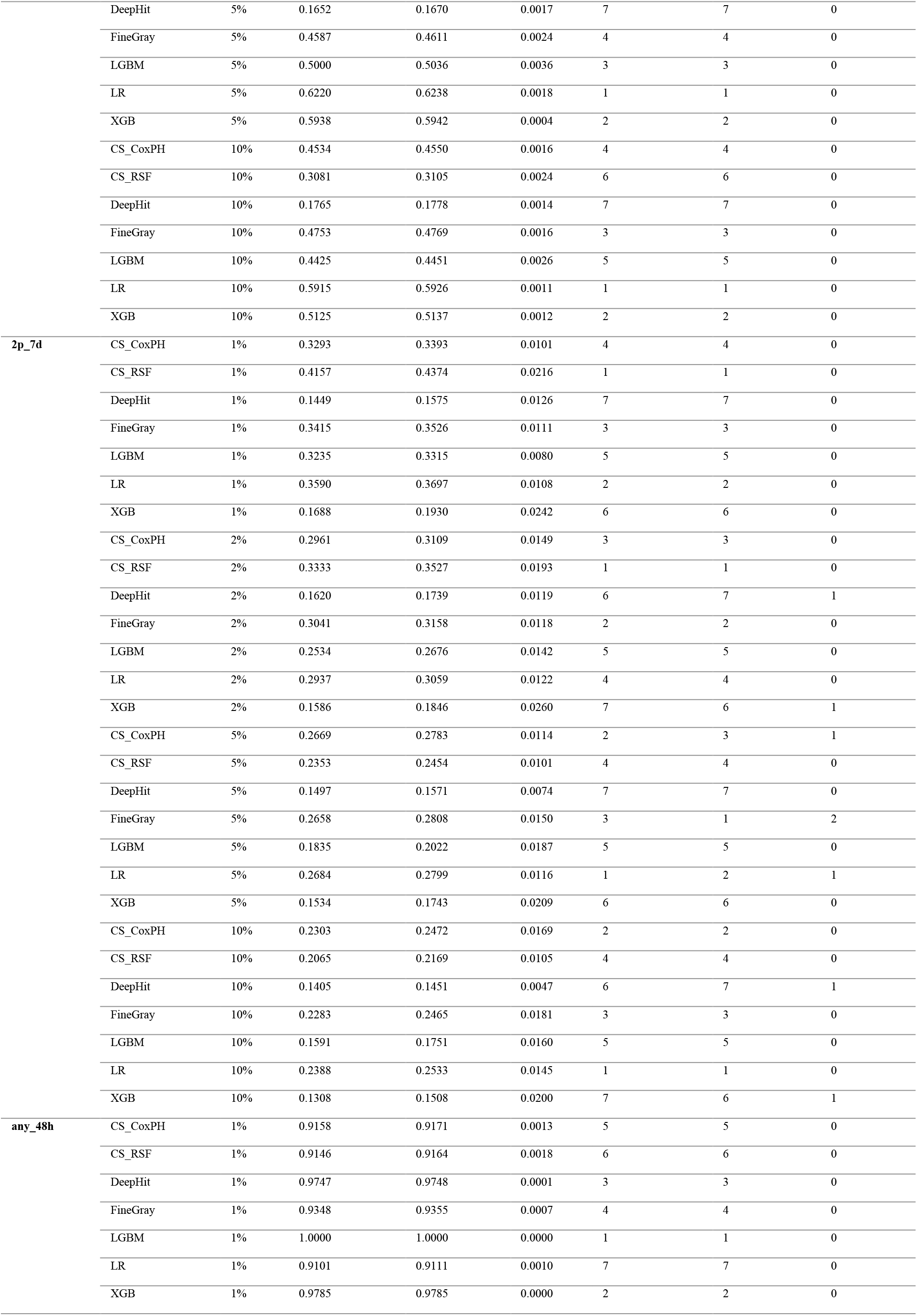
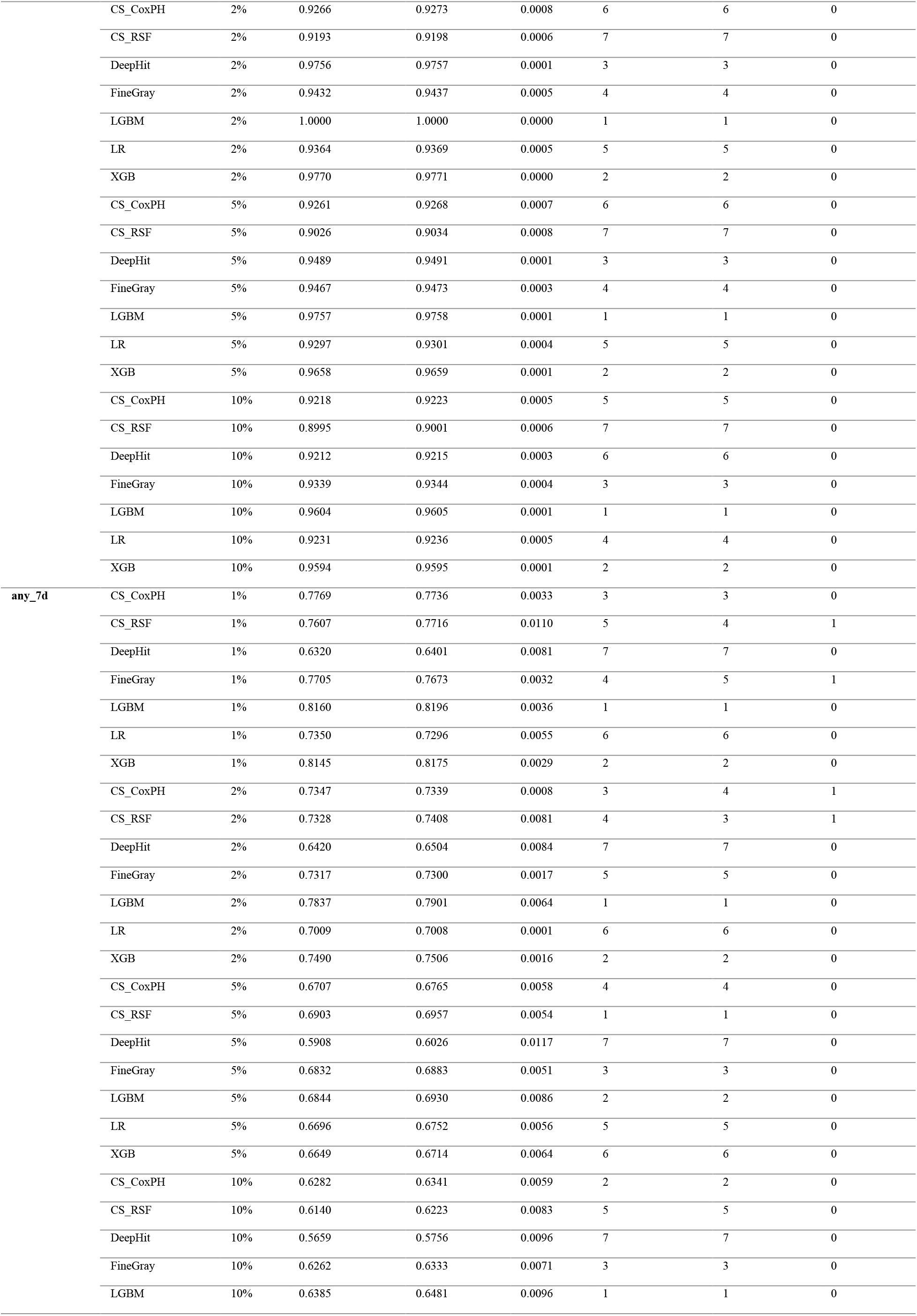
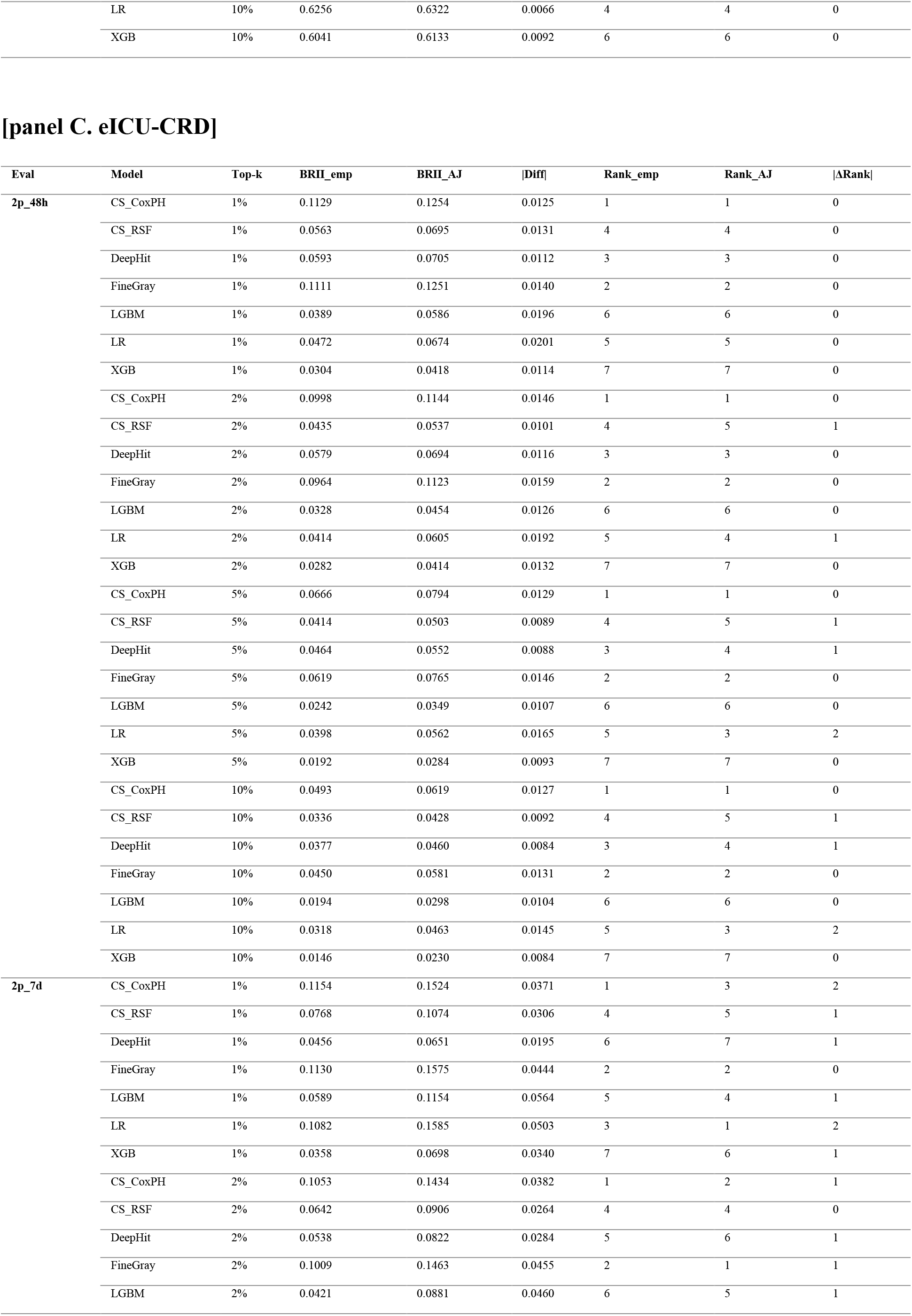
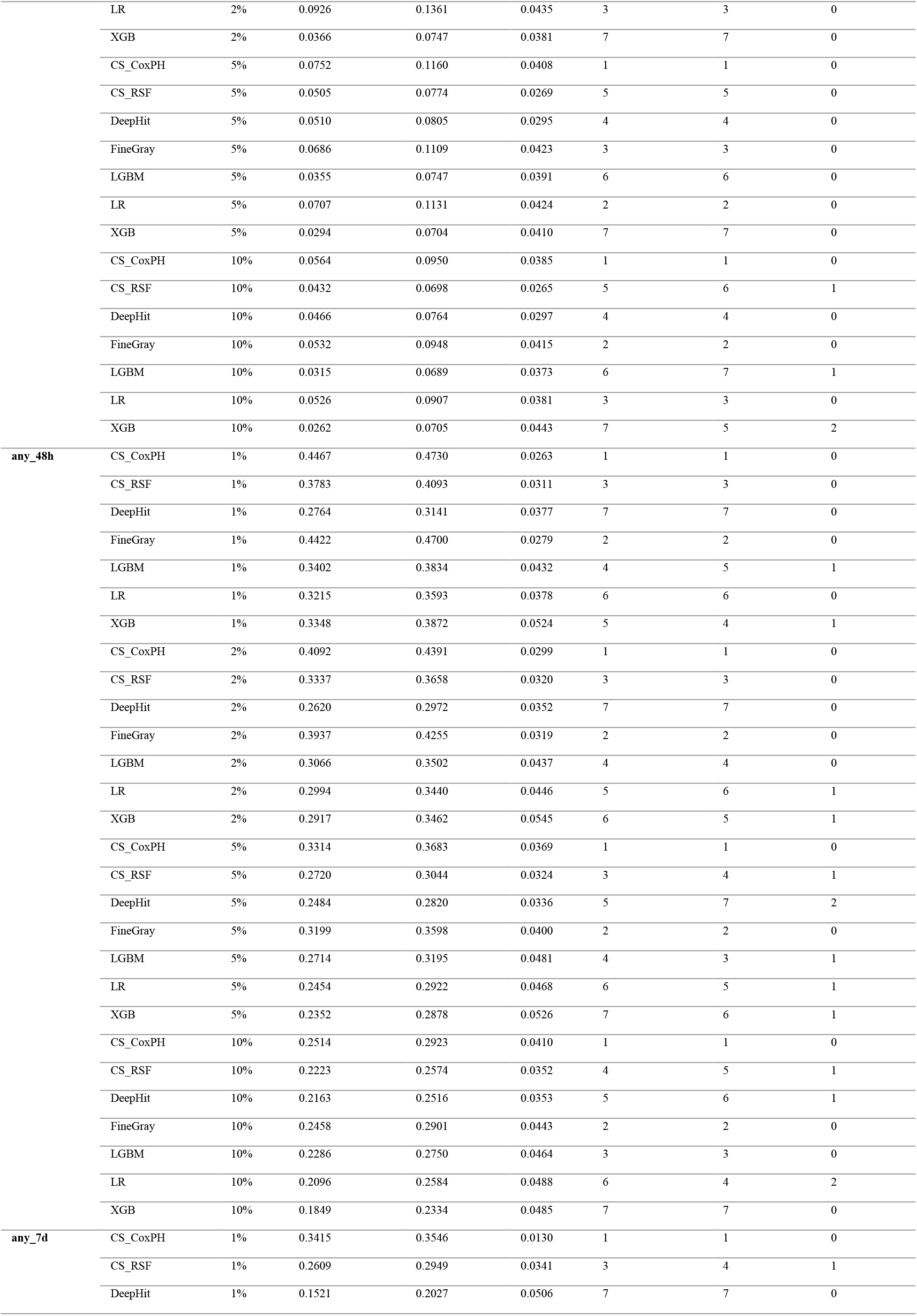
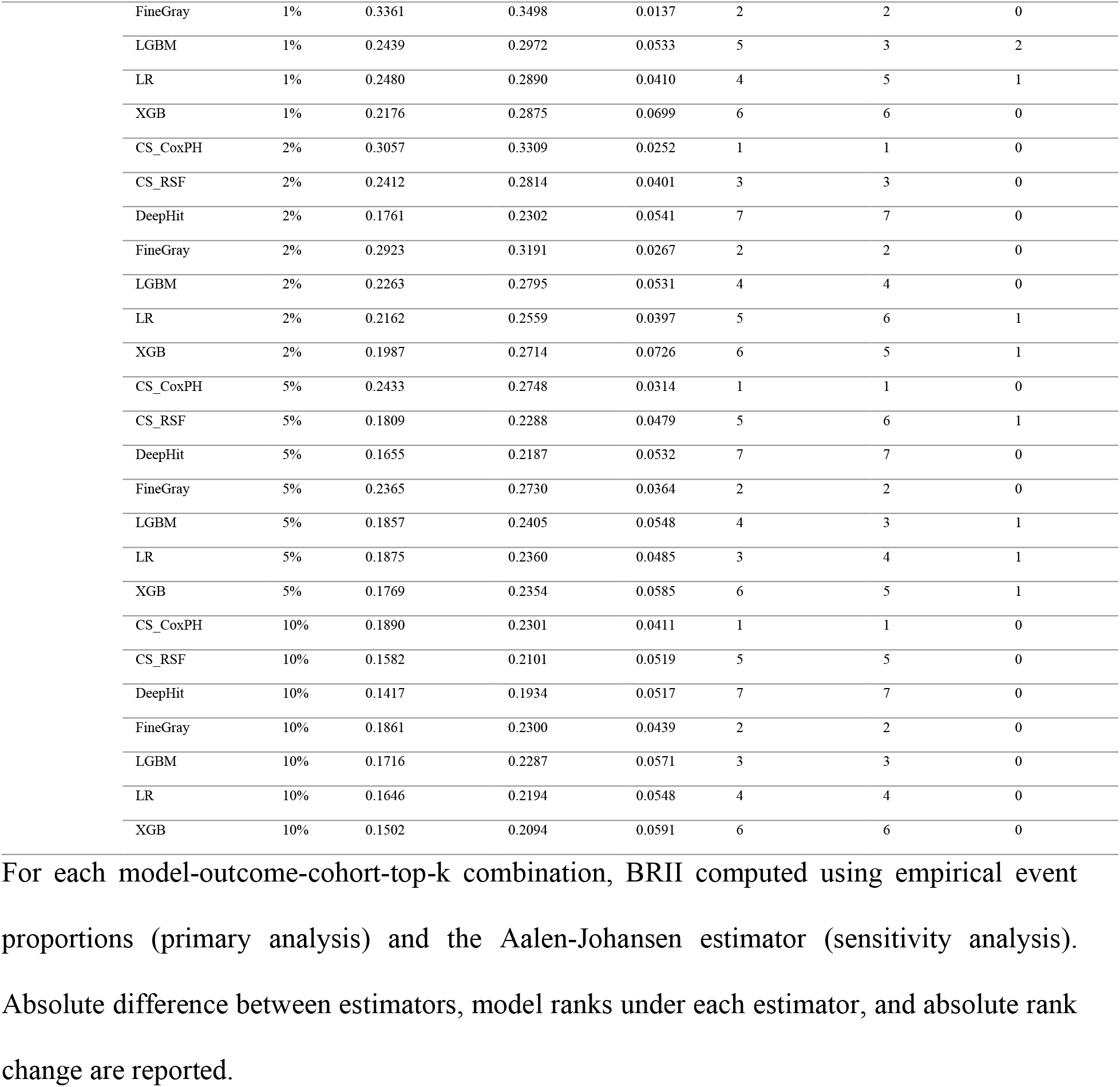
Estimator sensitivity analysis: empirical versus Aalen-Johansen BRII across all cohorts.

**Supplementary Table 2.**
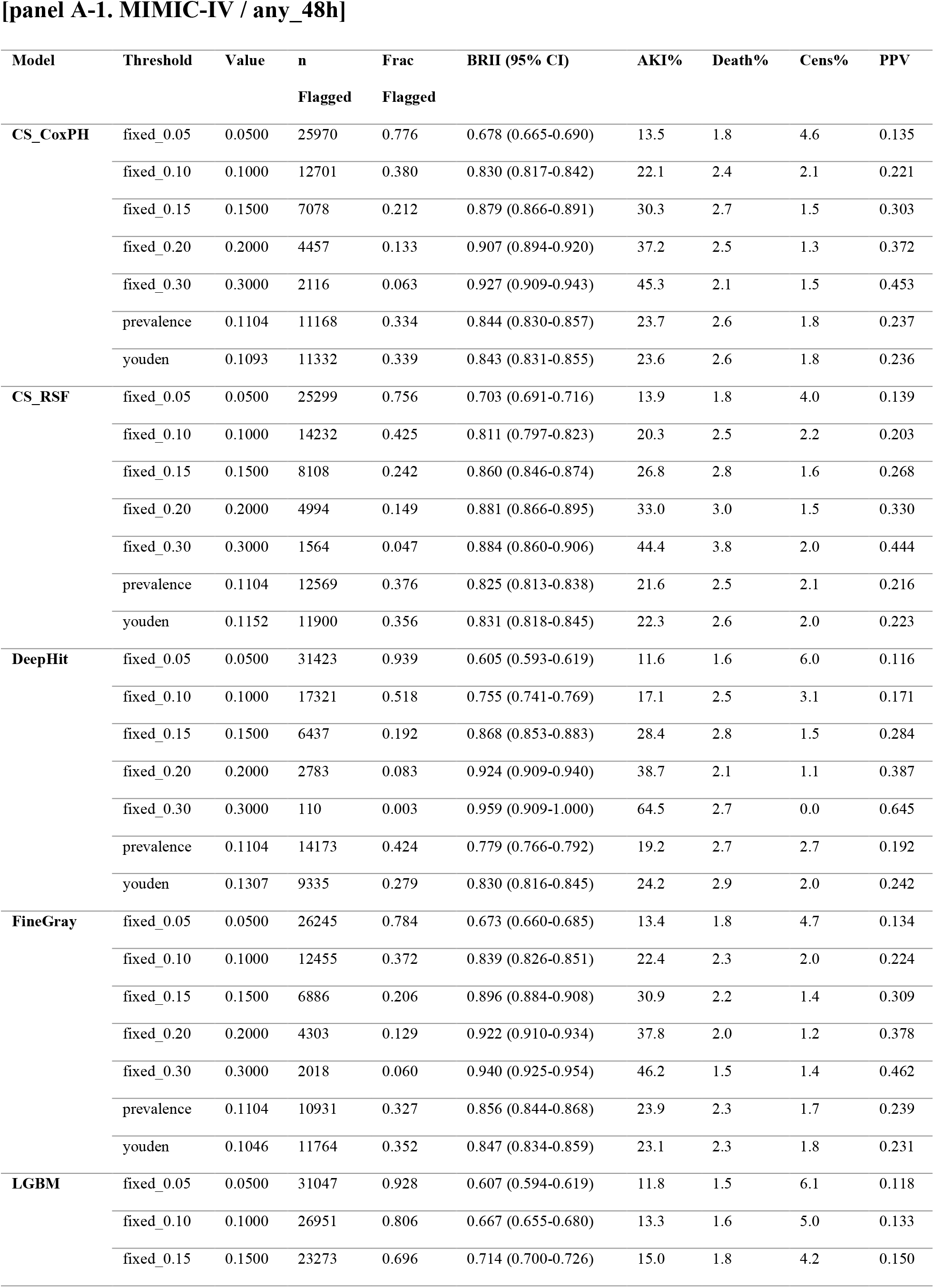
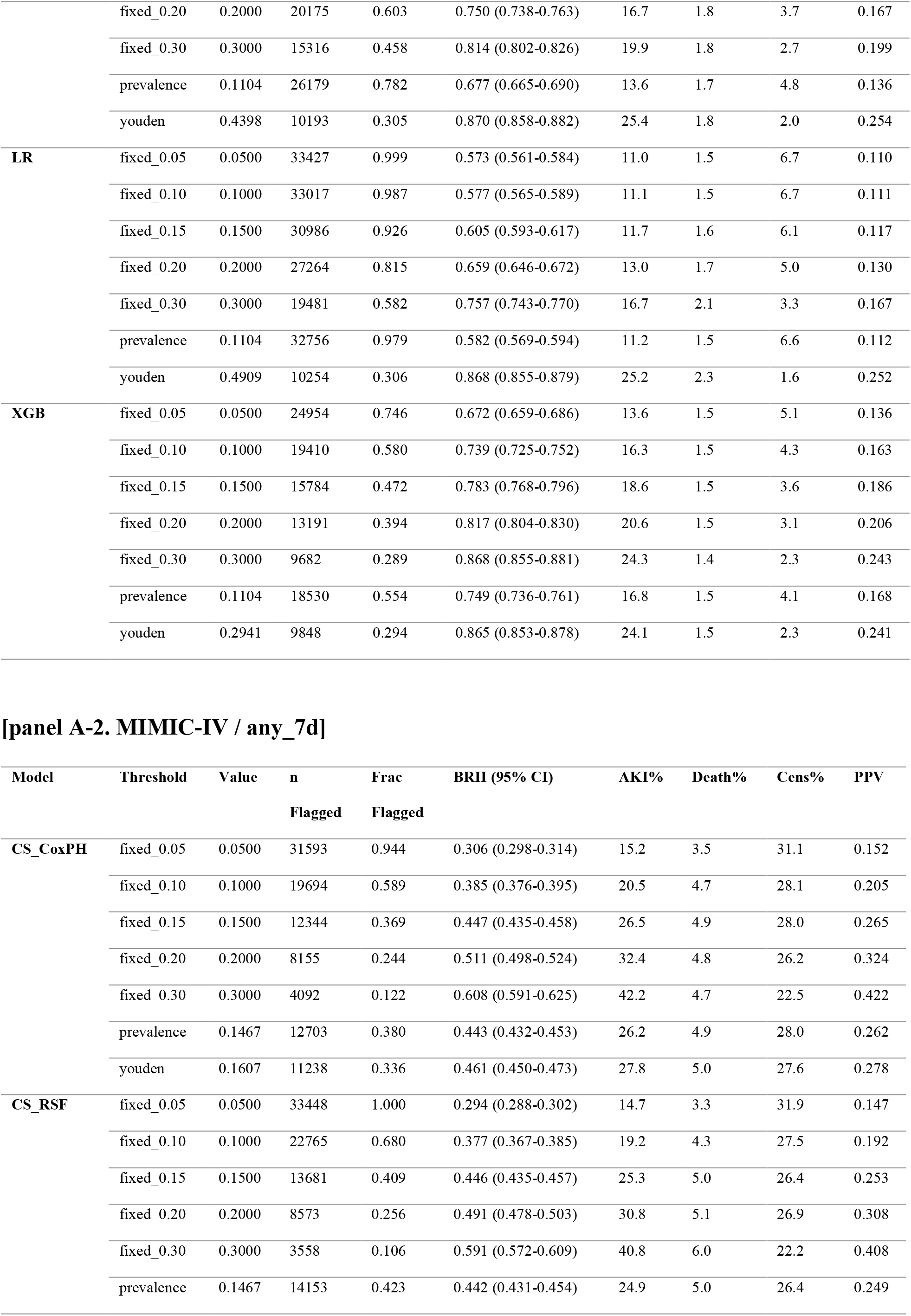
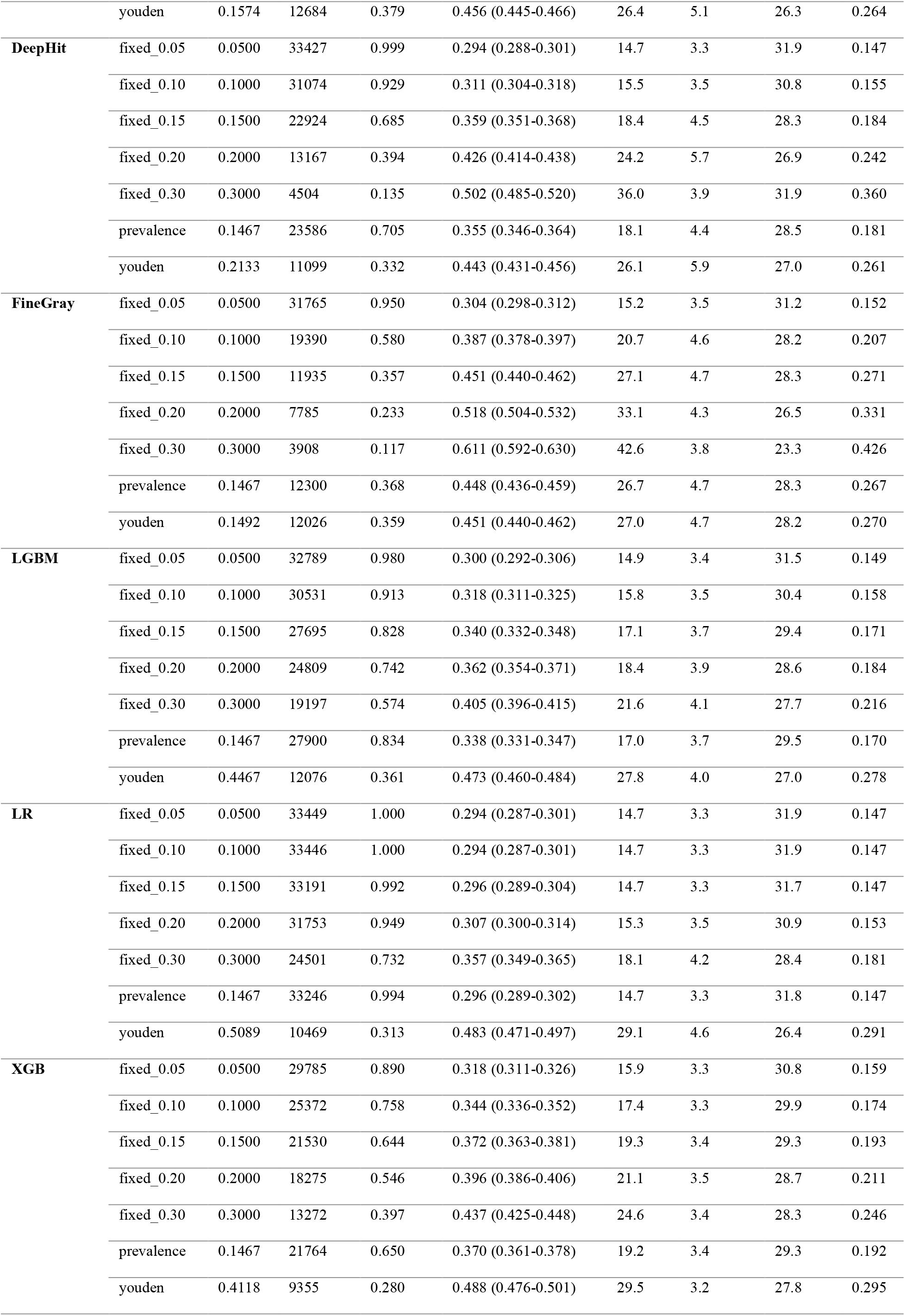
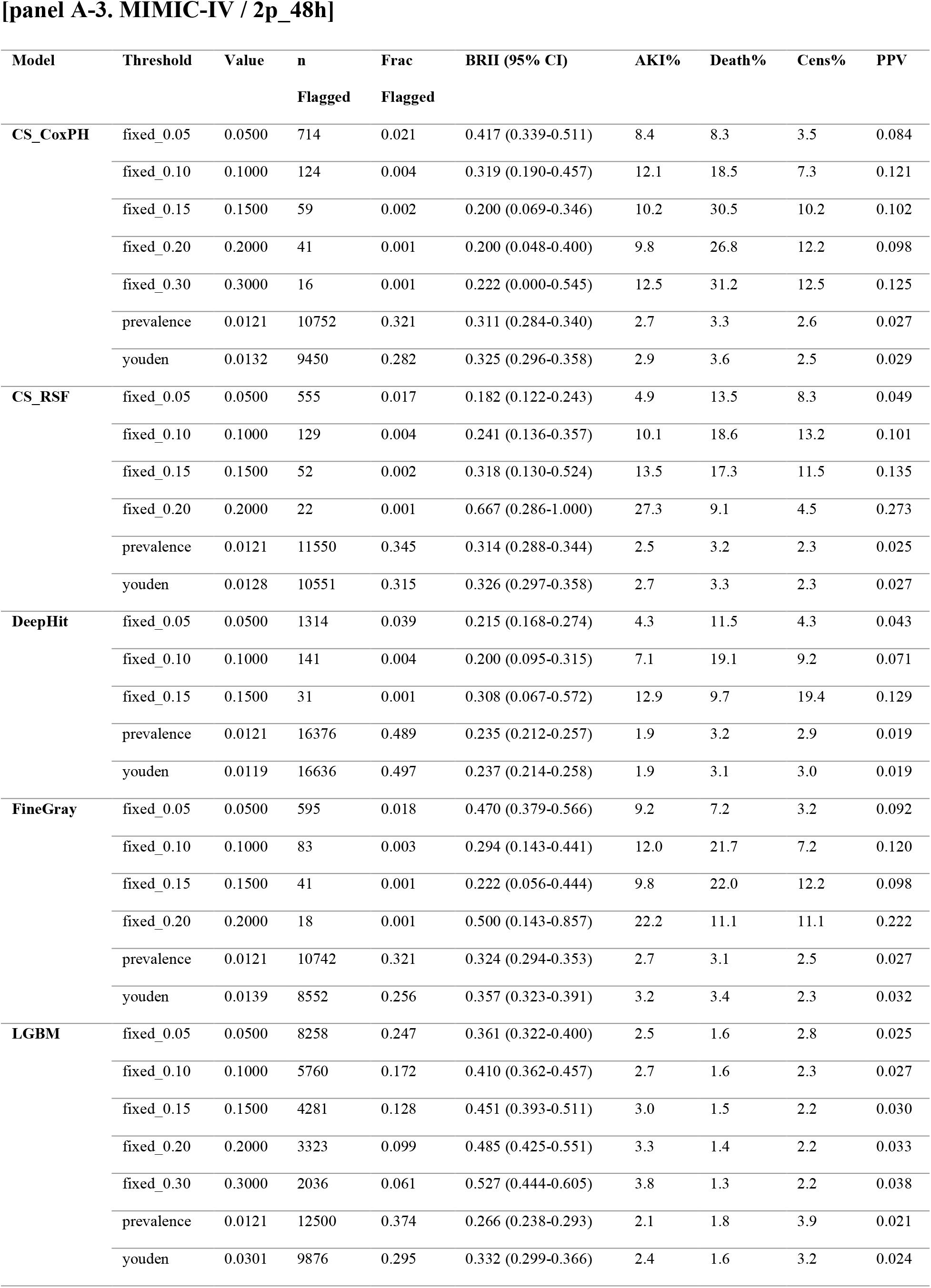
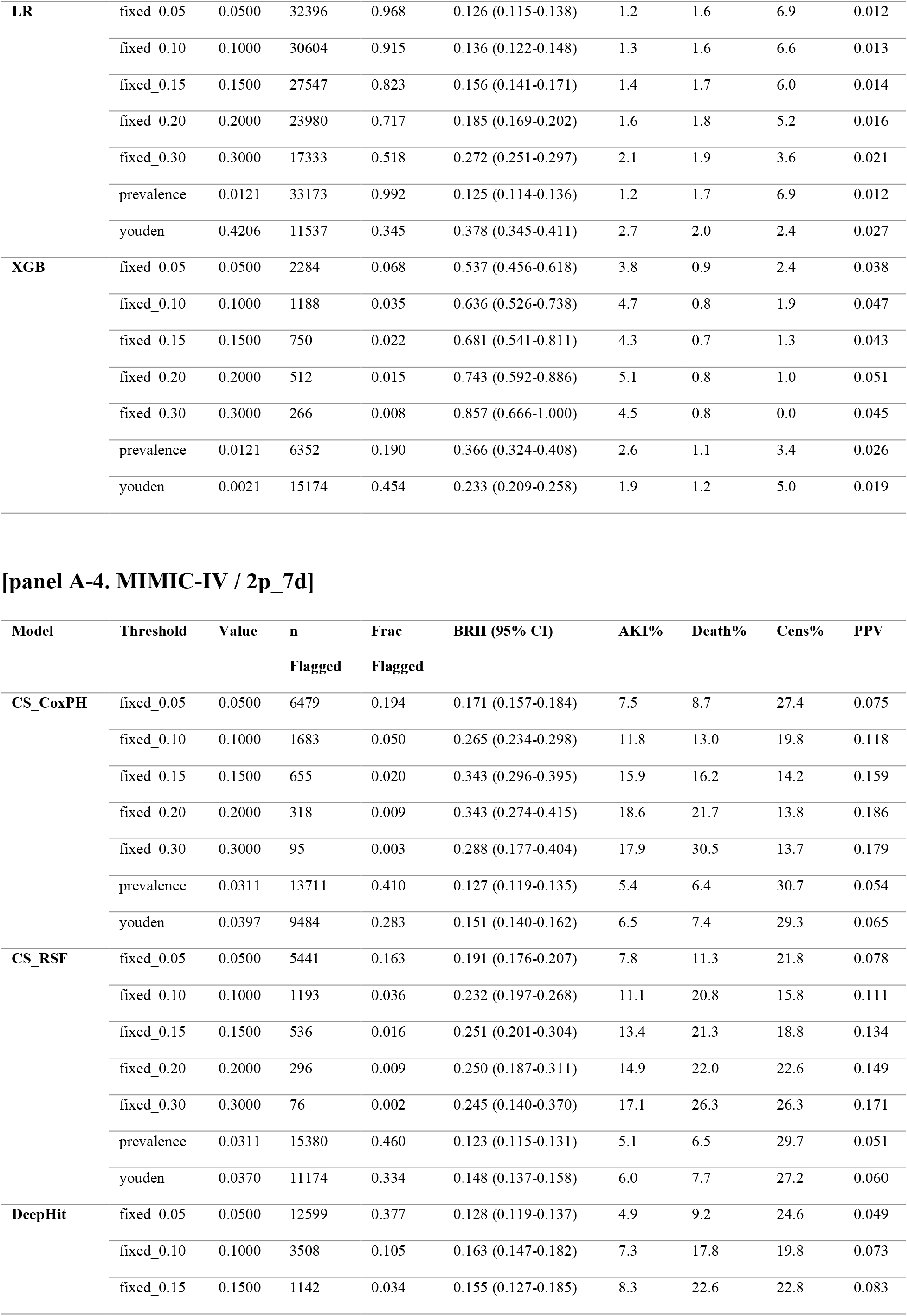
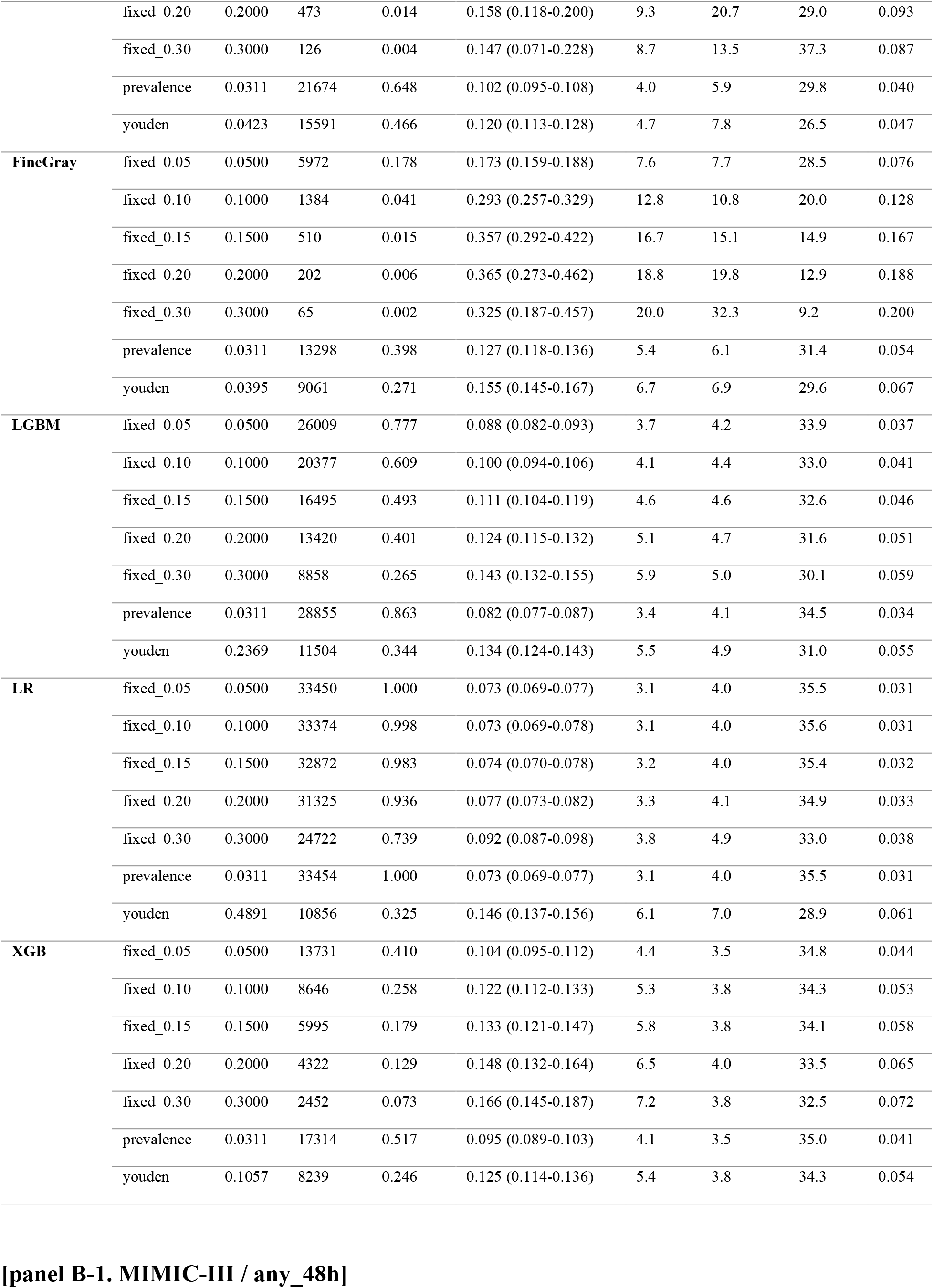
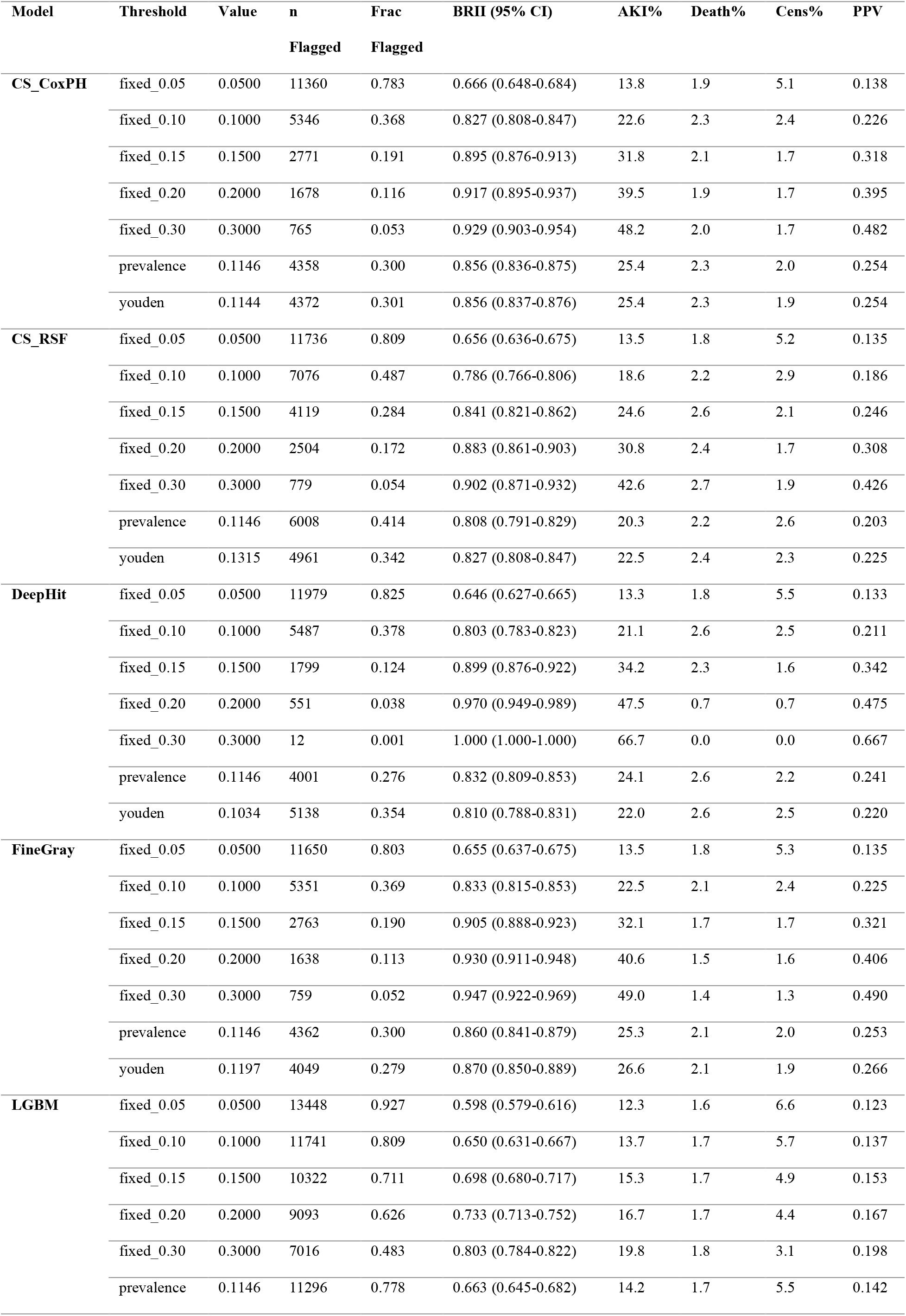
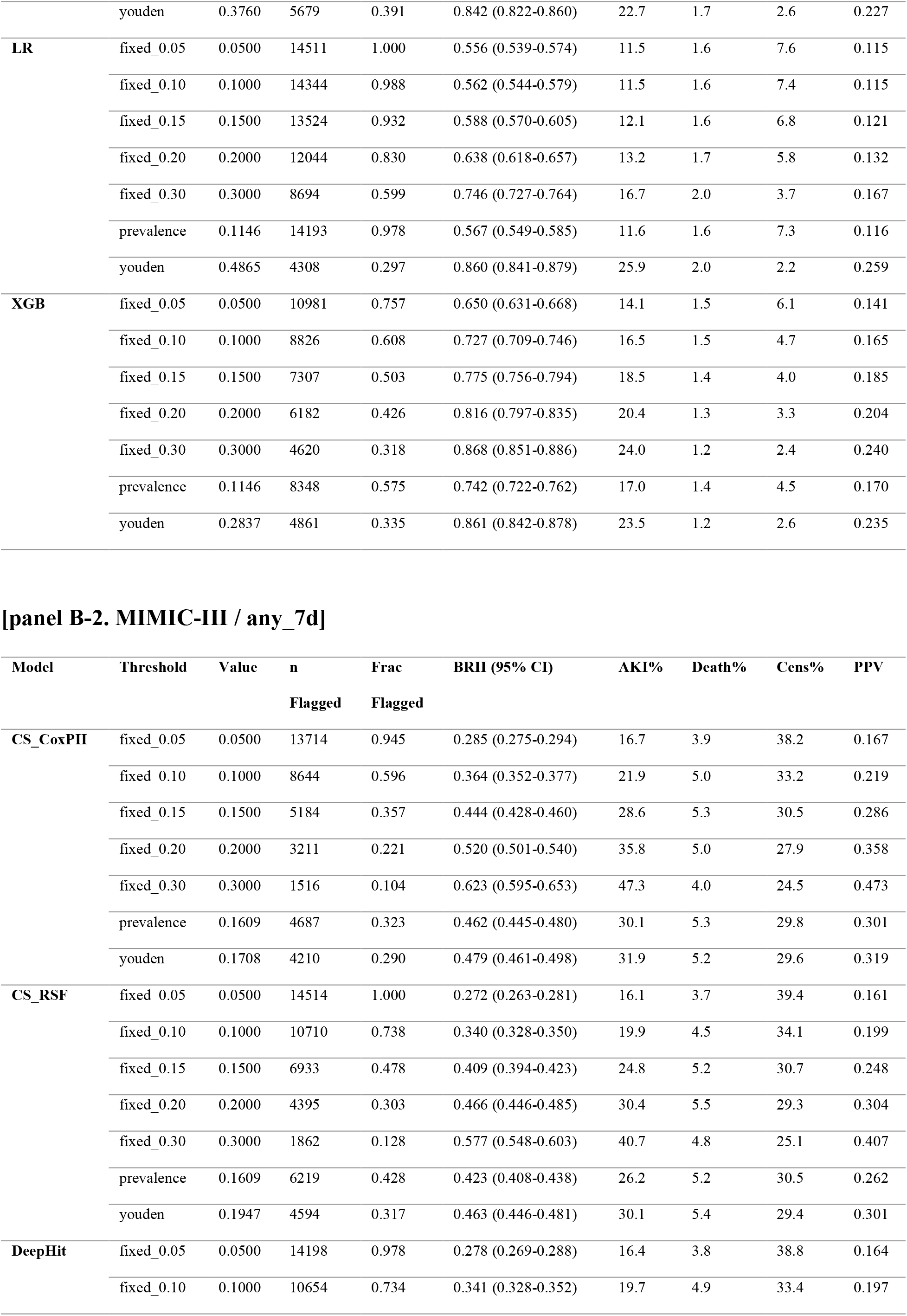
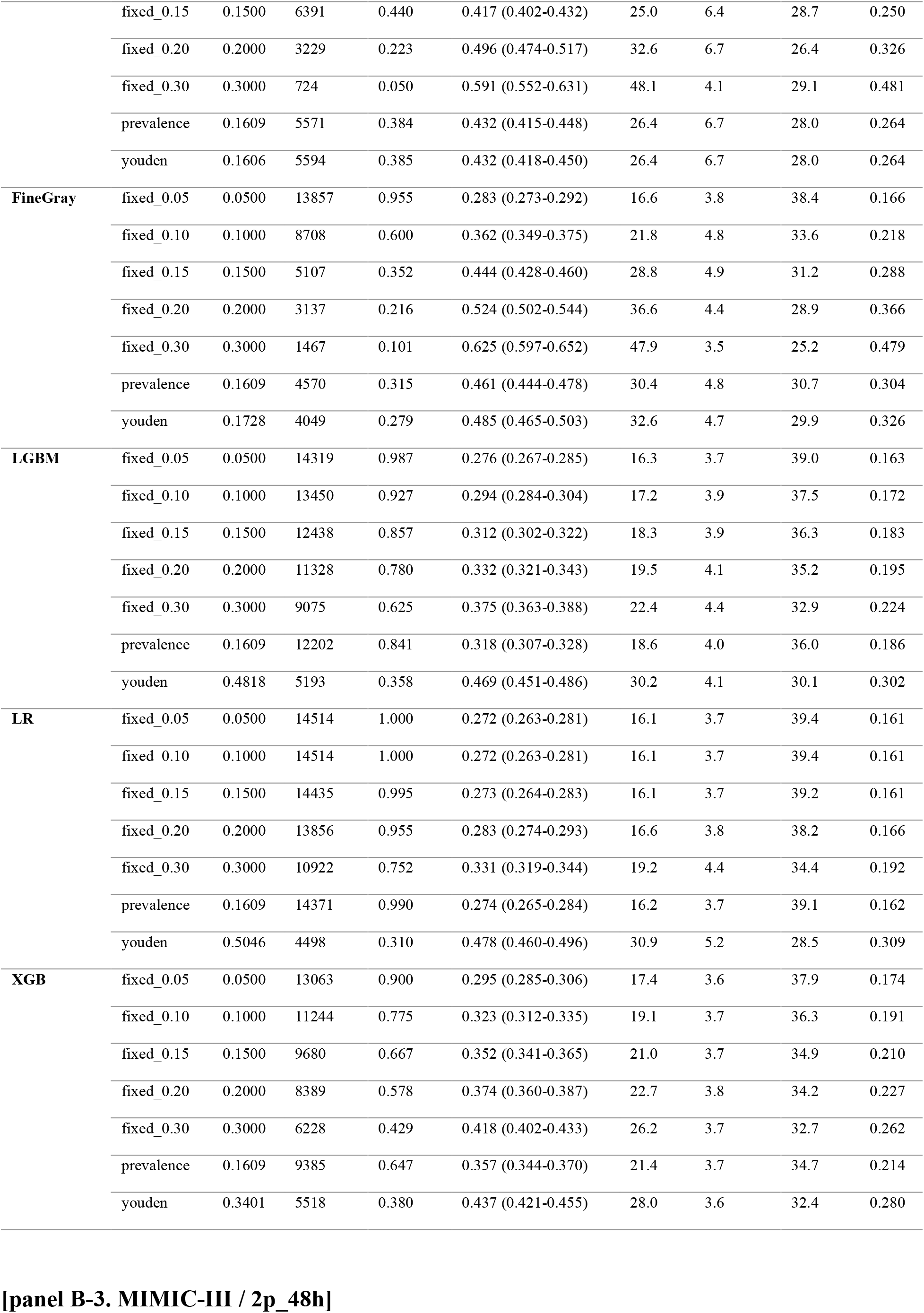
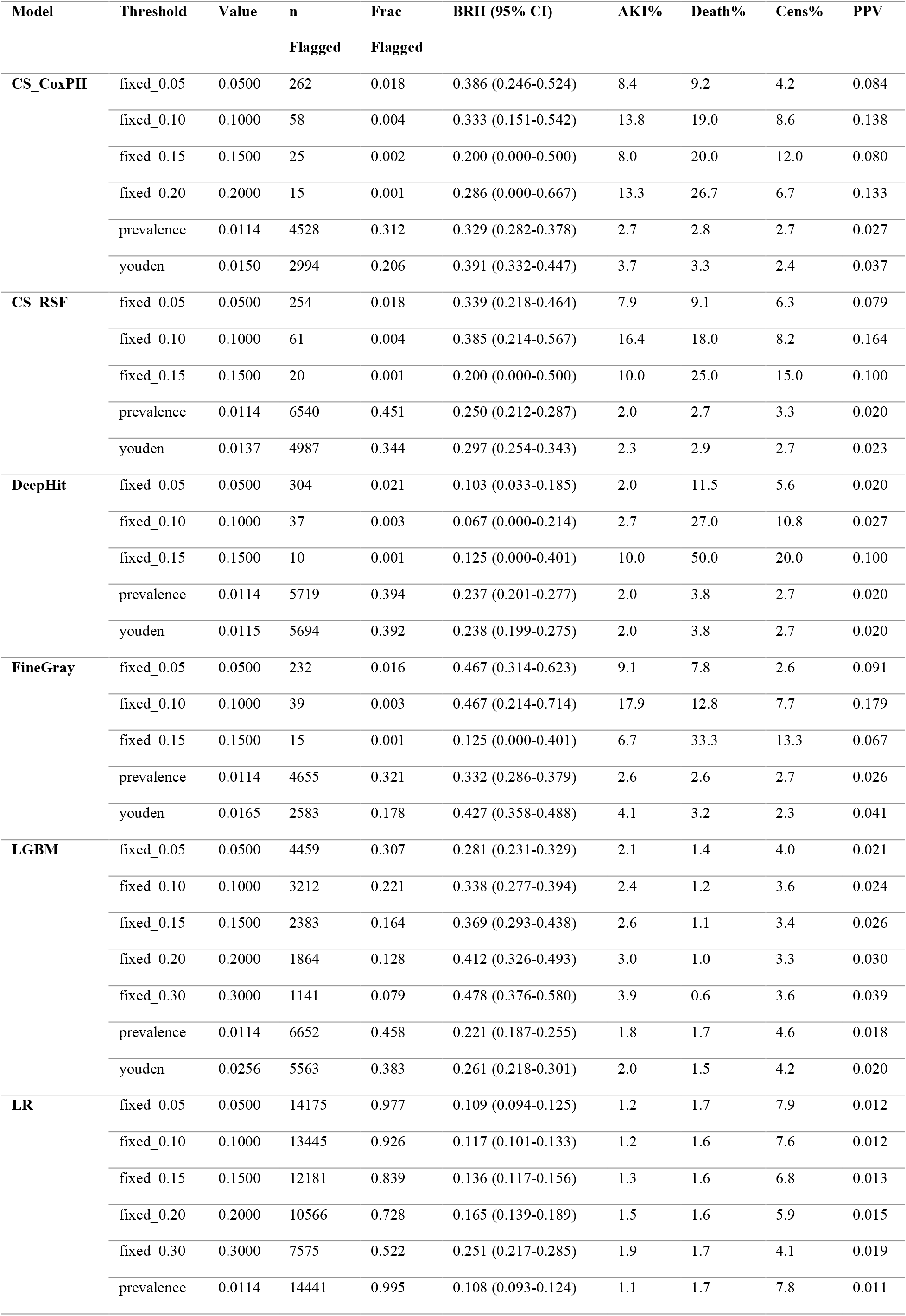
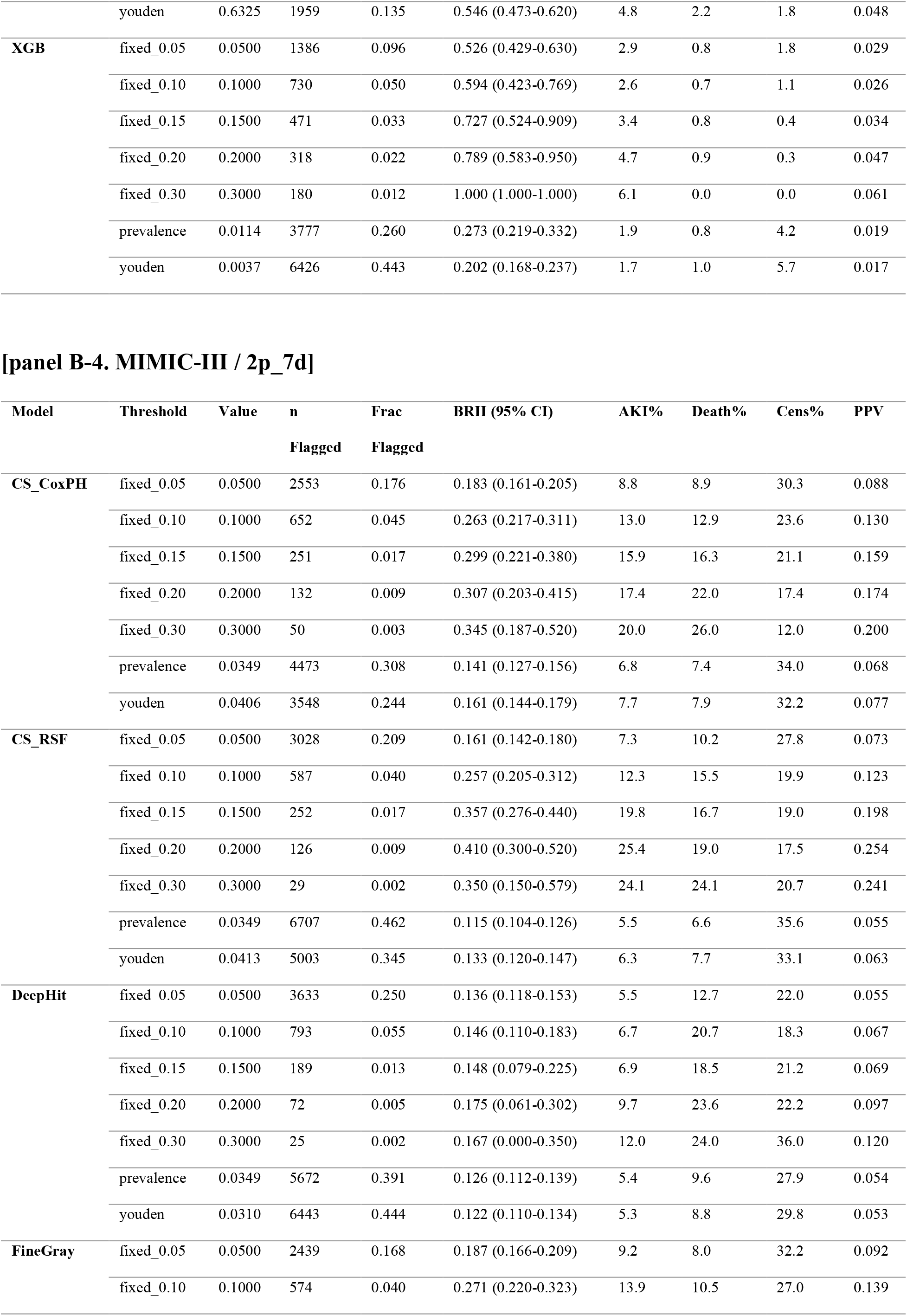
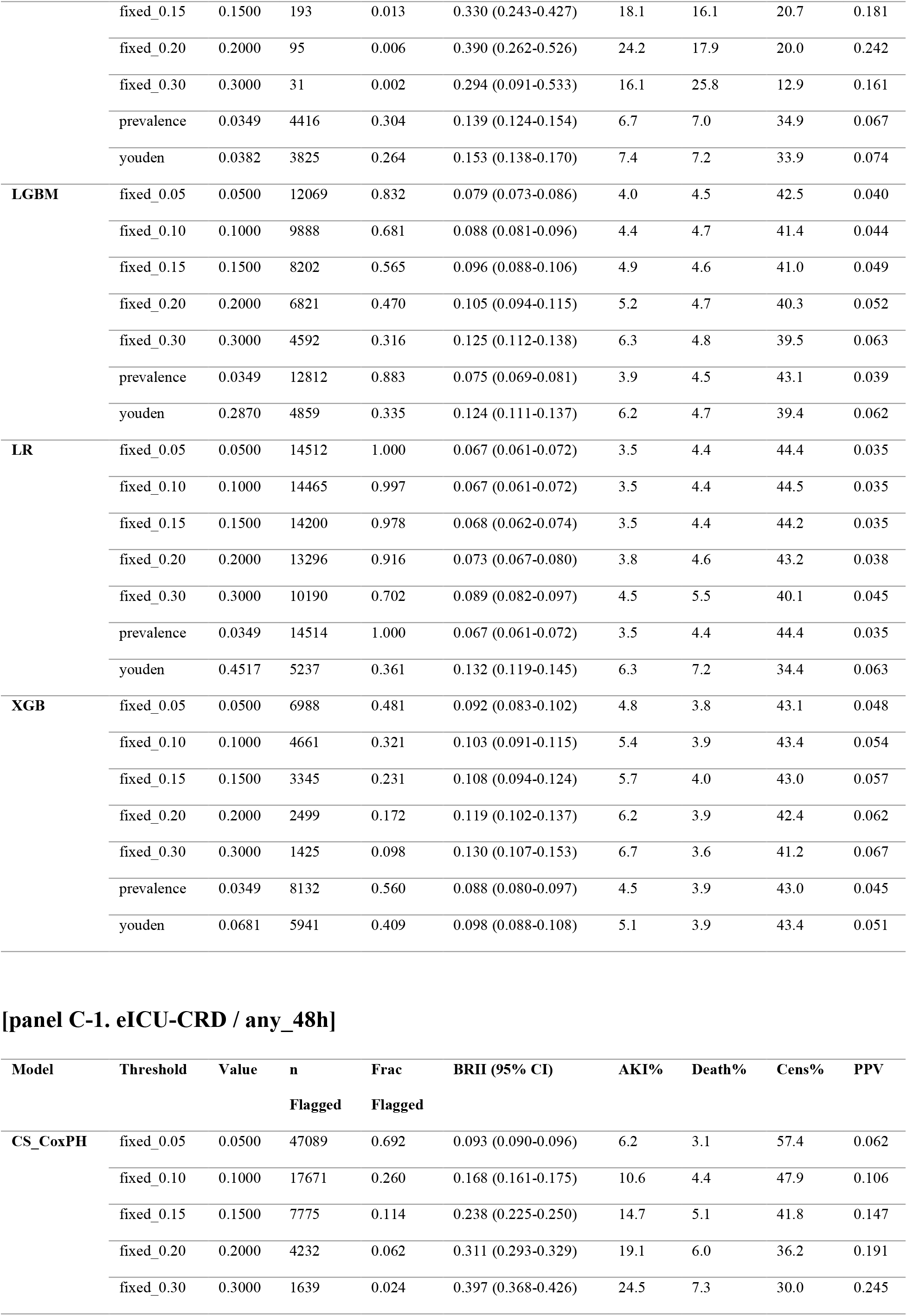
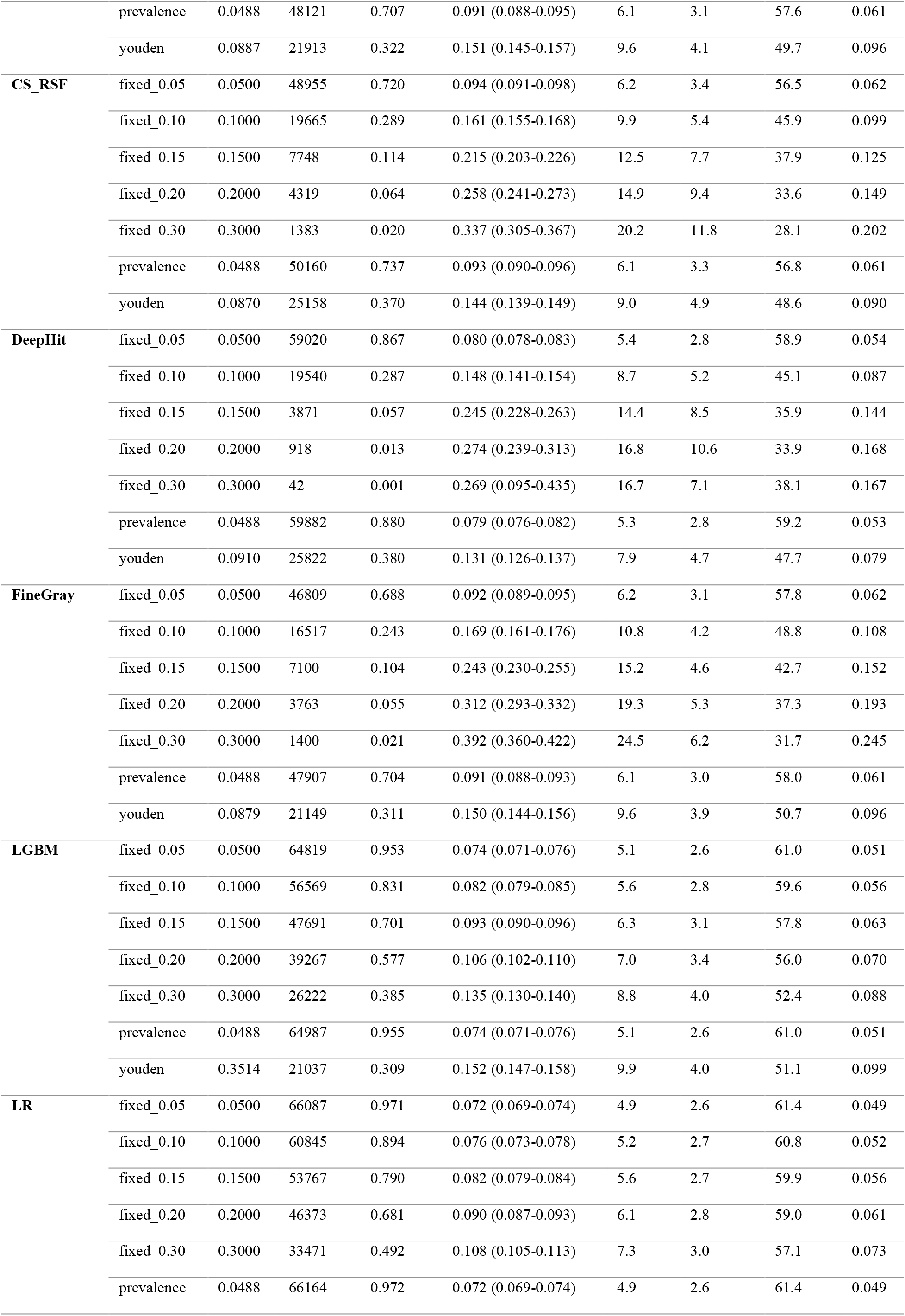
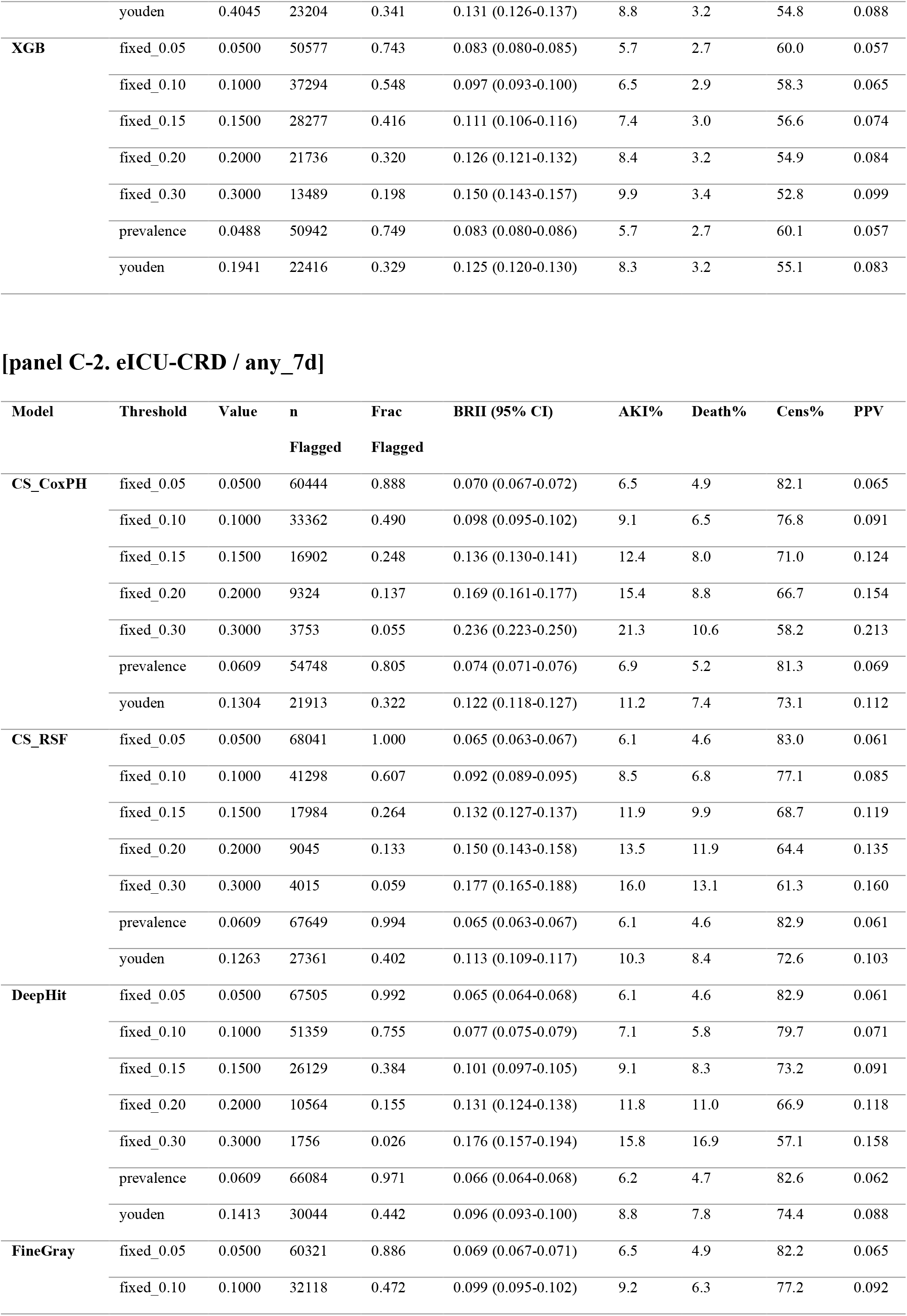
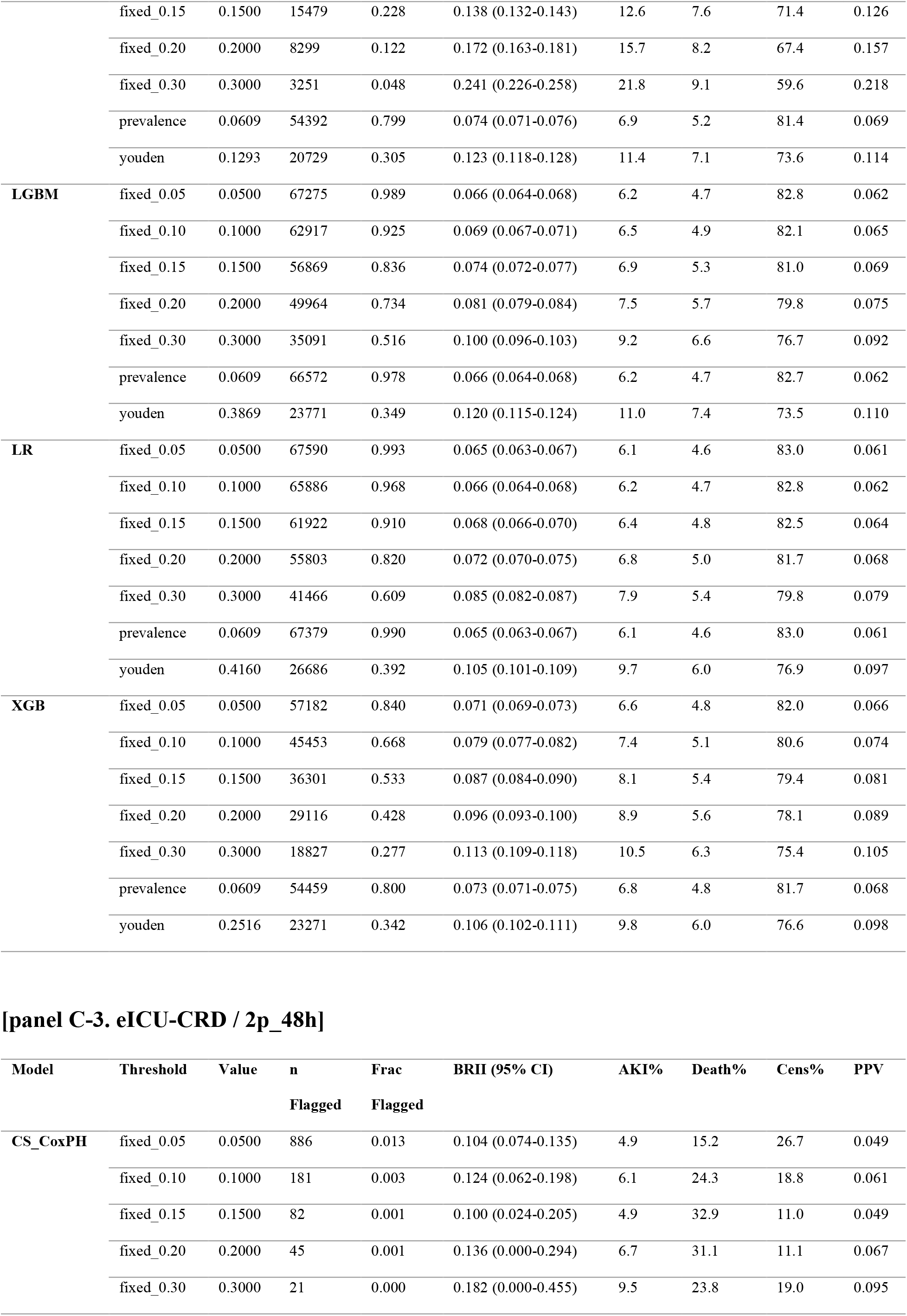
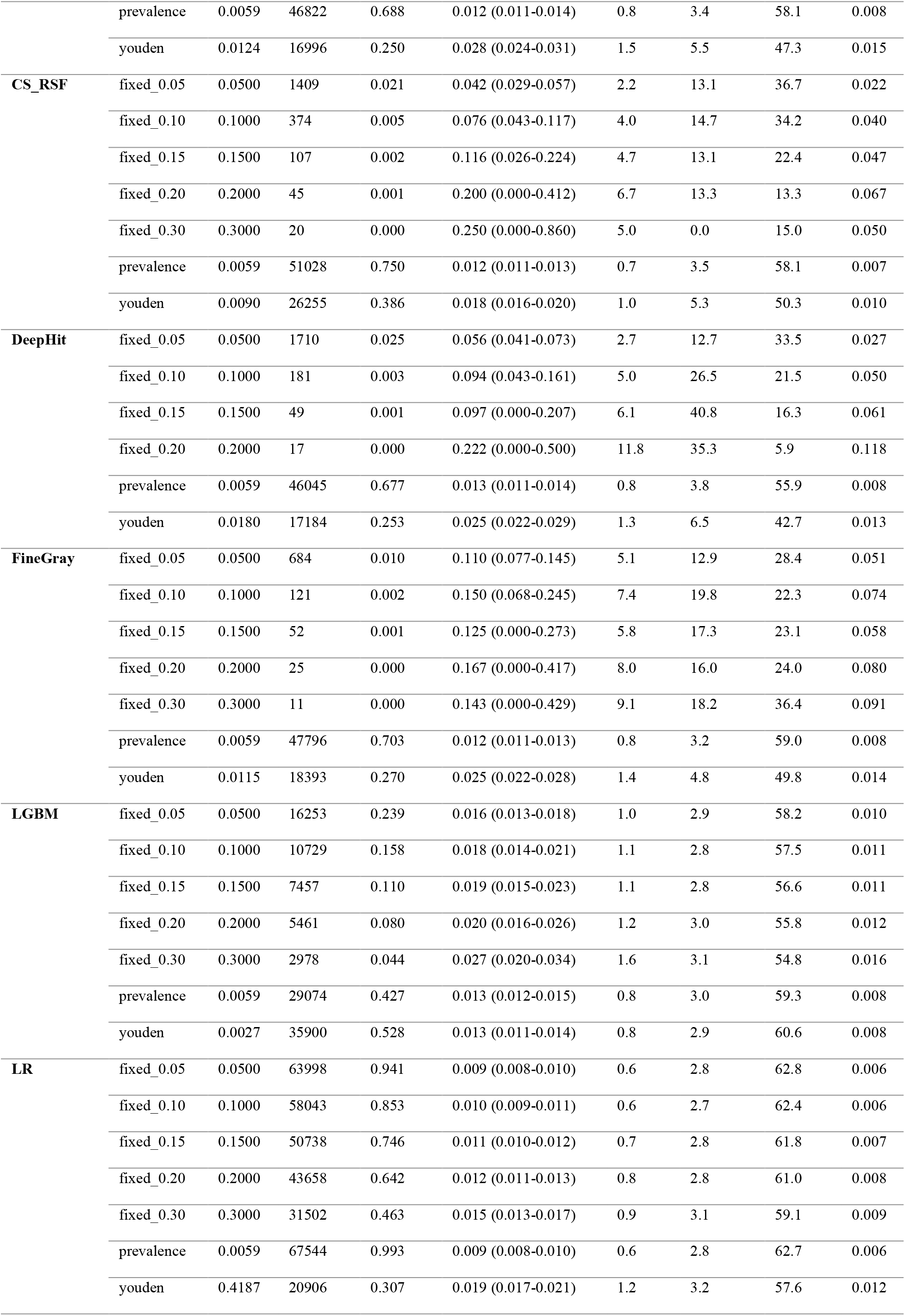
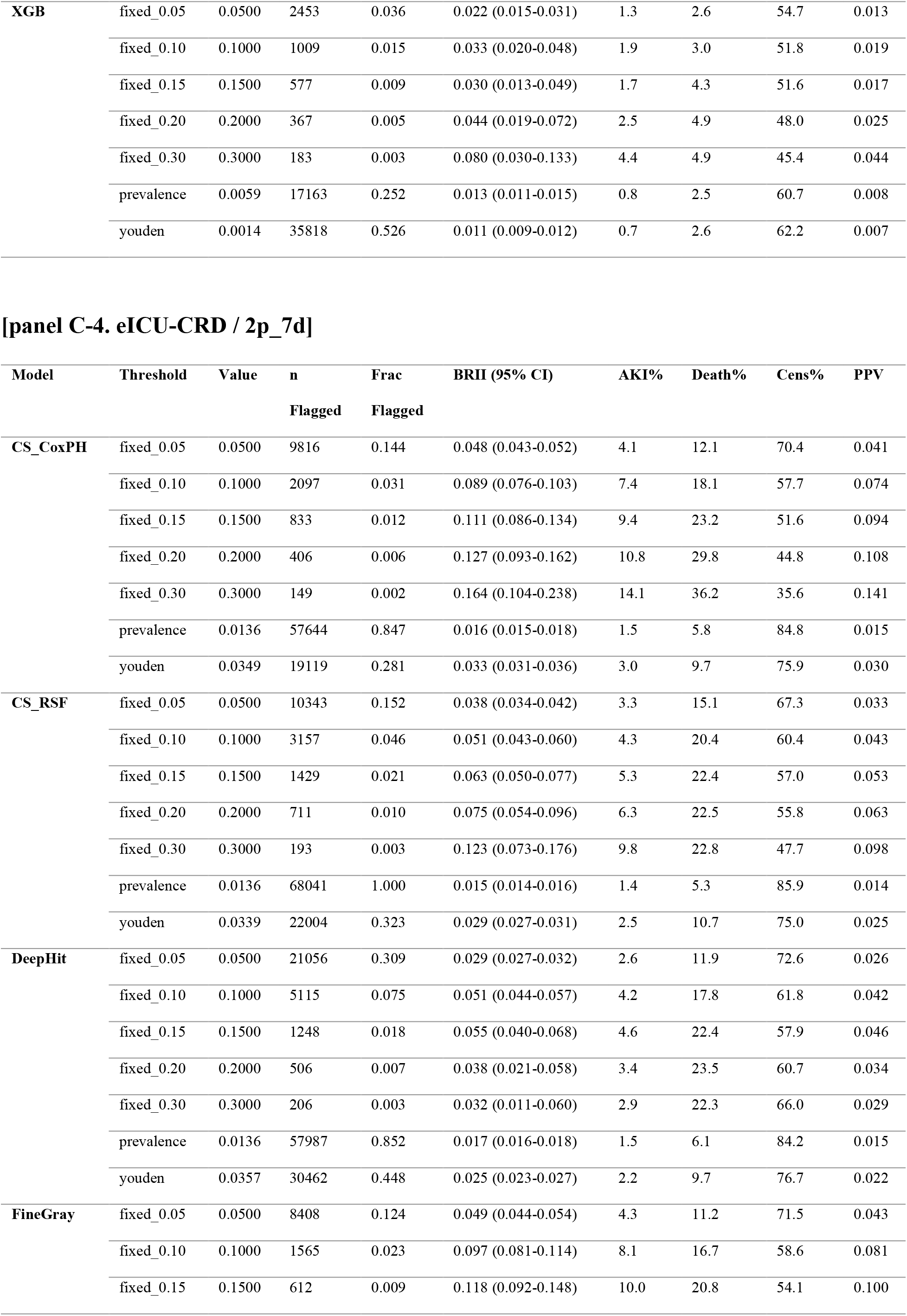
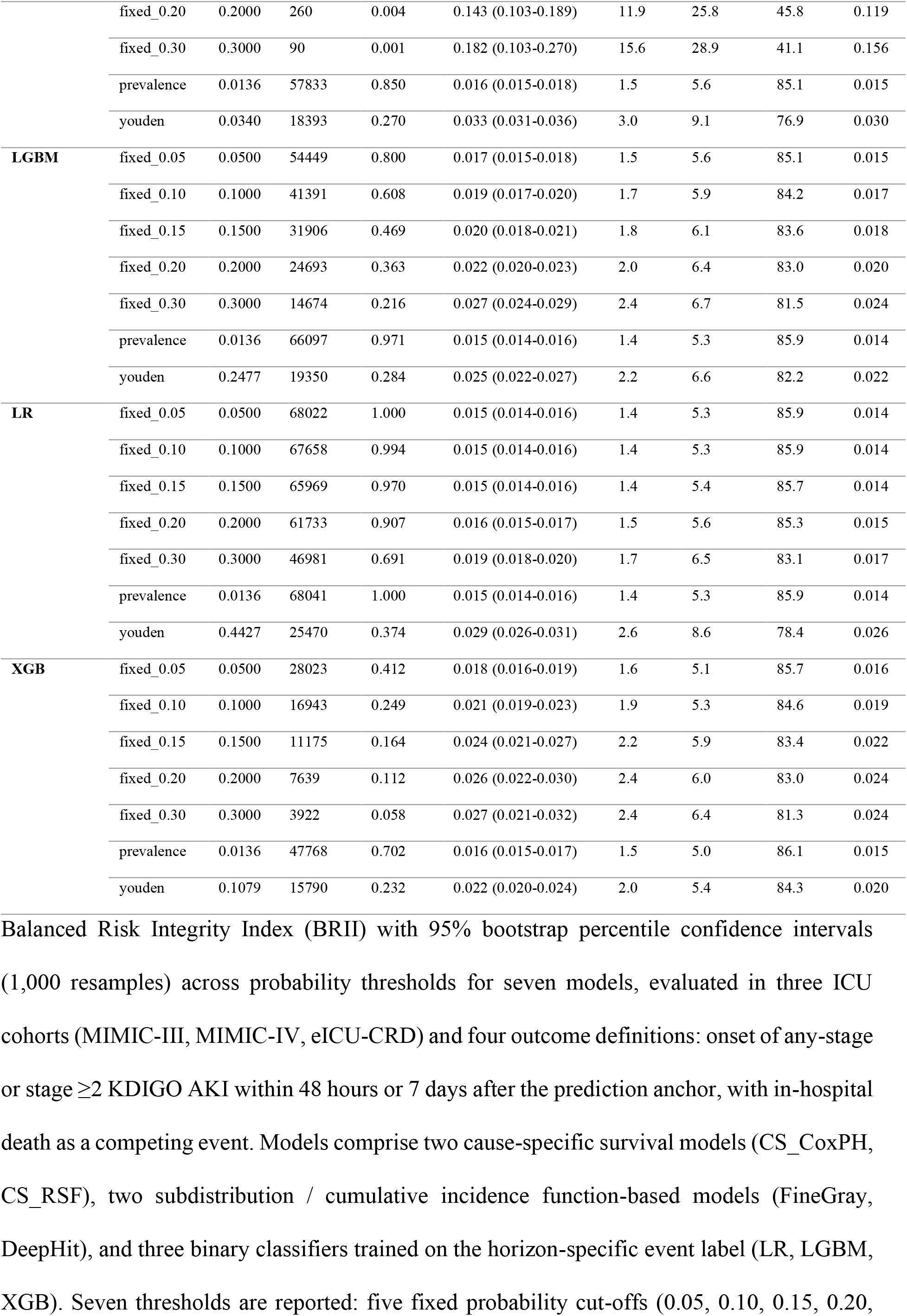
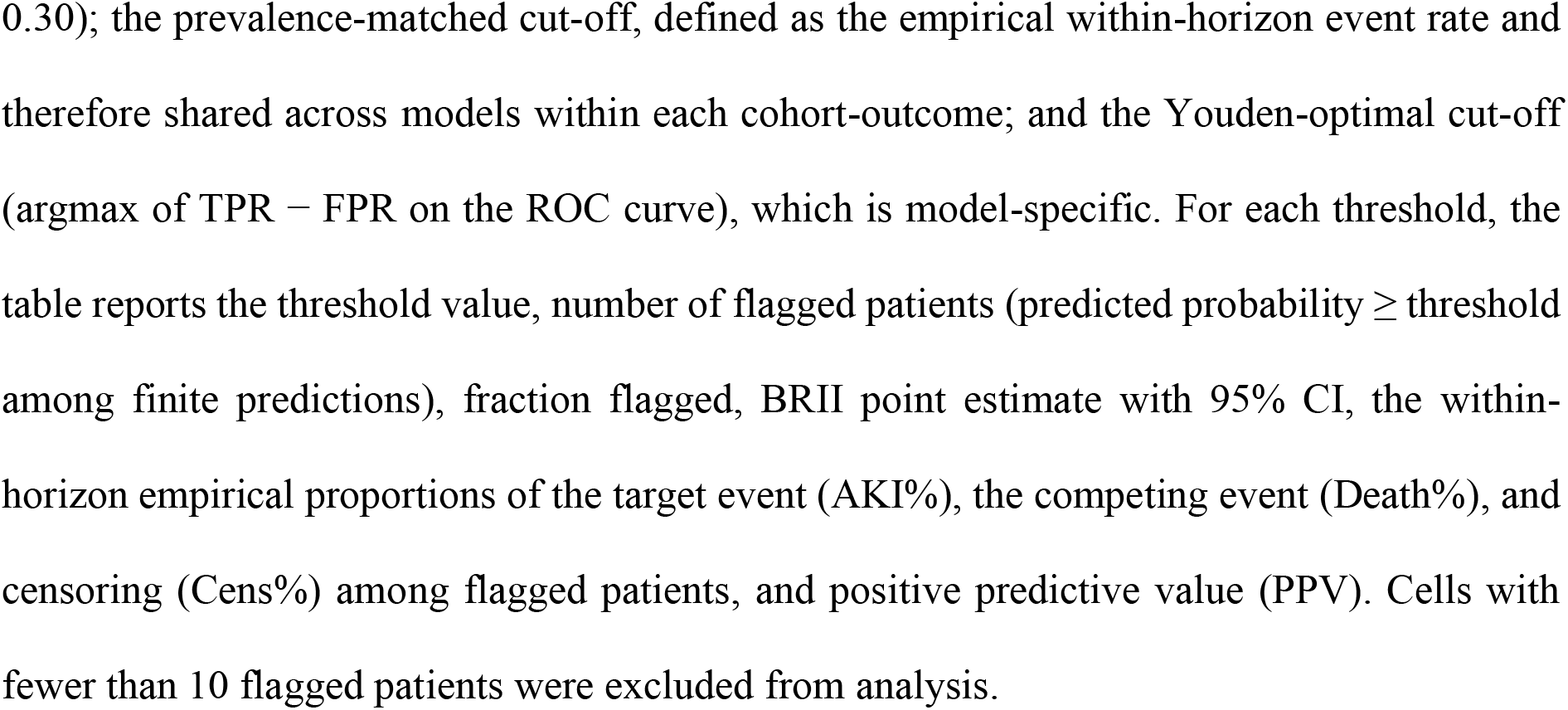
Threshold-based BRII across alternative stratum definitions.

**Supplementary Table 3.**
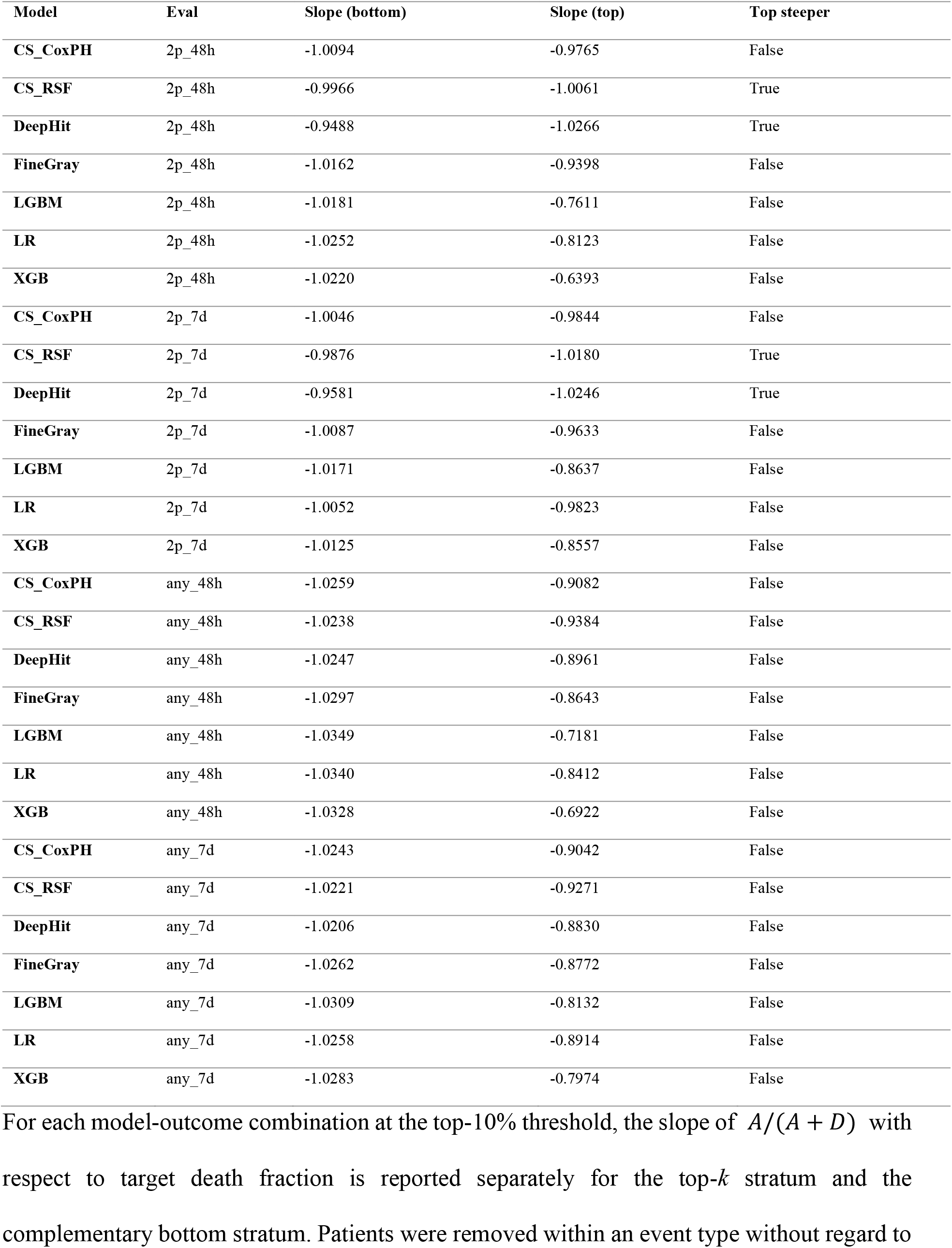
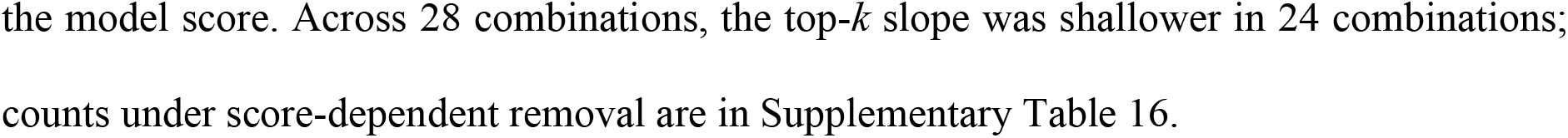
Top-*k* versus bottom stratum compositional slope comparison from the death-contamination sweep.

**Supplementary Table 4.**
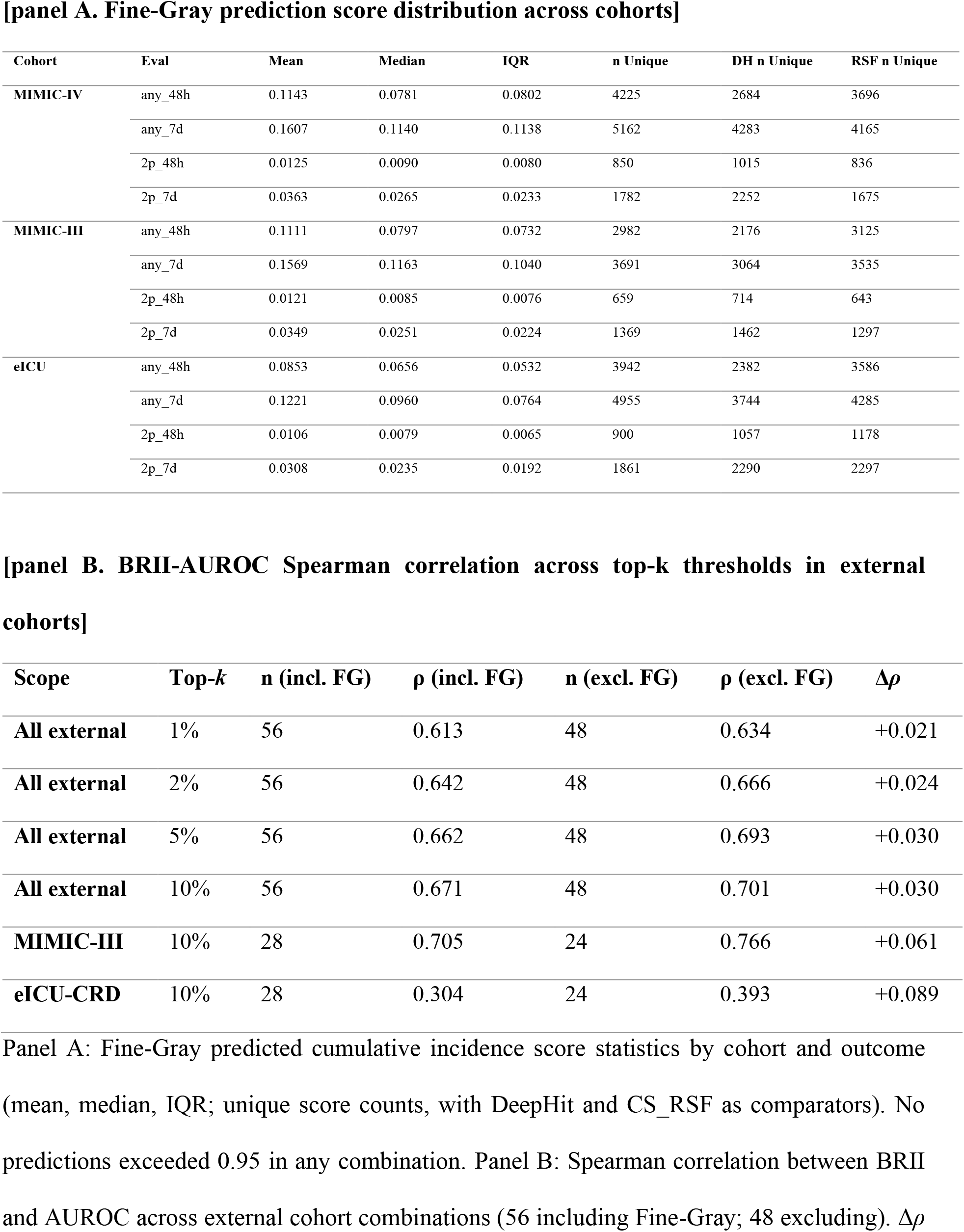
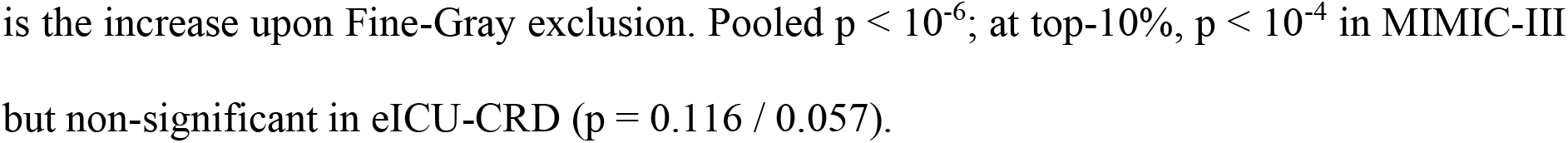
Fine-Gray prediction score distribution and its effect on BRII-AUROC rank correlation.

**Supplementary Table 5.** Cohort-level frequencies of the three outcomes.

| Cohort | Eval | $H$<br>(h) | n | AKI | Deaths | Early<br>Cens % | Med<br>FU<br>(h) | $p_0$ | CIF_AKI | CIF_Death | CIF_Censor | $S(H)$ | BRII_cohort |
| --- | --- | --- | --- | --- | --- | --- | --- | --- | --- | --- | --- | --- | --- |
| MIMIC-IV | any_48h | 48 | 33,455 | 3,692 | 493 | 6.7 | 48.0 | 0.118 | 0.1104 | 0.0147 | 0.0675 | 0.8074 | 0.5730 |
|  | any_7d | 168 | 33,455 | 4,907 | 1,103 | 31.9 | 168.0 | 0.184 | 0.1467 | 0.0330 | 0.3187 | 0.5016 | 0.2943 |
|  | 2p_48h | 48 | 33,455 | 405 | 558 | 6.9 | 48.0 | 0.579 | 0.0121 | 0.0167 | 0.0687 | 0.9025 | 0.1242 |
|  | 2p_7d | 168 | 33,455 | 1,041 | 1,329 | 35.5 | 168.0 | 0.561 | 0.0311 | 0.0397 | 0.3555 | 0.5737 | 0.0730 |
| MIMIC-III | any_48h | 48 | 14,514 | 1,663 | 225 | 7.6 | 48.0 | 0.119 | 0.1146 | 0.0155 | 0.0760 | 0.7939 | 0.5560 |
|  | any_7d | 168 | 14,514 | 2,335 | 534 | 39.4 | 168.0 | 0.186 | 0.1609 | 0.0368 | 0.3938 | 0.4085 | 0.2720 |
|  | 2p_48h | 48 | 14,514 | 166 | 246 | 7.8 | 48.0 | 0.597 | 0.0114 | 0.0169 | 0.0783 | 0.8933 | 0.1072 |
|  | 2p_7d | 168 | 14,514 | 506 | 635 | 44.4 | 168.0 | 0.557 | 0.0349 | 0.0438 | 0.4443 | 0.4771 | 0.0667 |
| eICU-CRD | any_48h | 48 | 68,041 | 3,320 | 1,739 | 61.5 | 27.1 | 0.344 | 0.0488 | 0.0256 | 0.6151 | 0.3105 | 0.0708 |
|  | any_7d | 168 | 68,041 | 4,145 | 3,136 | 83.0 | 25.8 | 0.431 | 0.0609 | 0.0461 | 0.8299 | 0.0631 | 0.0650 |
|  | 2p_48h | 48 | 68,041 | 402 | 1,910 | 62.6 | 28.5 | 0.826 | 0.0059 | 0.0281 | 0.6262 | 0.3398 | 0.0089 |
|  | 2p_7d | 168 | 68,041 | 922 | 3,632 | 85.9 | 27.0 | 0.798 | 0.0136 | 0.0534 | 0.8592 | 0.0738 | 0.0146 |

Aalen-Johansen estimates of cumulative incidence for target AKI (CIF\_AKI), competing death (CIF\_Death), and early censoring (CIF\_Censor), with residual event-free survival $S(H)$ at the prediction horizon, for each cohort-outcome combination. Percentage of patients censored before the horizon and median follow-up among patients without a target event or death are also reported.

**Supplementary Table 6.** Death-fraction threshold crossings from the death-contamination sweep.

**[panel A. any\_48h]**
| Model | Top-k % | $p_0$ @ BRII<0.80 | $p_0$ @ BRII<0.70 | $p_0$ @ BRII<0.50 | $p_0$ @ BRII<0.30 | $p_0$ @ BRII<0.20 |
| --- | --- | --- | --- | --- | --- | --- |
| CS_CoxPH | 5 | 0.287 | 0.405 | 0.613 | 0.786 | 0.864 |
|  | 10 | 0.254 | 0.356 | 0.561 | 0.741 | 0.832 |
|  | 15 | 0.215 | 0.311 | 0.510 | 0.705 | 0.802 |
|  | 20 | 0.194 | 0.288 | 0.479 | 0.678 | 0.783 |
| CS_RSf | 5 | 0.202 | 0.303 | 0.502 | 0.703 | 0.803 |
|  | 10 | 0.197 | 0.295 | 0.496 | 0.699 | 0.801 |
|  | 15 | 0.192 | 0.285 | 0.476 | 0.678 | 0.785 |
|  | 20 | 0.180 | 0.268 | 0.454 | 0.657 | 0.767 |
| DeepHit | 5 | 0.316 | 0.445 | 0.644 | 0.806 | 0.879 |
|  | 10 | 0.257 | 0.365 | 0.570 | 0.754 | 0.841 |
|  | 15 | 0.206 | 0.297 | 0.482 | 0.677 | 0.782 |
|  | 20 | 0.166 | 0.251 | 0.431 | 0.637 | 0.749 |
| FineGray | 5 | 0.335 | 0.463 | 0.665 | 0.824 | 0.892 |
|  | 10 | 0.289 | 0.400 | 0.598 | 0.769 | 0.851 |
|  | 15 | 0.237 | 0.342 | 0.545 | 0.732 | 0.824 |
|  | 20 | 0.211 | 0.308 | 0.503 | 0.699 | 0.799 |
| LGBM | 5 | 0.514 | 0.635 | 0.794 | 0.900 | NA |
|  | 10 | 0.375 | 0.504 | 0.694 | 0.840 | NA |
|  | 15 | 0.301 | 0.409 | 0.606 | 0.781 | 0.859 |
|  | 20 | 0.243 | 0.347 | 0.543 | 0.730 | 0.822 |
| LR | 5 | 0.310 | 0.433 | 0.643 | 0.807 | 0.880 |
|  | 10 | 0.287 | 0.405 | 0.610 | 0.782 | 0.859 |
|  | 15 | 0.245 | 0.349 | 0.548 | 0.738 | 0.828 |
|  | 20 | 0.213 | 0.312 | 0.511 | 0.706 | 0.805 |
| XGB | 5 | 0.490 | 0.622 | 0.798 | NA | NA |
|  | 10 | 0.345 | 0.473 | 0.671 | 0.828 | 0.894 |
|  | 15 | 0.283 | 0.394 | 0.591 | 0.772 | 0.853 |
|  | 20 | 0.236 | 0.338 | 0.536 | 0.724 | 0.819 |

**[panel B. any\_7d]**
| Model | Top-k % | $p_0$ @ BRII<0.80 | $p_0$ @ BRII<0.70 | $p_0$ @ BRII<0.50 | $p_0$ @ BRII<0.30 | $p_0$ @ BRII<0.20 |
| --- | --- | --- | --- | --- | --- | --- |
| CS_CoxPH | 5 | 0.000 | 0.122 | 0.319 | 0.509 | 0.637 |
|  | 10 | 0.000 | 0.000 | 0.270 | 0.447 | 0.574 |
|  | 15 | 0.000 | 0.000 | 0.238 | 0.399 | 0.526 |
|  | 20 | 0.000 | 0.000 | 0.210 | 0.369 | 0.490 |
| CS_RSf | 5 | 0.000 | 0.081 | 0.288 | 0.475 | 0.606 |
|  | 10 | 0.000 | 0.000 | 0.248 | 0.421 | 0.553 |
|  | 15 | 0.000 | 0.000 | 0.222 | 0.383 | 0.506 |
|  | 20 | 0.000 | 0.000 | 0.185 | 0.354 | 0.478 |
| DeepHit | 5 | 0.000 | 0.000 | 0.221 | 0.395 | 0.524 |
|  | 10 | 0.000 | 0.000 | 0.171 | 0.357 | 0.484 |
|  | 15 | 0.000 | 0.000 | 0.132 | 0.342 | 0.468 |
|  | 20 | 0.000 | 0.000 | 0.101 | 0.324 | 0.449 |
| FineGray | 5 | 0.000 | 0.120 | 0.322 | 0.508 | 0.636 |
|  | 10 | 0.000 | 0.000 | 0.268 | 0.445 | 0.573 |
|  | 15 | 0.000 | 0.000 | 0.235 | 0.398 | 0.525 |
|  | 20 | 0.000 | 0.000 | 0.209 | 0.367 | 0.489 |
| LGBM | 5 | 0.000 | 0.108 | 0.309 | 0.488 | 0.612 |
|  | 10 | 0.000 | 0.000 | 0.258 | 0.435 | 0.564 |
|  | 15 | 0.000 | 0.000 | 0.232 | 0.395 | 0.524 |
|  | 20 | 0.000 | 0.000 | 0.212 | 0.375 | 0.499 |
| LR | 5 | 0.000 | 0.121 | 0.317 | 0.508 | 0.637 |
|  | 10 | 0.000 | 0.000 | 0.271 | 0.450 | 0.576 |
|  | 15 | 0.000 | 0.000 | 0.241 | 0.407 | 0.536 |
|  | 20 | 0.000 | 0.000 | 0.213 | 0.376 | 0.498 |
| XGB | 5 | 0.000 | 0.000 | 0.290 | 0.477 | 0.602 |
|  | 10 | 0.000 | 0.000 | 0.244 | 0.413 | 0.543 |
|  | 15 | 0.000 | 0.000 | 0.215 | 0.379 | 0.503 |
|  | 20 | 0.000 | 0.000 | 0.190 | 0.358 | 0.482 |

[panel C. 2p\_48h]
| Model | Top-k % | $p_0$ @ BRII<0.80 | $p_0$ @ BRII<0.70 | $p_0$ @ BRII<0.50 | $p_0$ @ BRII<0.30 | $p_0$ @ BRII<0.20 |
| --- | --- | --- | --- | --- | --- | --- |
| CS_CoxPH | 5 | 0.000 | 0.060 | 0.436 | 0.686 | 0.787 |
|  | 10 | 0.000 | 0.000 | 0.361 | 0.661 | 0.768 |
|  | 15 | 0.000 | 0.000 | 0.315 | 0.646 | 0.756 |
|  | 20 | 0.000 | 0.000 | 0.253 | 0.624 | 0.741 |
| **CS_RSf** | 5 | 0.000 | 0.000 | 0.023 | 0.464 | 0.637 |
|  | 10 | 0.000 | 0.000 | 0.200 | 0.586 | 0.716 |
|  | 15 | 0.000 | 0.000 | 0.261 | 0.619 | 0.737 |
|  | 20 | 0.000 | 0.000 | 0.238 | 0.619 | 0.735 |
| **DeepHit** | 5 | 0.000 | 0.000 | 0.092 | 0.462 | 0.631 |
|  | 10 | 0.000 | 0.000 | 0.027 | 0.425 | 0.613 |
|  | 15 | 0.000 | 0.000 | 0.000 | 0.413 | 0.605 |
|  | 20 | 0.000 | 0.000 | 0.000 | 0.430 | 0.622 |
| **FineGray** | 5 | 0.000 | 0.186 | 0.554 | 0.750 | 0.834 |
|  | 10 | 0.000 | 0.000 | 0.457 | 0.705 | 0.803 |
|  | 15 | 0.000 | 0.000 | 0.391 | 0.681 | 0.787 |
|  | 20 | 0.000 | 0.000 | 0.324 | 0.658 | 0.767 |
| **LGBM** | 5 | 0.000 | 0.000 | 0.610 | 0.783 | 0.860 |
|  | 10 | 0.000 | 0.000 | 0.520 | 0.747 | 0.832 |
|  | 15 | 0.000 | 0.000 | 0.358 | 0.712 | 0.807 |
|  | 20 | 0.000 | 0.000 | 0.183 | 0.686 | 0.788 |
| **LR** | 5 | 0.086 | 0.377 | 0.662 | 0.817 | 0.886 |
|  | 10 | 0.000 | 0.224 | 0.622 | 0.797 | 0.870 |
|  | 15 | 0.000 | 0.138 | 0.606 | 0.780 | 0.858 |
|  | 20 | 0.000 | 0.000 | 0.528 | 0.743 | 0.832 |
| **XGB** | 5 | 0.000 | 0.000 | 0.664 | 0.819 | 0.885 |
|  | 10 | 0.000 | 0.000 | 0.540 | 0.764 | 0.846 |
|  | 15 | 0.000 | 0.000 | 0.000 | 0.691 | 0.791 |
|  | 20 | 0.000 | 0.000 | 0.000 | 0.640 | 0.751 |

[panel D. 2p\_7d]
| **Model** | **Top-k %** | **$p_0$ @ BRII<0.80** | **$p_0$ @ BRII<0.70** | **$p_0$ @ BRII<0.50** | **$p_0$ @ BRII<0.30** | **$p_0$ @ BRII<0.20** |
| --- | --- | --- | --- | --- | --- | --- |
| **CS_CoxPH** | 5 | 0.000 | 0.000 | 0.000 | 0.418 | 0.648 |
|  | 10 | 0.000 | 0.000 | 0.000 | 0.000 | 0.559 |
|  | 15 | 0.000 | 0.000 | 0.000 | 0.000 | 0.425 |
|  | 20 | 0.000 | 0.000 | 0.000 | 0.000 | 0.163 |
| **CS_RSf** | 5 | 0.000 | 0.000 | 0.000 | 0.352 | 0.611 |
|  | 10 | 0.000 | 0.000 | 0.000 | 0.057 | 0.566 |
|  | 15 | 0.000 | 0.000 | 0.000 | 0.000 | 0.520 |
|  | 20 | 0.000 | 0.000 | 0.000 | 0.000 | 0.404 |
| **DeepHit** | 5 | 0.000 | 0.000 | 0.000 | 0.000 | 0.430 |
|  | 10 | 0.000 | 0.000 | 0.000 | 0.000 | 0.409 |
|  | 15 | 0.000 | 0.000 | 0.000 | 0.000 | 0.304 |
|  | 20 | 0.000 | 0.000 | 0.000 | 0.000 | 0.178 |
| **FineGray** | 5 | 0.000 | 0.000 | 0.000 | 0.464 | 0.663 |
|  | 10 | 0.000 | 0.000 | 0.000 | 0.000 | 0.567 |
|  | 15 | 0.000 | 0.000 | 0.000 | 0.000 | 0.411 |
|  | 20 | 0.000 | 0.000 | 0.000 | 0.000 | 0.000 |
| **LGBM** | 5 | 0.000 | 0.000 | 0.000 | 0.000 | 0.620 |
|  | 10 | 0.000 | 0.000 | 0.000 | 0.000 | 0.552 |
|  | 15 | 0.000 | 0.000 | 0.000 | 0.000 | 0.000 |
|  | 20 | 0.000 | 0.000 | 0.000 | 0.000 | 0.000 |
| **LR** | 5 | 0.000 | 0.000 | 0.000 | 0.480 | 0.661 |
|  | 10 | 0.000 | 0.000 | 0.000 | 0.000 | 0.598 |
|  | 15 | 0.000 | 0.000 | 0.000 | 0.000 | 0.503 |
|  | 20 | 0.000 | 0.000 | 0.000 | 0.000 | 0.302 |
| **XGB** | 5 | 0.000 | 0.000 | 0.000 | 0.000 | 0.000 |
|  | 10 | 0.000 | 0.000 | 0.000 | 0.000 | 0.000 |
|  | 15 | 0.000 | 0.000 | 0.000 | 0.000 | 0.000 |
|  | 20 | 0.000 | 0.000 | 0.000 | 0.000 | 0.000 |

**Supplementary Table 7.**
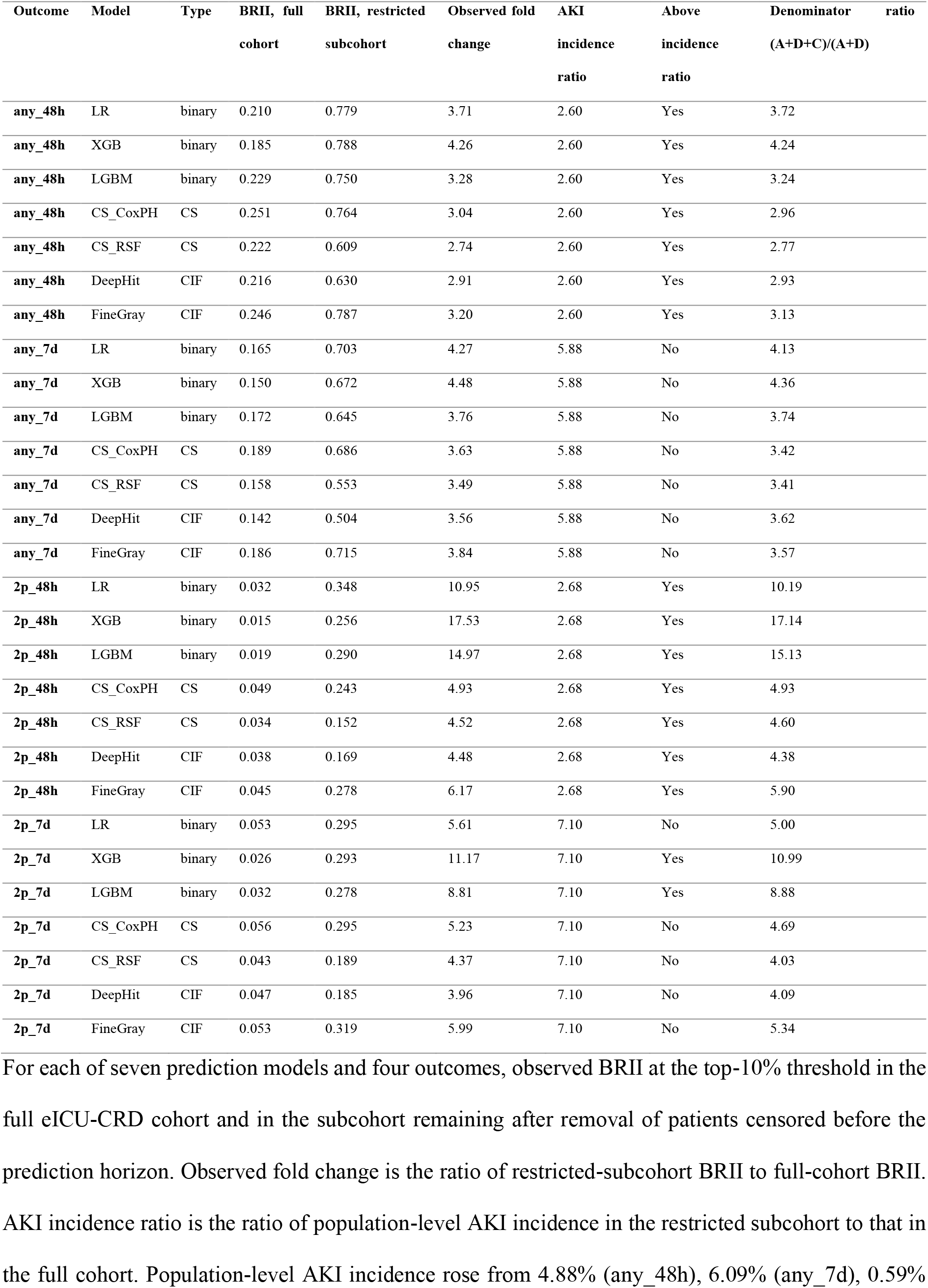
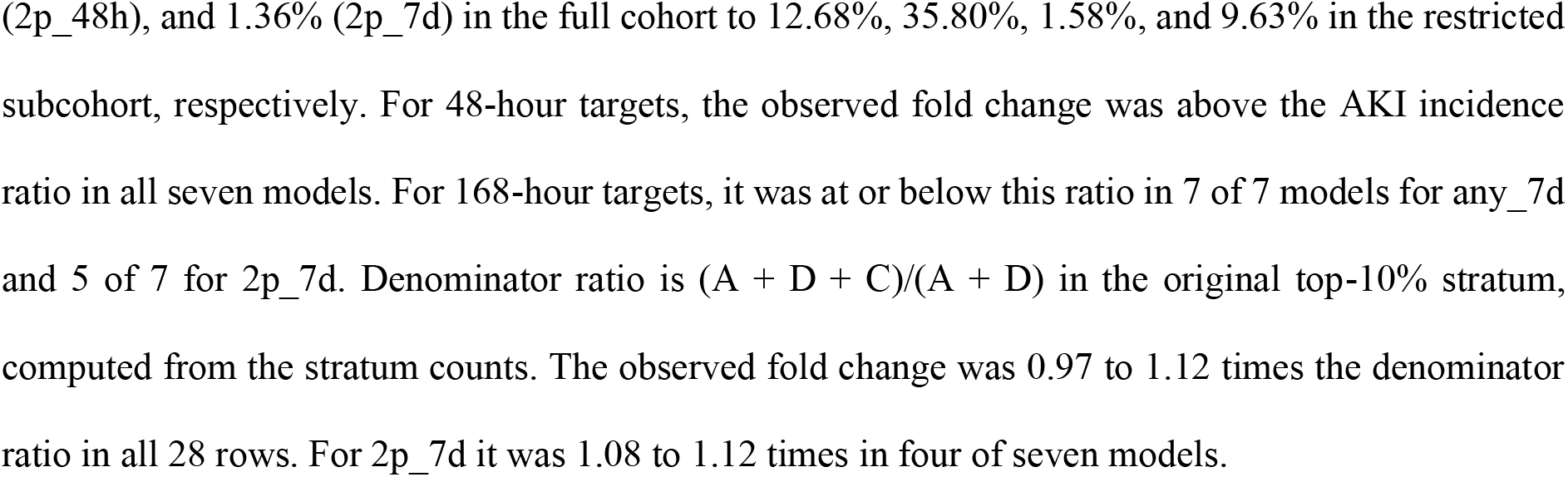
BRII fold change after removal of early-censored patients in eICU-CRD and two reference ratios.

| Outcome | Model | Type | BRII, full cohort | BRII, restricted subcohort | Observed fold change | AKI incidence ratio | Above incidence ratio | Denominator (A+D+C)/(A+D) |
| --- | --- | --- | --- | --- | --- | --- | --- | --- |
| any_48h | LR | binary | 0.210 | 0.779 | 3.71 | 2.60 | Yes | 3.72 |
| any_48h | XGB | binary | 0.185 | 0.788 | 4.26 | 2.60 | Yes | 4.24 |
| any_48h | LGBM | binary | 0.229 | 0.750 | 3.28 | 2.60 | Yes | 3.24 |
| any_48h | CS_CoxPH | CS | 0.251 | 0.764 | 3.04 | 2.60 | Yes | 2.96 |
| any_48h | CS_RSF | CS | 0.222 | 0.609 | 2.74 | 2.60 | Yes | 2.77 |
| any_48h | DeepHit | CIF | 0.216 | 0.630 | 2.91 | 2.60 | Yes | 2.93 |
| any_48h | FineGray | CIF | 0.246 | 0.787 | 3.20 | 2.60 | Yes | 3.13 |
| any_7d | LR | binary | 0.165 | 0.703 | 4.27 | 5.88 | No | 4.13 |
| any_7d | XGB | binary | 0.150 | 0.672 | 4.48 | 5.88 | No | 4.36 |
| any_7d | LGBM | binary | 0.172 | 0.645 | 3.76 | 5.88 | No | 3.74 |
| any_7d | CS_CoxPH | CS | 0.189 | 0.686 | 3.63 | 5.88 | No | 3.42 |
| any_7d | CS_RSF | CS | 0.158 | 0.553 | 3.49 | 5.88 | No | 3.41 |
| any_7d | DeepHit | CIF | 0.142 | 0.504 | 3.56 | 5.88 | No | 3.62 |
| any_7d | FineGray | CIF | 0.186 | 0.715 | 3.84 | 5.88 | No | 3.57 |
| 2p_48h | LR | binary | 0.032 | 0.348 | 10.95 | 2.68 | Yes | 10.19 |
| 2p_48h | XGB | binary | 0.015 | 0.256 | 17.53 | 2.68 | Yes | 17.14 |
| 2p_48h | LGBM | binary | 0.019 | 0.290 | 14.97 | 2.68 | Yes | 15.13 |
| 2p_48h | CS_CoxPH | CS | 0.049 | 0.243 | 4.93 | 2.68 | Yes | 4.93 |
| 2p_48h | CS_RSF | CS | 0.034 | 0.152 | 4.52 | 2.68 | Yes | 4.60 |
| 2p_48h | DeepHit | CIF | 0.038 | 0.169 | 4.48 | 2.68 | Yes | 4.38 |
| 2p_48h | FineGray | CIF | 0.045 | 0.278 | 6.17 | 2.68 | Yes | 5.90 |
| 2p_7d | LR | binary | 0.053 | 0.295 | 5.61 | 7.10 | No | 5.00 |
| 2p_7d | XGB | binary | 0.026 | 0.293 | 11.17 | 7.10 | Yes | 10.99 |
| 2p_7d | LGBM | binary | 0.032 | 0.278 | 8.81 | 7.10 | Yes | 8.88 |
| 2p_7d | CS_CoxPH | CS | 0.056 | 0.295 | 5.23 | 7.10 | No | 4.69 |
| 2p_7d | CS_RSF | CS | 0.043 | 0.189 | 4.37 | 7.10 | No | 4.03 |
| 2p_7d | DeepHit | CIF | 0.047 | 0.185 | 3.96 | 7.10 | No | 4.09 |
| 2p_7d | FineGray | CIF | 0.053 | 0.319 | 5.99 | 7.10 | No | 5.34 |

For each of seven prediction models and four outcomes, observed BRII at the top-10% threshold in the full eICU-CRD cohort and in the subcohort remaining after removal of patients censored before the prediction horizon. Observed fold change is the ratio of restricted-subcohort BRII to full-cohort BRII. AKI incidence ratio is the ratio of population-level AKI incidence in the restricted subcohort to that in the full cohort. Population-level AKI incidence rose from 4.88% (any\_48h), 6.09% (any\_7d), 0.59%
(2p\_48h), and 1.36% (2p\_7d) in the full cohort to 12.68%, 35.80%, 1.58%, and 9.63% in the restricted subcohort, respectively. For 48-hour targets, the observed fold change was above the AKI incidence ratio in all seven models. For 168-hour targets, it was at or below this ratio in 7 of 7 models for any\_7d and 5 of 7 for 2p\_7d. Denominator ratio is $(A + D + C)/(A + D)$ in the original top-10% stratum, computed from the stratum counts. The observed fold change was 0.97 to 1.12 times the denominator ratio in all 28 rows. For 2p\_7d it was 1.08 to 1.12 times in four of seven models.

**Supplementary Table 8.** BRII zone classification at top-10% threshold.

**[panel A. MIMIC-IV]**
| Eval | Model | Type | BRII | Zone |
| --- | --- | --- | --- | --- |
| 2p_48h | CS_CoxPH | CS | 0.3772 | C |
|  | CS_RSf | CS | 0.3079 | C |
|  | DeepHit | CIF | 0.2236 | D |
|  | FineGray | CIF | 0.4266 | C |
|  | LGBM | binary | 0.4825 | C |
|  | LR | binary | 0.5452 | B |
|  | XGB | binary | 0.4978 | C |
| 2p_7d | CS_CoxPH | CS | 0.2112 | D |
|  | CS_RSf | CS | 0.2114 | D |
|  | DeepHit | CIF | 0.1650 | D |
|  | FineGray | CIF | 0.2144 | D |
|  | LGBM | binary | 0.2097 | D |
|  | LR | binary | 0.2272 | D |
|  | XGB | binary | 0.1598 | D |
| any_48h | CS_CoxPH | CS | 0.9198 | A |
|  | CS_RSf | CS | 0.8808 | A |
|  | DeepHit | CIF | 0.9208 | A |
|  | FineGray | CIF | 0.9338 | A |
|  | LGBM | binary | 0.9616 | A |
|  | LR | binary | 0.9310 | A |
|  | XGB | binary | 0.9553 | A |
| any_7d | CS_CoxPH | CS | 0.6338 | B |
|  | CS_RSf | CS | 0.6003 | B |
|  | DeepHit | CIF | 0.5175 | B |
|  | FineGray | CIF | 0.6302 | B |
|  | LGBM | binary | 0.6113 | B |
|  | LR | binary | 0.6348 | B |
|  | XGB | binary | 0.5943 | B |

**[panel B. MIMIC-III]**
| Eval | Model | Type | BRII | Zone |
| --- | --- | --- | --- | --- |
| **2p_48h** | CS_CoxPH | CS | 0.4534 | C |
|  | CS_RSf | CS | 0.3081 | C |
|  | DeepHit | CIF | 0.1765 | D |
|  | FineGray | CIF | 0.4753 | C |
|  | LGBM | binary | 0.4425 | C |
|  | LR | binary | 0.5915 | B |
|  | XGB | binary | 0.5125 | B |
| **2p_7d** | CS_CoxPH | CS | 0.2303 | D |
|  | CS_RSf | CS | 0.2065 | D |
|  | DeepHit | CIF | 0.1405 | D |
|  | FineGray | CIF | 0.2283 | D |
|  | LGBM | binary | 0.1591 | D |
|  | LR | binary | 0.2388 | D |
|  | XGB | binary | 0.1308 | D |
| **any_48h** | CS_CoxPH | CS | 0.9218 | A |
|  | CS_RSf | CS | 0.8995 | A |
|  | DeepHit | CIF | 0.9212 | A |
|  | FineGray | CIF | 0.9339 | A |
|  | LGBM | binary | 0.9604 | A |
|  | LR | binary | 0.9231 | A |
|  | XGB | binary | 0.9594 | A |
| **any_7d** | CS_CoxPH | CS | 0.6282 | B |
|  | CS_RSf | CS | 0.6140 | B |
|  | DeepHit | CIF | 0.5659 | B |
|  | FineGray | CIF | 0.6262 | B |
|  | LGBM | binary | 0.6385 | B |
|  | LR | binary | 0.6256 | B |
|  | XGB | binary | 0.6041 | B |

**[panel C. eICU-CRD]**
| Eval | Model | Type | BRII | Zone |
| --- | --- | --- | --- | --- |
| **2p_48h** | CS_CoxPH | CS | 0.0493 | D |
|  | CS_RSf | CS | 0.0336 | D |
|  | DeepHit | CIF | 0.0377 | D |
|  | FineGray | CIF | 0.0450 | D |
|  | LGBM | binary | 0.0194 | D |
|  | LR | binary | 0.0318 | D |
|  | XGB | binary | 0.0146 | D |
| **2p_7d** | CS_CoxPH | CS | 0.0564 | D |
|  | CS_RSf | CS | 0.0432 | D |
|  | DeepHit | CIF | 0.0466 | D |
|  | FineGray | CIF | 0.0532 | D |
|  | LGBM | binary | 0.0315 | D |
|  | LR | binary | 0.0526 | D |
|  | XGB | binary | 0.0262 | D |
| **any_48h** | CS_CoxPH | CS | 0.2514 | D |
|  | CS_RSf | CS | 0.2223 | D |
|  | DeepHit | CIF | 0.2163 | D |
|  | FineGray | CIF | 0.2458 | D |
|  | LGBM | binary | 0.2286 | D |
|  | LR | binary | 0.2096 | D |
|  | XGB | binary | 0.1849 | D |
| **any_7d** | CS_CoxPH | CS | 0.1890 | D |
|  | CS_RSf | CS | 0.1582 | D |
|  | DeepHit | CIF | 0.1417 | D |
|  | FineGray | CIF | 0.1861 | D |
|  | LGBM | binary | 0.1716 | D |
|  | LR | binary | 0.1646 | D |
|  | XGB | binary | 0.1502 | D |

Each model-cohort-outcome combination classified into Zone A ( $\text{BRII} \geq 0.80$ ), Zone B (0.50-
0.80), Zone C (0.30-0.50), or Zone D ( $< 0.30$ ). Classification based on point-estimate BRII
from Tables 2 and 3.

**Supplementary Table 9.** BRII and performance metrics at 1%, 2%, and 5% top-*k* thresholds.

**[panel A. MIMIC-IV]**
| Eval | Model | Type | Top-k | BRII (95% CI) | AUROC | PPV | Sens | F1 | NB | AKI% | Death% | Cens% |
| --- | --- | --- | --- | --- | --- | --- | --- | --- | --- | --- | --- | --- |
| Any 48h | LR | binary | 1% | 0.891 (0.847-0.930) | 0.788 | 0.563 | 0.051 | 0.093 | -0.05988 | 56.3 | 3.9 | 3.0 |
|  | XGB | binary | 1% | 0.981 (0.961-0.995) | 0.758 | 0.611 | 0.055 | 0.101 | -0.03636 | 61.1 | 0.6 | 0.6 |
|  | LGBM | binary | 1% | 0.995 (0.984-1.000) | 0.795 | 0.614 | 0.056 | 0.102 | -0.02126 | 61.4 | 0.3 | 0.0 |
|  | CS_CoxPH | CS | 1% | 0.896 (0.855-0.934) | 0.784 | 0.569 | 0.051 | 0.094 | 0.00015 | 56.9 | 4.2 | 2.4 |
|  | CS_RSf | CS | 1% | 0.842 (0.798-0.886) | 0.774 | 0.560 | 0.051 | 0.093 | 0.00257 | 56.0 | 6.3 | 4.2 |
|  | DeepHit | CIF | 1% | 0.957 (0.925-0.984) | 0.747 | 0.539 | 0.049 | 0.089 | 0.00366 | 53.9 | 2.1 | 0.3 |
|  | FineGray | CIF | 1% | 0.910 (0.872-0.947) | 0.784 | 0.575 | 0.052 | 0.095 | 0.00062 | 57.5 | 3.3 | 2.4 |
|  | LR | binary | 2% | 0.914 (0.885-0.941) | 0.788 | 0.525 | 0.095 | 0.161 | -0.08266 | 52.5 | 3.0 | 1.9 |
|  | XGB | binary | 2% | 0.987 (0.975-0.997) | 0.758 | 0.565 | 0.102 | 0.173 | -0.05062 | 56.5 | 0.4 | 0.3 |
|  | LGBM | binary | 2% | 0.990 (0.978-0.998) | 0.795 | 0.577 | 0.105 | 0.177 | -0.03606 | 57.7 | 0.4 | 0.1 |
|  | CS_CoxPH | CS | 2% | 0.921 (0.895-0.948) | 0.784 | 0.526 | 0.095 | 0.161 | 0.00210 | 52.6 | 2.7 | 1.8 |
|  | CS_RSf | CS | 2% | 0.860 (0.826-0.892) | 0.774 | 0.513 | 0.093 | 0.157 | 0.00483 | 51.3 | 5.2 | 3.1 |
|  | DeepHit | CIF | 2% | 0.964 (0.946-0.982) | 0.747 | 0.528 | 0.096 | 0.162 | 0.00734 | 52.8 | 1.3 | 0.6 |
|  | FineGray | CIF | 2% | 0.937 (0.912-0.961) | 0.784 | 0.532 | 0.096 | 0.163 | 0.00263 | 53.2 | 2.2 | 1.3 |
|  | LR | binary | 5% | 0.934 (0.917-0.951) | 0.788 | 0.480 | 0.217 | 0.299 | -0.11162 | 48.0 | 1.8 | 1.6 |
|  | XGB | binary | 5% | 0.970 (0.957-0.982) | 0.758 | 0.467 | 0.211 | 0.291 | -0.07401 | 46.7 | 1.0 | 0.4 |
|  | LGBM | binary | 5% | 0.982 (0.972-0.990) | 0.795 | 0.511 | 0.231 | 0.318 | -0.06630 | 51.1 | 0.7 | 0.2 |
|  | CS_CoxPH | CS | 5% | 0.926 (0.910-0.943) | 0.784 | 0.475 | 0.215 | 0.296 | 0.01031 | 47.5 | 2.2 | 1.6 |
|  | CS_RSf | CS | 5% | 0.884 (0.863-0.905) | 0.774 | 0.437 | 0.198 | 0.273 | 0.01007 | 43.7 | 3.6 | 2.1 |
|  | DeepHit | CIF | 5% | 0.939 (0.924-0.956) | 0.747 | 0.436 | 0.197 | 0.272 | 0.01369 | 43.6 | 1.9 | 1.0 |
|  | FineGray | CIF | 5% | 0.940 (0.924-0.956) | 0.784 | 0.481 | 0.218 | 0.300 | 0.01128 | 48.1 | 1.6 | 1.5 |
| Any 7d | LR | binary | 1% | 0.723 (0.667-0.777) | 0.738 | 0.548 | 0.037 | 0.070 | -0.03994 | 54.8 | 6.3 | 14.7 |
|  | XGB | binary | 1% | 0.779 (0.722-0.830) | 0.717 | 0.611 | 0.042 | 0.078 | -0.03453 | 61.1 | 0.9 | 16.5 |
|  | LGBM | binary | 1% | 0.776 (0.728-0.826) | 0.754 | 0.644 | 0.044 | 0.082 | -0.01478 | 64.4 | 0.9 | 17.7 |
|  | CS_CoxPH | CS | 1% | 0.730 (0.676-0.787) | 0.733 | 0.584 | 0.040 | 0.074 | -0.00443 | 58.4 | 7.2 | 14.4 |
|  | CS_RSf | CS | 1% | 0.649 (0.591-0.705) | 0.738 | 0.503 | 0.034 | 0.064 | 0.00038 | 50.3 | 12.6 | 14.7 |
|  | DeepHit | CIF | 1% | 0.654 (0.594-0.709) | 0.704 | 0.527 | 0.036 | 0.067 | 0.00103 | 52.7 | 1.8 | 26.0 |
|  | FineGray | CIF | 1% | 0.743 (0.691-0.796) | 0.733 | 0.590 | 0.040 | 0.075 | -0.00331 | 59.0 | 5.1 | 15.3 |
|  | LR | binary | 2% | 0.732 (0.693-0.773) | 0.738 | 0.538 | 0.073 | 0.129 | -0.05727 | 53.8 | 4.2 | 15.5 |
|  | XGB | binary | 2% | 0.730 (0.694-0.769) | 0.717 | 0.558 | 0.076 | 0.134 | -0.05090 | 55.8 | 1.3 | 19.3 |
|  | LGBM | binary | 2% | 0.741 (0.704-0.777) | 0.754 | 0.599 | 0.082 | 0.144 | -0.02640 | 59.9 | 1.6 | 19.3 |
|  | CS_CoxPH | CS | 2% | 0.728 (0.690-0.767) | 0.733 | 0.553 | 0.075 | 0.133 | -0.00328 | 55.3 | 4.3 | 16.3 |
|  | CS_RSf | CS | 2% | 0.690 (0.648-0.730) | 0.738 | 0.535 | 0.073 | 0.128 | 0.00358 | 53.5 | 9.1 | 14.9 |
|  | DeepHit | CIF | 2% | 0.630 (0.584-0.668) | 0.704 | 0.502 | 0.068 | 0.121 | 0.00196 | 50.2 | 1.9 | 27.5 |
|  | FineGray | CIF | 2% | 0.726 (0.686-0.763) | 0.733 | 0.550 | 0.075 | 0.132 | -0.00264 | 55.0 | 3.9 | 16.9 |
|  | LR | binary | 5% | 0.688 (0.663-0.713) | 0.738 | 0.499 | 0.170 | 0.254 | -0.08288 | 49.9 | 4.3 | 18.4 |
|  | XGB | binary | 5% | 0.664 (0.638-0.689) | 0.717 | 0.483 | 0.165 | 0.246 | -0.07355 | 48.3 | 2.5 | 22.0 |
|  | LGBM | binary | 5% | 0.695 (0.670-0.719) | 0.754 | 0.532 | 0.181 | 0.271 | -0.05142 | 53.2 | 1.7 | 21.7 |
|  | CS_CoxPH | CS | 5% | 0.687 (0.663-0.714) | 0.733 | 0.510 | 0.174 | 0.259 | 0.00439 | 51.0 | 4.5 | 18.7 |
|  | CS_RSf | CS | 5% | 0.663 (0.637-0.688) | 0.738 | 0.483 | 0.165 | 0.246 | 0.00931 | 48.3 | 7.2 | 17.4 |
|  | DeepHit | CIF | 5% | 0.561 (0.533-0.586) | 0.704 | 0.439 | 0.150 | 0.223 | 0.00310 | 43.9 | 2.1 | 32.3 |
|  | FineGray | CIF | 5% | 0.692 (0.665-0.717) | 0.733 | 0.516 | 0.176 | 0.262 | 0.00615 | 51.6 | 3.5 | 19.6 |
| 2p 48h | LR | binary | 1% | 0.547 (0.421-0.677) | 0.781 | 0.105 | 0.086 | 0.095 | -0.09696 | 10.5 | 5.7 | 3.0 |
|  | XGB | binary | 1% | 0.810 (0.625-0.958) | 0.667 | 0.051 | 0.042 | 0.046 | -0.00291 | 5.1 | 1.2 | 0.0 |
|  | LGBM | binary | 1% | 0.586 (0.407-0.767) | 0.666 | 0.051 | 0.042 | 0.046 | -0.01503 | 5.1 | 1.2 | 2.4 |
|  | CS_CoxPH | CS | 1% | 0.398 (0.293-0.500) | 0.763 | 0.117 | 0.096 | 0.106 | 0.00051 | 11.7 | 12.3 | 5.4 |
|  | CS_RSf | CS | 1% | 0.214 (0.138-0.300) | 0.747 | 0.066 | 0.054 | 0.060 | 0.00002 | 6.6 | 13.5 | 10.8 |
|  | DeepHit | CIF | 1% | 0.186 (0.112-0.263) | 0.686 | 0.057 | 0.047 | 0.051 | -0.00023 | 5.7 | 17.4 | 7.5 |
|  | FineGray | CIF | 1% | 0.471 (0.372-0.575) | 0.770 | 0.123 | 0.101 | 0.111 | 0.00065 | 12.3 | 9.3 | 4.5 |
|  | LR | binary | 2% | 0.634 (0.542-0.732) | 0.781 | 0.088 | 0.146 | 0.110 | -0.13747 | 8.8 | 3.3 | 1.8 |
|  | XGB | binary | 2% | 0.689 (0.553-0.822) | 0.667 | 0.046 | 0.077 | 0.058 | -0.00280 | 4.6 | 0.7 | 1.3 |
|  | LGBM | binary | 2% | 0.603 (0.473-0.724) | 0.666 | 0.057 | 0.094 | 0.071 | -0.01811 | 5.7 | 1.6 | 2.1 |
|  | CS_CoxPH | CS | 2% | 0.406 (0.328-0.489) | 0.763 | 0.084 | 0.138 | 0.104 | 0.00068 | 8.4 | 8.7 | 3.6 |
|  | CS_RSf | CS | 2% | 0.189 (0.131-0.249) | 0.747 | 0.046 | 0.077 | 0.058 | 0.00002 | 4.6 | 12.3 | 7.6 |
|  | DeepHit | CIF | 2% | 0.200 (0.144-0.268) | 0.686 | 0.051 | 0.084 | 0.063 | -0.00026 | 5.1 | 14.1 | 6.3 |
|  | FineGray | CIF | 2% | 0.468 (0.388-0.552) | 0.770 | 0.088 | 0.146 | 0.110 | 0.00086 | 8.8 | 7.0 | 3.0 |
|  | LR | binary | 5% | 0.591 (0.521-0.659) | 0.781 | 0.066 | 0.272 | 0.106 | -0.19077 | 6.6 | 3.2 | 1.4 |
|  | XGB | binary | 5% | 0.590 (0.500-0.675) | 0.667 | 0.043 | 0.178 | 0.069 | -0.00152 | 4.3 | 0.7 | 2.3 |
|  | LGBM | binary | 5% | 0.532 (0.448-0.621) | 0.666 | 0.039 | 0.163 | 0.064 | -0.02235 | 3.9 | 1.4 | 2.1 |
|  | CS_CoxPH | CS | 5% | 0.403 (0.342-0.466) | 0.763 | 0.059 | 0.242 | 0.094 | 0.00128 | 5.9 | 6.5 | 2.2 |
|  | CS_RSf | CS | 5% | 0.241 (0.194-0.291) | 0.747 | 0.042 | 0.175 | 0.068 | 0.00063 | 4.2 | 9.2 | 4.2 |
|  | DeepHit | CIF | 5% | 0.236 (0.191-0.286) | 0.686 | 0.044 | 0.180 | 0.070 | -0.00011 | 4.4 | 10.5 | 3.6 |
|  | FineGray | CIF | 5% | 0.486 (0.421-0.552) | 0.770 | 0.062 | 0.254 | 0.099 | 0.00152 | 6.2 | 4.7 | 1.8 |
| 2p 7d | LR | binary | 1% | 0.306 (0.234-0.376) | 0.714 | 0.156 | 0.050 | 0.076 | -0.05858 | 15.6 | 22.8 | 12.6 |
|  | XGB | binary | 1% | 0.247 (0.181-0.318) | 0.625 | 0.111 | 0.036 | 0.054 | -0.01398 | 11.1 | 3.9 | 29.9 |
|  | LGBM | binary | 1% | 0.324 (0.254-0.407) | 0.681 | 0.135 | 0.043 | 0.065 | -0.02349 | 13.5 | 6.9 | 21.3 |
|  | CS_CoxPH | CS | 1% | 0.349 (0.276-0.424) | 0.709 | 0.183 | 0.059 | 0.089 | -0.00017 | 18.3 | 21.0 | 13.2 |
|  | CS_RSf | CS | 1% | 0.263 (0.200-0.328) | 0.709 | 0.153 | 0.049 | 0.074 | -0.00042 | 15.3 | 22.2 | 20.7 |
|  | DeepHit | CIF | 1% | 0.158 (0.116-0.208) | 0.663 | 0.096 | 0.031 | 0.047 | -0.00169 | 9.6 | 21.0 | 30.2 |
|  | FineGray | CIF | 1% | 0.371 (0.297-0.449) | 0.709 | 0.177 | 0.057 | 0.086 | 0.00003 | 17.7 | 16.5 | 13.5 |
|  | LR | binary | 2% | 0.320 (0.268-0.368) | 0.714 | 0.148 | 0.095 | 0.116 | -0.08397 | 14.8 | 17.0 | 14.3 |
|  | XGB | binary | 2% | 0.207 (0.161-0.254) | 0.625 | 0.091 | 0.059 | 0.071 | -0.01828 | 9.1 | 4.5 | 30.3 |
|  | LGBM | binary | 2% | 0.321 (0.270-0.374) | 0.681 | 0.133 | 0.085 | 0.104 | -0.03601 | 13.3 | 5.8 | 22.3 |
|  | CS_CoxPH | CS | 2% | 0.342 (0.290-0.392) | 0.709 | 0.158 | 0.102 | 0.124 | 0.00022 | 15.8 | 16.3 | 14.2 |
|  | CS_RSf | CS | 2% | 0.240 (0.196-0.285) | 0.709 | 0.123 | 0.079 | 0.096 | -0.00025 | 12.3 | 21.4 | 17.5 |
|  | DeepHit | CIF | 2% | 0.150 (0.113-0.185) | 0.663 | 0.085 | 0.055 | 0.067 | -0.00227 | 8.5 | 21.7 | 26.6 |
|  | FineGray | CIF | 2% | 0.353 (0.301-0.410) | 0.709 | 0.158 | 0.102 | 0.124 | 0.00055 | 15.8 | 13.9 | 15.1 |
|  | LR | binary | 5% | 0.280 (0.250-0.311) | 0.714 | 0.122 | 0.196 | 0.150 | -0.13133 | 12.2 | 12.9 | 18.5 |
|  | XGB | binary | 5% | 0.173 (0.147-0.201) | 0.625 | 0.077 | 0.124 | 0.095 | -0.02297 | 7.7 | 4.1 | 32.7 |
|  | LGBM | binary | 5% | 0.243 (0.213-0.274) | 0.681 | 0.106 | 0.170 | 0.130 | -0.05920 | 10.6 | 6.0 | 26.9 |
|  | CS_CoxPH | CS | 5% | 0.266 (0.236-0.298) | 0.709 | 0.119 | 0.191 | 0.147 | 0.00104 | 11.9 | 13.0 | 19.8 |
|  | CS_RSf | CS | 5% | 0.234 (0.203-0.262) | 0.709 | 0.108 | 0.173 | 0.133 | 0.00127 | 10.8 | 19.1 | 16.2 |
|  | DeepHit | CIF | 5% | 0.162 (0.137-0.187) | 0.663 | 0.081 | 0.131 | 0.100 | -0.00296 | 8.1 | 21.7 | 20.4 |
|  | FineGray | CIF | 5% | 0.282 (0.250-0.315) | 0.709 | 0.122 | 0.196 | 0.150 | 0.00160 | 12.2 | 10.1 | 20.9 |

[panel B. MIMIC-III]
| Eval | Model | Type | Top-k | BRII (95% CI) | AUROC | PPV | Sens | F1 | NB | AKI% | Death% | Cens% |
| --- | --- | --- | --- | --- | --- | --- | --- | --- | --- | --- | --- | --- |
| Any 48h | LR | binary | 1% | 0.910 (0.847-0.966) | 0.773 | 0.559 | 0.049 | 0.090 | -0.04916 | 55.9 | 3.4 | 2.1 |
|  | XGB | binary | 1% | 0.978 (0.946-1.000) | 0.759 | 0.628 | 0.055 | 0.101 | -0.03551 | 62.8 | 1.4 | 0.0 |
|  | LGBM | binary | 1% | 1.000 (1.000-1.000) | 0.790 | 0.697 | 0.061 | 0.112 | -0.01362 | 69.7 | 0.0 | 0.0 |
|  | CS_CoxPH | CS | 1% | 0.916 (0.854-0.968) | 0.768 | 0.600 | 0.052 | 0.096 | 0.00177 | 60.0 | 3.4 | 2.1 |
|  | CS_RSf | CS | 1% | 0.915 (0.855-0.974) | 0.751 | 0.517 | 0.045 | 0.083 | 0.00192 | 51.7 | 2.1 | 2.8 |
|  | DeepHit | CIF | 1% | 0.975 (0.938-1.000) | 0.744 | 0.531 | 0.046 | 0.085 | 0.00372 | 53.1 | 0.7 | 0.7 |
|  | FineGray | CIF | 1% | 0.935 (0.881-0.979) | 0.768 | 0.593 | 0.052 | 0.095 | 0.00169 | 59.3 | 2.8 | 1.4 |
|  | LR | binary | 2% | 0.936 (0.897-0.971) | 0.773 | 0.559 | 0.097 | 0.166 | -0.06235 | 55.9 | 2.4 | 1.4 |
|  | XGB | binary | 2% | 0.977 (0.950-0.994) | 0.759 | 0.586 | 0.102 | 0.174 | -0.04989 | 58.6 | 1.0 | 0.3 |
|  | LGBM | binary | 2% | 1.000 (1.000-1.000) | 0.790 | 0.614 | 0.107 | 0.182 | -0.03041 | 61.4 | 0.0 | 0.0 |
|  | CS_CoxPH | CS | 2% | 0.927 (0.886-0.964) | 0.768 | 0.566 | 0.099 | 0.168 | 0.00471 | 56.6 | 2.8 | 1.7 |
|  | CS_RSf | CS | 2% | 0.919 (0.876-0.961) | 0.751 | 0.510 | 0.089 | 0.152 | 0.00454 | 51.0 | 3.1 | 1.4 |
|  | DeepHit | CIF | 2% | 0.976 (0.953-0.994) | 0.744 | 0.552 | 0.096 | 0.164 | 0.00840 | 55.2 | 0.7 | 0.7 |
|  | FineGray | CIF | 2% | 0.943 (0.908-0.976) | 0.768 | 0.572 | 0.100 | 0.170 | 0.00498 | 57.2 | 2.1 | 1.4 |
|  | LR | binary | 5% | 0.930 (0.905-0.954) | 0.773 | 0.492 | 0.215 | 0.299 | -0.09275 | 49.2 | 2.3 | 1.4 |
|  | XGB | binary | 5% | 0.966 (0.946-0.983) | 0.759 | 0.468 | 0.204 | 0.284 | -0.07798 | 46.8 | 1.1 | 0.6 |
|  | LGBM | binary | 5% | 0.976 (0.960-0.990) | 0.790 | 0.499 | 0.218 | 0.303 | -0.06678 | 49.9 | 0.6 | 0.7 |
|  | CS_CoxPH | CS | 5% | 0.926 (0.900-0.952) | 0.768 | 0.484 | 0.211 | 0.294 | 0.01269 | 48.4 | 2.1 | 1.8 |
|  | CS_RSf | CS | 5% | 0.903 (0.872-0.933) | 0.751 | 0.434 | 0.189 | 0.264 | 0.00926 | 43.4 | 2.8 | 1.9 |
|  | DeepHit | CIF | 5% | 0.949 (0.924-0.972) | 0.744 | 0.436 | 0.190 | 0.265 | 0.01520 | 43.6 | 1.4 | 1.0 |
|  | FineGray | CIF | 5% | 0.947 (0.923-0.968) | 0.768 | 0.492 | 0.215 | 0.299 | 0.01334 | 49.2 | 1.5 | 1.2 |
| Any 7d | LR | binary | 1% | 0.735 (0.656-0.815) | 0.723 | 0.593 | 0.037 | 0.069 | -0.03141 | 59.3 | 8.3 | 13.1 |
|  | XGB | binary | 1% | 0.815 (0.748-0.881) | 0.724 | 0.697 | 0.043 | 0.081 | -0.02447 | 69.7 | 4.1 | 11.7 |
|  | LGBM | binary | 1% | 0.816 (0.752-0.884) | 0.753 | 0.703 | 0.044 | 0.082 | -0.00971 | 70.3 | 2.1 | 13.8 |
|  | CS_CoxPH | CS | 1% | 0.777 (0.698-0.850) | 0.722 | 0.648 | 0.040 | 0.076 | -0.00040 | 64.8 | 6.9 | 11.7 |
|  | CS_RSf | CS | 1% | 0.761 (0.686-0.832) | 0.726 | 0.614 | 0.038 | 0.072 | 0.00248 | 61.4 | 3.4 | 15.9 |
|  | DeepHit | CIF | 1% | 0.632 (0.545-0.711) | 0.708 | 0.545 | 0.034 | 0.064 | 0.00250 | 54.5 | 3.4 | 28.3 |
|  | FineGray | CIF | 1% | 0.770 (0.689-0.840) | 0.721 | 0.648 | 0.040 | 0.076 | -0.00017 | 64.8 | 5.5 | 13.8 |
|  | LR | binary | 2% | 0.701 (0.636-0.759) | 0.723 | 0.566 | 0.070 | 0.125 | -0.04588 | 56.6 | 6.6 | 17.6 |
|  | XGB | binary | 2% | 0.749 (0.692-0.803) | 0.724 | 0.628 | 0.078 | 0.139 | -0.04170 | 62.8 | 3.4 | 17.6 |
|  | LGBM | binary | 2% | 0.784 (0.733-0.838) | 0.753 | 0.662 | 0.082 | 0.146 | -0.01827 | 66.2 | 2.1 | 16.2 |
|  | CS_CoxPH | CS | 2% | 0.735 (0.678-0.790) | 0.722 | 0.621 | 0.077 | 0.137 | 0.00225 | 62.1 | 6.9 | 15.5 |
|  | CS_RSf | CS | 2% | 0.733 (0.678-0.790) | 0.726 | 0.586 | 0.073 | 0.130 | 0.00524 | 58.6 | 4.5 | 16.9 |
|  | DeepHit | CIF | 2% | 0.642 (0.581-0.700) | 0.708 | 0.538 | 0.067 | 0.119 | 0.00562 | 53.8 | 4.5 | 25.5 |
|  | FineGray | CIF | 2% | 0.732 (0.672-0.784) | 0.721 | 0.621 | 0.077 | 0.137 | 0.00246 | 62.1 | 5.2 | 17.6 |
|  | LR | binary | 5% | 0.670 (0.630-0.707) | 0.723 | 0.528 | 0.164 | 0.250 | -0.06810 | 52.8 | 4.4 | 21.7 |
|  | XGB | binary | 5% | 0.665 (0.624-0.704) | 0.724 | 0.531 | 0.165 | 0.252 | -0.06603 | 53.1 | 3.0 | 23.7 |
|  | LGBM | binary | 5% | 0.684 (0.644-0.723) | 0.753 | 0.550 | 0.171 | 0.261 | -0.04996 | 55.0 | 1.8 | 23.6 |
|  | CS_CoxPH | CS | 5% | 0.671 (0.631-0.707) | 0.722 | 0.539 | 0.167 | 0.256 | 0.00986 | 53.9 | 4.4 | 22.1 |
|  | CS_RSf | CS | 5% | 0.690 (0.650-0.730) | 0.726 | 0.510 | 0.158 | 0.242 | 0.01048 | 51.0 | 5.7 | 17.2 |
|  | DeepHit | CIF | 5% | 0.591 (0.551-0.628) | 0.708 | 0.480 | 0.149 | 0.227 | 0.01285 | 48.0 | 4.1 | 29.1 |
|  | FineGray | CIF | 5% | 0.683 (0.646-0.720) | 0.721 | 0.550 | 0.171 | 0.261 | 0.01114 | 55.0 | 3.6 | 21.9 |
| 2p 48h | LR | binary | 1% | 0.586 (0.407-0.778) | 0.790 | 0.117 | 0.102 | 0.109 | -0.08609 | 11.7 | 5.5 | 2.8 |
|  | XGB | binary | 1% | 1.000 (1.000-1.000) | 0.634 | 0.062 | 0.054 | 0.058 | -0.00388 | 6.2 | 0.0 | 0.0 |
|  | LGBM | binary | 1% | 0.571 (0.286-0.833) | 0.685 | 0.055 | 0.048 | 0.051 | -0.01778 | 5.5 | 0.0 | 4.1 |
|  | CS_CoxPH | CS | 1% | 0.425 (0.275-0.571) | 0.790 | 0.117 | 0.102 | 0.109 | 0.00052 | 11.7 | 11.7 | 4.1 |
|  | CS_RSf | CS | 1% | 0.364 (0.227-0.513) | 0.729 | 0.110 | 0.096 | 0.103 | 0.00046 | 11.0 | 11.7 | 7.6 |
|  | DeepHit | CIF | 1% | 0.088 (0.000-0.189) | 0.682 | 0.021 | 0.018 | 0.019 | -0.00046 | 2.1 | 12.4 | 9.0 |
|  | FineGray | CIF | 1% | 0.514 (0.351-0.675) | 0.794 | 0.124 | 0.108 | 0.116 | 0.00068 | 12.4 | 8.3 | 3.4 |
|  | LR | binary | 2% | 0.630 (0.488-0.780) | 0.790 | 0.100 | 0.175 | 0.127 | -0.11839 | 10.0 | 3.8 | 2.1 |
|  | XGB | binary | 2% | 0.824 (0.600-1.000) | 0.634 | 0.048 | 0.084 | 0.061 | -0.00415 | 4.8 | 0.7 | 0.3 |
|  | LGBM | binary | 2% | 0.571 (0.400-0.759) | 0.685 | 0.055 | 0.096 | 0.070 | -0.02295 | 5.5 | 0.3 | 3.8 |
|  | CS_CoxPH | CS | 2% | 0.393 (0.277-0.525) | 0.790 | 0.083 | 0.145 | 0.105 | 0.00073 | 8.3 | 8.3 | 4.5 |
|  | CS_RSf | CS | 2% | 0.333 (0.224-0.467) | 0.729 | 0.072 | 0.127 | 0.092 | 0.00052 | 7.2 | 8.3 | 6.2 |
|  | DeepHit | CIF | 2% | 0.091 (0.020-0.178) | 0.682 | 0.017 | 0.030 | 0.022 | -0.00070 | 1.7 | 11.4 | 5.9 |
|  | FineGray | CIF | 2% | 0.500 (0.365-0.633) | 0.794 | 0.090 | 0.157 | 0.114 | 0.00093 | 9.0 | 6.9 | 2.1 |
|  | LR | binary | 5% | 0.622 (0.523-0.722) | 0.790 | 0.070 | 0.307 | 0.114 | -0.16960 | 7.0 | 2.3 | 1.9 |
|  | XGB | binary | 5% | 0.594 (0.419-0.778) | 0.634 | 0.026 | 0.114 | 0.043 | -0.00417 | 2.6 | 0.7 | 1.1 |
|  | LGBM | binary | 5% | 0.500 (0.378-0.627) | 0.685 | 0.043 | 0.187 | 0.070 | -0.02907 | 4.3 | 0.6 | 3.7 |
|  | CS_CoxPH | CS | 5% | 0.440 (0.346-0.530) | 0.790 | 0.066 | 0.289 | 0.108 | 0.00176 | 6.6 | 5.8 | 2.6 |
|  | CS_RSf | CS | 5% | 0.311 (0.225-0.402) | 0.729 | 0.044 | 0.193 | 0.072 | 0.00067 | 4.4 | 5.8 | 4.0 |
|  | DeepHit | CIF | 5% | 0.165 (0.105-0.237) | 0.682 | 0.026 | 0.114 | 0.043 | -0.00058 | 2.6 | 9.5 | 3.7 |
|  | FineGray | CIF | 5% | 0.459 (0.367-0.545) | 0.794 | 0.069 | 0.301 | 0.112 | 0.00196 | 6.9 | 5.4 | 2.8 |
| 2p 7d | LR | binary | 1% | 0.359 (0.260-0.463) | 0.710 | 0.193 | 0.055 | 0.086 | -0.05493 | 19.3 | 18.6 | 15.9 |
|  | XGB | binary | 1% | 0.169 (0.095-0.263) | 0.621 | 0.090 | 0.026 | 0.040 | -0.01634 | 9.0 | 6.2 | 37.9 |
|  | LGBM | binary | 1% | 0.324 (0.217-0.440) | 0.681 | 0.152 | 0.043 | 0.068 | -0.02301 | 15.2 | 4.8 | 26.9 |
|  | CS_CoxPH | CS | 1% | 0.329 (0.227-0.437) | 0.709 | 0.186 | 0.053 | 0.083 | -0.00011 | 18.6 | 20.7 | 17.2 |
|  | CS_RSf | CS | 1% | 0.416 (0.313-0.517) | 0.690 | 0.255 | 0.073 | 0.114 | 0.00078 | 25.5 | 17.9 | 17.9 |
|  | DeepHit | CIF | 1% | 0.145 (0.068-0.236) | 0.649 | 0.069 | 0.020 | 0.031 | -0.00112 | 6.9 | 19.3 | 21.4 |
|  | FineGray | CIF | 1% | 0.341 (0.241-0.437) | 0.712 | 0.193 | 0.055 | 0.086 | 0.00028 | 19.3 | 16.6 | 20.7 |
|  | LR | binary | 2% | 0.294 (0.220-0.367) | 0.710 | 0.145 | 0.083 | 0.106 | -0.08185 | 14.5 | 15.9 | 19.0 |
|  | XGB | binary | 2% | 0.159 (0.101-0.221) | 0.621 | 0.079 | 0.045 | 0.058 | -0.02271 | 7.9 | 4.5 | 37.6 |
|  | LGBM | binary | 2% | 0.253 (0.187-0.326) | 0.681 | 0.128 | 0.073 | 0.093 | -0.03711 | 12.8 | 5.5 | 32.1 |
|  | CS_CoxPH | CS | 2% | 0.296 (0.222-0.369) | 0.709 | 0.155 | 0.089 | 0.113 | 0.00036 | 15.5 | 14.5 | 22.4 |
|  | CS_RSf | CS | 2% | 0.333 (0.264-0.405) | 0.690 | 0.179 | 0.103 | 0.131 | 0.00087 | 17.9 | 15.5 | 20.3 |
|  | DeepHit | CIF | 2% | 0.162 (0.103-0.223) | 0.649 | 0.079 | 0.045 | 0.058 | -0.00123 | 7.9 | 21.7 | 19.3 |
|  | FineGray | CIF | 2% | 0.304 (0.227-0.380) | 0.712 | 0.155 | 0.089 | 0.113 | 0.00061 | 15.5 | 12.4 | 23.1 |
|  | LR | binary | 5% | 0.268 (0.222-0.311) | 0.710 | 0.131 | 0.188 | 0.154 | -0.12634 | 13.1 | 12.8 | 22.9 |
|  | XGB | binary | 5% | 0.153 (0.118-0.190) | 0.621 | 0.077 | 0.111 | 0.091 | -0.02940 | 7.7 | 4.1 | 38.5 |
|  | LGBM | binary | 5% | 0.183 (0.146-0.223) | 0.681 | 0.098 | 0.140 | 0.115 | -0.06712 | 9.8 | 4.8 | 38.8 |
|  | CS_CoxPH | CS | 5% | 0.267 (0.219-0.313) | 0.709 | 0.131 | 0.188 | 0.154 | 0.00198 | 13.1 | 12.4 | 23.6 |
|  | CS_RSf | CS | 5% | 0.235 (0.191-0.280) | 0.690 | 0.110 | 0.158 | 0.130 | 0.00109 | 11.0 | 16.1 | 19.7 |
|  | DeepHit | CIF | 5% | 0.150 (0.115-0.188) | 0.649 | 0.069 | 0.099 | 0.081 | -0.00187 | 6.9 | 21.2 | 17.9 |
|  | FineGray | CIF | 5% | 0.266 (0.219-0.310) | 0.712 | 0.134 | 0.192 | 0.158 | 0.00242 | 13.4 | 10.8 | 26.2 |

[panel C. eICU-CRD]
| Eval | Model | Type | Top-k | BRII (95% CI) | AUROC | PPV | Sens | F1 | NB | AKI% | Death% | Cens% |
| --- | --- | --- | --- | --- | --- | --- | --- | --- | --- | --- | --- | --- |
| Any 48h | LR | binary | 1% | 0.322 (0.278-0.364) | 0.685 | 0.200 | 0.041 | 0.068 | -0.15153 | 20.0 | 6.0 | 36.2 |
|  | XGB | binary | 1% | 0.335 (0.293-0.376) | 0.656 | 0.221 | 0.045 | 0.075 | -0.02693 | 22.1 | 4.4 | 39.4 |
|  | LGBM | binary | 1% | 0.340 (0.298-0.385) | 0.720 | 0.219 | 0.045 | 0.074 | -0.02407 | 21.9 | 7.4 | 35.1 |
|  | CS_CoxPH | CS | 1% | 0.447 (0.400-0.493) | 0.713 | 0.290 | 0.059 | 0.099 | -0.00210 | 29.0 | 8.8 | 27.1 |
|  | CS_RSf | CS | 1% | 0.378 (0.329-0.425) | 0.715 | 0.235 | 0.048 | 0.080 | -0.00191 | 23.5 | 13.8 | 24.9 |
|  | DeepHit | CIF | 1% | 0.276 (0.233-0.321) | 0.672 | 0.169 | 0.035 | 0.058 | -0.00053 | 16.9 | 10.4 | 33.8 |
|  | FineGray | CIF | 1% | 0.442 (0.396-0.488) | 0.708 | 0.287 | 0.059 | 0.098 | -0.00171 | 28.7 | 7.1 | 29.1 |
|  | LR | binary | 2% | 0.299 (0.271-0.330) | 0.685 | 0.188 | 0.077 | 0.109 | -0.16129 | 18.8 | 4.9 | 39.1 |
|  | XGB | binary | 2% | 0.292 (0.264-0.320) | 0.656 | 0.194 | 0.080 | 0.113 | -0.03550 | 19.4 | 4.0 | 43.1 |
|  | LGBM | binary | 2% | 0.307 (0.280-0.336) | 0.720 | 0.202 | 0.083 | 0.118 | -0.03749 | 20.2 | 6.8 | 38.9 |
|  | CS_CoxPH | CS | 2% | 0.409 (0.376-0.443) | 0.713 | 0.255 | 0.105 | 0.148 | -0.00200 | 25.5 | 7.5 | 29.3 |
|  | CS_RSF | CS | 2% | 0.334 (0.302-0.367) | 0.715 | 0.201 | 0.083 | 0.117 | -0.00286 | 20.1 | 11.9 | 28.3 |
|  | DeepHit | CIF | 2% | 0.262 (0.233-0.292) | 0.672 | 0.161 | 0.066 | 0.094 | -0.00061 | 16.1 | 10.7 | 34.6 |
|  | FineGray | CIF | 2% | 0.394 (0.362-0.427) | 0.708 | 0.246 | 0.101 | 0.143 | -0.00161 | 24.6 | 6.3 | 31.6 |
|  | LR | binary | 5% | 0.245 (0.228-0.263) | 0.685 | 0.157 | 0.161 | 0.159 | -0.18032 | 15.7 | 4.5 | 43.9 |
|  | XGB | binary | 5% | 0.235 (0.218-0.253) | 0.656 | 0.156 | 0.160 | 0.158 | -0.04737 | 15.6 | 3.9 | 47.0 |
|  | LGBM | binary | 5% | 0.271 (0.253-0.290) | 0.720 | 0.179 | 0.183 | 0.181 | -0.06326 | 17.9 | 5.8 | 42.3 |
|  | CS_CoxPH | CS | 5% | 0.331 (0.311-0.352) | 0.713 | 0.203 | 0.208 | 0.205 | -0.00113 | 20.3 | 6.3 | 34.6 |
|  | CS_RSF | CS | 5% | 0.272 (0.253-0.291) | 0.715 | 0.158 | 0.162 | 0.160 | -0.00403 | 15.8 | 10.1 | 32.2 |
|  | DeepHit | CIF | 5% | 0.248 (0.228-0.266) | 0.672 | 0.146 | 0.150 | 0.148 | -0.00047 | 14.6 | 8.8 | 35.5 |
|  | FineGray | CIF | 5% | 0.320 (0.301-0.340) | 0.708 | 0.198 | 0.203 | 0.200 | -0.00071 | 19.8 | 5.5 | 36.6 |
| Any 7d | LR | binary | 1% | 0.248 (0.214-0.285) | 0.660 | 0.225 | 0.037 | 0.063 | -0.09775 | 22.5 | 8.7 | 59.6 |
|  | XGB | binary | 1% | 0.218 (0.186-0.252) | 0.645 | 0.200 | 0.033 | 0.056 | -0.03975 | 20.0 | 7.4 | 64.6 |
|  | LGBM | binary | 1% | 0.244 (0.210-0.280) | 0.701 | 0.222 | 0.036 | 0.063 | -0.02476 | 22.2 | 9.9 | 59.0 |
|  | CS_CoxPH | CS | 1% | 0.342 (0.306-0.379) | 0.688 | 0.306 | 0.050 | 0.086 | -0.00548 | 30.6 | 13.8 | 45.1 |
|  | CS_RSF | CS | 1% | 0.261 (0.224-0.294) | 0.697 | 0.229 | 0.038 | 0.065 | -0.00417 | 22.9 | 18.4 | 46.6 |
|  | DeepHit | CIF | 1% | 0.152 (0.126-0.181) | 0.644 | 0.138 | 0.023 | 0.039 | -0.00371 | 13.8 | 19.4 | 57.6 |
|  | FineGray | CIF | 1% | 0.336 (0.299-0.374) | 0.682 | 0.301 | 0.049 | 0.085 | -0.00461 | 30.1 | 11.6 | 47.9 |
|  | LR | binary | 2% | 0.216 (0.193-0.239) | 0.660 | 0.196 | 0.064 | 0.097 | -0.12033 | 19.6 | 7.9 | 63.3 |
|  | XGB | binary | 2% | 0.199 (0.177-0.220) | 0.645 | 0.184 | 0.060 | 0.091 | -0.05217 | 18.4 | 7.5 | 66.6 |
|  | LGBM | binary | 2% | 0.226 (0.205-0.250) | 0.701 | 0.207 | 0.068 | 0.102 | -0.04090 | 20.7 | 10.0 | 60.9 |
|  | CS_CoxPH | CS | 2% | 0.306 (0.280-0.331) | 0.688 | 0.274 | 0.090 | 0.136 | -0.00609 | 27.4 | 11.8 | 50.5 |
|  | CS_RSF | CS | 2% | 0.241 (0.216-0.266) | 0.697 | 0.212 | 0.070 | 0.105 | -0.00628 | 21.2 | 15.7 | 51.1 |
|  | DeepHit | CIF | 2% | 0.176 (0.156-0.197) | 0.644 | 0.160 | 0.052 | 0.079 | -0.00461 | 16.0 | 17.4 | 57.3 |
|  | FineGray | CIF | 2% | 0.292 (0.268-0.317) | 0.682 | 0.264 | 0.087 | 0.130 | -0.00516 | 26.4 | 10.3 | 53.6 |
|  | LR | binary | 5% | 0.188 (0.174-0.202) | 0.660 | 0.171 | 0.140 | 0.154 | -0.15118 | 17.1 | 7.3 | 66.8 |
|  | XGB | binary | 5% | 0.177 (0.163-0.190) | 0.645 | 0.162 | 0.133 | 0.146 | -0.07031 | 16.2 | 7.9 | 67.5 |
|  | LGBM | binary | 5% | 0.186 (0.172-0.200) | 0.701 | 0.170 | 0.139 | 0.153 | -0.07472 | 17.0 | 9.3 | 65.1 |
|  | CS_CoxPH | CS | 5% | 0.243 (0.229-0.258) | 0.688 | 0.220 | 0.180 | 0.198 | -0.00672 | 22.0 | 10.7 | 57.6 |
|  | CS_RSF | CS | 5% | 0.181 (0.167-0.195) | 0.697 | 0.163 | 0.134 | 0.147 | -0.01141 | 16.3 | 13.4 | 60.4 |
|  | DeepHit | CIF | 5% | 0.165 (0.152-0.178) | 0.644 | 0.149 | 0.122 | 0.134 | -0.00762 | 14.9 | 13.9 | 61.3 |
|  | FineGray | CIF | 5% | 0.237 (0.221-0.251) | 0.682 | 0.214 | 0.176 | 0.193 | -0.00570 | 21.4 | 9.3 | 59.8 |
| 2p 48h | LR | binary | 1% | 0.047 (0.026-0.070) | 0.693 | 0.026 | 0.045 | 0.033 | -0.19407 | 2.6 | 6.0 | 47.4 |
|  | XGB | binary | 1% | 0.030 (0.015-0.048) | 0.565 | 0.018 | 0.030 | 0.022 | -0.00134 | 1.8 | 4.0 | 52.4 |
|  | LGBM | binary | 1% | 0.039 (0.021-0.058) | 0.624 | 0.024 | 0.040 | 0.030 | -0.01225 | 2.4 | 2.8 | 55.3 |
|  | CS_CoxPH | CS | 1% | 0.113 (0.079-0.148) | 0.747 | 0.051 | 0.087 | 0.065 | -0.00006 | 5.1 | 16.5 | 24.0 |
|  | CS_RSF | CS | 1% | 0.056 (0.034-0.082) | 0.701 | 0.029 | 0.050 | 0.037 | -0.00051 | 2.9 | 14.6 | 34.7 |
|  | DeepHit | CIF | 1% | 0.059 (0.034-0.085) | 0.701 | 0.031 | 0.052 | 0.039 | -0.00037 | 3.1 | 18.7 | 30.3 |
|  | FineGray | CIF | 1% | 0.111 (0.076-0.148) | 0.741 | 0.051 | 0.087 | 0.065 | 0.00002 | 5.1 | 12.9 | 28.2 |
|  | LR | binary | 2% | 0.041 (0.028-0.056) | 0.693 | 0.023 | 0.077 | 0.035 | -0.19196 | 2.3 | 5.1 | 47.7 |
|  | XGB | binary | 2% | 0.028 (0.017-0.041) | 0.565 | 0.016 | 0.055 | 0.025 | -0.00137 | 1.6 | 2.9 | 52.8 |
|  | LGBM | binary | 2% | 0.033 (0.021-0.046) | 0.624 | 0.019 | 0.065 | 0.030 | -0.01482 | 1.9 | 2.9 | 53.5 |
|  | CS_CoxPH | CS | 2% | 0.100 (0.078-0.122) | 0.747 | 0.046 | 0.154 | 0.070 | 0.00010 | 4.6 | 12.9 | 28.2 |
|  | CS_RSF | CS | 2% | 0.044 (0.029-0.059) | 0.701 | 0.023 | 0.077 | 0.035 | -0.00060 | 2.3 | 13.1 | 37.0 |
|  | DeepHit | CIF | 2% | 0.058 (0.041-0.076) | 0.701 | 0.029 | 0.097 | 0.044 | -0.00053 | 2.9 | 14.1 | 32.6 |
|  | FineGray | CIF | 2% | 0.096 (0.074-0.119) | 0.741 | 0.045 | 0.152 | 0.069 | 0.00017 | 4.5 | 10.7 | 31.4 |
|  | LR | binary | 5% | 0.040 (0.031-0.048) | 0.693 | 0.022 | 0.189 | 0.040 | -0.20269 | 2.2 | 4.1 | 49.8 |
|  | XGB | binary | 5% | 0.019 (0.014-0.025) | 0.565 | 0.011 | 0.097 | 0.021 | -0.00137 | 1.1 | 2.6 | 56.0 |
|  | LGBM | binary | 5% | 0.024 (0.018-0.031) | 0.624 | 0.014 | 0.122 | 0.026 | -0.01812 | 1.4 | 3.1 | 55.0 |
|  | CS_CoxPH | CS | 5% | 0.067 (0.054-0.079) | 0.747 | 0.031 | 0.266 | 0.056 | 0.00022 | 3.1 | 9.7 | 34.4 |
|  | CS_RSF | CS | 5% | 0.041 (0.033-0.052) | 0.701 | 0.020 | 0.172 | 0.036 | -0.00043 | 2.0 | 11.0 | 36.0 |
|  | DeepHit | CIF | 5% | 0.046 (0.036-0.057) | 0.701 | 0.022 | 0.189 | 0.040 | -0.00090 | 2.2 | 11.1 | 34.8 |
|  | FineGray | CIF | 5% | 0.062 (0.050-0.073) | 0.741 | 0.030 | 0.254 | 0.054 | 0.00026 | 3.0 | 8.1 | 37.3 |
| 2p 7d | LR | binary | 1% | 0.108 (0.082-0.133) | 0.719 | 0.090 | 0.066 | 0.076 | -0.06739 | 9.0 | 21.3 | 52.6 |
|  | XGB | binary | 1% | 0.036 (0.022-0.051) | 0.565 | 0.031 | 0.023 | 0.026 | -0.01250 | 3.1 | 7.8 | 75.4 |
|  | LGBM | binary | 1% | 0.059 (0.040-0.078) | 0.626 | 0.050 | 0.037 | 0.042 | -0.02200 | 5.0 | 8.2 | 71.6 |
|  | CS_CoxPH | CS | 1% | 0.115 (0.091-0.143) | 0.721 | 0.097 | 0.072 | 0.082 | -0.00080 | 9.7 | 25.4 | 49.0 |
|  | CS_RSF | CS | 1% | 0.077 (0.056-0.101) | 0.694 | 0.065 | 0.048 | 0.055 | -0.00172 | 6.5 | 22.1 | 55.7 |
|  | DeepHit | CIF | 1% | 0.046 (0.030-0.061) | 0.696 | 0.040 | 0.029 | 0.034 | -0.00174 | 4.0 | 23.5 | 59.6 |
|  | FineGray | CIF | 1% | 0.113 (0.089-0.138) | 0.714 | 0.096 | 0.070 | 0.081 | -0.00054 | 9.6 | 20.4 | 54.6 |
|  | LR | binary | 2% | 0.093 (0.076-0.109) | 0.719 | 0.077 | 0.114 | 0.092 | -0.08913 | 7.7 | 18.8 | 56.8 |
|  | XGB | binary | 2% | 0.037 (0.026-0.049) | 0.565 | 0.032 | 0.048 | 0.039 | -0.01632 | 3.2 | 7.4 | 77.7 |
|  | LGBM | binary | 2% | 0.042 (0.031-0.053) | 0.626 | 0.037 | 0.054 | 0.044 | -0.03198 | 3.7 | 8.2 | 75.4 |
|  | CS_CoxPH | CS | 2% | 0.105 (0.088-0.124) | 0.721 | 0.087 | 0.128 | 0.103 | -0.00076 | 8.7 | 20.4 | 53.3 |
|  | CS_RSF | CS | 2% | 0.064 (0.051-0.078) | 0.694 | 0.054 | 0.080 | 0.065 | -0.00234 | 5.4 | 22.6 | 56.7 |
|  | DeepHit | CIF | 2% | 0.054 (0.041-0.068) | 0.696 | 0.046 | 0.067 | 0.054 | -0.00237 | 4.6 | 21.8 | 58.3 |
|  | FineGray | CIF | 2% | 0.101 (0.084-0.119) | 0.714 | 0.085 | 0.125 | 0.101 | -0.00047 | 8.5 | 17.3 | 58.1 |
|  | LR | binary | 5% | 0.071 (0.061-0.080) | 0.719 | 0.060 | 0.220 | 0.094 | -0.13383 | 6.0 | 15.0 | 63.4 |
|  | XGB | binary | 5% | 0.029 (0.023-0.036) | 0.565 | 0.026 | 0.098 | 0.042 | -0.02199 | 2.6 | 6.4 | 80.8 |
|  | LGBM | binary | 5% | 0.036 (0.029-0.042) | 0.626 | 0.032 | 0.117 | 0.050 | -0.05198 | 3.2 | 8.3 | 77.8 |
|  | CS_CoxPH | CS | 5% | 0.075 (0.066-0.084) | 0.721 | 0.063 | 0.234 | 0.100 | -0.00094 | 6.3 | 16.2 | 61.9 |
|  | CS_RSF | CS | 5% | 0.051 (0.043-0.058) | 0.694 | 0.043 | 0.158 | 0.068 | -0.00293 | 4.3 | 20.1 | 60.6 |
|  | DeepHit | CIF | 5% | 0.051 (0.043-0.059) | 0.696 | 0.043 | 0.158 | 0.068 | -0.00398 | 4.3 | 19.4 | 60.6 |
|  | FineGray | CIF | 5% | 0.069 (0.060-0.078) | 0.714 | 0.058 | 0.216 | 0.092 | -0.00077 | 5.8 | 13.9 | 65.5 |

BRII with 95% bootstrap confidence intervals, AUROC, PPV, sensitivity, F1, net benefit, and
BRII decomposition components for seven models across all cohort-outcome combinations at
three additional top-k thresholds. Format and estimator conventions follow Table 2.

**Supplementary Table 10.**
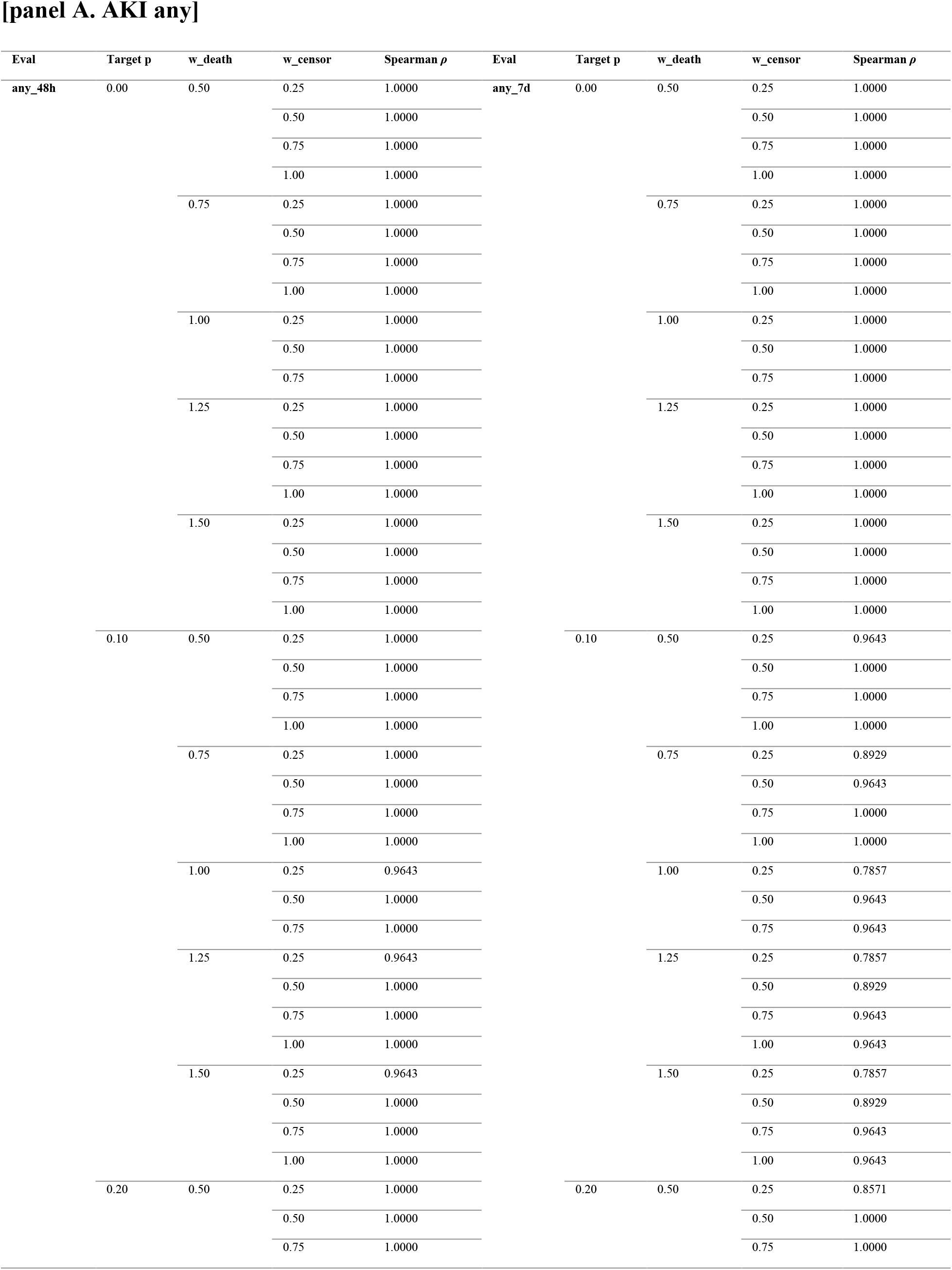
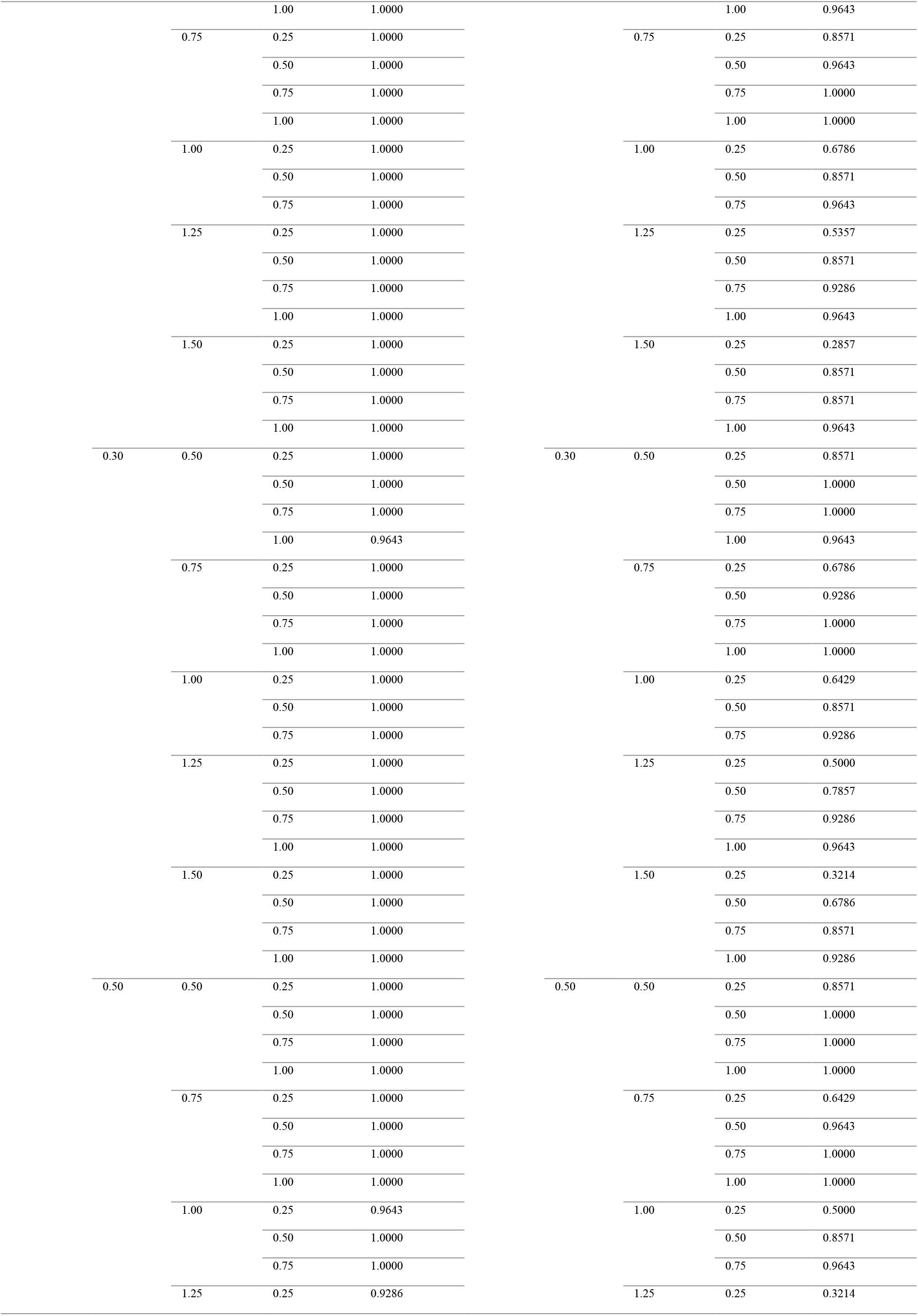
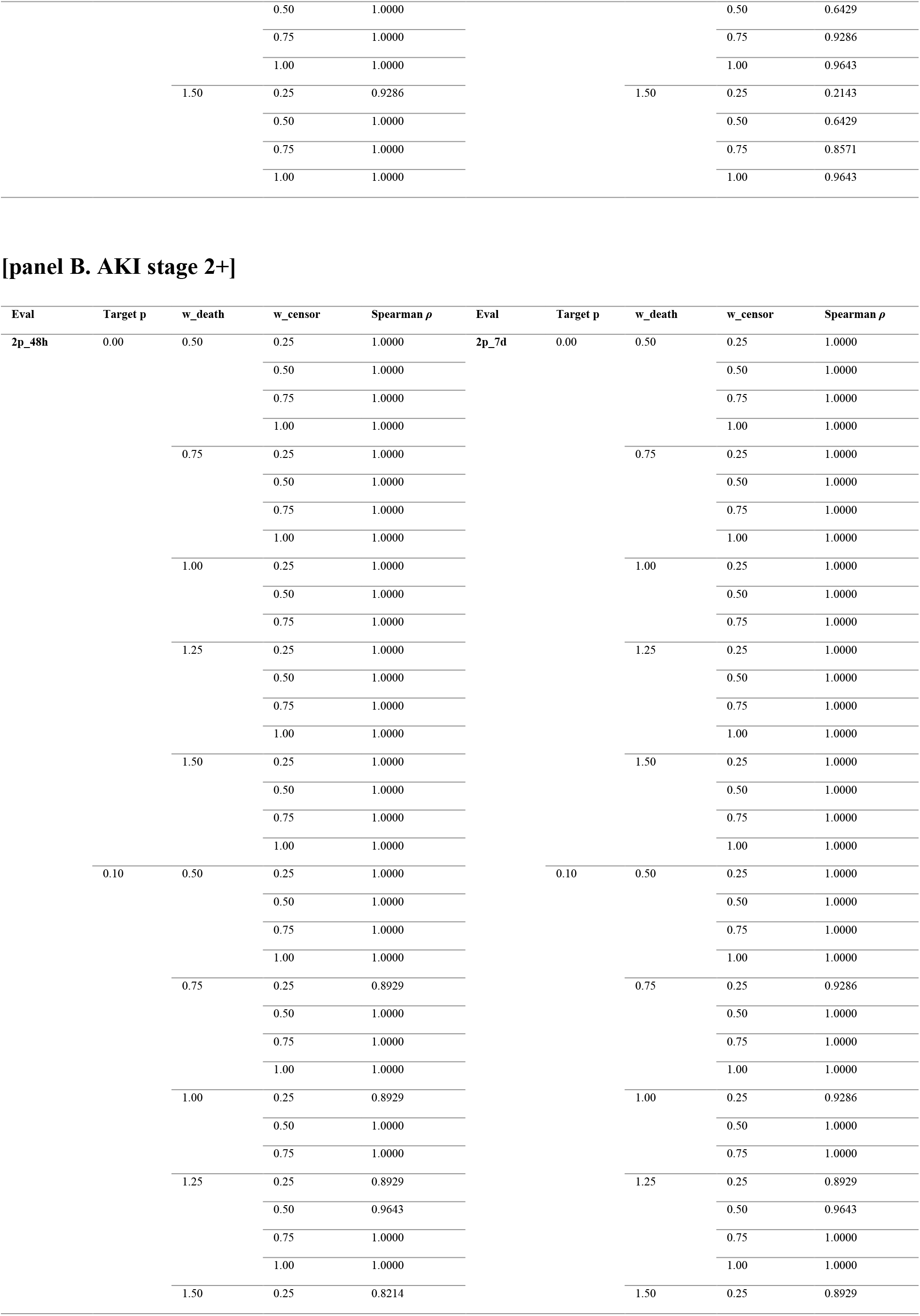
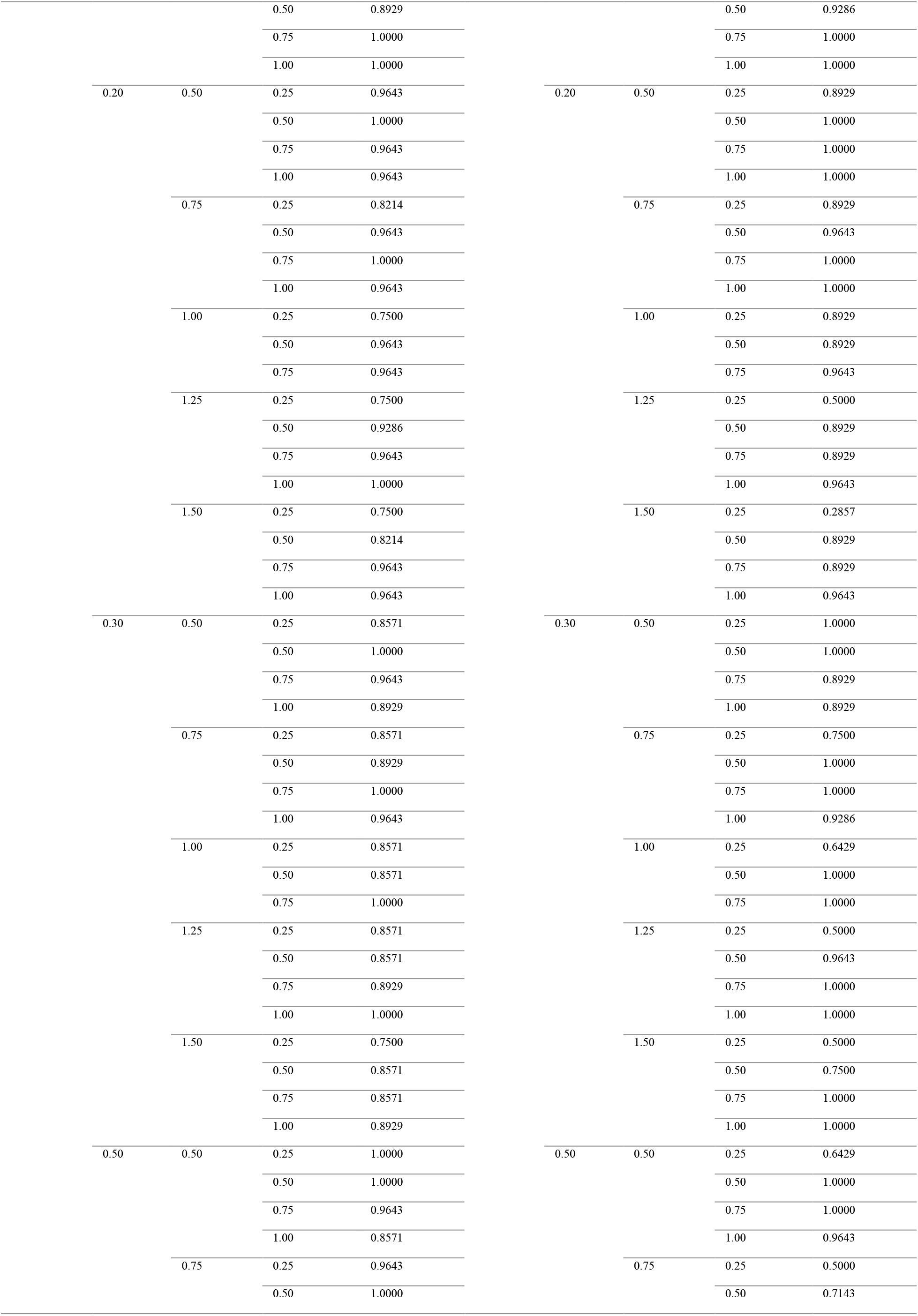
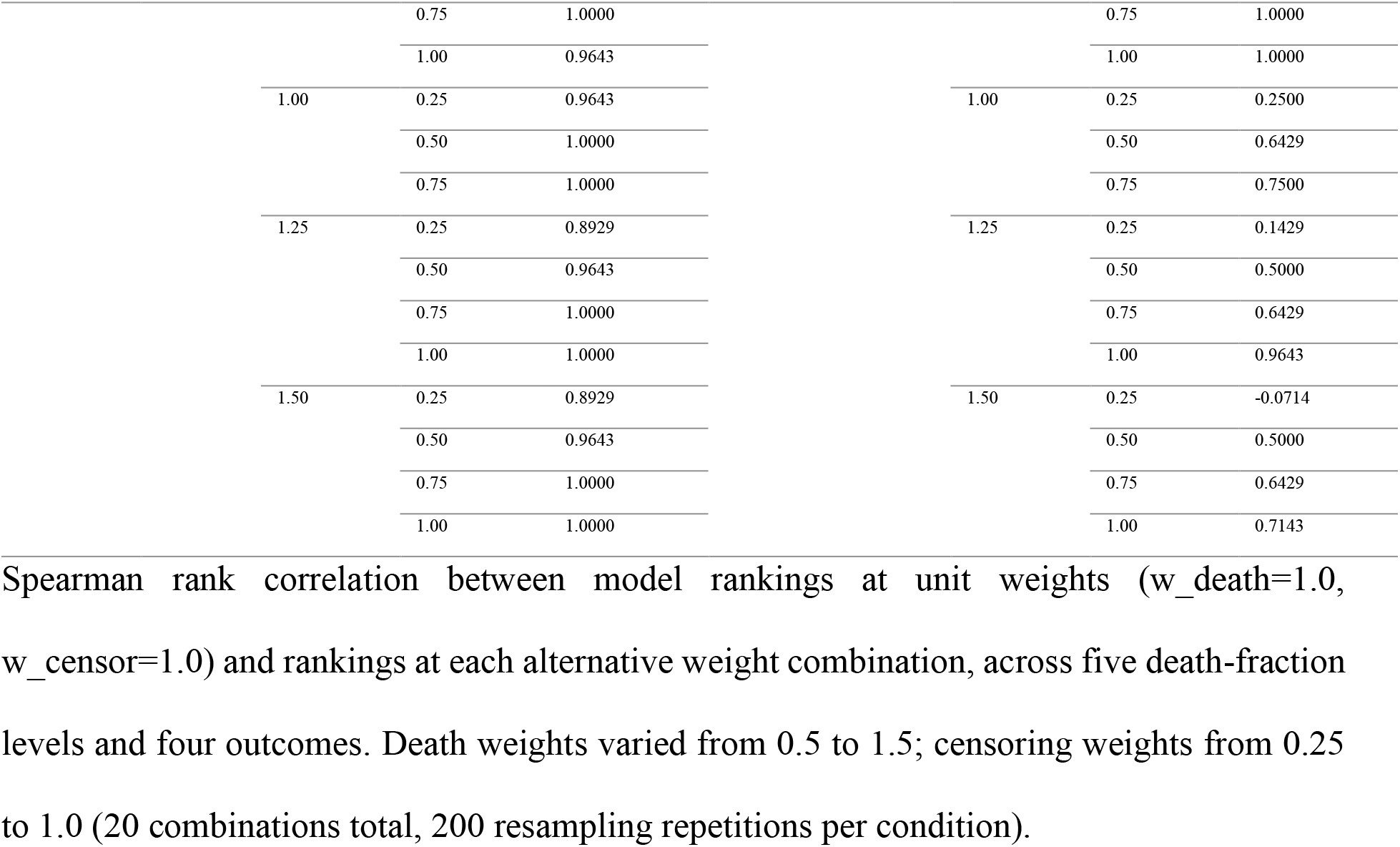
BRII ranking stability under alternative death and censoring weights.

**Supplementary Table 11.** Censoring-sweep sensitivity to bias signal choice.

**[panel A. any\_48h]**
| Censor Type | Censor Rate | Mean \|Diff\| | Max \|Diff\| | Spearman $\rho$ |
| --- | --- | --- | --- | --- |
| informative_healthy | 0.0 | 0.00000 | 0.00000 | 1.0000 |
|  | 0.1 | 0.01961 | 0.04146 | 0.7857 |
|  | 0.2 | 0.03661 | 0.06780 | 0.4643 |
|  | 0.3 | 0.05152 | 0.08990 | 0.4643 |
|  | 0.4 | 0.06508 | 0.11017 | 0.4643 |
|  | 0.5 | 0.07735 | 0.13588 | 0.4286 |
|  | 0.6 | 0.08785 | 0.14814 | 0.3929 |
|  | 0.7 | 0.09646 | 0.16660 | 0.3214 |
|  | 0.8 | 0.10537 | 0.17700 | 0.1429 |
|  | 0.9 | 0.11160 | 0.18285 | 0.1429 |
| informative_sick | 0.0 | 0.00000 | 0.00000 | 1.0000 |
|  | 0.1 | 0.01537 | 0.05268 | 0.6071 |
|  | 0.2 | 0.01870 | 0.05734 | 0.5714 |
|  | 0.3 | 0.01919 | 0.04858 | 0.5714 |
|  | 0.4 | 0.01825 | 0.04698 | 0.5000 |
|  | 0.5 | 0.01701 | 0.04542 | 0.0714 |
|  | 0.6 | 0.00853 | 0.03122 | 0.9643 |
|  | 0.7 | 0.00566 | 0.02606 | 1.0000 |
|  | 0.8 | 0.00559 | 0.02555 | 1.0000 |
|  | 0.9 | 0.00521 | 0.01994 | 1.0000 |

**[panel B. any\_7d]**
| Censor Type | Censor Rate | Mean \|Diff\| | Max \|Diff\| | Spearman $\rho$ |
| --- | --- | --- | --- | --- |
| informative_healthy | 0.0 | 0.00000 | 0.00000 | 1.0000 |
|  | 0.1 | 0.00829 | 0.02097 | 0.9286 |
|  | 0.2 | 0.01618 | 0.03844 | 0.9286 |
|  | 0.3 | 0.02367 | 0.05085 | 0.7143 |
|  | 0.4 | 0.03079 | 0.06249 | 0.7143 |
|  | 0.5 | 0.03775 | 0.07470 | 0.7143 |
|  | 0.6 | 0.04316 | 0.07926 | 0.7857 |
|  | 0.7 | 0.04796 | 0.08242 | 0.7857 |
|  | 0.8 | 0.05172 | 0.08661 | 0.7857 |
|  | 0.9 | 0.05499 | 0.08874 | 0.7500 |
| **informative_sick** | 0.0 | 0.00000 | 0.00000 | 1.0000 |
|  | 0.1 | 0.00839 | 0.03220 | 0.9643 |
|  | 0.2 | 0.01333 | 0.04021 | 0.9643 |
|  | 0.3 | 0.01721 | 0.05349 | 0.6429 |
|  | 0.4 | 0.01992 | 0.04903 | 0.2857 |
|  | 0.5 | 0.02124 | 0.05622 | 0.2143 |
|  | 0.6 | 0.01564 | 0.05590 | 0.6429 |
|  | 0.7 | 0.01155 | 0.05060 | 0.9286 |
|  | 0.8 | 0.01016 | 0.04302 | 0.9286 |
|  | 0.9 | 0.00949 | 0.04543 | 0.9286 |

[panel C. 2p\_48h]
| Censor Type | Censor Rate | Mean \|Diff\| | Max \|Diff\| | Spearman $\rho$ |
| --- | --- | --- | --- | --- |
| **informative_healthy** | 0.0 | 0.00000 | 0.00000 | 1.0000 |
|  | 0.1 | 0.08938 | 0.19349 | 0.5357 |
|  | 0.2 | 0.11510 | 0.22524 | 0.1071 |
|  | 0.3 | 0.12267 | 0.23730 | -0.2857 |
|  | 0.4 | 0.12269 | 0.23377 | -0.2857 |
|  | 0.5 | 0.12044 | 0.21537 | -0.2857 |
|  | 0.6 | 0.11512 | 0.20856 | -0.3571 |
|  | 0.7 | 0.10903 | 0.20856 | -0.3571 |
|  | 0.8 | 0.10325 | 0.20030 | -0.3571 |
|  | 0.9 | 0.09709 | 0.18707 | -0.3571 |
| **informative_sick** | 0.0 | 0.00000 | 0.00000 | 1.0000 |
|  | 0.1 | 0.02516 | 0.05966 | 0.5000 |
|  | 0.2 | 0.01928 | 0.04740 | 0.3929 |
|  | 0.3 | 0.01509 | 0.03786 | 0.0714 |
|  | 0.4 | 0.01176 | 0.02840 | -0.1429 |
|  | 0.5 | 0.00959 | 0.02089 | -0.3929 |
|  | 0.6 | 0.00652 | 0.01772 | -0.2500 |
|  | 0.7 | 0.00500 | 0.01246 | 0.4643 |
|  | 0.8 | 0.00421 | 0.01144 | 0.8214 |
|  | 0.9 | 0.00372 | 0.00996 | 0.8571 |

**[panel D. 2p\_7d]**
| Censor Type | Censor Rate | Mean \|Diff\| | Max \|Diff\| | Spearman $\rho$ |
| --- | --- | --- | --- | --- |
| informative_healthy | 0.0 | 0.00000 | 0.00000 | 1.0000 |
|  | 0.1 | 0.01046 | 0.02536 | 0.6071 |
|  | 0.2 | 0.01908 | 0.03935 | 0.6071 |
|  | 0.3 | 0.02611 | 0.05012 | 0.6071 |
|  | 0.4 | 0.03260 | 0.06124 | 0.6071 |
|  | 0.5 | 0.03788 | 0.07486 | 0.6071 |
|  | 0.6 | 0.04229 | 0.08046 | 0.6071 |
|  | 0.7 | 0.04547 | 0.08297 | 0.6071 |
|  | 0.8 | 0.04746 | 0.08491 | 0.6071 |
|  | 0.9 | 0.04906 | 0.08956 | 0.6071 |
| informative_sick | 0.0 | 0.00000 | 0.00000 | 1.0000 |
|  | 0.1 | 0.00776 | 0.02624 | 0.7143 |
|  | 0.2 | 0.01081 | 0.03096 | 0.3929 |
|  | 0.3 | 0.01191 | 0.03270 | 0.1786 |
|  | 0.4 | 0.01190 | 0.02759 | 0.1786 |
|  | 0.5 | 0.01143 | 0.02690 | -0.3929 |
|  | 0.6 | 0.00847 | 0.02108 | 0.0714 |
|  | 0.7 | 0.00596 | 0.01759 | 0.2857 |
|  | 0.8 | 0.00470 | 0.01382 | 0.6071 |
|  | 0.9 | 0.00391 | 0.01407 | 0.8929 |

BRII under informative censoring computed using Cox proportional hazards predictions as the bias signal, compared with the primary analysis using LightGBM predictions. Rows labeled “informative\_healthy” and “informative\_sick” denote informative censoring biased toward lower-risk and higher-risk patients, respectively. For each model-outcome-censoring-rate combination, absolute BRII difference and within-condition Spearman rank correlation between the two bias signals are reported.

**Supplementary Table 12.** Net-benefit-matched bins with divergent BRII decomposition.

**[panel A. any\_48h]**
| NB range | n | BRII range (2.5th to 97.5th percentile) | Death-share range | Censor-share range |
| --- | --- | --- | --- | --- |
| (−0.001017, 0.000030] | 1,000 | 0.1584 | 0.2098 | 0.1540 |
| (0.000030, 0.000084] | 538 | 0.2441 | 0.2600 | 0.1741 |
| (0.000084, 0.000151] | 620 | 0.2685 | 0.2640 | 0.1742 |
| (0.000151, 0.000285] | 680 | 0.2088 | 0.2199 | 0.1311 |
| (0.000285, 0.000420] | 737 | 0.2301 | 0.2244 | 0.1078 |
| (0.000420, 0.000652] | 645 | 0.2245 | 0.2210 | 0.0989 |
| (0.000652, 0.000890] | 680 | 0.2316 | 0.2094 | 0.1015 |
| (0.000890, 0.001310] | 702 | 0.2340 | 0.2069 | 0.0688 |
| (0.001310, 0.001850] | 758 | 0.2080 | 0.1855 | 0.0467 |
| (0.001850, 0.002590] | 640 | 0.2074 | 0.1732 | 0.0550 |
| (0.002590, 0.003530] | 737 | 0.1772 | 0.1580 | 0.0357 |
| (0.003530, 0.004820] | 663 | 0.1797 | 0.1416 | 0.0453 |
| (0.004820, 0.006960] | 724 | 0.1505 | 0.1223 | 0.0343 |
| (0.006960, 0.009780] | 676 | 0.1075 | 0.0794 | 0.0286 |
| (0.009780, 0.015100] | 700 | 0.1148 | 0.0880 | 0.0268 |
| (0.015100, 0.019700] | 700 | 0.0659 | 0.0463 | 0.0205 |
| (0.019700, 0.032000] | 712 | 0.0810 | 0.0695 | 0.0182 |
| (0.032000, 0.035300] | 688 | 0.0205 | 0.0231 | 0.0321 |
| (0.035300, 0.036000] | 726 | 0.0617 | 0.0454 | 0.0164 |
| (0.036000, 0.036800] | 674 | 0.0315 | 0.0323 | 0.0036 |

**[panel B. any\_7d]**
| NB range | n | BRII range (2.5th to 97.5th percentile) | Death-share range | Censor-share range |
| --- | --- | --- | --- | --- |
| (−0.001018, 0.000122] | 775 | 0.0201 | 0.0846 | 0.0926 |
| (0.000122, 0.000262] | 630 | 0.0391 | 0.0845 | 0.0954 |
| (0.000262, 0.000538] | 800 | 0.0440 | 0.0817 | 0.0988 |
| (0.000538, 0.000853] | 602 | 0.0326 | 0.0829 | 0.0939 |
| (0.000853, 0.001410] | 759 | 0.0516 | 0.0811 | 0.1083 |
| (0.001410, 0.001970] | 637 | 0.0317 | 0.0818 | 0.0922 |
| (0.001970, 0.002930] | 698 | 0.0670 | 0.0765 | 0.1148 |
| (0.002930, 0.003850] | 701 | 0.0355 | 0.0749 | 0.0904 |
| (0.003850, 0.005460] | 707 | 0.0760 | 0.0714 | 0.1221 |
| (0.005460, 0.007170] | 691 | 0.0297 | 0.0677 | 0.0738 |
| (0.007170, 0.009860] | 713 | 0.0917 | 0.0657 | 0.1290 |
| (0.009860, 0.012900] | 687 | 0.0335 | 0.0428 | 0.0661 |
| (0.012900, 0.018700] | 703 | 0.1060 | 0.0593 | 0.1330 |
| (0.018700, 0.023600] | 697 | 0.0286 | 0.0318 | 0.0528 |
| (0.023600, 0.036800] | 737 | 0.0887 | 0.0717 | 0.1308 |
| (0.036800, 0.038000] | 712 | 0.0789 | 0.0680 | 0.1329 |
| (0.038000, 0.040200] | 751 | 0.0207 | 0.0680 | 0.0551 |
| (0.040200, 0.044300] | 612 | 0.0312 | 0.0493 | 0.0280 |
| (0.044300, 0.045400] | 788 | 0.0176 | 0.0357 | 0.0491 |
| (0.045400, 0.046600] | 600 | 0.0138 | 0.0000 | 0.0139 |

[panel C. 2p\_48h]
| NB range | n | BRII range (2.5th to 97.5th percentile) | Death-share range | Censor-share range |
| --- | --- | --- | --- | --- |
| (-0.000118, 0.000027] | 1,036 | 0.2103 | 0.4756 | 0.3809 |
| (0.000027, 0.000057] | 471 | 0.2383 | 0.4983 | 0.3810 |
| (0.000057, 0.000118] | 608 | 0.3048 | 0.4989 | 0.3113 |
| (0.000118, 0.000194] | 802 | 0.3015 | 0.4945 | 0.3057 |
| (0.000194, 0.000311] | 624 | 0.2701 | 0.4614 | 0.2245 |
| (0.000311, 0.000419] | 695 | 0.2818 | 0.4400 | 0.1703 |
| (0.000419, 0.000430] | 800 | 0.0589 | 0.1024 | 0.0436 |
| (0.000430, 0.000450] | 666 | 0.3737 | 0.5785 | 0.2099 |
| (0.000450, 0.000460] | 600 | 0.0699 | 0.1164 | 0.0464 |
| (0.000460, 0.000480] | 788 | 0.3912 | 0.5741 | 0.1849 |
| (0.000480, 0.000523] | 614 | 0.1635 | 0.1782 | 0.1050 |
| (0.000523, 0.000582] | 761 | 0.1340 | 0.2940 | 0.1700 |
| (0.000582, 0.000615] | 899 | 0.1402 | 0.2369 | 0.1515 |
| (0.000615, 0.000719] | 436 | 0.0619 | 0.0671 | 0.0904 |
| (0.000719, 0.001070] | 871 | 0.2986 | 0.2981 | 0.0279 |
| (0.001070, 0.001076] | 600 | 0.0809 | 0.1070 | 0.0261 |
| (0.001076, 0.001218] | 740 | 0.3953 | 0.4332 | 0.0470 |
| (0.001218, 0.001256] | 591 | 0.1232 | 0.1416 | 0.0436 |
| (0.001256, 0.001290] | 785 | 0.1556 | 0.1815 | 0.0446 |
| (0.001290, 0.001293] | 613 | 0.0897 | 0.0745 | 0.0441 |

[panel D. 2p\_7d]
| NB range | n | BRII range (2.5th to 97.5th percentile) | Death-share range | Censor-share range |
| --- | --- | --- | --- | --- |
| (-0.001021, 0.000072] | 763 | 0.0241 | 0.3642 | 0.3699 |
| (0.000072, 0.000195] | 664 | 0.0336 | 0.3518 | 0.3780 |
| (0.000195, 0.000421] | 781 | 0.0545 | 0.3510 | 0.3601 |
| (0.000421, 0.000575] | 621 | 0.0298 | 0.3397 | 0.3495 |
| (0.000575, 0.001090] | 672 | 0.0614 | 0.3399 | 0.3465 |
| (0.001090, 0.001330] | 714 | 0.0259 | 0.3221 | 0.3292 |
| (0.001330, 0.001640] | 780 | 0.0427 | 0.3657 | 0.3446 |
| (0.001640, 0.001650] | 803 | 0.0061 | 0.0349 | 0.0288 |
| (0.001650, 0.002080] | 506 | 0.0270 | 0.1047 | 0.0877 |
| (0.002080, 0.002420] | 807 | 0.0689 | 0.2978 | 0.2370 |
| (0.002420, 0.002730] | 603 | 0.0803 | 0.2556 | 0.1940 |
| (0.002730, 0.003080] | 687 | 0.0702 | 0.0897 | 0.1203 |
| (0.003080, 0.003230] | 799 | 0.0532 | 0.1112 | 0.1458 |
| (0.003230, 0.003280] | 671 | 0.0621 | 0.0871 | 0.1303 |
| (0.003280, 0.003430] | 729 | 0.0694 | 0.2081 | 0.2051 |
| (0.003430, 0.003640] | 615 | 0.0329 | 0.0968 | 0.0836 |
| (0.003640, 0.003760] | 785 | 0.0382 | 0.1801 | 0.1420 |
| (0.003760, 0.003860] | 653 | 0.0343 | 0.1413 | 0.1130 |
| (0.003860, 0.004000] | 747 | 0.0237 | 0.0981 | 0.0978 |
| (0.004000, 0.004290] | 600 | 0.0172 | 0.0663 | 0.0491 |

For each outcome, net benefit across death-contamination levels (seven models, 200 repetitions per death-fraction level) was grouped into 20 quantile bins. For each bin, the number of observations and the 2.5th to 97.5th percentile ranges of BRII, the death share, and the censoring share within the bin are reported. A bin is counted as divergent when the BRII range exceeds 0.05 or the death-share range exceeds 0.10: 18 of 20 bins for any\_48h, 7 for any\_7d, 20 for 2p\_48h, and 16 for 2p\_7d. The Spearman correlation between BRII and net benefit was 0.982, 0.995, 0.732, and 0.930.

**Supplementary Table 13.** Rank agreement of BRII with PPV and with the within-stratum Aalen-Johansen ratio.

| Cohort | Outcome | Top-k | PPV:<br>concordant | PPV:<br>discordant | PPV:<br>tied | R:<br>concordant | R:<br>discordant | R: tied | $\rho$ (BRII, R) |
| --- | --- | --- | --- | --- | --- | --- | --- | --- | --- |
| MIMIC-IV | any_48h | 1% | 17 | 4 | 0 | 20 | 1 | 0 | 0.96 |
| MIMIC-IV | any_48h | 2% | 20 | 1 | 0 | 21 | 0 | 0 | 1.00 |
| MIMIC-IV | any_48h | 5% | 15 | 6 | 0 | 20 | 1 | 0 | 0.96 |
| MIMIC-IV | any_48h | 10% | 14 | 6 | 1 | 19 | 2 | 0 | 0.93 |
| MIMIC-IV | any_7d | 1% | 20 | 1 | 0 | 16 | 5 | 0 | 0.71 |
| MIMIC-IV | any_7d | 2% | 18 | 3 | 0 | 13 | 8 | 0 | 0.36 |
| MIMIC-IV | any_7d | 5% | 19 | 1 | 1 | 13 | 8 | 0 | 0.29 |
| MIMIC-IV | any_7d | 10% | 17 | 3 | 1 | 8 | 13 | 0 | -0.32 |
| MIMIC-IV | 2p_48h | 1% | 8 | 12 | 1 | 21 | 0 | 0 | 1.00 |
| MIMIC-IV | 2p_48h | 2% | 12 | 7 | 2 | 19 | 2 | 0 | 0.93 |
| MIMIC-IV | 2p_48h | 5% | 13 | 8 | 0 | 19 | 2 | 0 | 0.89 |
| MIMIC-IV | 2p_48h | 10% | 14 | 6 | 1 | 19 | 2 | 0 | 0.89 |
| MIMIC-IV | 2p_7d | 1% | 18 | 3 | 0 | 13 | 8 | 0 | 0.32 |
| MIMIC-IV | 2p_7d | 2% | 19 | 1 | 1 | 15 | 6 | 0 | 0.46 |
| MIMIC-IV | 2p_7d | 5% | 18 | 2 | 1 | 13 | 8 | 0 | 0.25 |
| MIMIC-IV | 2p_7d | 10% | 19 | 0 | 2 | 9 | 12 | 0 | -0.18 |
| MIMIC-III | any_48h | 1% | 16 | 5 | 0 | 18 | 3 | 0 | 0.86 |
| MIMIC-III | any_48h | 2% | 17 | 4 | 0 | 20 | 1 | 0 | 0.96 |
| MIMIC-III | any_48h | 5% | 14 | 6 | 1 | 19 | 2 | 0 | 0.93 |
| MIMIC-III | any_48h | 10% | 13 | 7 | 1 | 21 | 0 | 0 | 1.00 |
| MIMIC-III | any_7d | 1% | 20 | 0 | 1 | 14 | 7 | 0 | 0.46 |
| MIMIC-III | any_7d | 2% | 19 | 1 | 1 | 16 | 5 | 0 | 0.64 |
| MIMIC-III | any_7d | 5% | 14 | 6 | 1 | 12 | 9 | 0 | 0.04 |
| MIMIC-III | any_7d | 10% | 18 | 2 | 1 | 16 | 5 | 0 | 0.61 |
| MIMIC-III | 2p_48h | 1% | 12 | 8 | 1 | 19 | 1 | 1 | 0.94 |
| MIMIC-III | 2p_48h | 2% | 13 | 8 | 0 | 19 | 2 | 0 | 0.89 |
| MIMIC-III | 2p_48h | 5% | 13 | 7 | 1 | 18 | 3 | 0 | 0.86 |
| MIMIC-III | 2p_48h | 10% | 16 | 5 | 0 | 16 | 5 | 0 | 0.61 |
| MIMIC-III | 2p_7d | 1% | 20 | 0 | 1 | 12 | 9 | 0 | 0.11 |
| MIMIC-III | 2p_7d | 2% | 19 | 0 | 2 | 11 | 10 | 0 | -0.07 |
| MIMIC-III | 2p_7d | 5% | 18 | 2 | 1 | 10 | 11 | 0 | 0.00 |
| **MIMIC-III** | 2p_7d | 10% | 18 | 2 | 1 | 8 | 13 | 0 | -0.25 |
| **eICU-CRD** | any_48h | 1% | 20 | 1 | 0 | 12 | 9 | 0 | 0.21 |
| **eICU-CRD** | any_48h | 2% | 19 | 2 | 0 | 10 | 11 | 0 | 0.04 |
| **eICU-CRD** | any_48h | 5% | 18 | 3 | 0 | 8 | 13 | 0 | -0.29 |
| **eICU-CRD** | any_48h | 10% | 19 | 2 | 0 | 9 | 12 | 0 | -0.32 |
| **eICU-CRD** | any_7d | 1% | 21 | 0 | 0 | 10 | 11 | 0 | 0.00 |
| **eICU-CRD** | any_7d | 2% | 21 | 0 | 0 | 12 | 9 | 0 | 0.29 |
| **eICU-CRD** | any_7d | 5% | 21 | 0 | 0 | 17 | 4 | 0 | 0.79 |
| **eICU-CRD** | any_7d | 10% | 21 | 0 | 0 | 12 | 9 | 0 | 0.29 |
| **eICU-CRD** | 2p_48h | 1% | 20 | 0 | 1 | 6 | 15 | 0 | -0.61 |
| **eICU-CRD** | 2p_48h | 2% | 20 | 0 | 1 | 6 | 15 | 0 | -0.64 |
| **eICU-CRD** | 2p_48h | 5% | 19 | 1 | 1 | 8 | 13 | 0 | -0.54 |
| **eICU-CRD** | 2p_48h | 10% | 19 | 2 | 0 | 10 | 11 | 0 | -0.14 |
| **eICU-CRD** | 2p_7d | 1% | 21 | 0 | 0 | 13 | 8 | 0 | 0.39 |
| **eICU-CRD** | 2p_7d | 2% | 21 | 0 | 0 | 13 | 8 | 0 | 0.32 |
| **eICU-CRD** | 2p_7d | 5% | 20 | 0 | 1 | 13 | 8 | 0 | 0.54 |
| **eICU-CRD** | 2p_7d | 10% | 20 | 0 | 1 | 12 | 9 | 0 | 0.11 |

**Supplementary Table 14.**
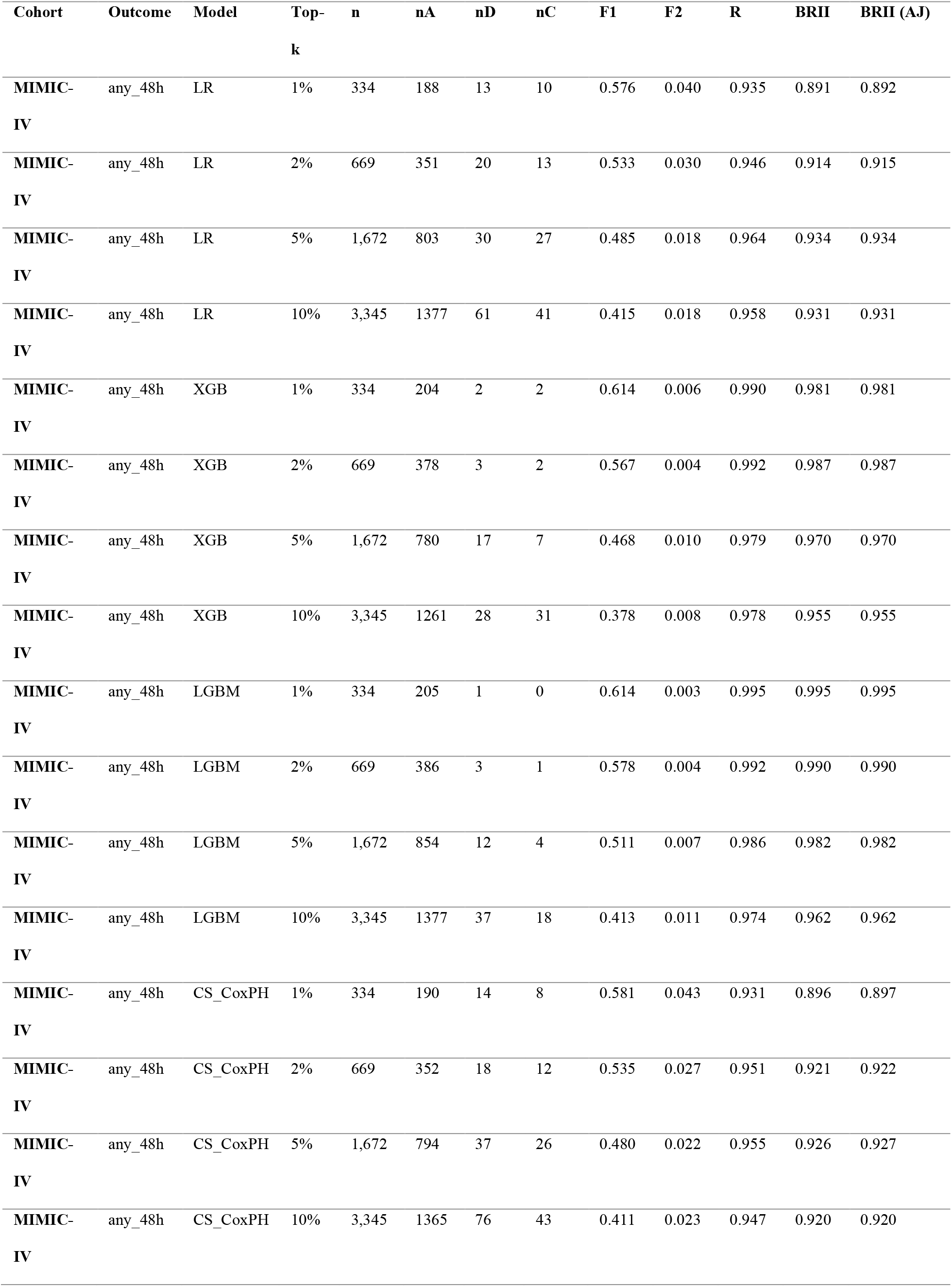
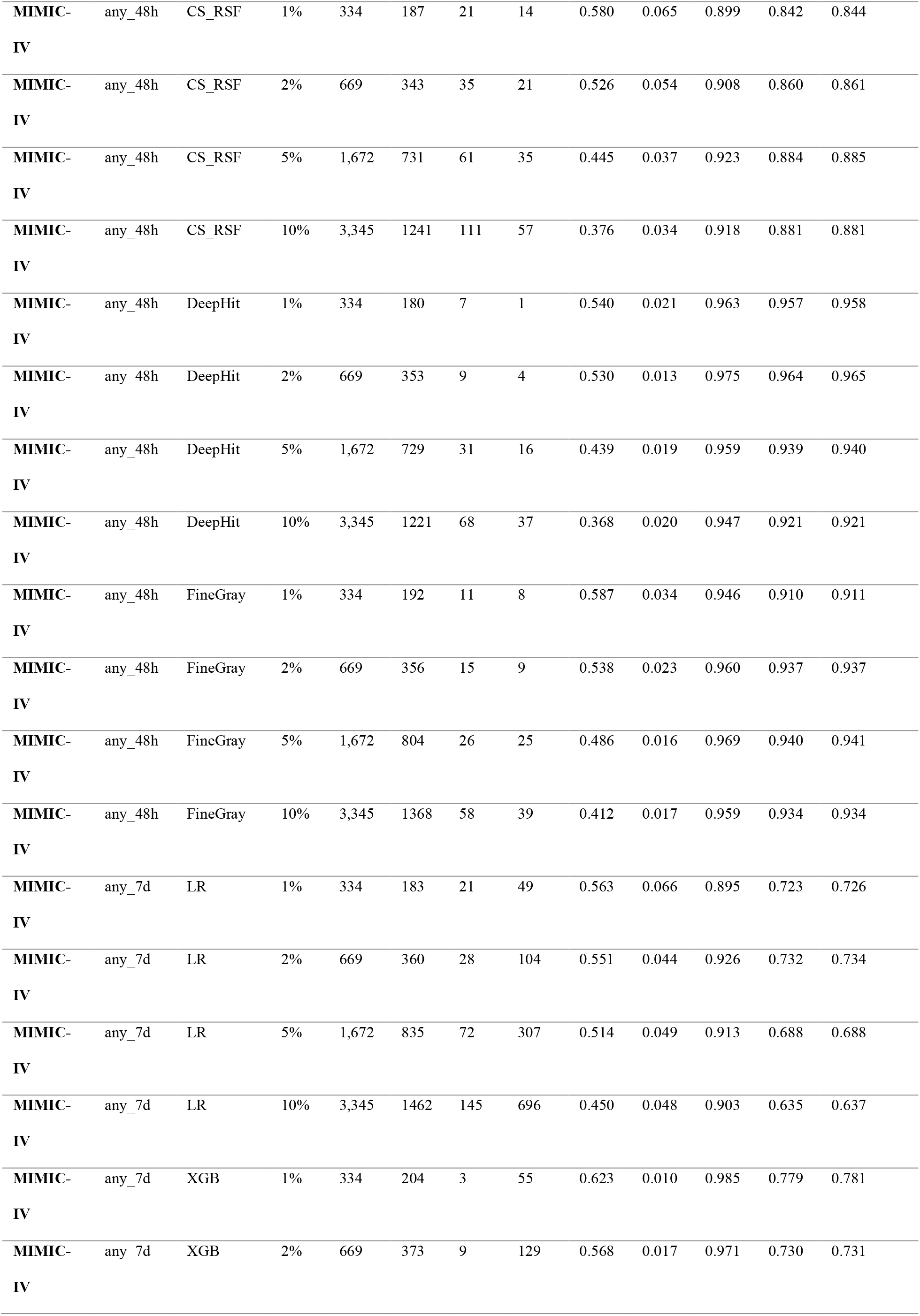
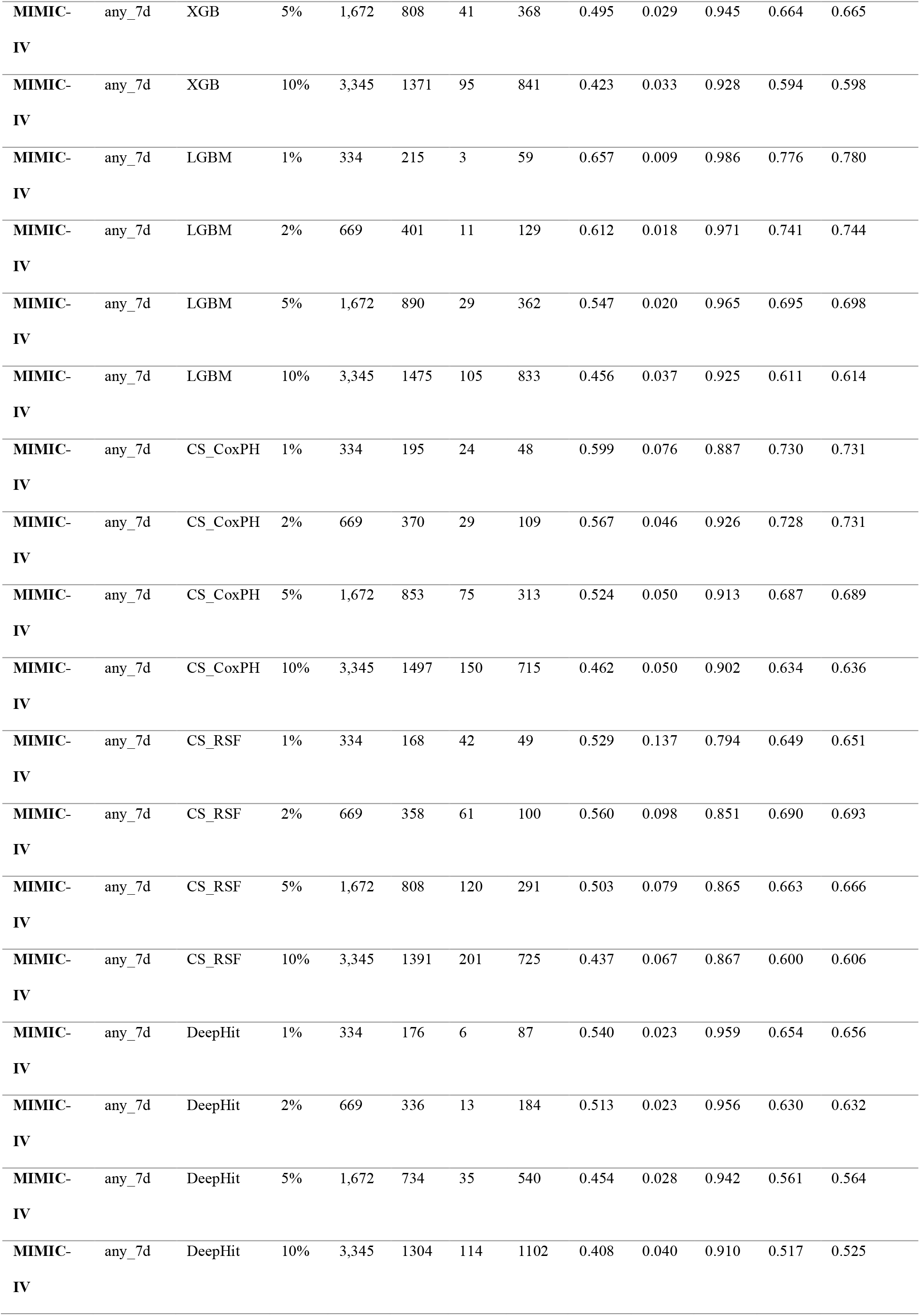
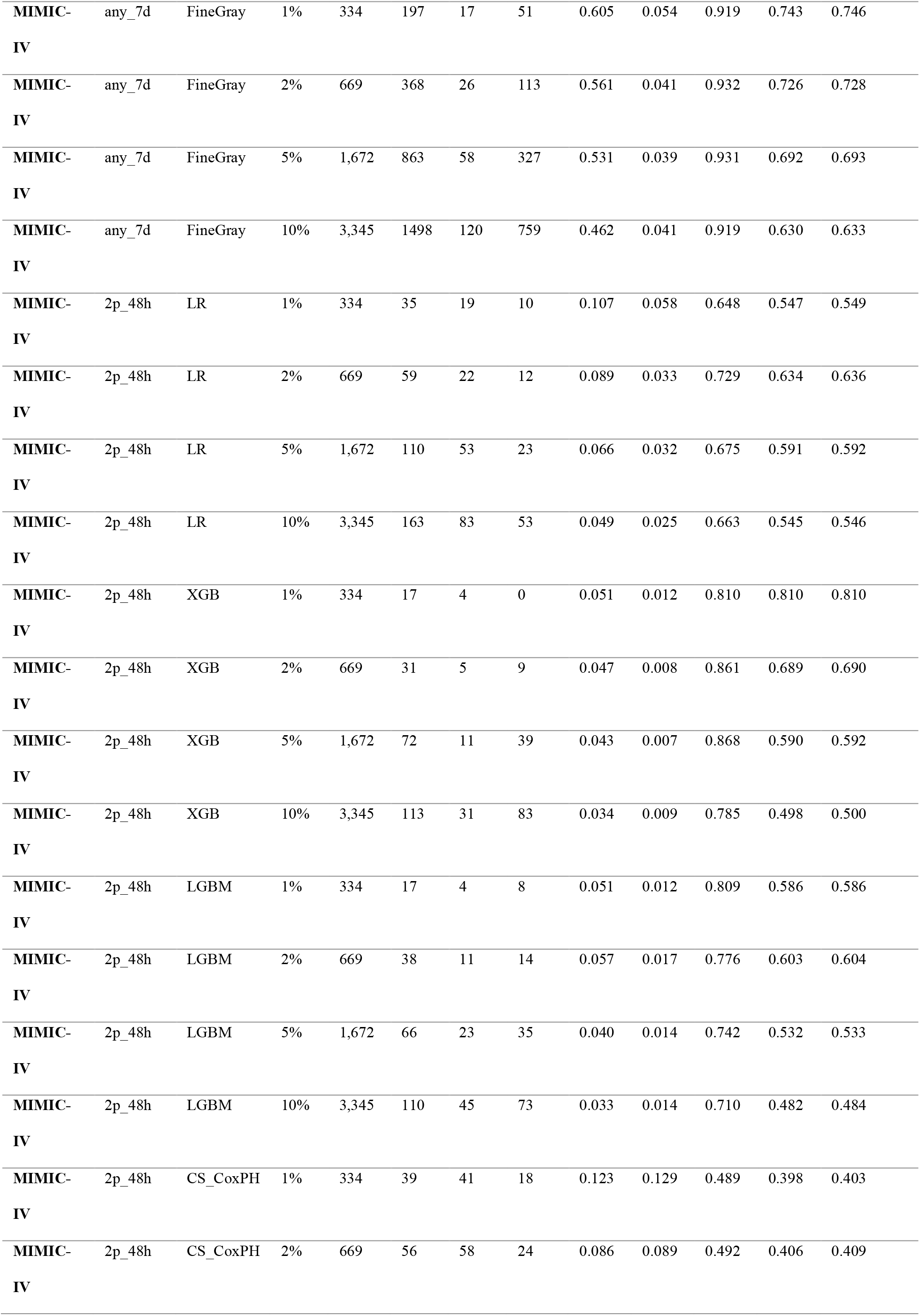
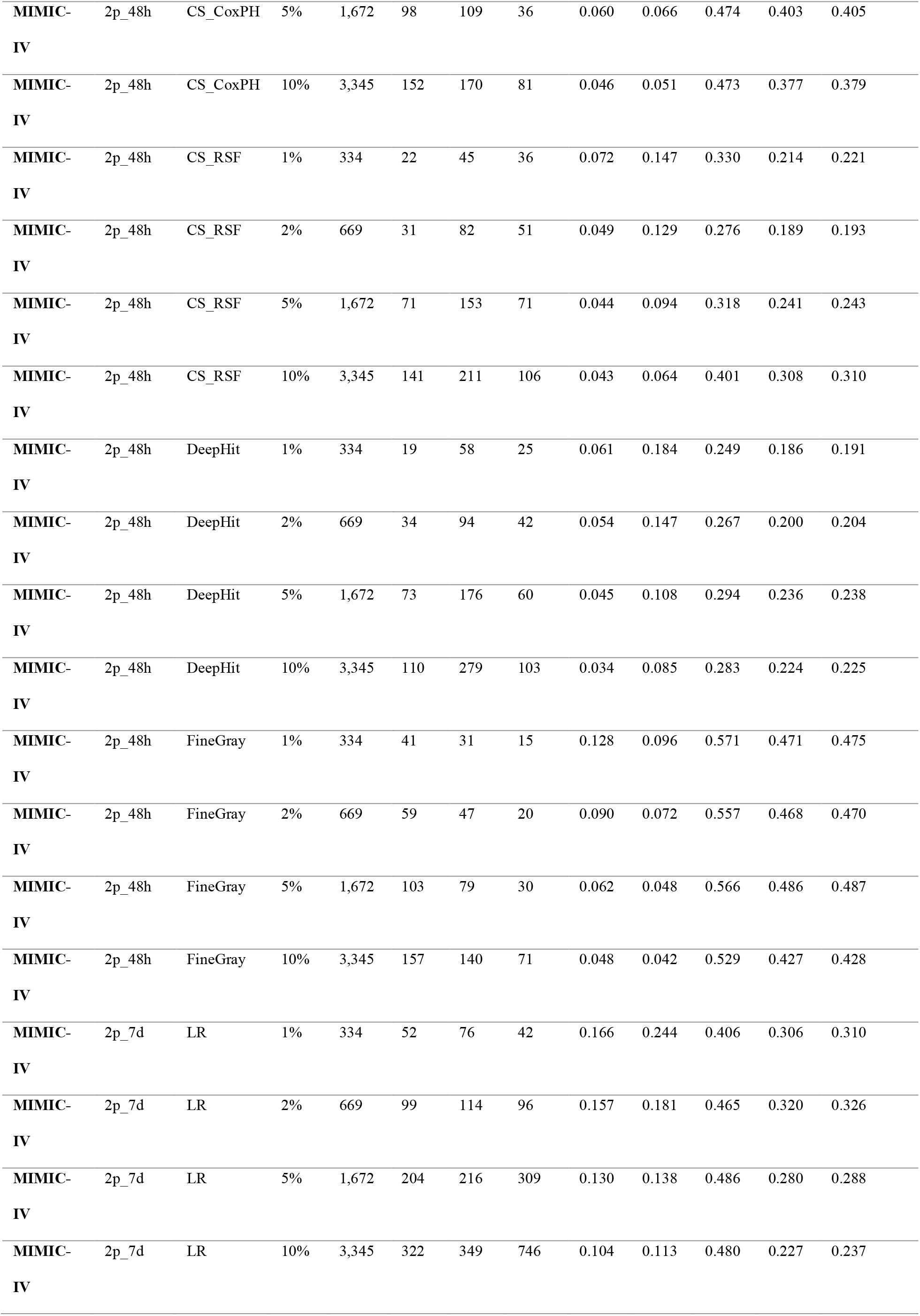
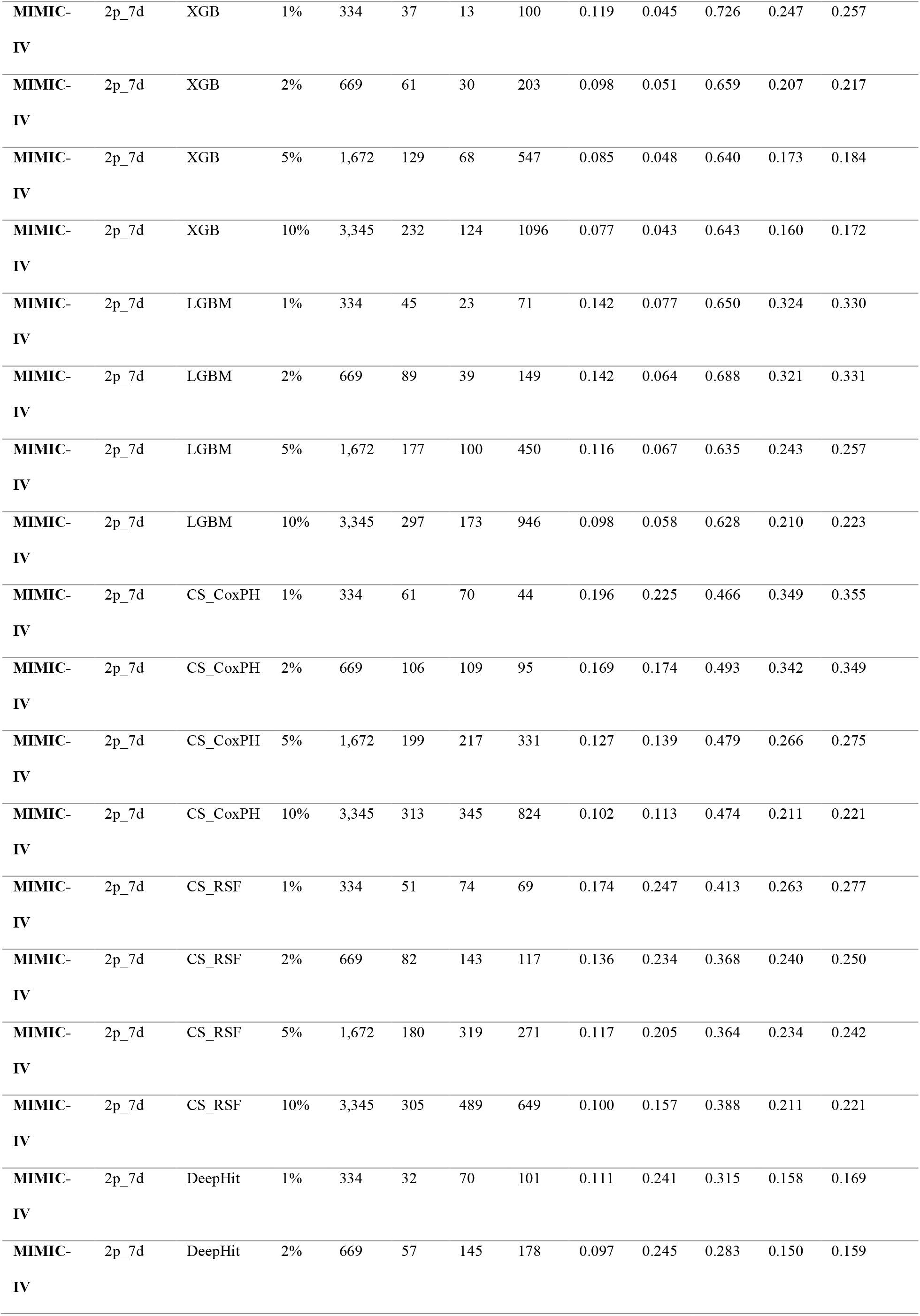
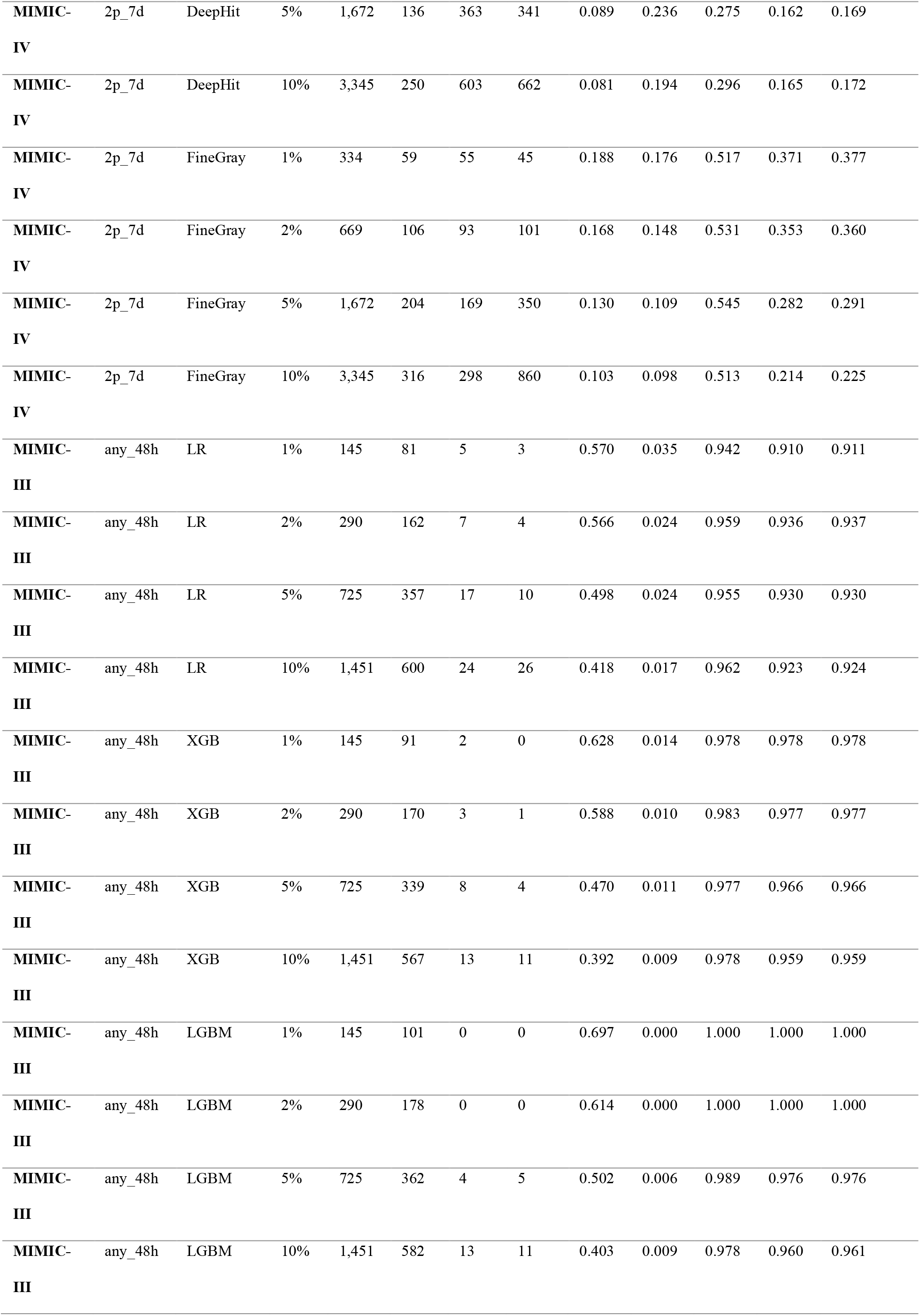
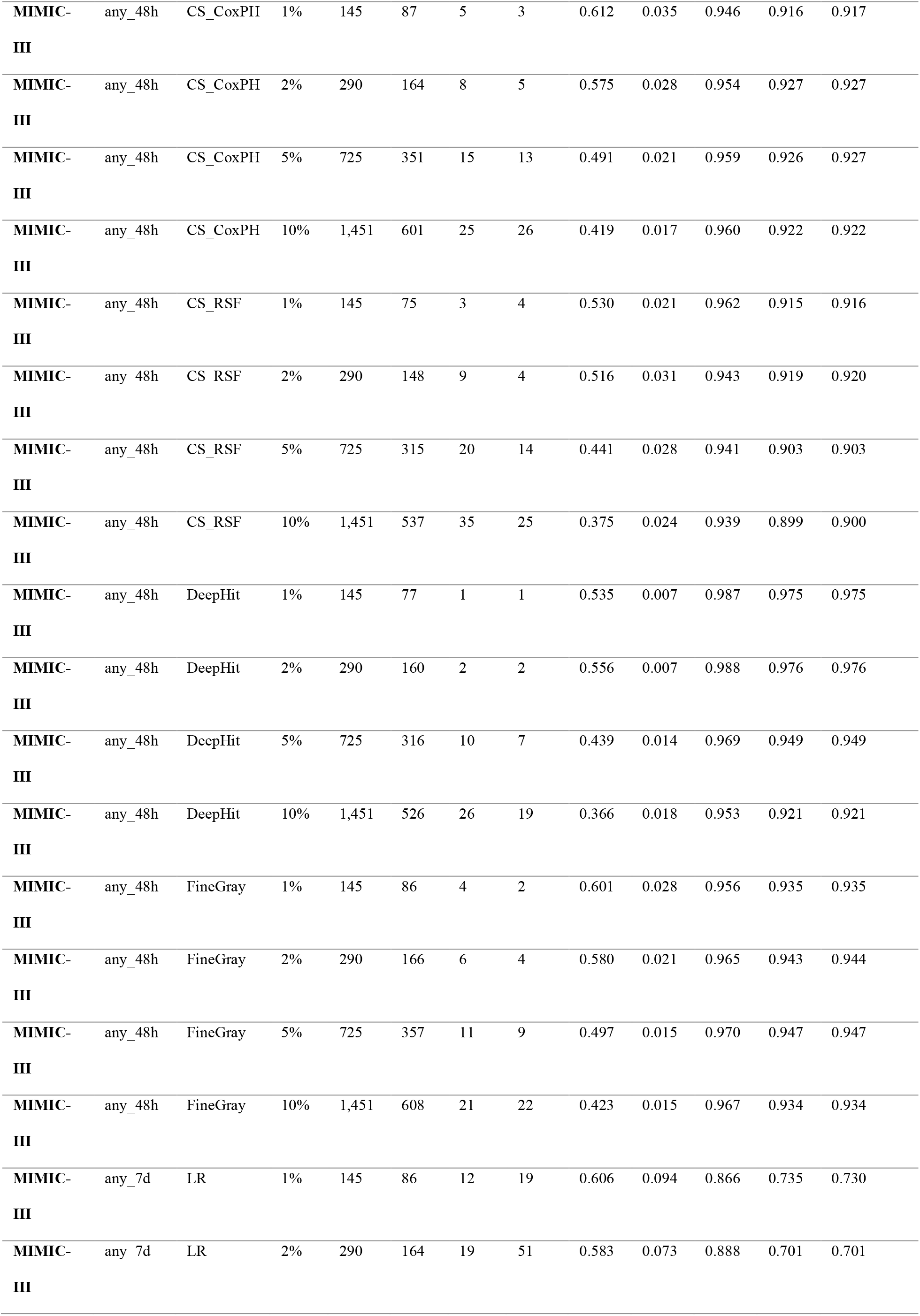
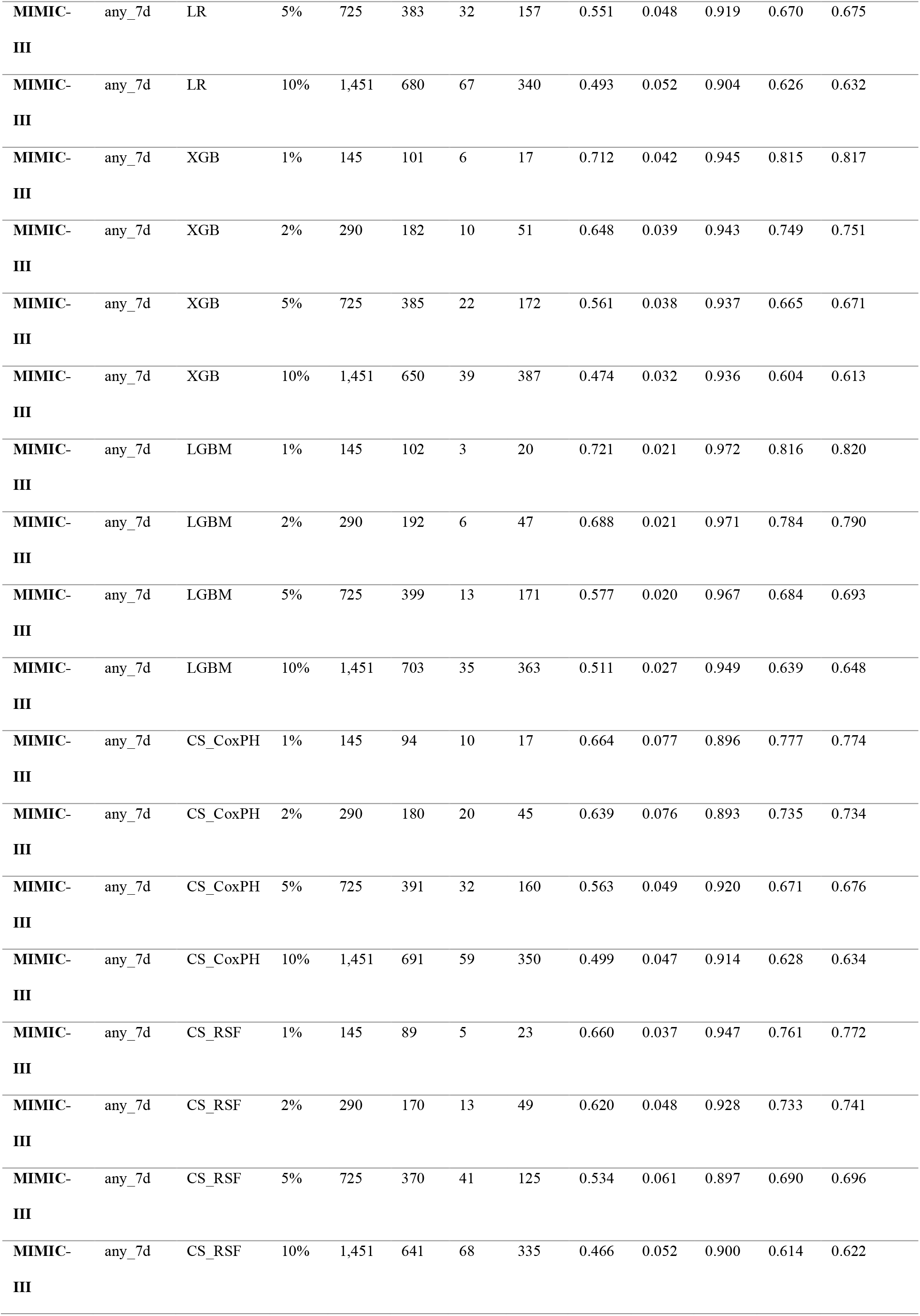
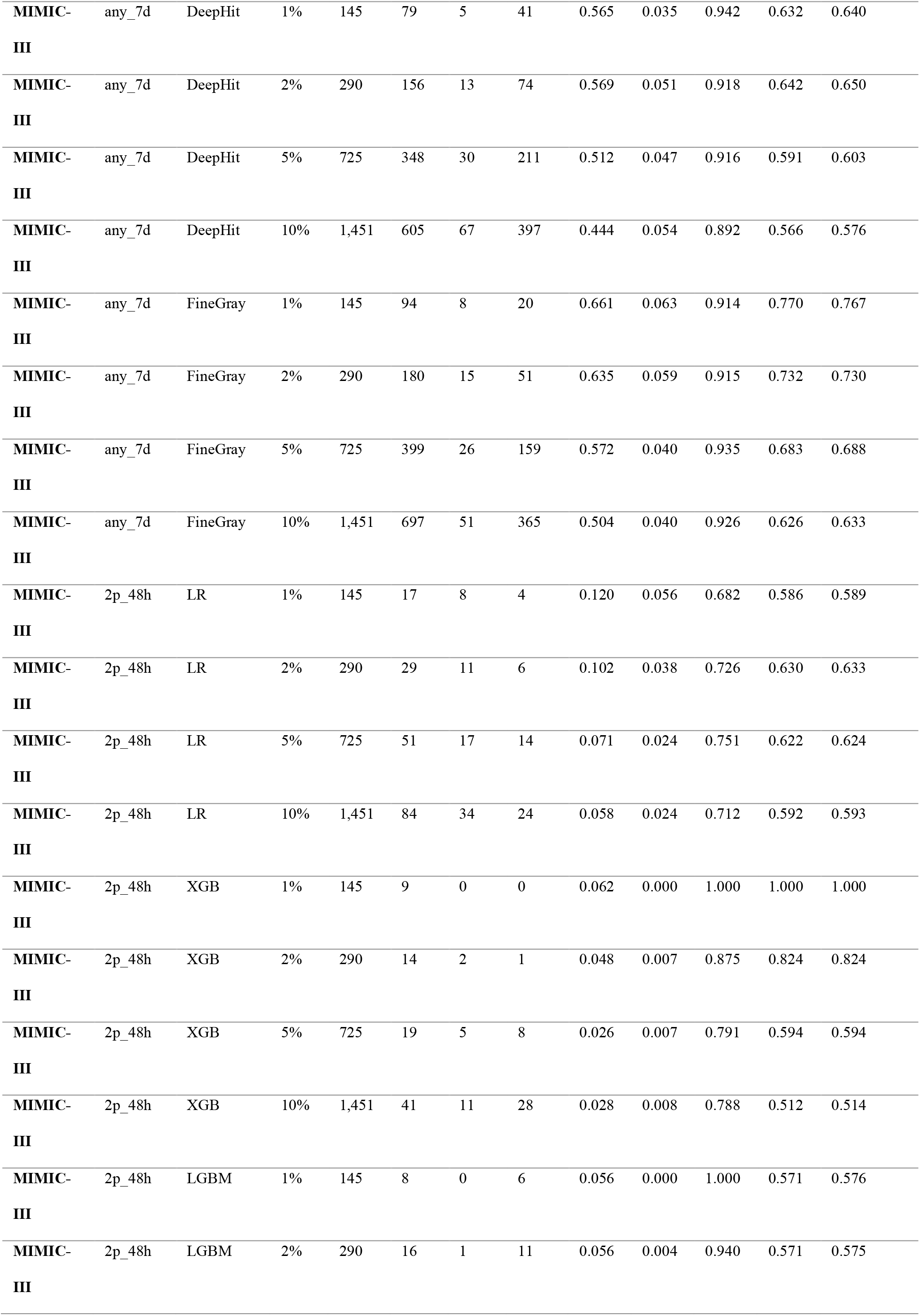
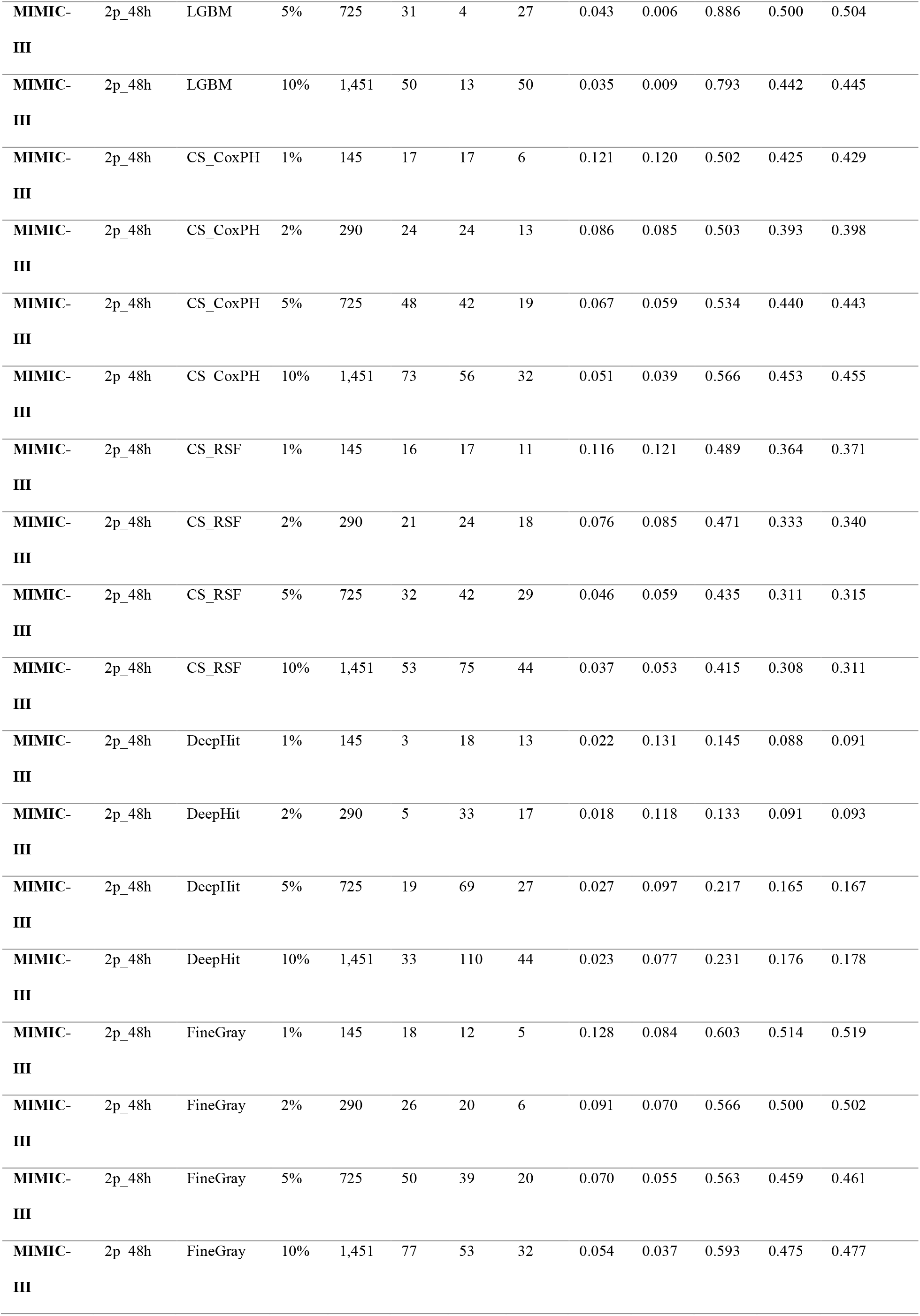
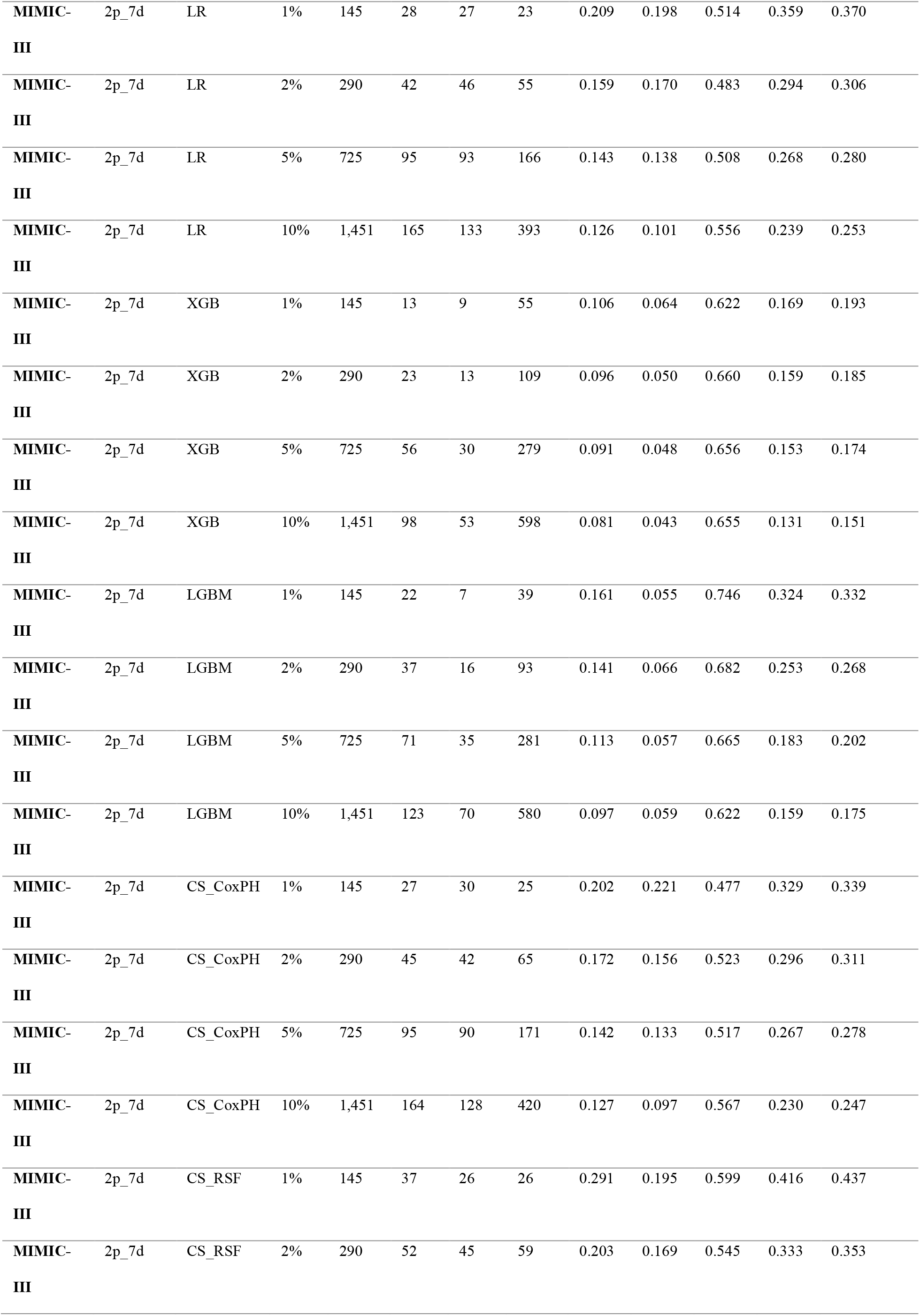
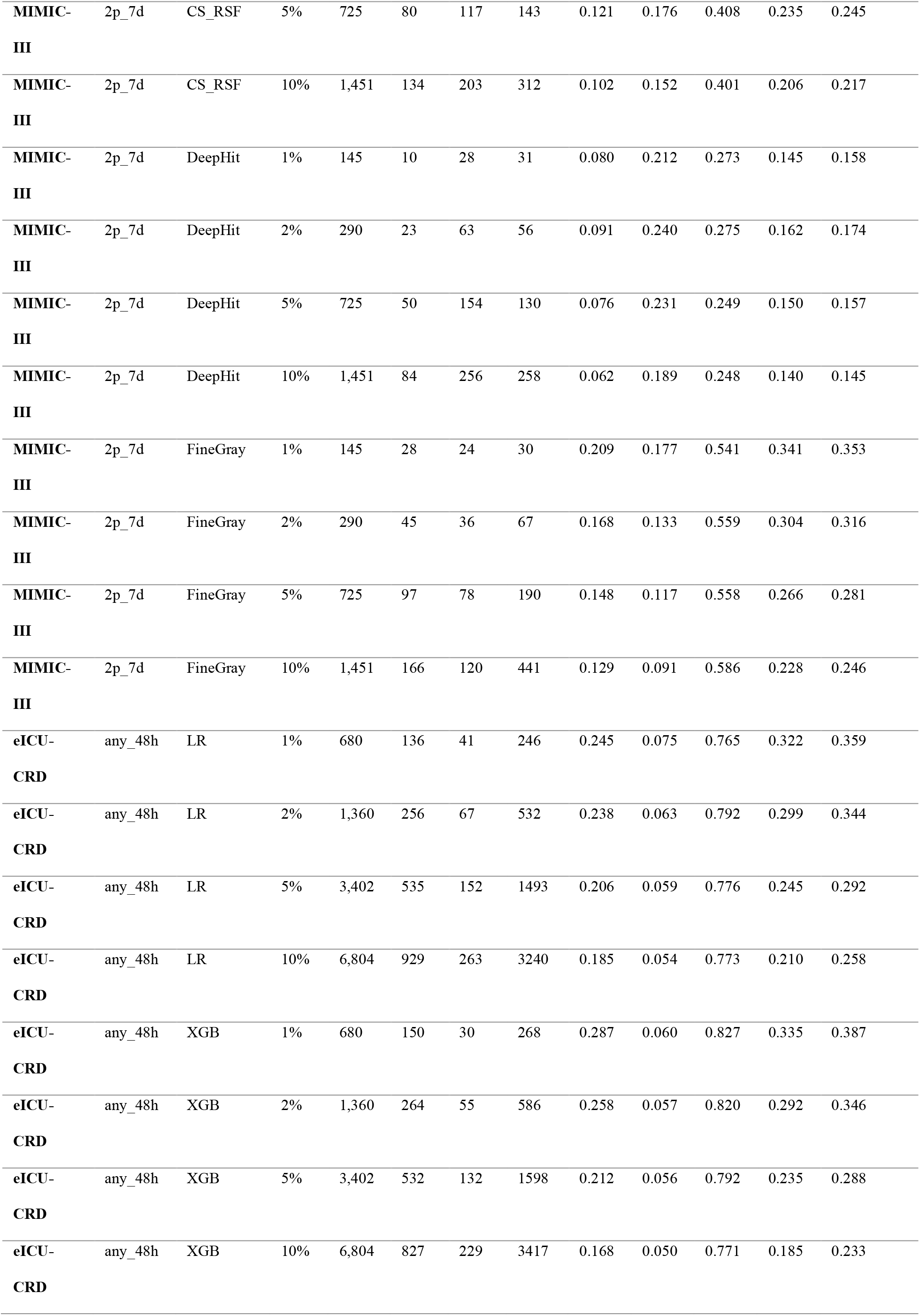
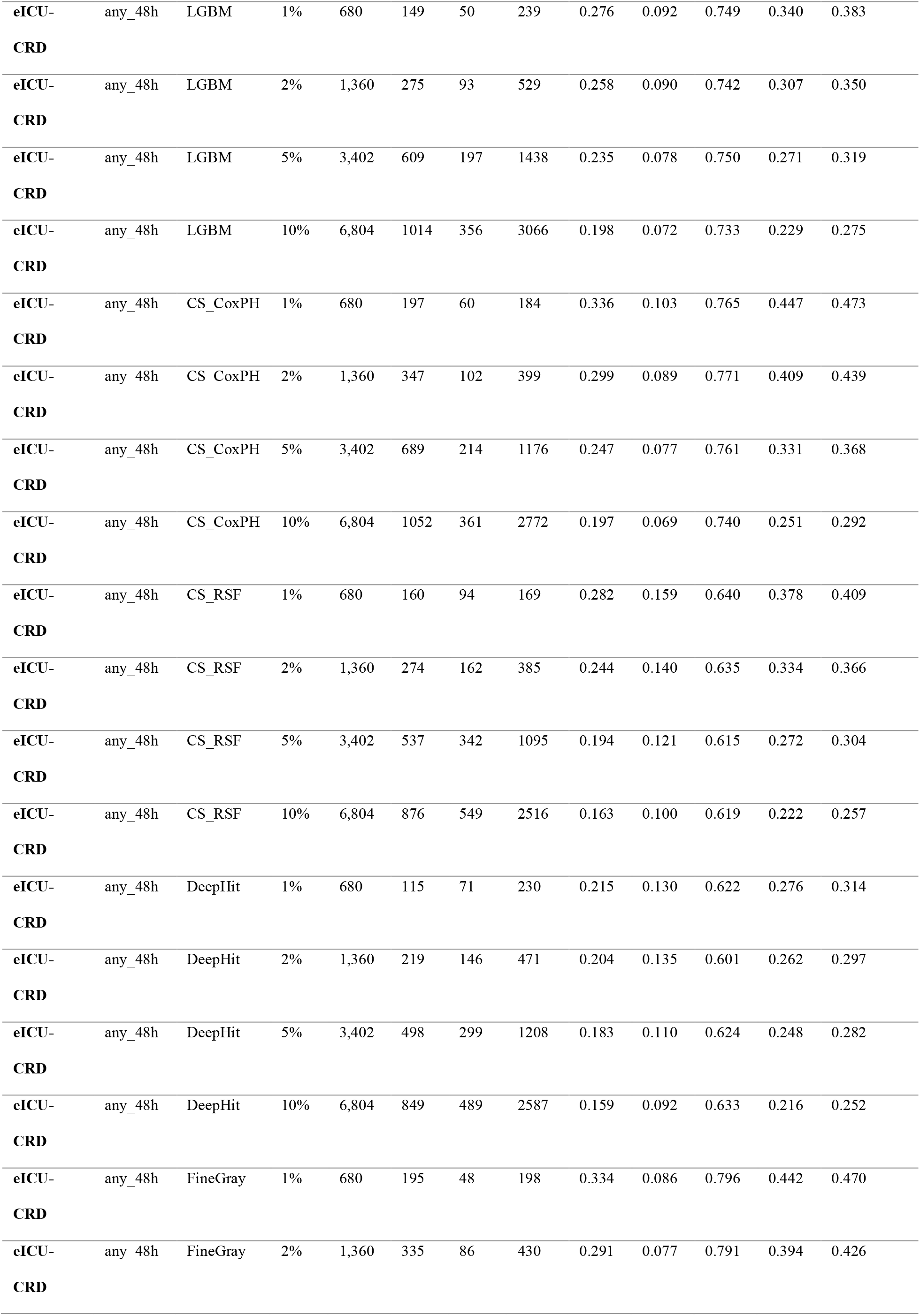
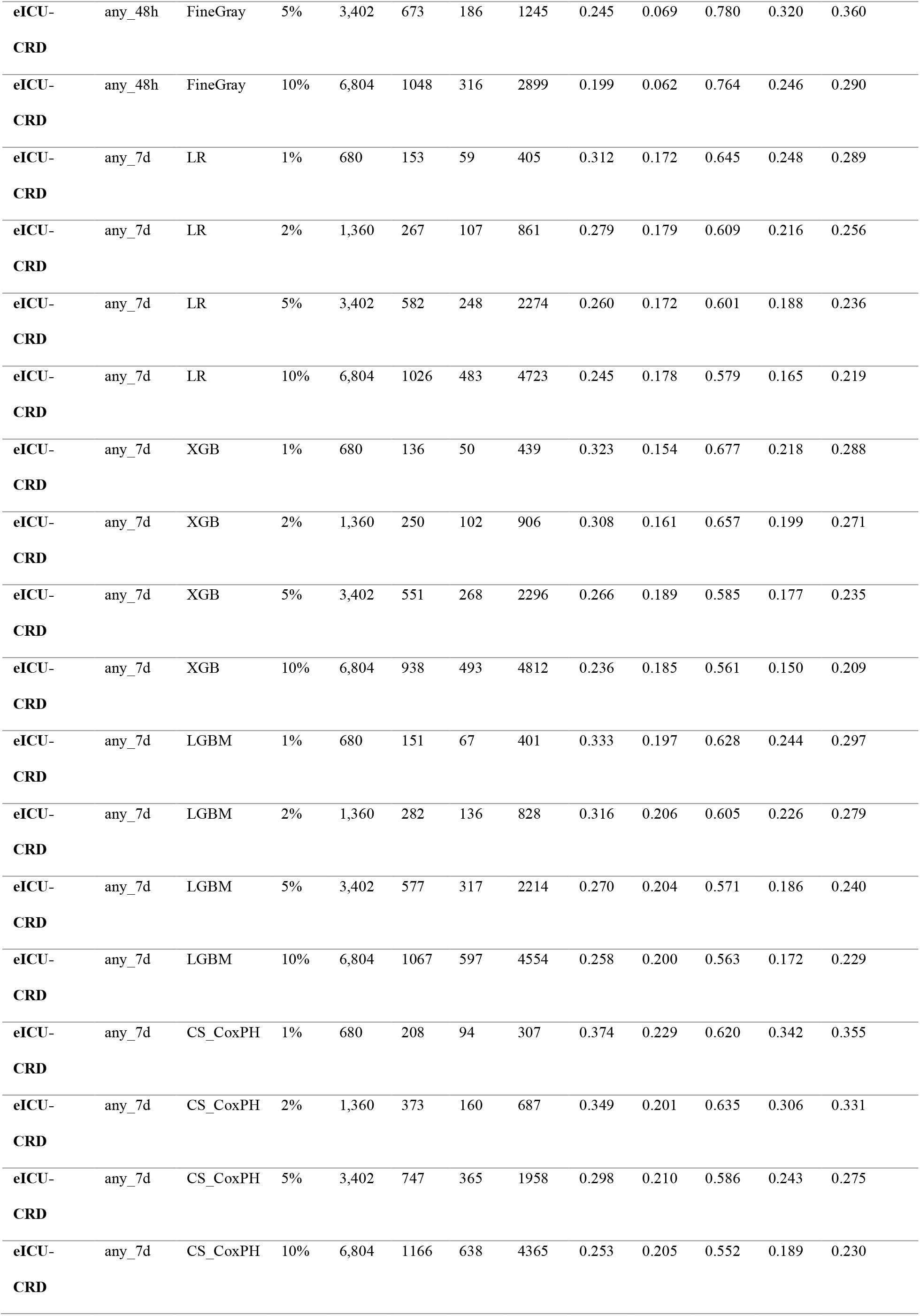
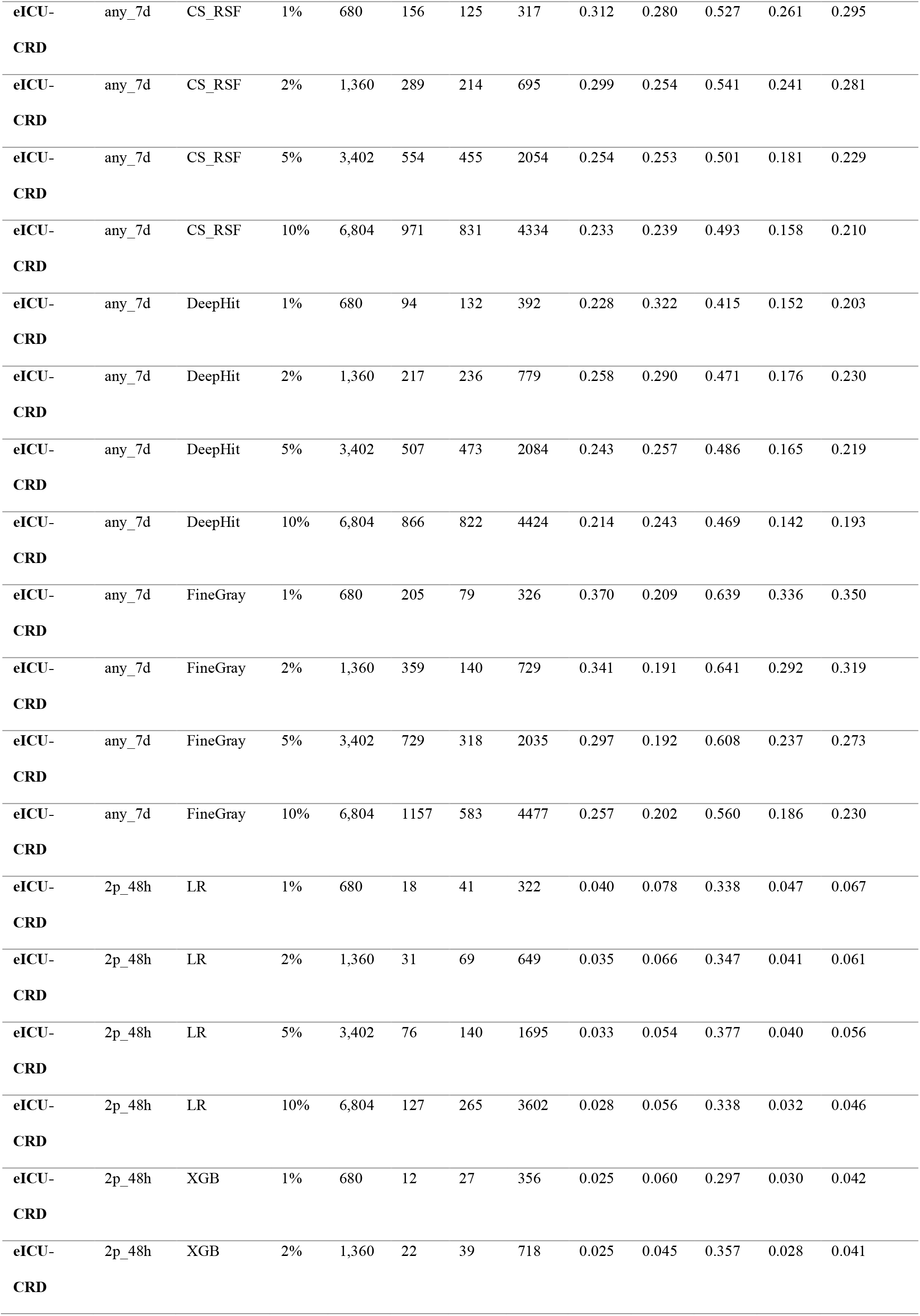
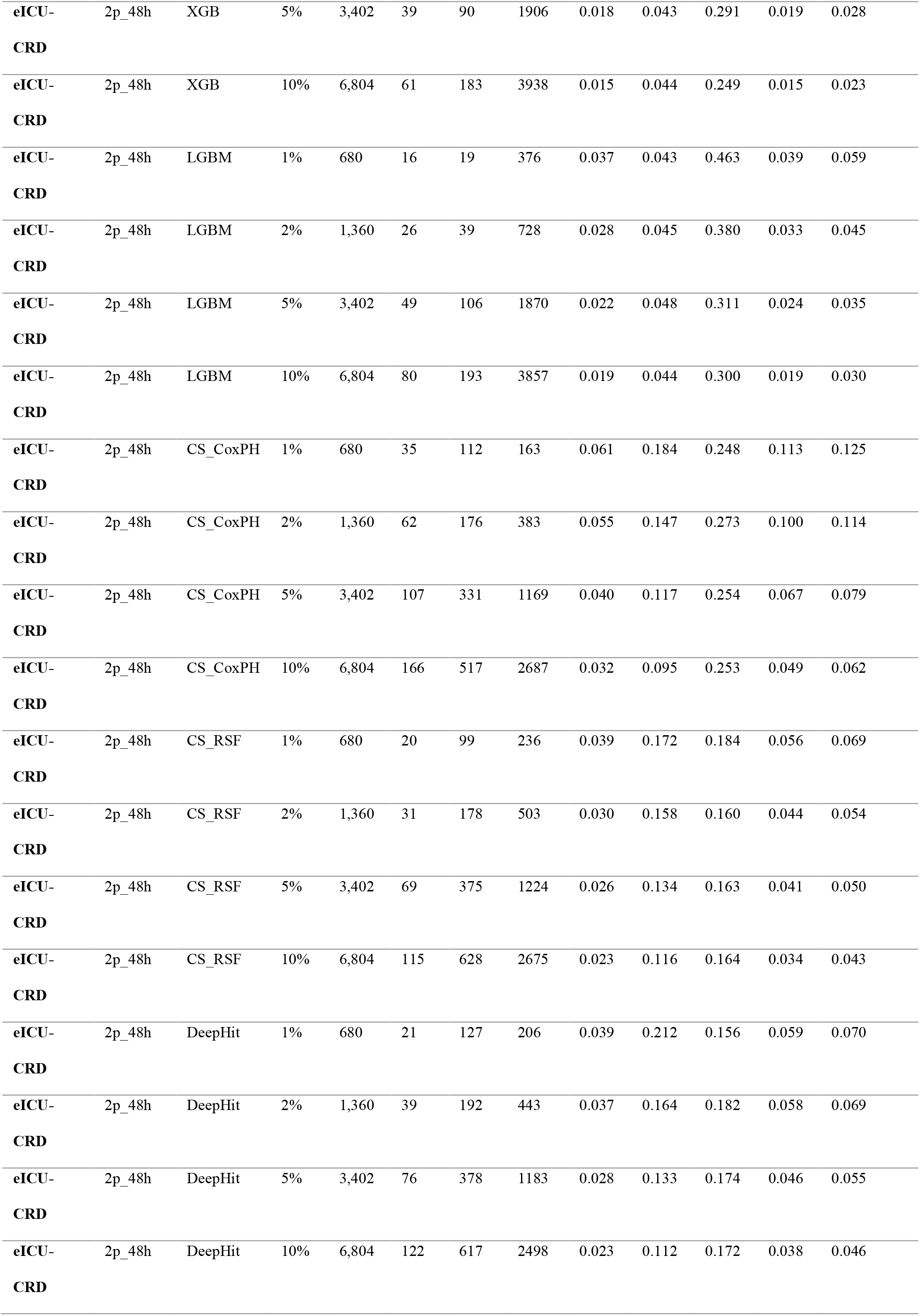
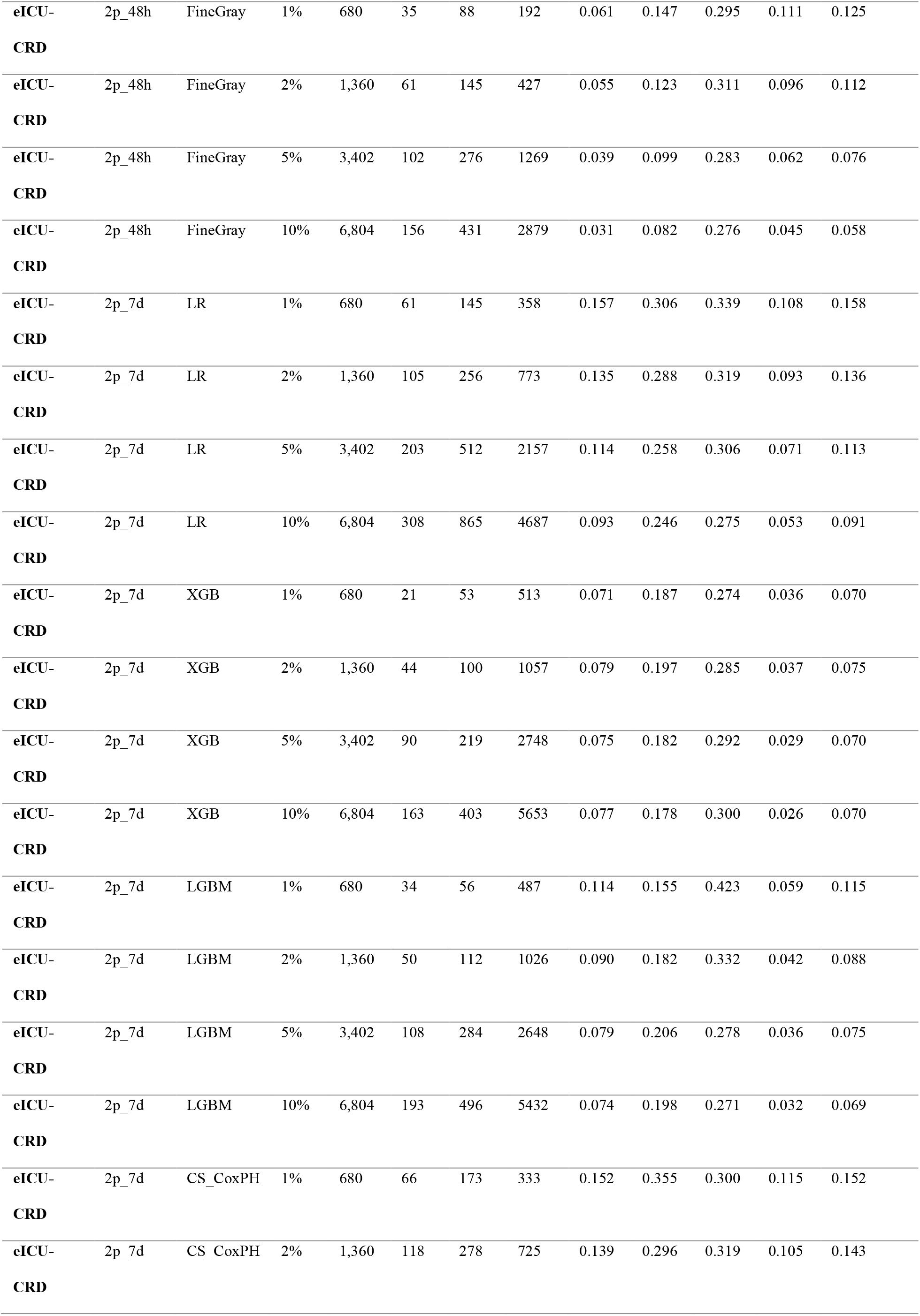
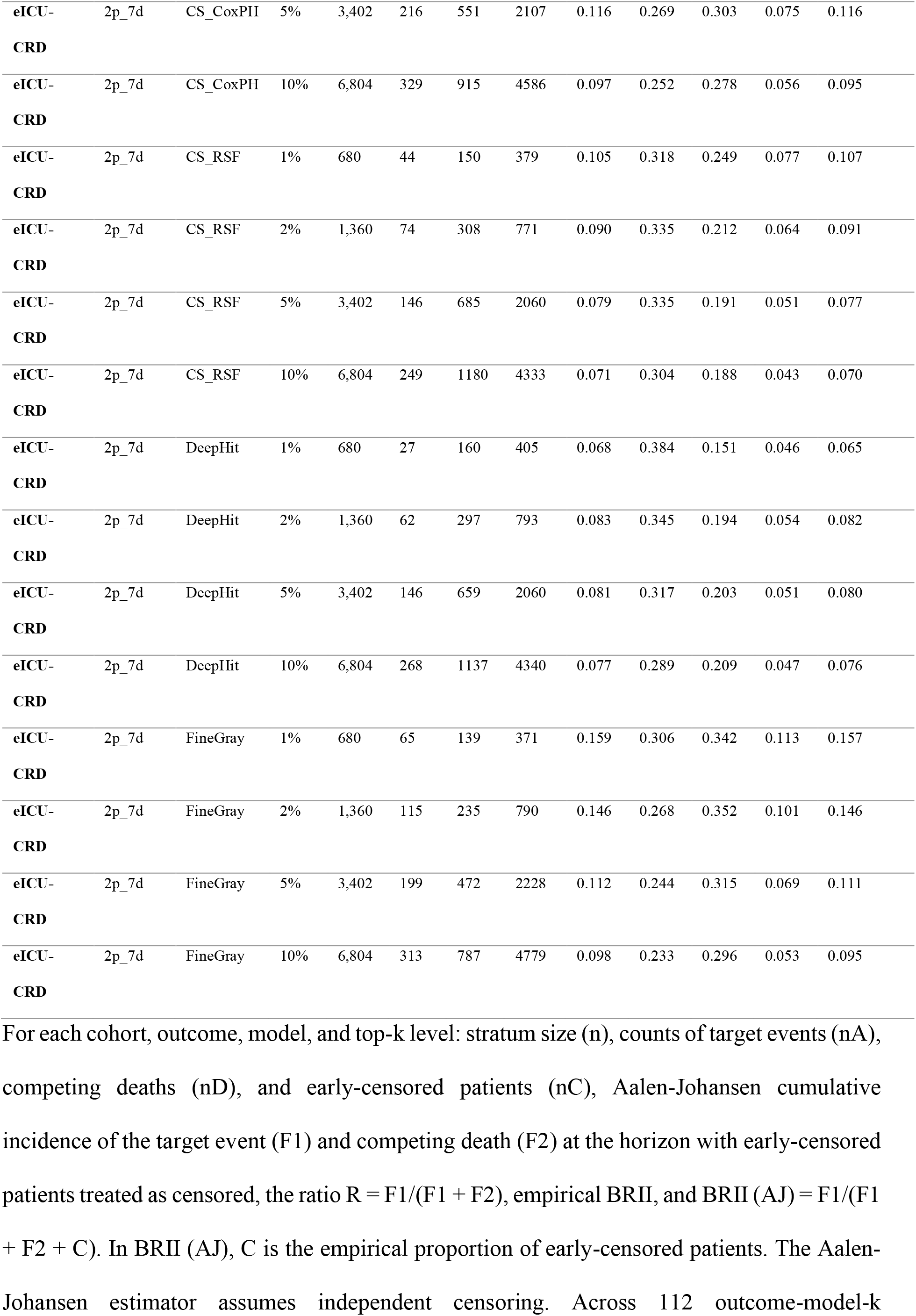
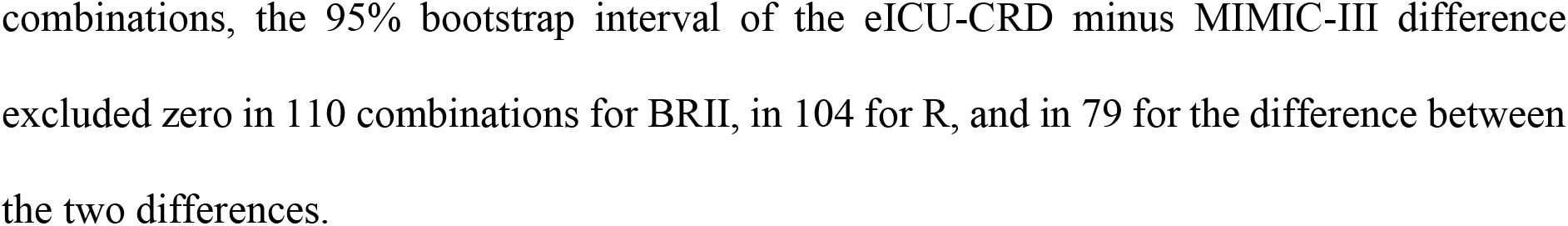
Within-stratum Aalen-Johansen cumulative incidences and BRII.

**Supplementary Table 15.**
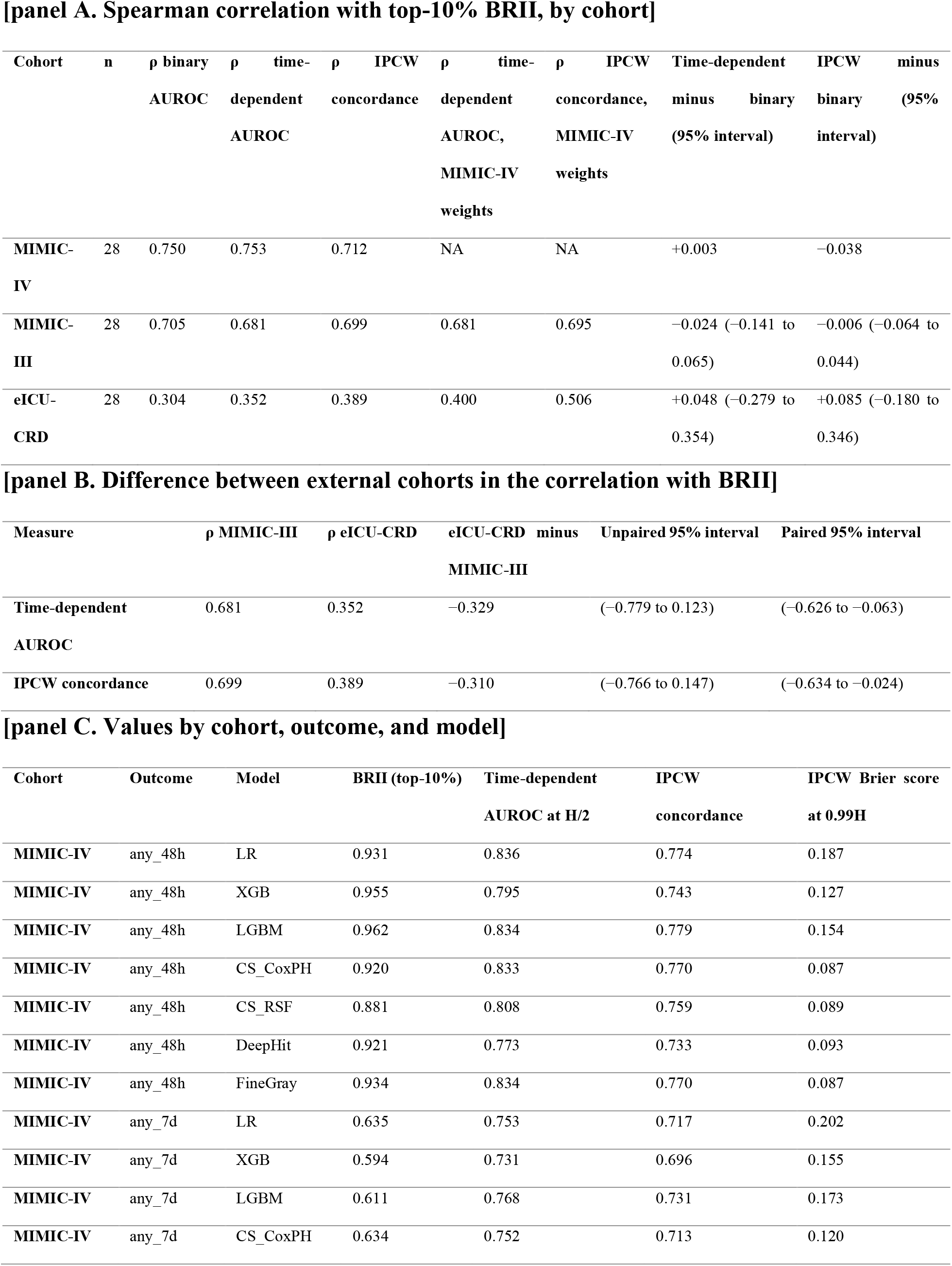
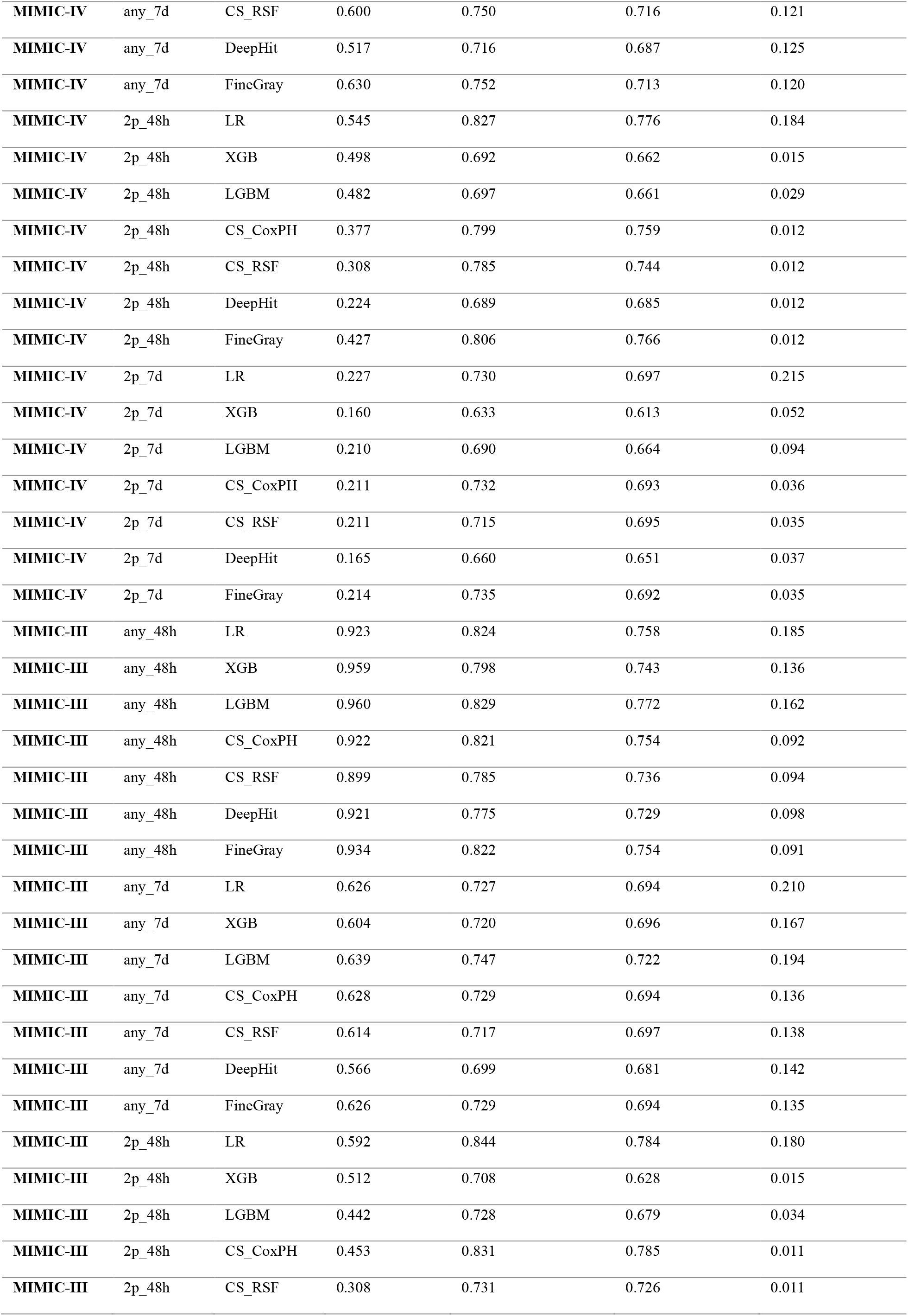
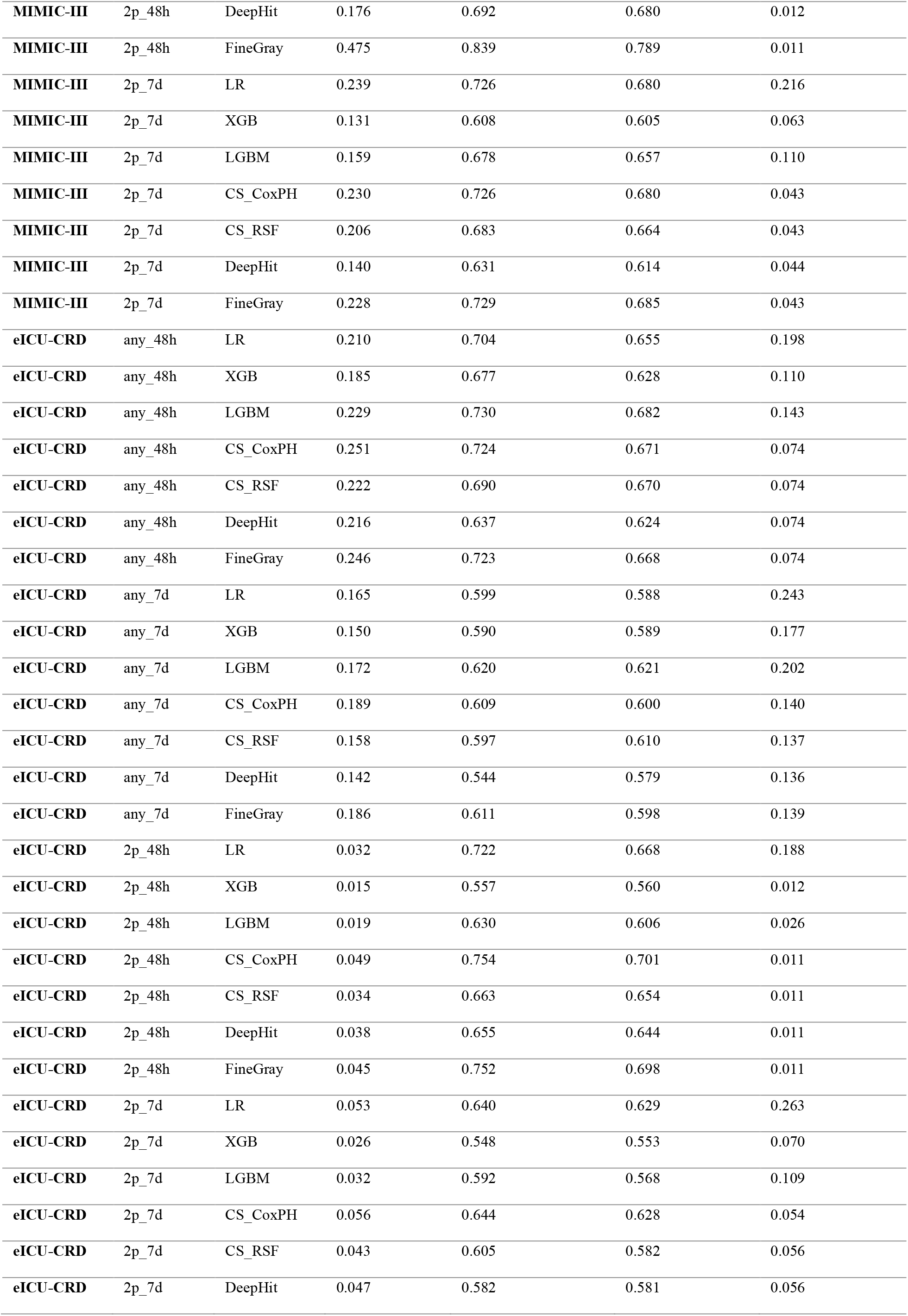
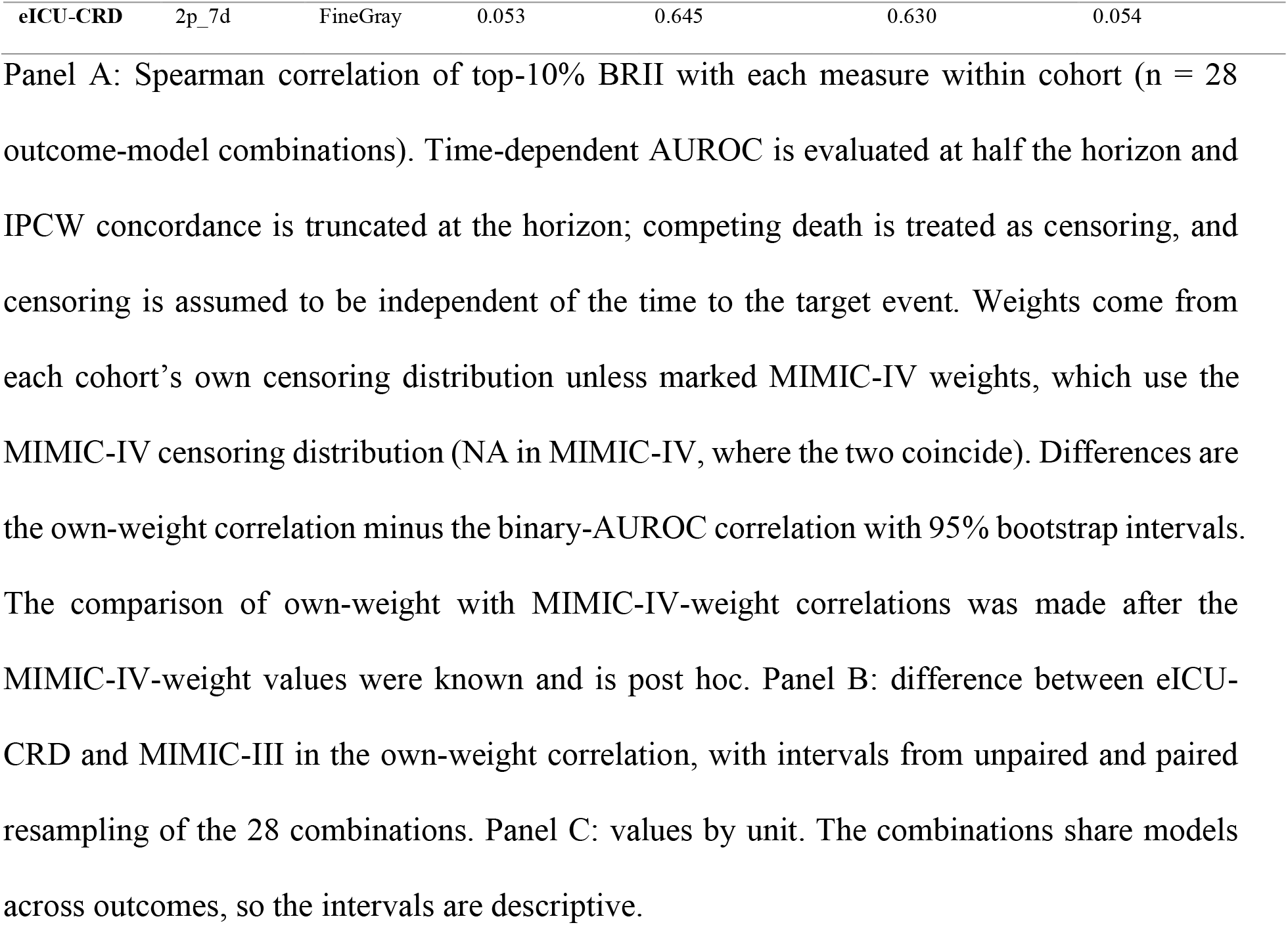
Correlation of BRII with time-dependent AUROC, IPCW concordance, and binary AUROC.

**Supplementary Table 16.** Death-contamination sweep under score-dependent removal.

**[panel A. Summary by removal design]**
| Removal design | Combinations with a less steep top-10% slope | Mean slope difference (top minus bottom) | Combinations with a steeper or equal top-10% slope |
| --- | --- | --- | --- |
| Score-independent removal (primary design) | 24 of 28 | 0.129 | 4 |
| Toward higher scores, linear weight, LightGBM score | 2 of 28 | −0.172 | 26 |
| Toward higher scores, cubic weight, LightGBM score | 1 of 28 | −0.285 | 27 |
| Toward higher scores, linear weight, own model score | 1 of 28 | −0.263 | 27 |
| Toward lower scores, linear weight, LightGBM score | 28 of 28 | 0.495 | 0 |
| Toward lower scores, cubic weight, LightGBM score | 28 of 28 | 0.524 | 0 |
| Toward lower scores, linear weight, own model score | 28 of 28 | 0.667 | 0 |

**[panel B. Slope difference (top-10% minus bottom 90%) by outcome and model]**
| Outcome | Model | Independent | High, linear | High, cubic | High, own | Low, linear | Low, cubic | Low, own |
| --- | --- | --- | --- | --- | --- | --- | --- | --- |
| any_48h | LR | 0.193 | −0.259 | −0.401 | −0.365 | 0.574 | 0.569 | 0.573 |
| any_48h | XGB | 0.341 | −0.251 | −0.448 | −0.375 | 0.750 | 0.730 | 0.753 |
| any_48h | LGBM | 0.317 | −0.291 | −0.449 | −0.289 | 0.697 | 0.633 | 0.697 |
| any_48h | CS_CoxPH | 0.118 | −0.284 | −0.397 | −0.376 | 0.505 | 0.503 | 0.504 |
| any_48h | CS_RSf | 0.085 | −0.290 | −0.356 | −0.338 | 0.521 | 0.505 | 0.517 |
| any_48h | DeepHit | 0.129 | −0.267 | −0.365 | −0.381 | 0.560 | 0.551 | 0.574 |
| any_48h | FineGray | 0.165 | −0.271 | −0.400 | −0.380 | 0.556 | 0.552 | 0.550 |
| any_7d | LR | 0.134 | −0.255 | −0.399 | −0.361 | 0.591 | 0.618 | 0.711 |
| any_7d | XGB | 0.231 | −0.267 | −0.455 | −0.359 | 0.758 | 0.781 | 0.817 |
| any_7d | LGBM | 0.218 | −0.309 | −0.477 | −0.307 | 0.781 | 0.788 | 0.781 |
| any_7d | CS_CoxPH | 0.120 | −0.274 | −0.416 | −0.375 | 0.580 | 0.609 | 0.687 |
| any_7d | CS_RSf | 0.095 | −0.289 | −0.413 | −0.328 | 0.611 | 0.640 | 0.682 |
| any_7d | DeepHit | 0.138 | −0.268 | −0.403 | −0.375 | 0.662 | 0.689 | 0.760 |
| any_7d | FineGray | 0.149 | −0.269 | −0.422 | −0.380 | 0.612 | 0.640 | 0.712 |
| 2p_48h | LR | 0.213 | 0.080 | 0.025 | −0.003 | 0.427 | 0.472 | 0.787 |
| **2p_48h** | XGB | 0.383 | 0.109 | −0.055 | 0.068 | 0.725 | 0.780 | 0.890 |
| **2p_48h** | LGBM | 0.257 | −0.056 | −0.232 | −0.058 | 0.822 | 0.942 | 0.825 |
| **2p_48h** | CS_CoxPH | 0.033 | −0.061 | −0.104 | −0.184 | 0.202 | 0.242 | 0.635 |
| **2p_48h** | CS_RSF | −0.010 | −0.040 | −0.068 | −0.235 | 0.149 | 0.188 | 0.556 |
| **2p_48h** | DeepHit | −0.078 | −0.126 | −0.144 | −0.263 | 0.058 | 0.092 | 0.436 |
| **2p_48h** | FineGray | 0.076 | −0.023 | −0.072 | −0.142 | 0.241 | 0.279 | 0.682 |
| **2p_7d** | LR | 0.023 | −0.111 | −0.178 | −0.234 | 0.274 | 0.320 | 0.654 |
| **2p_7d** | XGB | 0.157 | −0.071 | −0.222 | −0.169 | 0.535 | 0.595 | 0.787 |
| **2p_7d** | LGBM | 0.153 | −0.136 | −0.326 | −0.138 | 0.776 | 0.896 | 0.776 |
| **2p_7d** | CS_CoxPH | 0.020 | −0.115 | −0.180 | −0.243 | 0.270 | 0.317 | 0.656 |
| **2p_7d** | CS_RSF | −0.030 | −0.171 | −0.239 | −0.283 | 0.220 | 0.269 | 0.556 |
| **2p_7d** | DeepHit | −0.067 | −0.164 | −0.208 | −0.272 | 0.090 | 0.118 | 0.445 |
| **2p_7d** | FineGray | 0.045 | −0.092 | −0.162 | −0.220 | 0.310 | 0.356 | 0.686 |

MIMIC-IV, top-10% threshold, 200 repetitions per death-fraction level. Panel A: for each removal design, the number of the 28 model-outcome combinations in which the slope of $A/(A$ $+ D)$ against the target death fraction was less steep in the top-10% stratum than in the bottom 90% stratum, the mean slope difference, and the number of remaining combinations. Panel B: slope difference by outcome and model; column labels give the direction of removal (toward higher or lower scores), the weight (linear or cubic), and the reference score (LightGBM unless marked own). In the score-independent design, the four combinations without a less steep top-10% slope were CS\_RSF and DeepHit for 2p\_48h and 2p\_7d.

**Supplementary Table 17.**
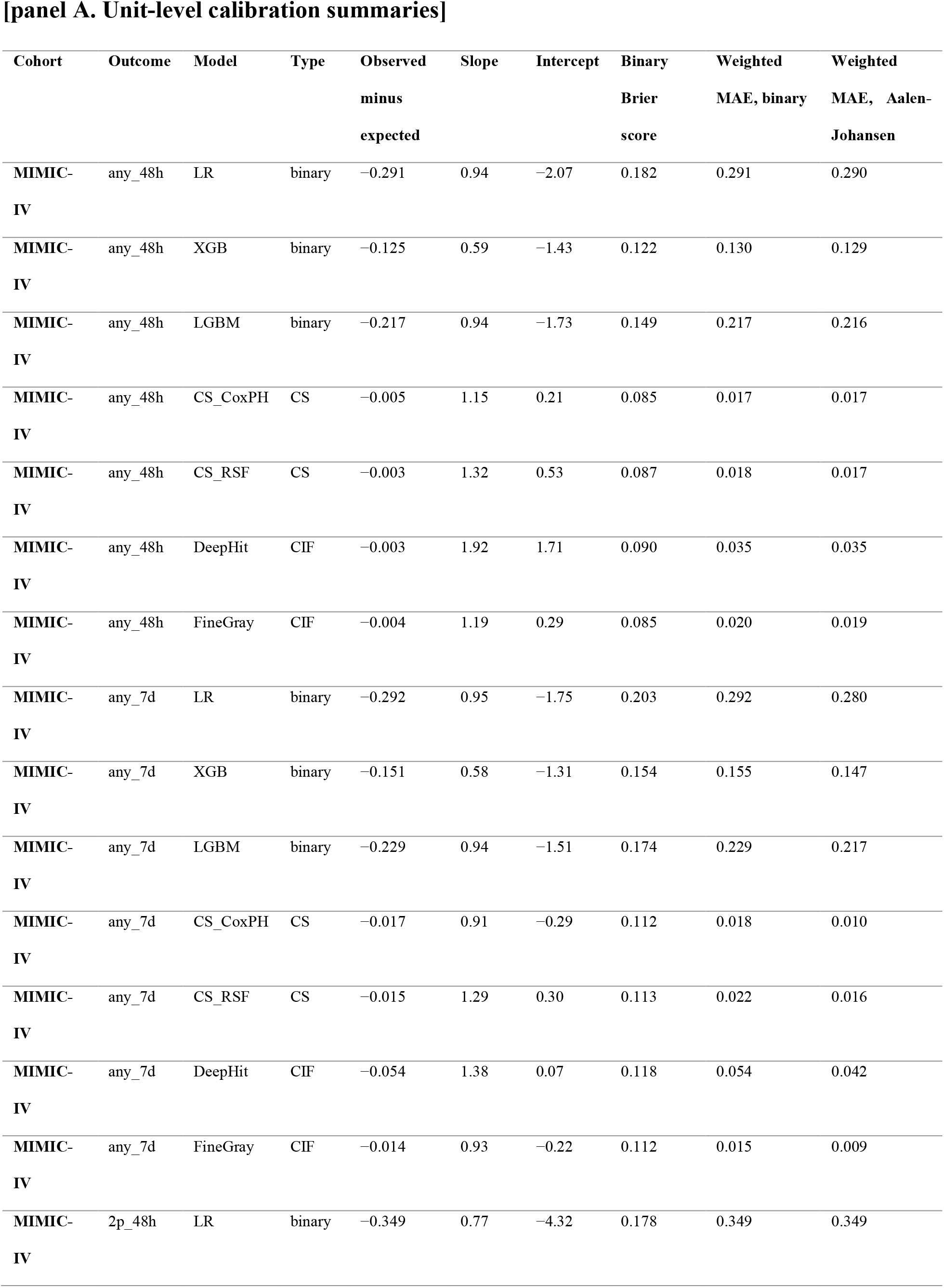
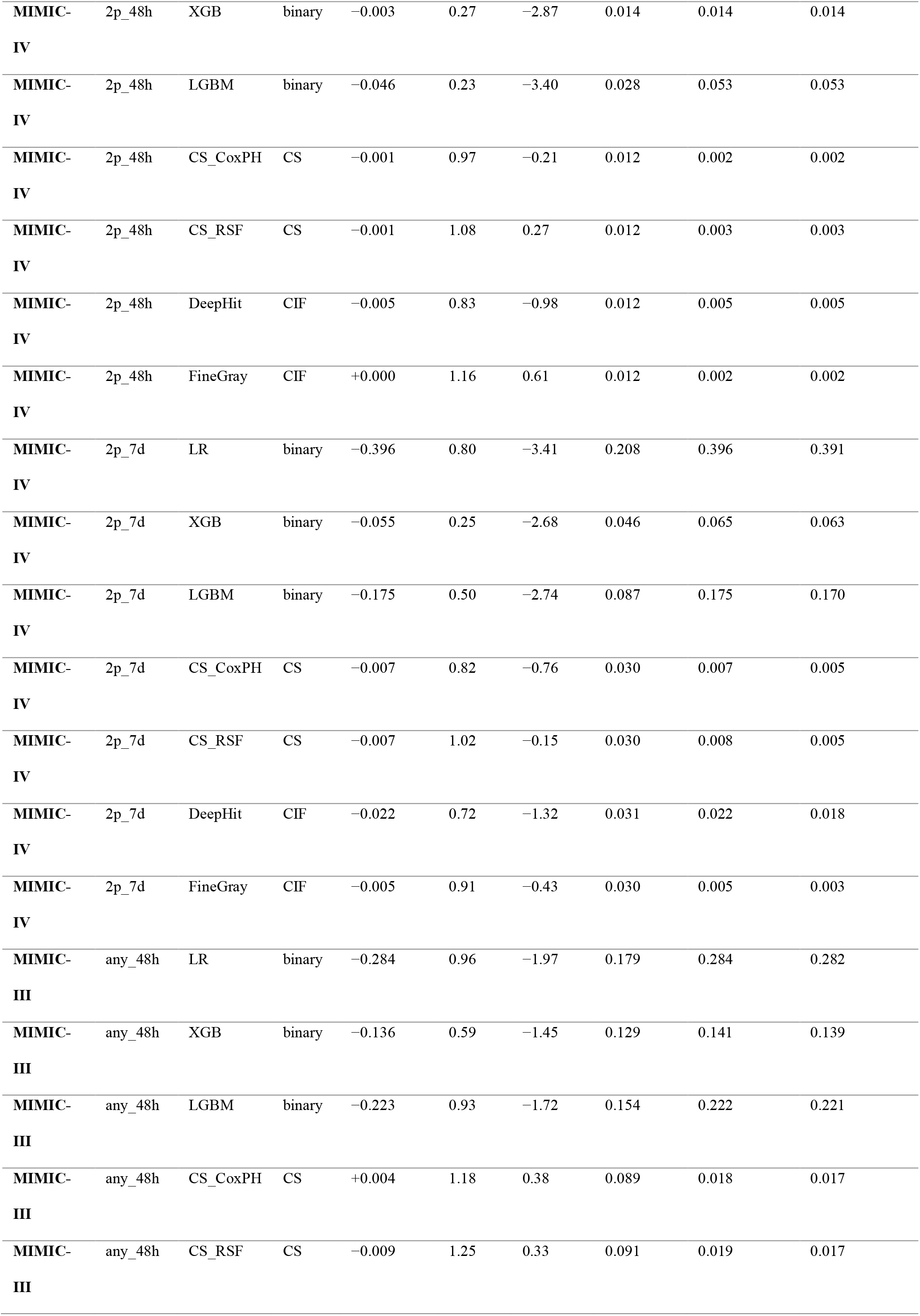
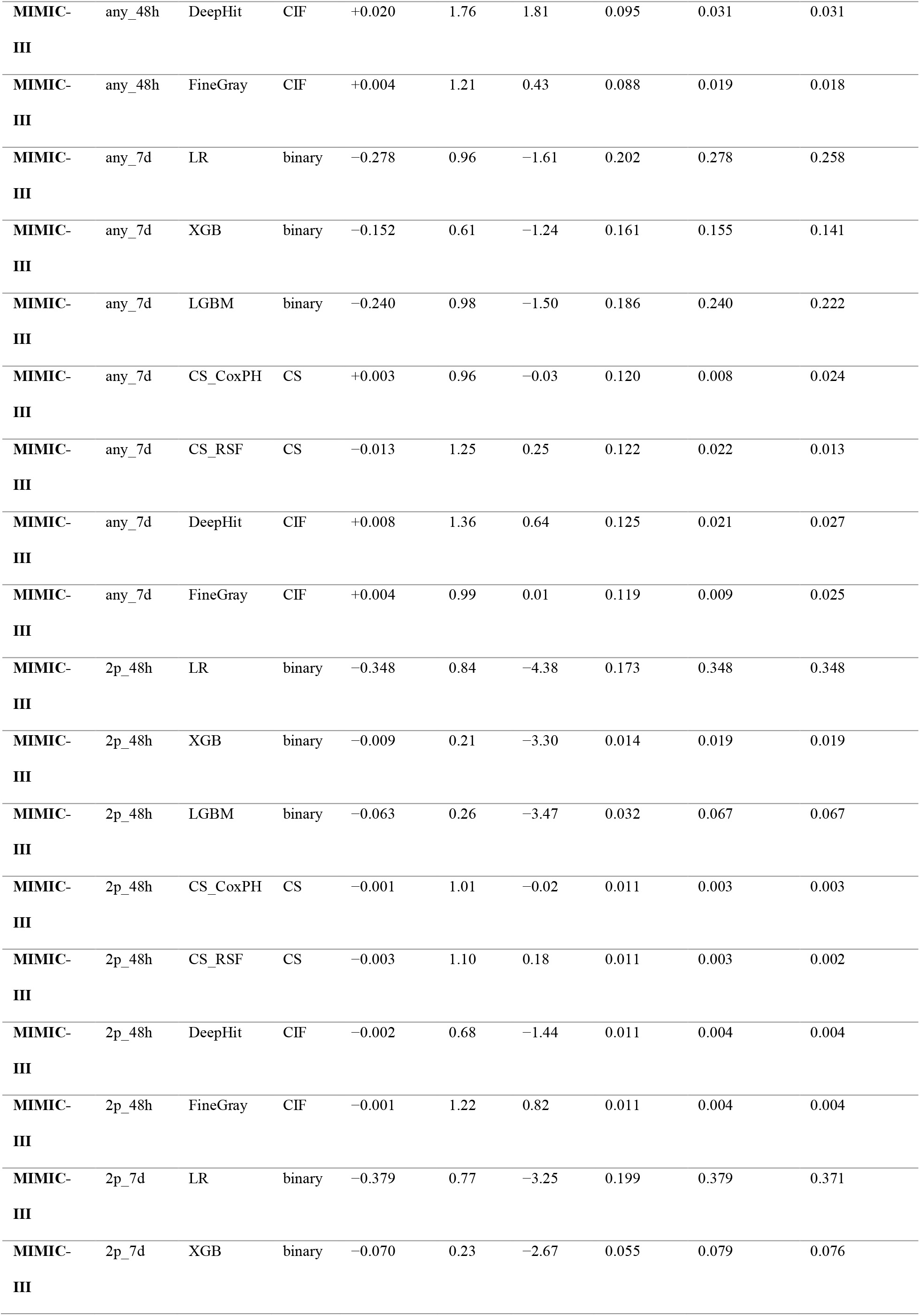
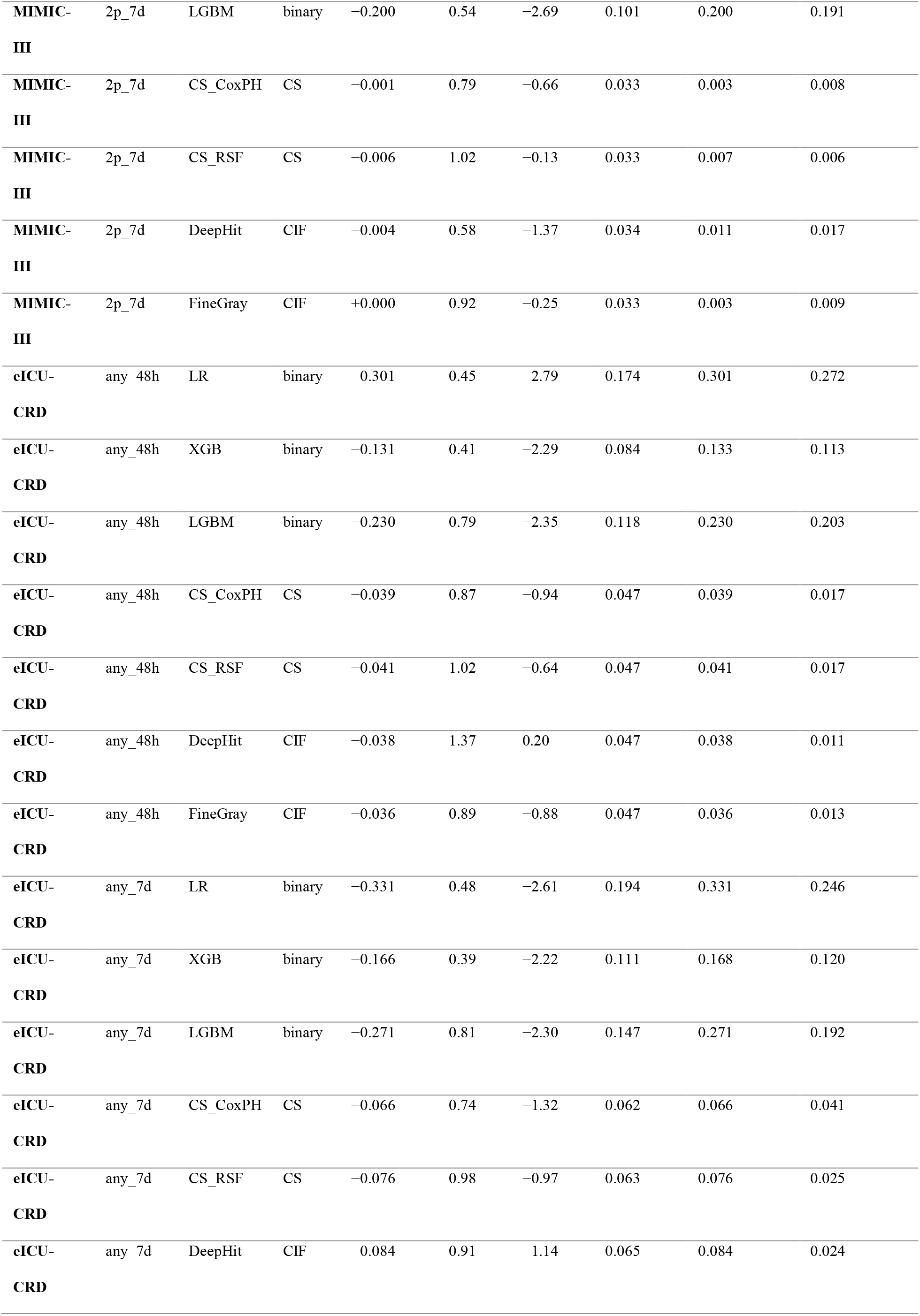
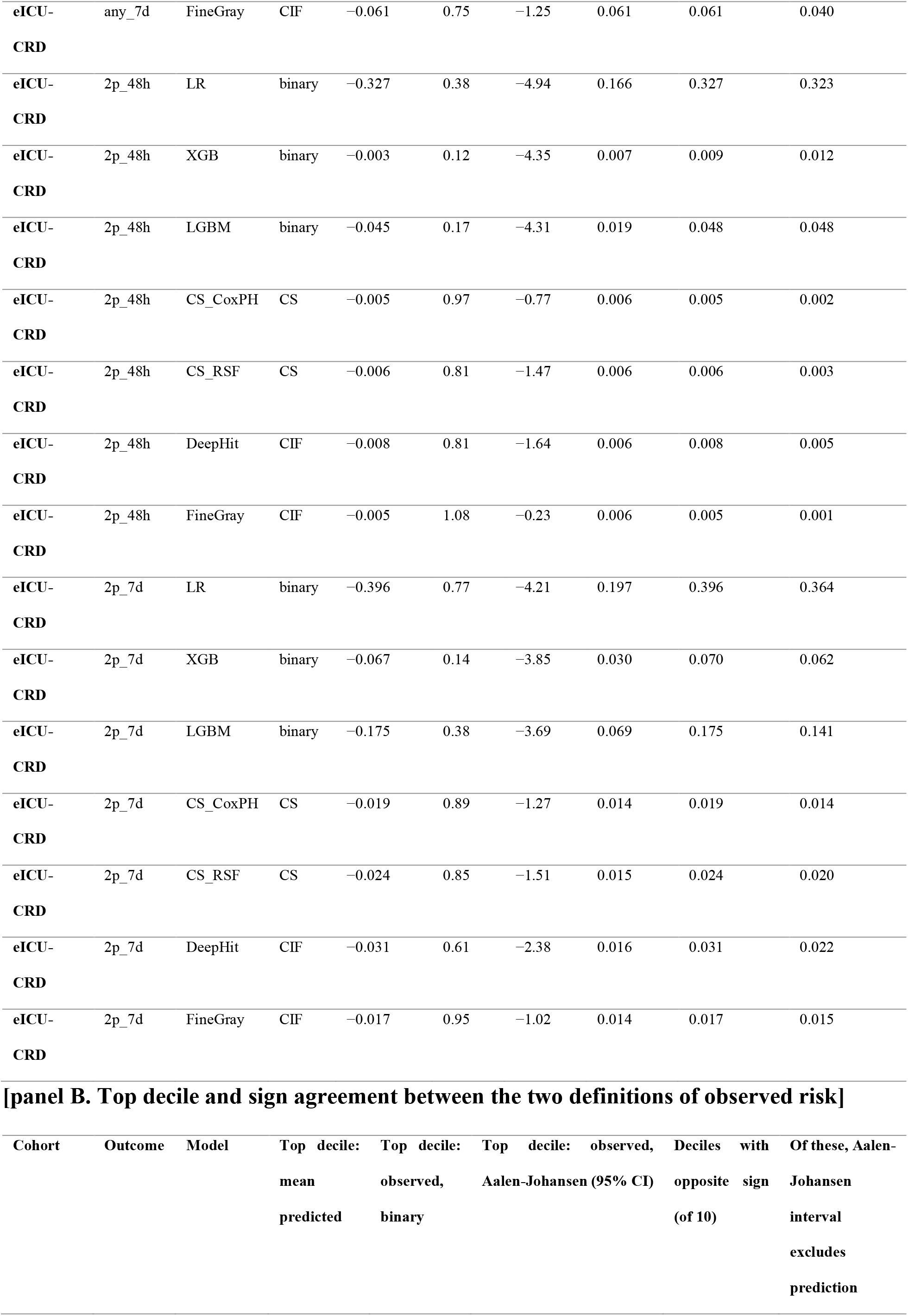
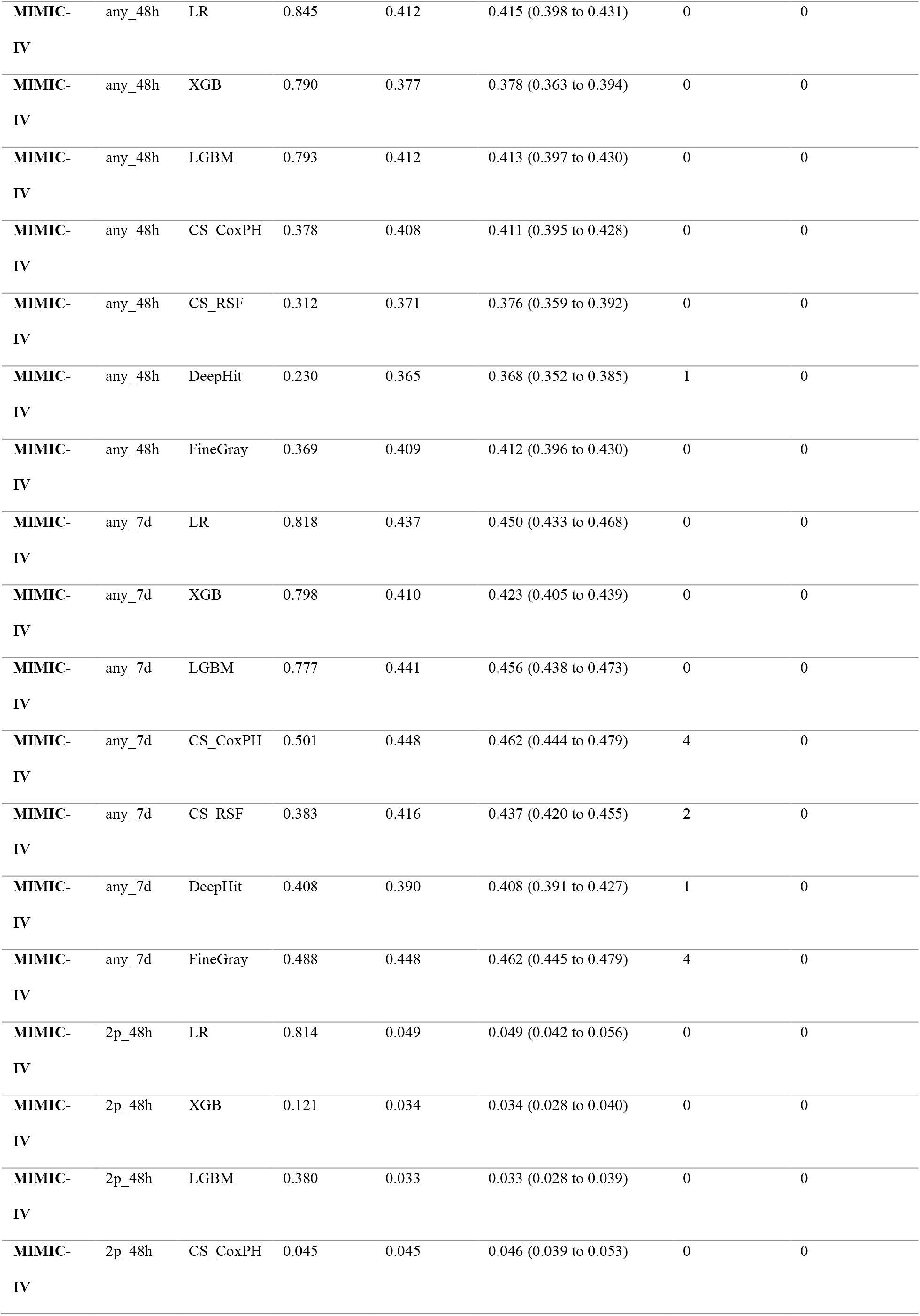
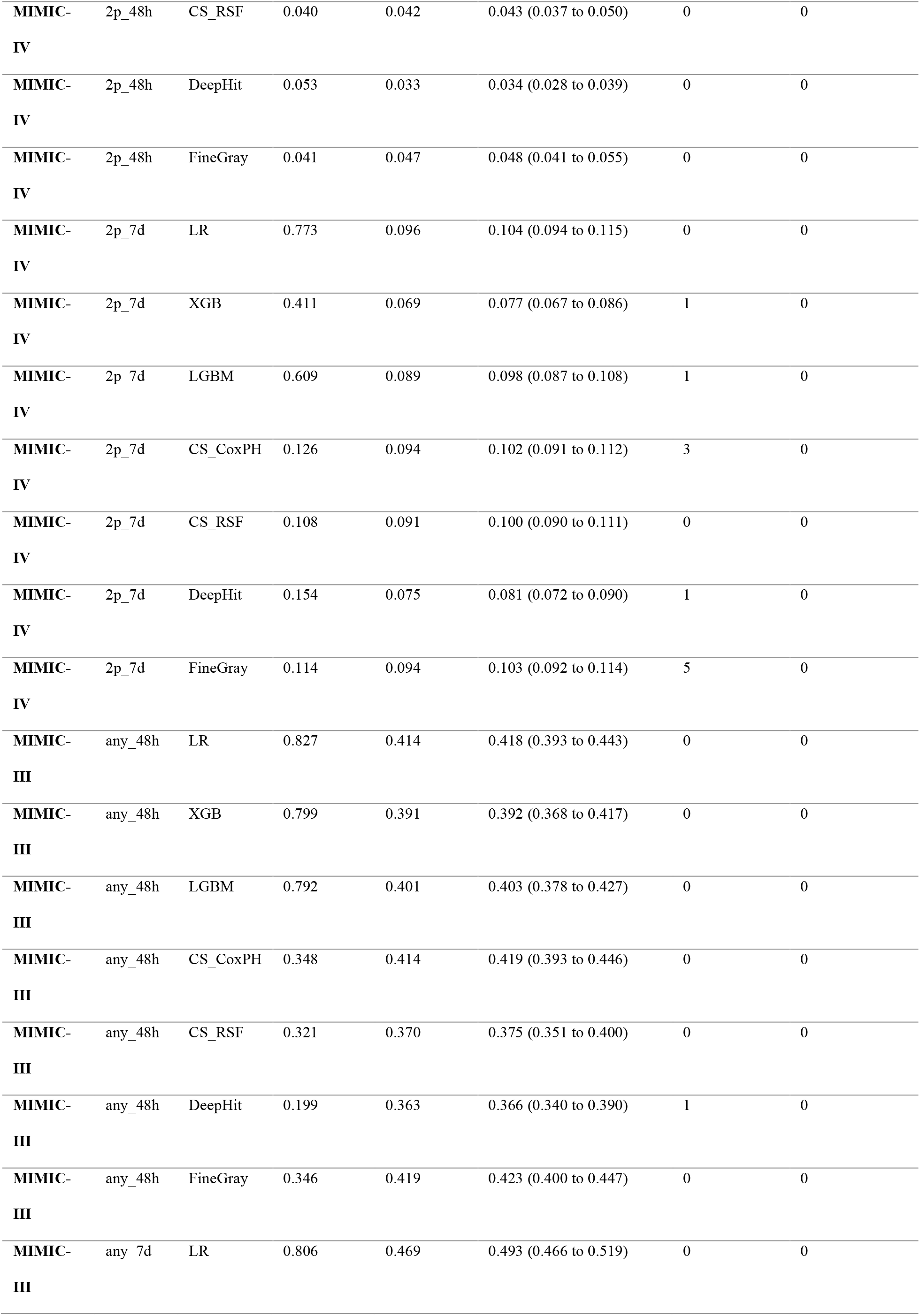
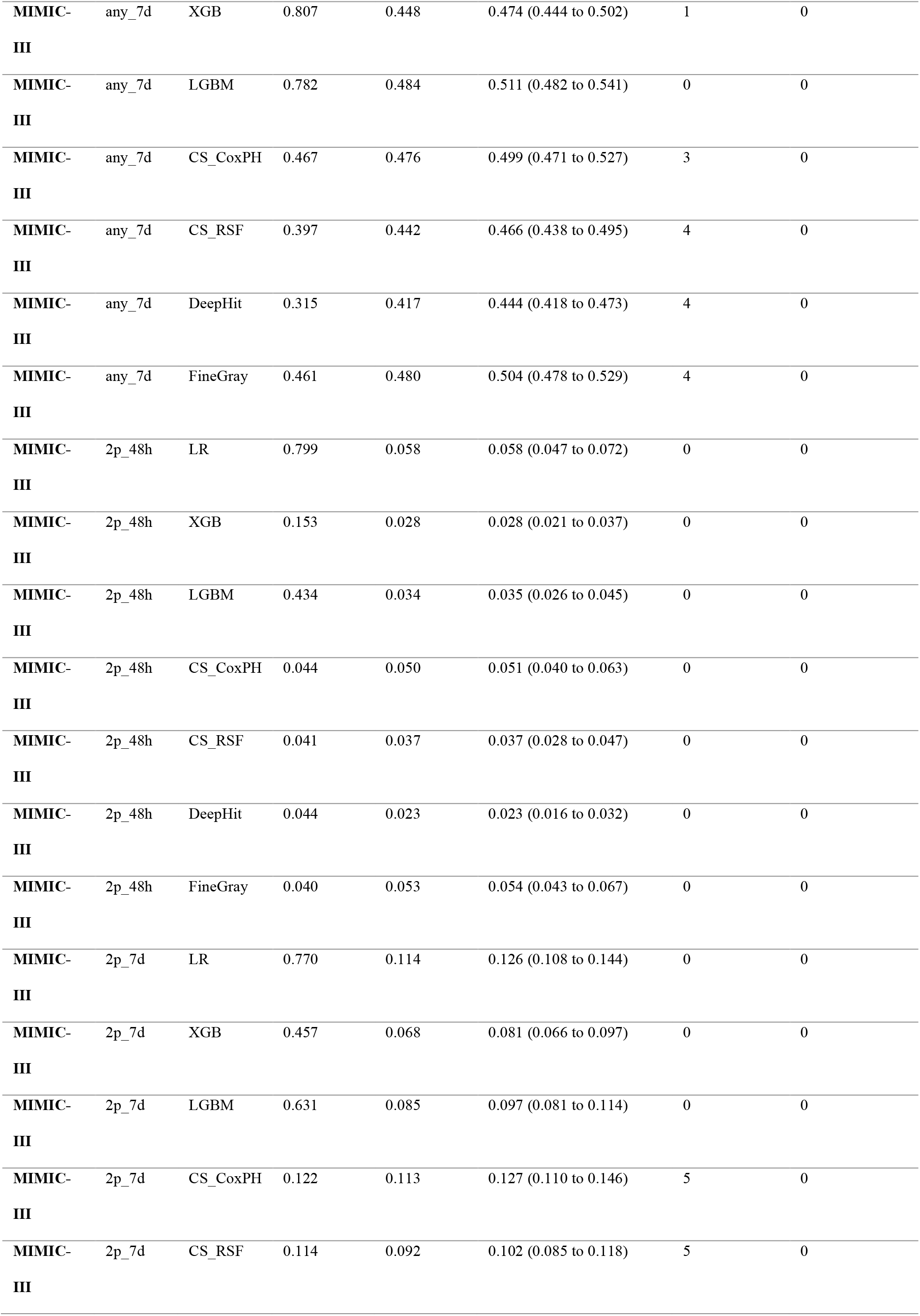
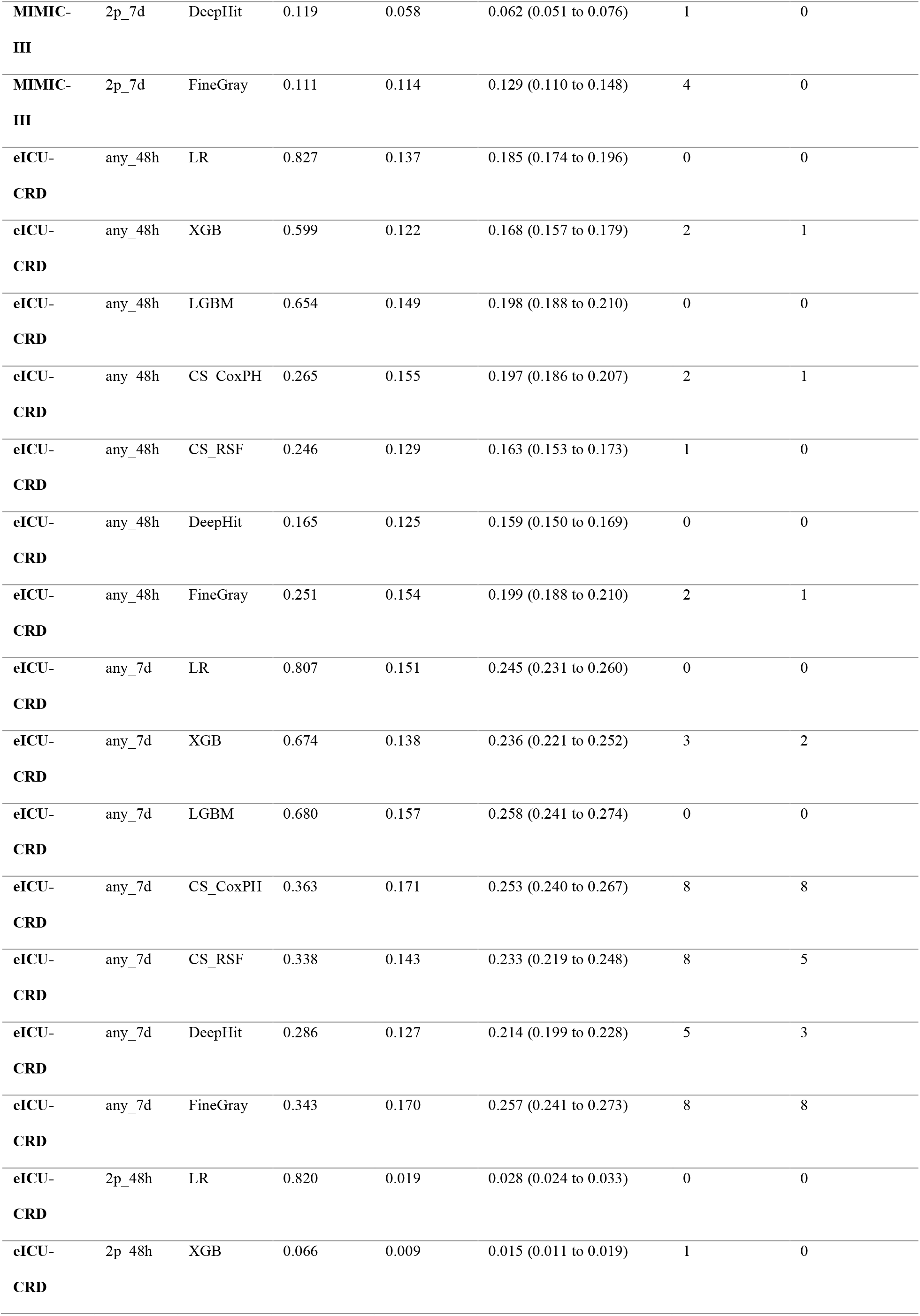
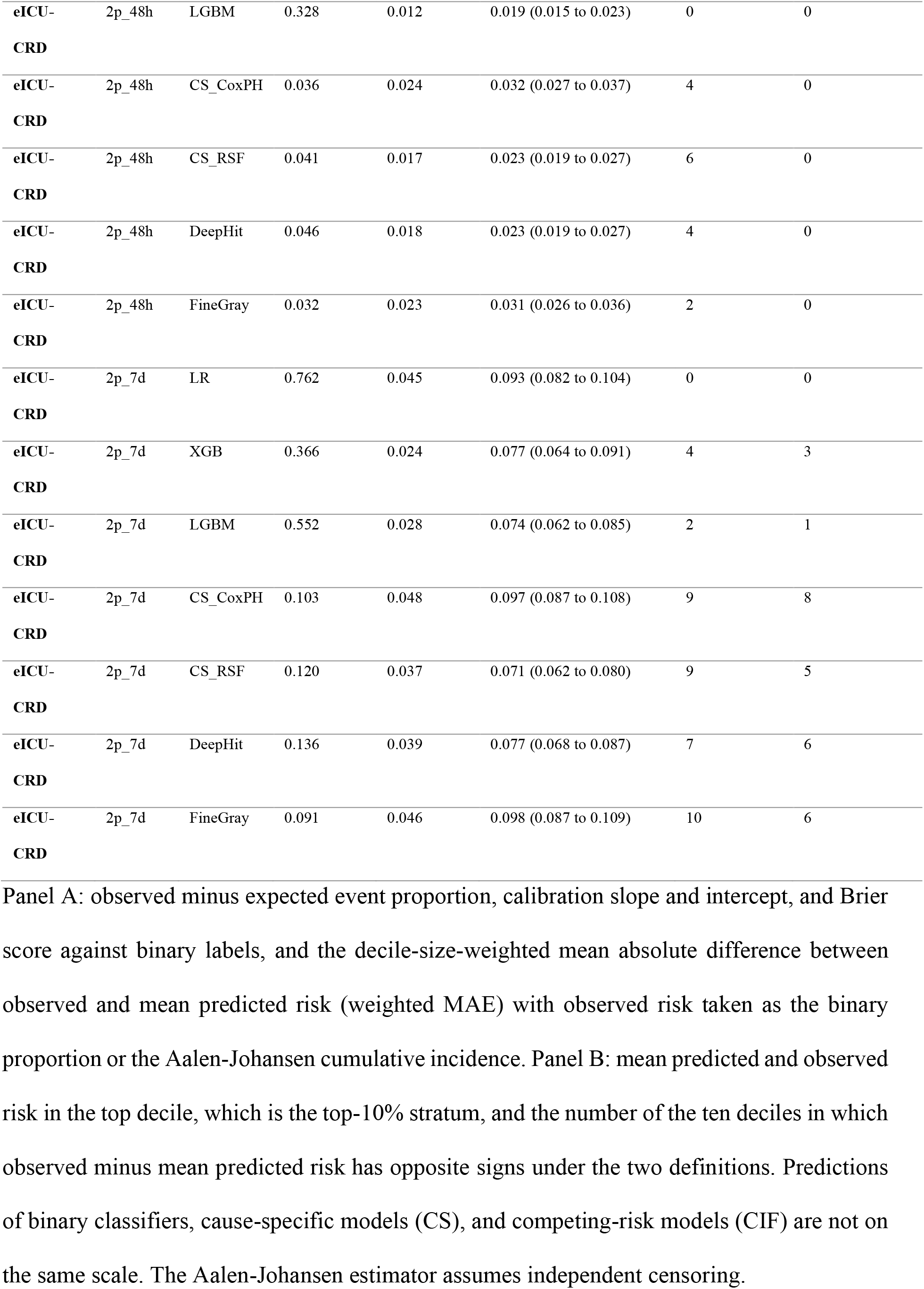
Decile calibration with binary and Aalen-Johansen observed risk.

**Supplementary Table 18.**
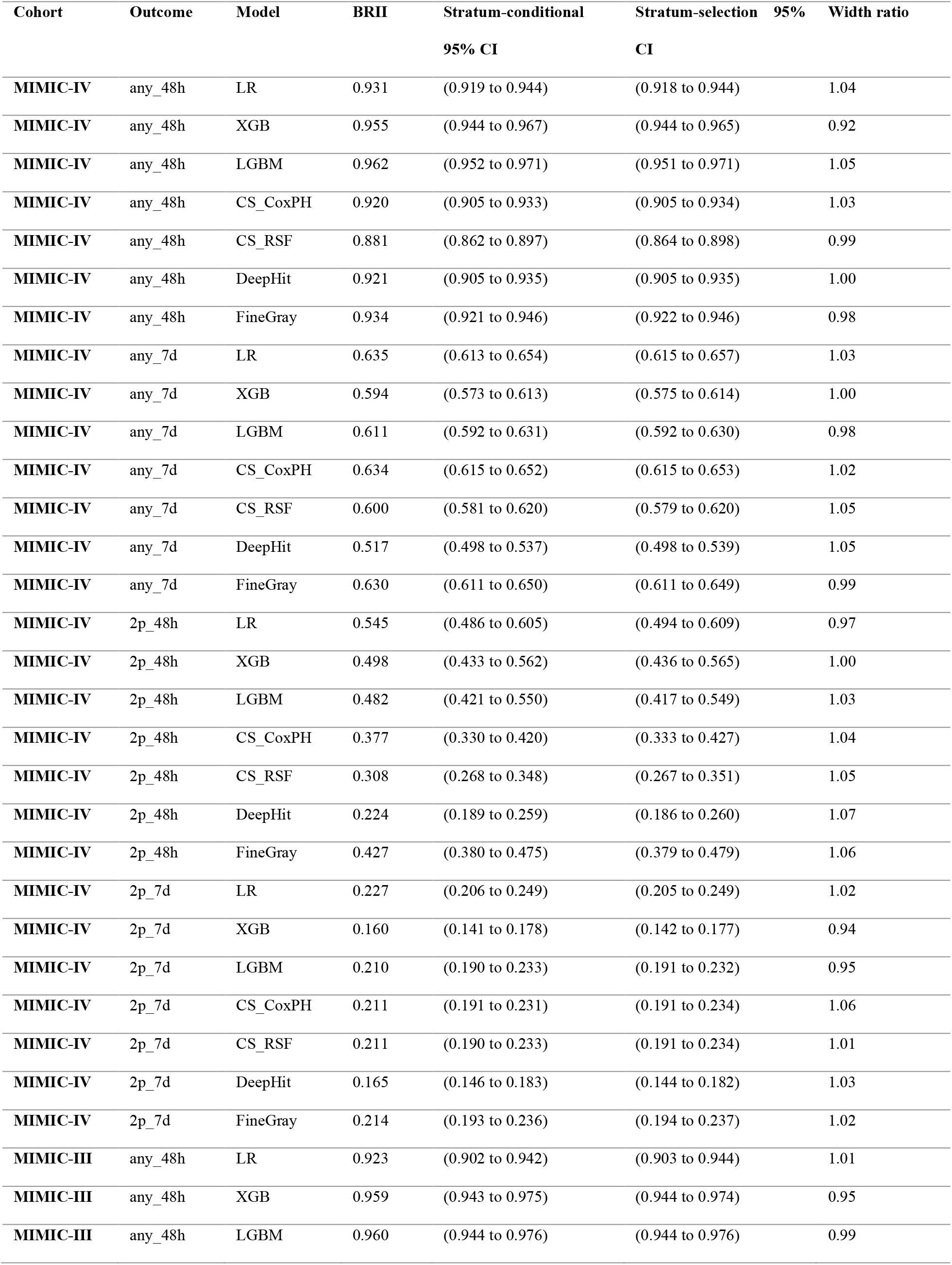
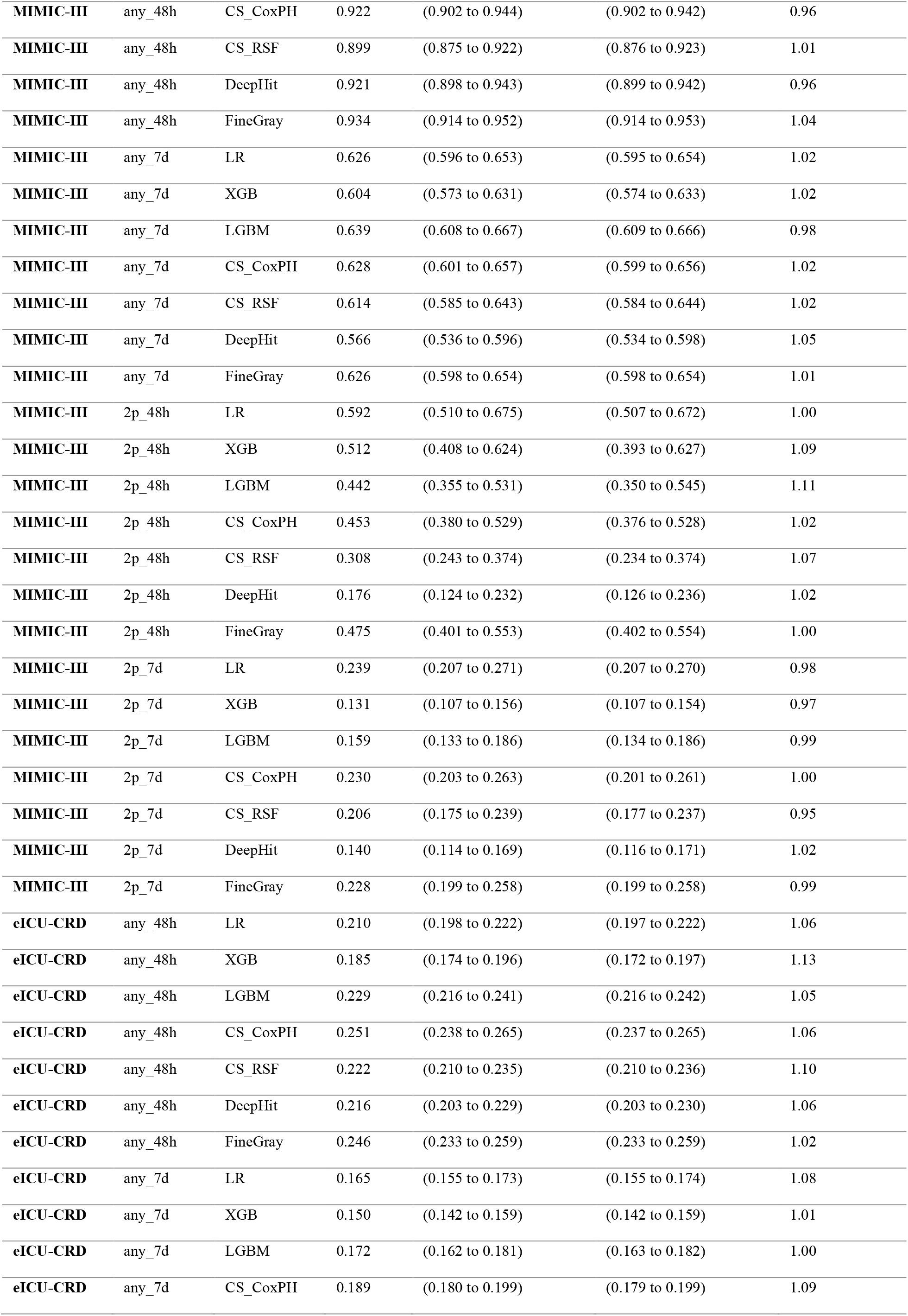
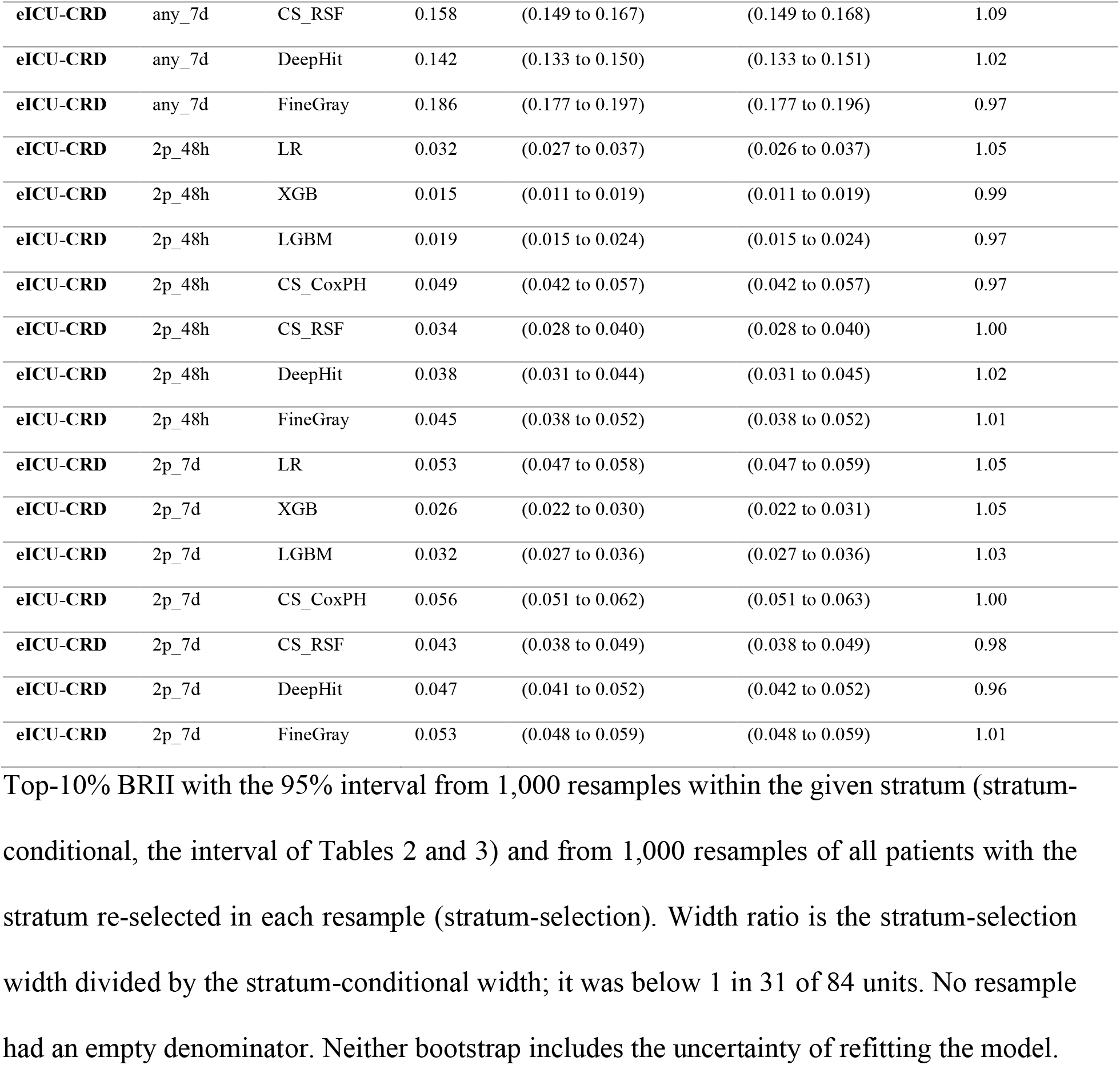
Stratum-conditional and stratum-selection bootstrap intervals for BRII.

**Supplementary Table 19.** Values of the target-event share under extreme assignments of early-censored patients.

| Cohort | Outcome | Model | A/(A+D+C) | A/(A+D) | (A+C)/(A+D+C) | C/(A+D+C) |
| --- | --- | --- | --- | --- | --- | --- |
| MIMIC-III | any_48h | LR | 0.923 | 0.962 | 0.963 | 0.040 |
| MIMIC-III | any_48h | XGB | 0.959 | 0.978 | 0.978 | 0.019 |
| MIMIC-III | any_48h | LGBM | 0.960 | 0.978 | 0.979 | 0.018 |
| MIMIC-III | any_48h | CS_CoxPH | 0.922 | 0.960 | 0.962 | 0.040 |
| MIMIC-III | any_48h | CS_RSf | 0.899 | 0.939 | 0.941 | 0.042 |
| MIMIC-III | any_48h | DeepHit | 0.921 | 0.953 | 0.954 | 0.033 |
| MIMIC-III | any_48h | FineGray | 0.934 | 0.967 | 0.968 | 0.034 |
| MIMIC-III | any_7d | LR | 0.626 | 0.910 | 0.938 | 0.313 |
| MIMIC-III | any_7d | XGB | 0.604 | 0.943 | 0.964 | 0.360 |
| MIMIC-III | any_7d | LGBM | 0.639 | 0.953 | 0.968 | 0.330 |
| MIMIC-III | any_7d | CS_CoxPH | 0.628 | 0.921 | 0.946 | 0.318 |
| MIMIC-III | any_7d | CS_RSf | 0.614 | 0.904 | 0.935 | 0.321 |
| MIMIC-III | any_7d | DeepHit | 0.566 | 0.900 | 0.937 | 0.371 |
| MIMIC-III | any_7d | FineGray | 0.626 | 0.932 | 0.954 | 0.328 |
| MIMIC-III | 2p_48h | LR | 0.592 | 0.712 | 0.761 | 0.169 |
| MIMIC-III | 2p_48h | XGB | 0.512 | 0.788 | 0.863 | 0.350 |
| MIMIC-III | 2p_48h | LGBM | 0.442 | 0.794 | 0.885 | 0.442 |
| MIMIC-III | 2p_48h | CS_CoxPH | 0.453 | 0.566 | 0.652 | 0.199 |
| MIMIC-III | 2p_48h | CS_RSf | 0.308 | 0.414 | 0.564 | 0.256 |
| MIMIC-III | 2p_48h | DeepHit | 0.176 | 0.231 | 0.412 | 0.235 |
| MIMIC-III | 2p_48h | FineGray | 0.475 | 0.592 | 0.673 | 0.198 |
| MIMIC-III | 2p_7d | LR | 0.239 | 0.554 | 0.808 | 0.569 |
| MIMIC-III | 2p_7d | XGB | 0.131 | 0.649 | 0.929 | 0.798 |
| MIMIC-III | 2p_7d | LGBM | 0.159 | 0.637 | 0.909 | 0.750 |
| MIMIC-III | 2p_7d | CS_CoxPH | 0.230 | 0.562 | 0.820 | 0.590 |
| MIMIC-III | 2p_7d | CS_RSf | 0.206 | 0.398 | 0.687 | 0.481 |
| MIMIC-III | 2p_7d | DeepHit | 0.140 | 0.247 | 0.572 | 0.431 |
| MIMIC-III | 2p_7d | FineGray | 0.228 | 0.580 | 0.835 | 0.607 |
| eICU-CRD | any_48h | LR | 0.210 | 0.779 | 0.941 | 0.731 |
| eICU-CRD | any_48h | XGB | 0.185 | 0.783 | 0.949 | 0.764 |
| eICU-CRD | any_48h | LGBM | 0.229 | 0.740 | 0.920 | 0.691 |
| eICU-CRD | any_48h | CS_CoxPH | 0.251 | 0.745 | 0.914 | 0.662 |
| **eICU-CRD** | any_48h | CS_RSF | 0.222 | 0.615 | 0.861 | 0.638 |
| **eICU-CRD** | any_48h | DeepHit | 0.216 | 0.635 | 0.875 | 0.659 |
| **eICU-CRD** | any_48h | FineGray | 0.246 | 0.768 | 0.926 | 0.680 |
| **eICU-CRD** | any_7d | LR | 0.165 | 0.680 | 0.922 | 0.758 |
| **eICU-CRD** | any_7d | XGB | 0.150 | 0.655 | 0.921 | 0.771 |
| **eICU-CRD** | any_7d | LGBM | 0.172 | 0.641 | 0.904 | 0.732 |
| **eICU-CRD** | any_7d | CS_CoxPH | 0.189 | 0.646 | 0.897 | 0.708 |
| **eICU-CRD** | any_7d | CS_RSF | 0.158 | 0.539 | 0.865 | 0.706 |
| **eICU-CRD** | any_7d | DeepHit | 0.142 | 0.513 | 0.866 | 0.724 |
| **eICU-CRD** | any_7d | FineGray | 0.186 | 0.665 | 0.906 | 0.720 |
| **eICU-CRD** | 2p_48h | LR | 0.032 | 0.324 | 0.934 | 0.902 |
| **eICU-CRD** | 2p_48h | XGB | 0.015 | 0.250 | 0.956 | 0.942 |
| **eICU-CRD** | 2p_48h | LGBM | 0.019 | 0.293 | 0.953 | 0.934 |
| **eICU-CRD** | 2p_48h | CS_CoxPH | 0.049 | 0.243 | 0.847 | 0.797 |
| **eICU-CRD** | 2p_48h | CS_RSF | 0.034 | 0.155 | 0.816 | 0.783 |
| **eICU-CRD** | 2p_48h | DeepHit | 0.038 | 0.165 | 0.809 | 0.772 |
| **eICU-CRD** | 2p_48h | FineGray | 0.045 | 0.266 | 0.876 | 0.831 |
| **eICU-CRD** | 2p_7d | LR | 0.053 | 0.263 | 0.852 | 0.800 |
| **eICU-CRD** | 2p_7d | XGB | 0.026 | 0.288 | 0.935 | 0.909 |
| **eICU-CRD** | 2p_7d | LGBM | 0.032 | 0.280 | 0.919 | 0.887 |
| **eICU-CRD** | 2p_7d | CS_CoxPH | 0.056 | 0.264 | 0.843 | 0.787 |
| **eICU-CRD** | 2p_7d | CS_RSF | 0.043 | 0.174 | 0.795 | 0.752 |
| **eICU-CRD** | 2p_7d | DeepHit | 0.047 | 0.191 | 0.802 | 0.755 |
| **eICU-CRD** | 2p_7d | FineGray | 0.053 | 0.285 | 0.866 | 0.813 |

For each top-10% stratum in the external cohorts: $A/(A + D + C)$ , which equals BRII and assigns all early-censored patients to no target event; $A/(A + D)$ , which leaves them out; $(A + C)/(A + D + C)$ , which assigns all of them to the target event; and $C/(A + D + C)$ , the distance between the first and third values. Values are computed from the stratum counts. The first and third values follow two extreme assumptions about target events that were not observed in early-censored patients; these values are not estimates of BRII under complete follow-up.

**Supplementary Table 20.** Normal-approximation half-width of a 95% interval for BRII by denominator size.

| **m** | **BRII = 0.1** | **BRII = 0.2** | **BRII = 0.3** | **BRII = 0.5** | **BRII = 0.7** | **BRII = 0.9** |
| --- | --- | --- | --- | --- | --- | --- |
| **25** | 0.118 | 0.157 | 0.180 | 0.196 | 0.180 | 0.118 |
| **50** | 0.083 | 0.111 | 0.127 | 0.139 | 0.127 | 0.083 |
| **100** | 0.059 | 0.078 | 0.090 | 0.098 | 0.090 | 0.059 |
| **200** | 0.042 | 0.055 | 0.064 | 0.069 | 0.064 | 0.042 |
| **500** | 0.026 | 0.035 | 0.040 | 0.044 | 0.040 | 0.026 |
| **1,000** | 0.019 | 0.025 | 0.028 | 0.031 | 0.028 | 0.019 |
| **2,000** | 0.013 | 0.018 | 0.020 | 0.022 | 0.020 | 0.013 |
| **5,000** | 0.008 | 0.011 | 0.013 | 0.014 | 0.013 | 0.008 |

Half-width $1.96 \times \sqrt{(BRII(1 - BRII)/m)}$ , where m is the number of patients in A + D + C. In the 84 top-10% units, the bootstrap half-width of Tables 2 and 3 equaled this half-width within rounding in 64 units, was wider in 7, and was narrower in 13; after allowing for the Monte Carlo error of 1,000 resamples, a post hoc calculation, the counts were 77, 1, and 6.

**Supplementary Table 21.** Weighted BRII for any_48h, including a censoring weight of zero.

| Cohort | Model | wC = 0.00 | wC = 0.25 | wC = 0.50 | wC = 0.75 | wC = 1.00 |
| --- | --- | --- | --- | --- | --- | --- |
| MIMIC-III | LR | 0.962 | 0.952 | 0.942 | 0.932 | 0.923 |
| MIMIC-III | XGB | 0.978 | 0.973 | 0.968 | 0.964 | 0.959 |
| MIMIC-III | LGBM | 0.978 | 0.974 | 0.969 | 0.965 | 0.960 |
| MIMIC-III | CS_CoxPH | 0.960 | 0.950 | 0.941 | 0.931 | 0.922 |
| MIMIC-III | CS_RSf | 0.939 | 0.929 | 0.919 | 0.909 | 0.899 |
| MIMIC-III | DeepHit | 0.953 | 0.945 | 0.937 | 0.929 | 0.921 |
| MIMIC-III | FineGray | 0.967 | 0.958 | 0.950 | 0.942 | 0.934 |
| eICU-CRD | LR | 0.779 | 0.464 | 0.330 | 0.256 | 0.210 |
| eICU-CRD | XGB | 0.783 | 0.433 | 0.299 | 0.229 | 0.185 |
| eICU-CRD | LGBM | 0.740 | 0.475 | 0.349 | 0.276 | 0.229 |
| eICU-CRD | CS_CoxPH | 0.745 | 0.500 | 0.376 | 0.301 | 0.251 |
| eICU-CRD | CS_RSf | 0.615 | 0.426 | 0.327 | 0.264 | 0.222 |
| eICU-CRD | DeepHit | 0.635 | 0.428 | 0.323 | 0.259 | 0.216 |
| eICU-CRD | FineGray | 0.768 | 0.502 | 0.372 | 0.296 | 0.246 |

$A/(A + D + wC \cdot C)$ for the any\_48h top-10% strata of the external cohorts, with the death
weight fixed at 1 and the censoring weight wC from 0 to 1, computed from the stratum counts.
wC = 1 is BROI and wC = 0 is $A/(A + D)$ .

### Supplementary Figures

**Supplementary Fig. 1.**
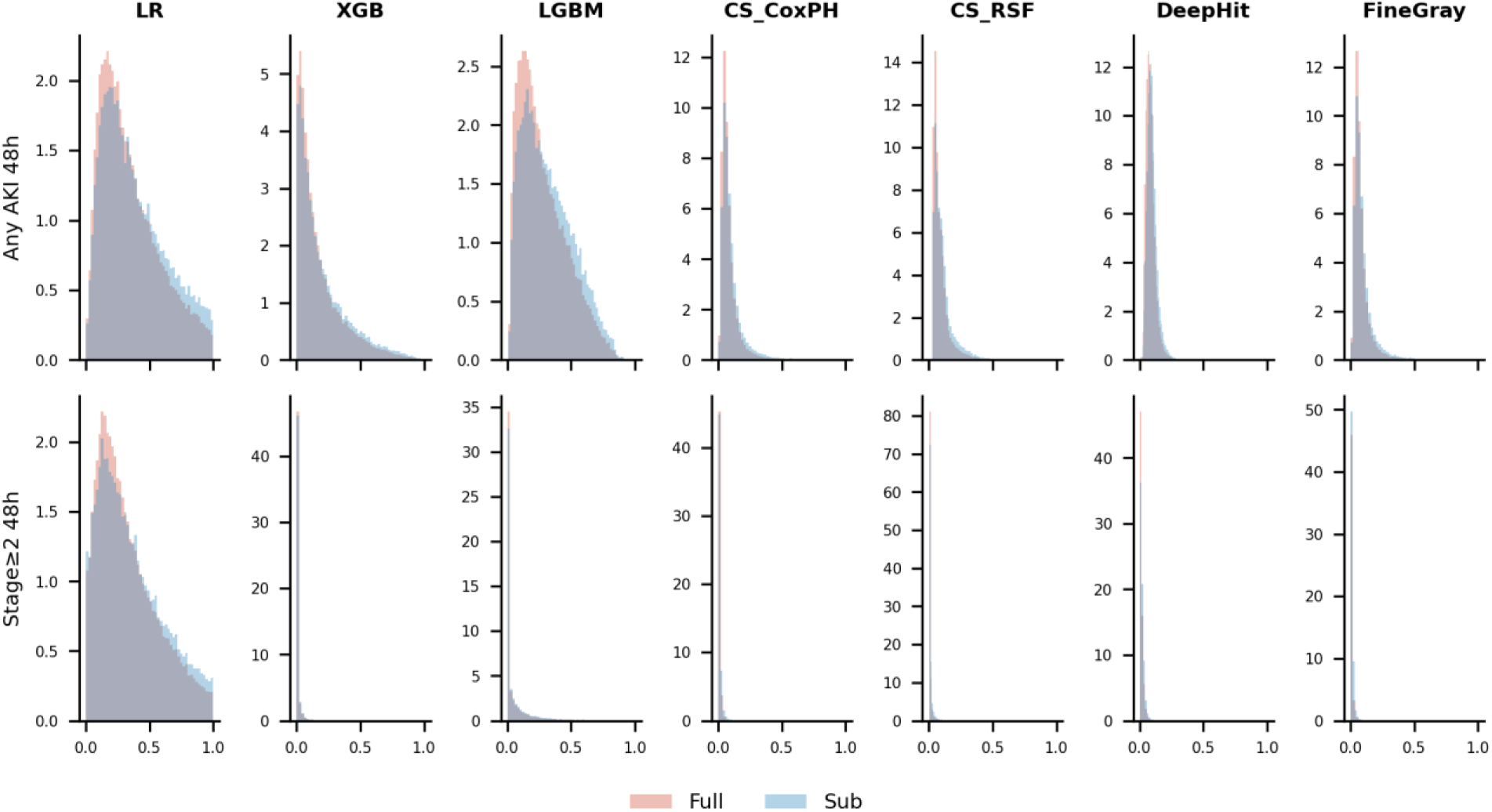
Model score distributions in full versus restricted eICU-CRD cohorts. Histograms of predicted risk scores for seven models in the full eICU-CRD cohort and the subcohort remaining after removal of patients censored before the horizon, for the any_48h and 2p_48h targets.

**Supplementary Fig. 2.**
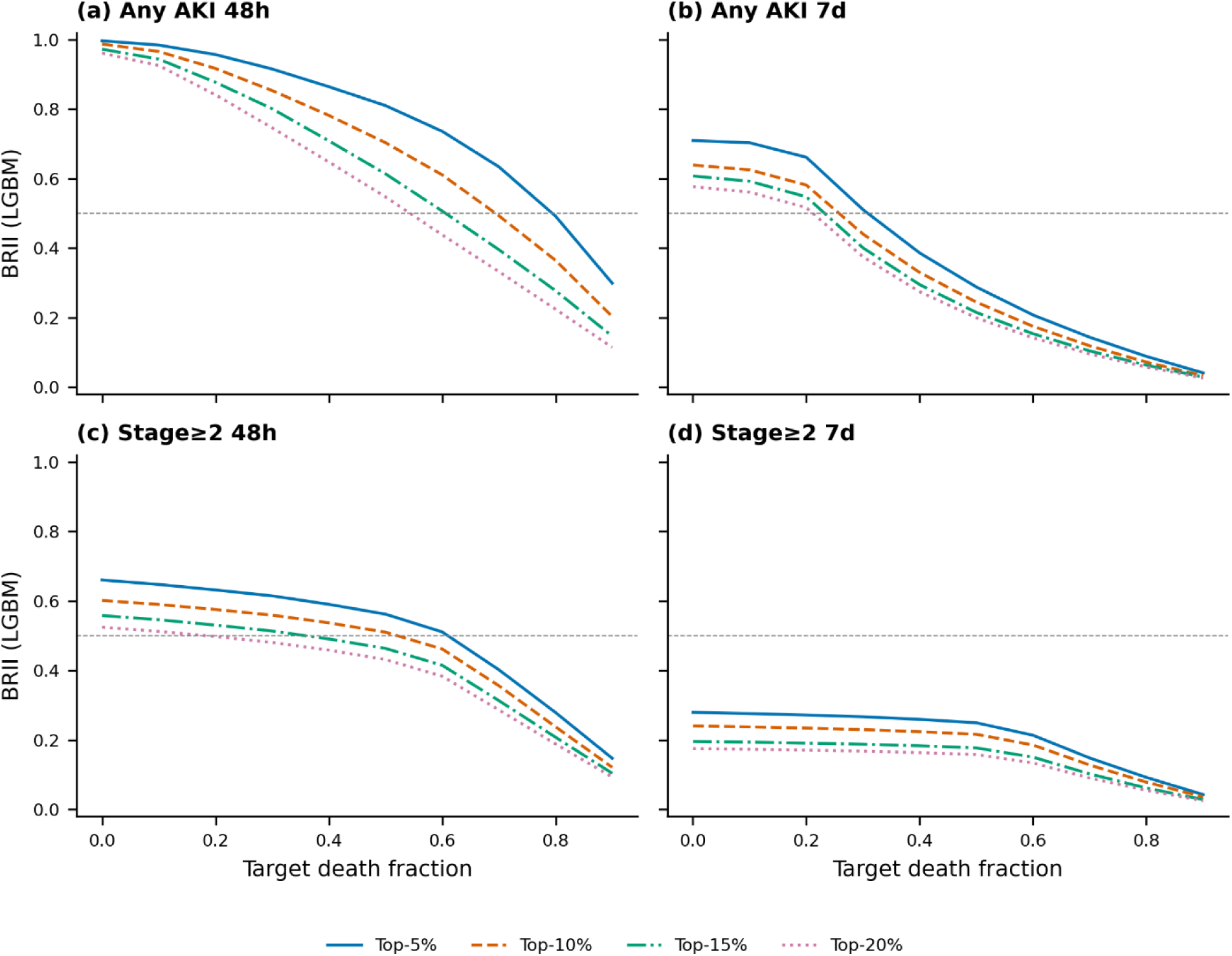
Effect of top-*k* threshold on BRII across death fractions. BRII plotted against target death fraction (0.0-0.9) at four top-*k* thresholds (5%, 10%, 15%, 20%) for a representative model (LightGBM). Panels: (a) any_48h, (b) any_7d, (c) 2p_48h, (d) 2p_7d. Smaller thresholds yield higher BRII.

**Supplementary Fig. 3.**
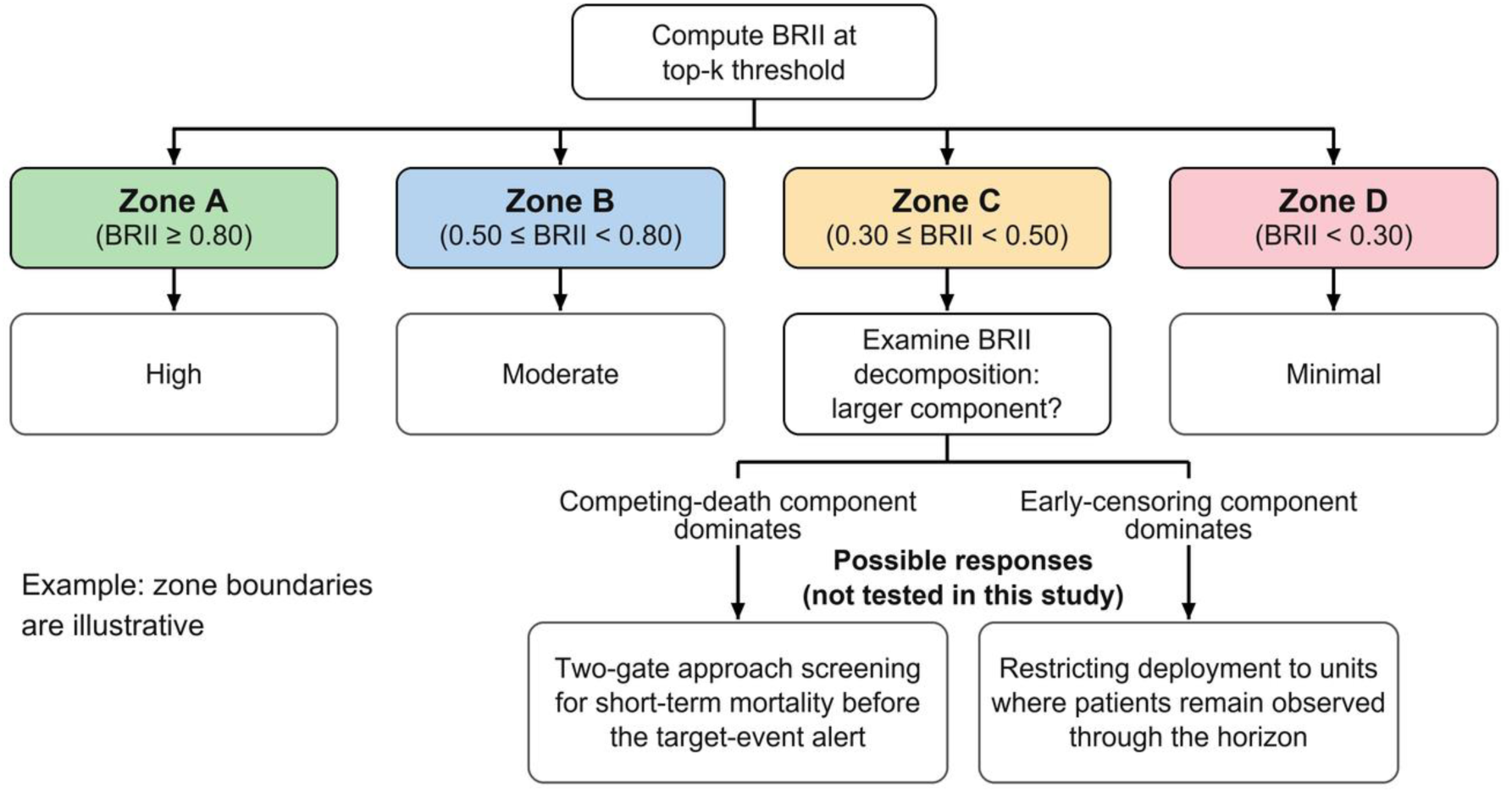
Example flowchart linking BRII zones and decomposition profiles to possible responses. Example flowchart mapping the exploratory BRII zones (Zone A through Zone D) to possible responses. The flowchart proceeds in three stages: zone identification, examination of the decomposition for Zone C, and a possible response for the larger component. The two responses are those discussed in the main text and were not tested in this study. The boxes below Zones A, B, and D give the zone labels of Supplementary Methods S5, and Zone C (limited) leads to the decomposition box. The zones were not externally validated and are not clinical decision thresholds.

**Supplementary Fig. 4.**
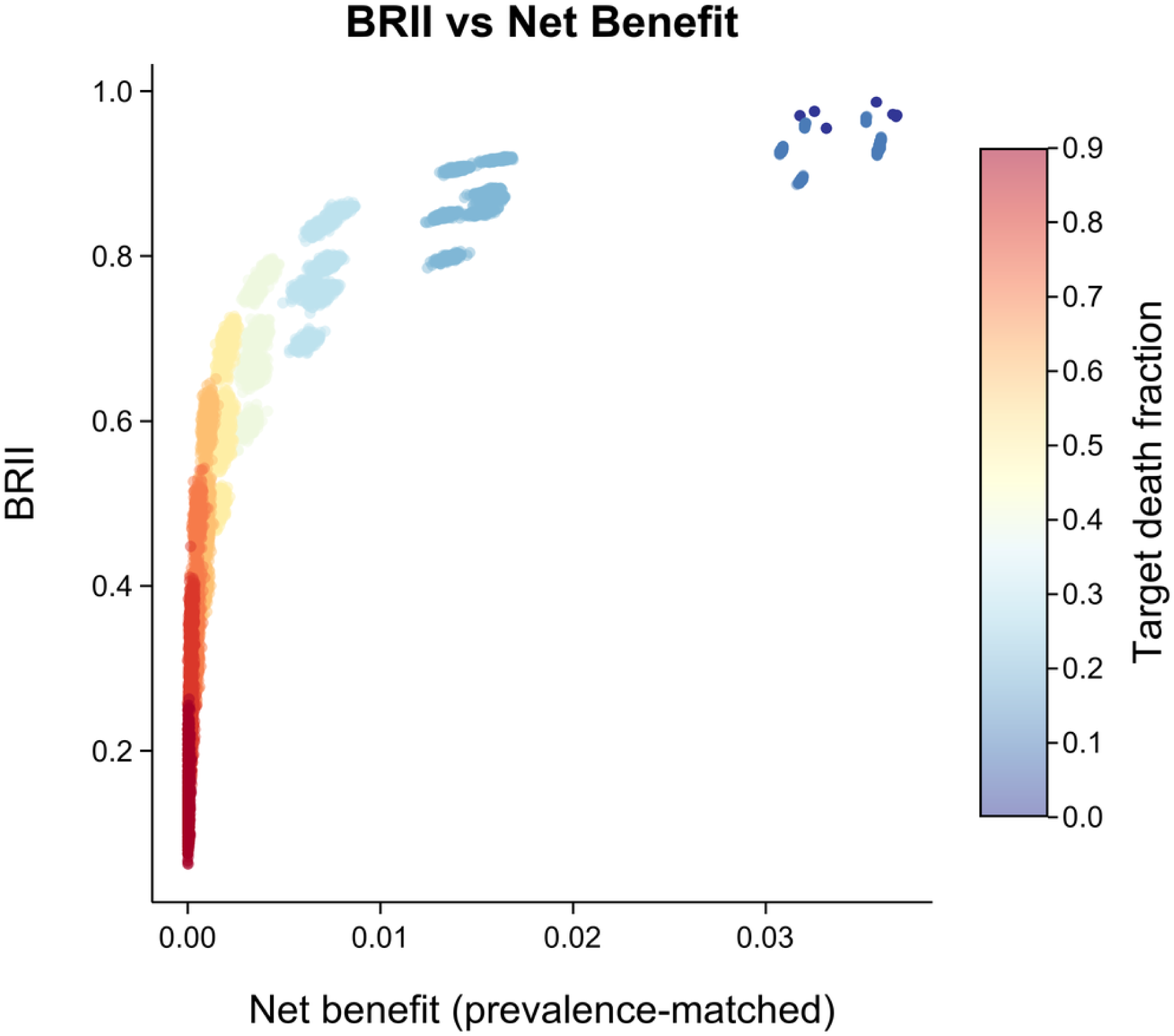
BRII versus net benefit under varying competing-death burden. Scatter plot of BRII against net benefit (at prevalence-matched threshold) across death-contamination levels for the any_48h target, with seven models shown (logistic regression, XGBoost, LightGBM, Cox proportional hazards, random survival forest, Fine-Gray, DeepHit; 200 repetitions per death-fraction level). Points colored by target death fraction. Overall, Spearman ρ = 0.982.

